# Molecular epidemiology of SARS-CoV-2 - a regional to global perspective

**DOI:** 10.1101/2021.01.25.21250447

**Authors:** Christian Brandt, Riccardo Spott, Martin Hölzer, Denise Kühnert, Stephan Fuchs, Mara Lohde, Mike Marquet, Adrian Viehweger, Dagmar Rimek, Mathias W. Pletz

## Abstract

**Background:** After a year of the global SARS-CoV-2 pandemic, a highly dynamic genetic diversity is surfacing. Among nearly 1000 reported virus lineages, dominant lineages such as B.1.1.7 or B.1.351 attract media attention with questions regarding vaccine efficiency and transmission potential. In response to the pandemic, the Jena University Hospital began sequencing SARS-CoV-2 samples in Thuringia in early 2020.

**Methods:** Viral RNA was sequenced in tiled amplicons using Nanopore sequencing. Subsequently, bioinformatic workflows were used to process the generated data. As a genomic background, 9,642 representative SARS-CoV-2 genomes (1,917 of German origin) were extracted from more than 300.000 genomes.

**Results:** In a comprehensive bioinformatics analysis, we have set Thuringian isolates in the German, European and global context. In Thuringia, a largely rural German region without an international airport and a population density below the German average, we discovered many of the common “EU lineages”. German samples are scattered across eight major clades, and Thuringian samples occupy four of them.

**Conclusion:** The rapid emergence and spread of novel variants are of great concern as these lineages could transmit more efficiently, evade current vaccine efforts or undermine diagnostic test accuracy. To anticipate and mitigate these threats, a continuous molecular surveillance is essential.

**Key messages:**

- Bioinformatics analysis of 1,917, 4,251, and 3,474 SARS-CoV-2 genomes from Germany, the EU (except Germany), and non-EU, respectively, subsampled from more than 300,000 public genomes and placed in the context of Thuringian sequences
- Constant antigenic drift for SARS-CoV-2 and no clear pattern or clustering is visible in Thuringia based on the current number of samples
- Currently over 100 described lineages are identified in Germany and only a subset (9) are detected in Thuringia so far, most likely due to genetic undersampling
- From a national perspective, it is likely that high-frequency lineages, which are currently spreading throughout Europe, will eventually also reach Thuringia
- Systematic and dense molecular surveillance via whole-genome sequencing is needed to detect concerning new lineages early, limit spread and adjust vaccines if necessary

## Introduction

In December 2019, health authorities in China reported an outbreak of pneumonia of unknown origin in Wuhan (1). A novel betacoronavirus called SARS-CoV-2 (severe acute respiratory syndrome coronavirus 2) was ultimately identified as the causative agent. SARS-CoV-2 has since spread around the world, causing a global pandemic with more than 80 million confirmed cases and over 1.8 million deaths worldwide in 2020. In response to the pandemic, many countries started to sequence SARS-CoV-2 genomes to track its evolution. To date (January 2021), more than 300,000 SARS-CoV-2 genomes are freely available in public databases such as GISAID or ENA.

Each time the virus replicates, mutants are created that convey genetic flexibility due to the high error rate of the RNA polymerase. Because the virus spread so quickly around the world, it has a huge effective population size, which provides ample opportunity for the virus to adapt to its environment and escape host immunity (2,3). In contrast to other RNA-viruses, SARS-CoV-2 includes a proofreading capability for its RNA polymerase (for viral replication) to reduce mutations (4). However, mutations in this polymerase gene itself have been described, which may affect its proofreading activity and potentially alter the mutation rates of SARS-CoV-2 (5). In general, the spontaneous mutation rate of SARS-CoV-2 is 0.80 - 2.38 × 10^−3^ nucleotide substitutions/site/year, the same order of magnitude as influenza A, which lacks a proofreading RNA polymerase (6,7). The higher the overall number of infections, the higher the likelihood that new virus variants, so-called “lineages” will arise, which may be better adapted and spread more effectively (8). The three main virus nomenclatures introduced for SARS-CoV-2 represent the respective clade names used by Pangolin, Nextstrain and GISAID (8–10) and there is an active discussion to unify and update the system (nature.com https://doi.org/10.1038/d41586-021-00097-w). SARS-CoV-2 is highly dynamic in this regard, reflected in more than 1000 described “lineages” (https://cov-lineages.org/lineages.html) (8). The UK observed over 1000 transmission lineages to date (11) and sets an example of molecular surveillance by contributing 151,860 of all 348,089 SARS-CoV-2 genomes stored on GISAID (data from 11.01.2021). So far they showed that larger regional epidemics had a greater diversity of lineages, and that the proximity to airports in UK areas led to an increase in lineages and their frequency (11).

An increasing number of variants of concern (VOC) have been arising recently, including B.1.1.7 first detected in the UK (12), B.1.351 first detected in South Africa (medRxiv https://doi.org/10.1101/2020.12.21.20248640), B.1.346 first detected in the US (https://virological.org/t/identification-of-a-novel-sars-cov-2-spike-69-70-deletion-lineage-circulating-in-the-united-states) and a mutated variant of B.1.1.28, first detected from four North Brazilian travelers in Japan and recently confirmed in Manaus, Brazil (currently labeled P.1; https://virological.org/t/genomic-characterisation-of-an-emergent-sars-cov-2-lineage-in-manaus-preliminary-findings). B.1.1.7 garnered high media attention due to its increasingly high prevalence in the South East of England and the increased detection of the lineage in other European countries including Germany. Noteworthy, lineage B.1.1.7 shows a large number of mutations with potentially immunologic significance, for instance, in the spike protein (such as N501Y and P681H), compared to the Wuhan strain in 2019. Early investigations indicate that this virus lineage possesses a significant transmission advantage (medRxiv https://doi.org/10.1101/2020.12.30.20249034). In particular, the N501Y mutation was also found in the emerging South African variant B.1.351 next to mutations K417N and E484K in the spike protein (medRxiv https://doi.org/10.1101/2020.12.21.20248640). All three mutations are located in the receptor-binding domain (RBD) of the spike protein and are involved in binding to the human ACE2 receptor. The mutated lineage B.1.1.28 (P.1) harbors, amongst others, the same three RBD mutations (K417N/E484K/N501Y). If these mutations in the RBD indeed provide a selective advantage for viral transmissibility, we expect an increasing frequency of such viral lineages around the world. Only limited changes to the epitope were observed for the strain B.1.1.7 so far, suggesting that it is highly unlikely that current RNA vaccines won’t be effective (medRxiv https://doi.org/10.1101/2021.01.06.20248960).

Overall, numerous lineages have increased and decreased in 2020. Multiple factors influence the spread, like seasonal changes, regional counter measurements, and people’s adherence to these. So far, there has been substantial heterogeneity in the transmissibility of SARS-CoV-2 infection. Adam et al. described that only 19% of cases were responsible for 80% of all SARS-CoV-2 transmissions in Hong Kong (13). Also, Endo et al. suggest a stronger focus on super spreading interventions due to transmission heterogeneity (14).

In early 2020, the Jena Universal Hospital (responsible for Thuringia, Germany) quickly implemented nanopore-based sequencing of SARS-CoV-2 to track viral diversity in the region. Here, we describe 40 Thuringian SARS-CoV-2 genomes and investigate their molecular context within Germany and the rest of the world.

## Methods

### Nanopore sequencing and genome reconstruction

Viral RNA was extracted directly from swabs, transcribed, and amplified via the ARTIC protocol using V3 amplicon primers (dx.doi.org/10.17504/protocols.io.bdp7i5rn). The sequencing was carried out with the MinION sequencer (Oxford Nanopore Technologies), which allows, similar to the more well-known Illumina sequencing, accurate construction of consensus-level SARS-CoV-2 sequences (15). All genomes sequenced at the Jena University Hospital are stored on GISAID under the search term “TH-IIMK-CaSe” (Field: Virus Name). The raw sequence data were processed using the standard settings of the poreCov workflow (v0.3.5; https://github.com/replikation/poreCov) (16), comprising base-calling, genome reconstruction, and SARS-CoV-2 lineage determination via Pangolin (https://github.com/cov-lineages/pangolin).

### Time tree creation

The Nextstrain workflow (10,17) was used for the phylogenetic analysis of the Thuringian samples. To this end, the following subsampling approach was used to reduce the 316,771 genomes from GISAID with a stepwise focus from a German to an EU to a global context. Genomes were grouped if the three criteria “division, year, and month” were the same. German groups were allowed to contain up to 200 genomes, other European countries up to 20, and other non-EU countries up to 10. Excess genomes were removed. This resulted in 1,917, 4,251, and 3,474 genomes from Germany, the EU (except Germany), and non-EU, respectively. Results were visualized via auspice (https://auspice.us/). The tree origin ist the strain “Wuhan/IVDC-HB-01/2019” (Accession Number: NC_045512.2).

## Results

Forty Thuringian SARS-CoV-2 isolates collected between December 2019 and January 2021 were analyzed. To bring the reconstructed SARS-CoV-2 genomes into a molecular context with the rest of Germany and the world, they were phylogenetically compared with 9,642 SARS-CoV-2 genomes. For this purpose, all GISAID sequences (316,771 genomes) were subsampled (for details, see method section), with an emphasis on Germany (n = 1,917). The results are visualized in a phylogenetic time tree (Fig. 1 - samples are placed according to collection date) and a divergence tree (Figure 2 - samples are placed according to nucleotide mutations), in which all German isolates are marked, and the Thuringian isolates are highlighted in red. Both trees are colored by the major Nextstrain clades. These major clades are named based on their year of occurrence and a letter (e.g., 2019 as 19A). A major clade needs to reach >20% global frequency or >30% regional frequency for two or more months to be added as a major clade. Certain clades such as 20 A/B/C are globally distributed and found in most countries (10).

**Fig 1:**
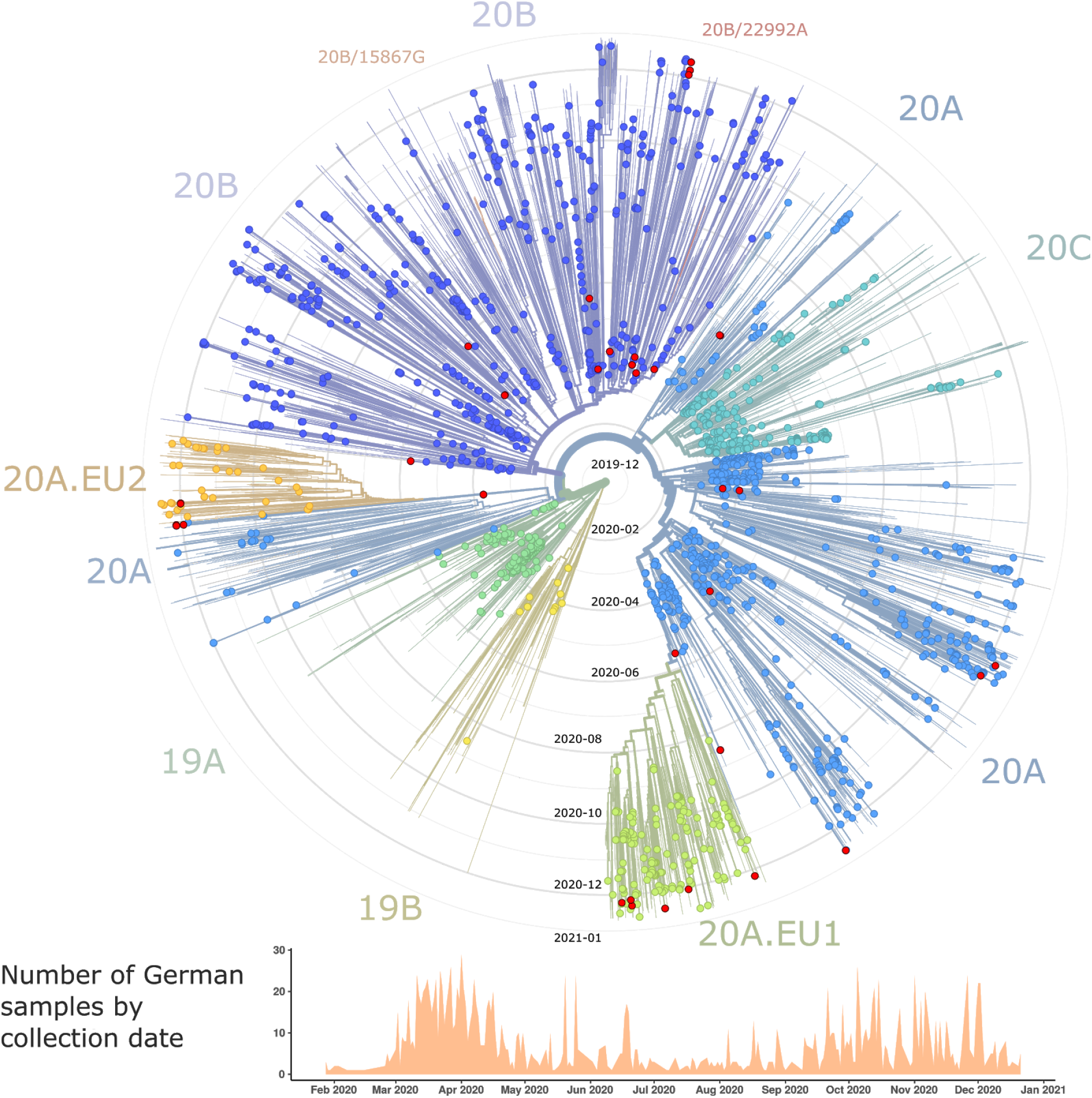
Radial time tree visualization of 9,642 SARS-CoV-2 genomes subsampled from 316,771 genomes. The tree contains 1,917 German samples (country - colored circles), including 40 Thuringian samples (region - red circles). The tree is colored and labeled by the major SARS-CoV-2 clades defined by Nextstrain. Tree origin (center of the tree) is the strain “Wuhan/IVDC-HB-01/2019” collected on 30-12-2019. Rings around the tree represent the time past and are labeled by year-month. The histogram shows the number of German samples included here by collection date.

**Figure 2:**
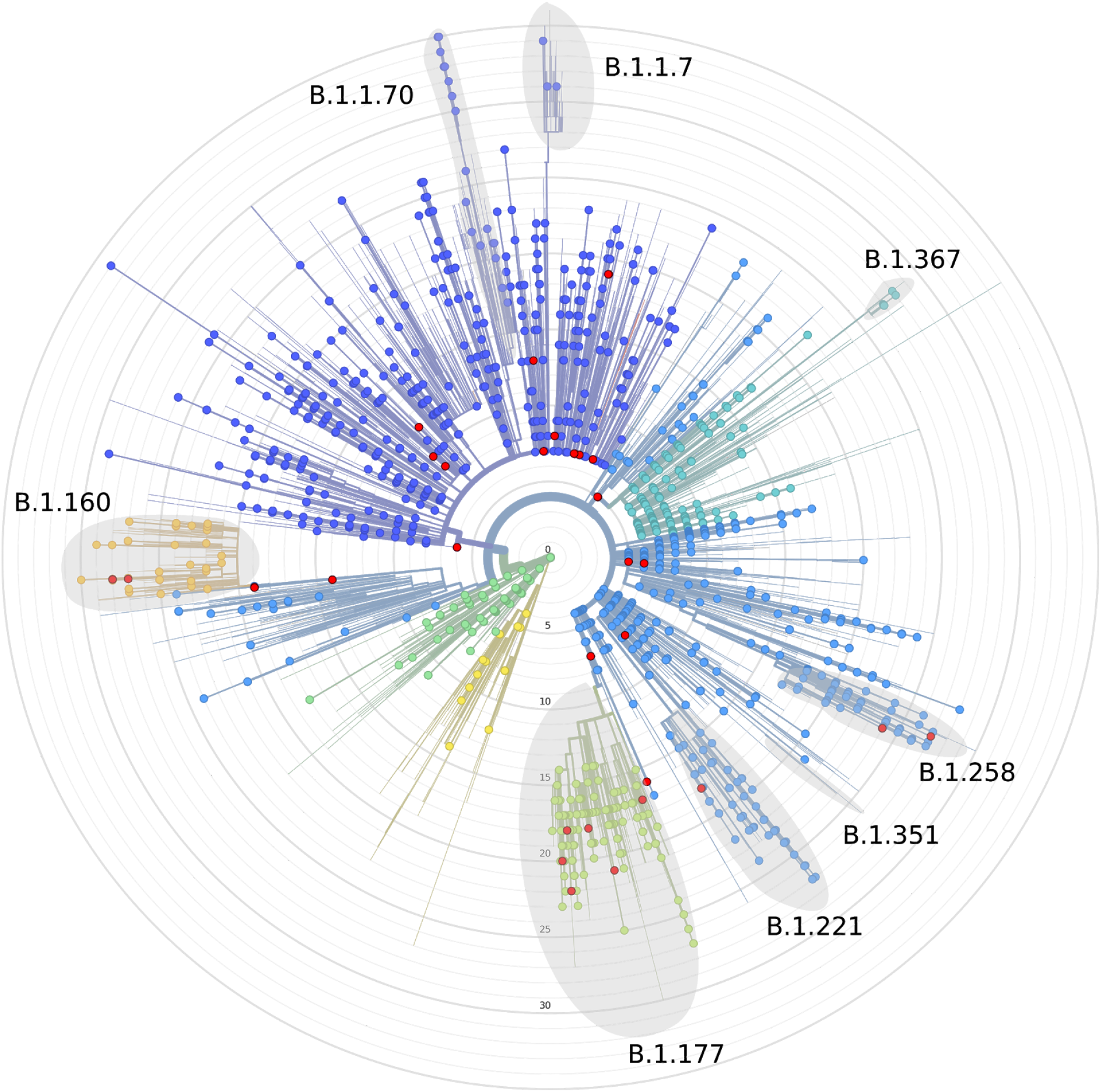
Radial divergence tree of 9,642 SARS-CoV-2 genomes representing the accumulated number of nucleotide mutations (grey rings) from “Wuhan/IVDC-HB-01/2019” submitted on 30-12-2019 (center of the tree). The tree contains 1,917 German samples (country - colored circles), including 40 Thuringian samples (region - red circles). Some lineages of higher divergence of “wave 2” are highlighted. The tree is colored by the major SARS-CoV-2 clades defined by Nextstrain (same color scheme as Figure 1).

### Most highly prevalent SARS-CoV-2 lineages in Germany detected in Thuringia

Thuringia, a region that is geographically in the middle of Germany and Europe, does not have its own international airport. We, therefore, expected lower genetic diversity/frequency in contrast to regions with higher density and international airports (11). However, Thuringian samples (colored red, Fig. 1) are scattered throughout the tree in close proximity to other German samples from other regions, with only a few noticeable exceptions like 19A/B and 20C. Up until now, no clear pattern or clustering is visible in Thuringia based on the current number of samples.

The 1,917 German samples are scattered throughout the major nextstrain clades 19A/B, 20A/B/C, 20A.EU1/2. Four of these clades can be found in Thuringian isolates (20A, 20A.EU1, 20A.EU2, and 20B). In contrast to the beginning of the year, certain major clades such as 19A, 19B are underrepresented in the 2nd wave in Germany up until now, even though isolates have been identified in other countries (colored lines Fig. 1). Noteworthy, a reduction in sampling is clearly visible around June to August when viral prevalence was lowest in Germany (Fig. 1).

More generally, German samples are scattered throughout all of the tree and clades with the exception of the smaller subclades 20B/15867G and 20B/22992A. It is worth noting that SARS-CoV-2 virus nomenclatures are continuously updated and reassigned throughout the pandemic to better reflect current trends and frequencies.

### Genetic divergence and current lineage distribution

Although SARS-CoV-2 is not a recombinant of any sarbecoviruses detected to date, recombination likely played a role in its emergence (18,19). A recombination event between SARS-CoV-2 lineages has, to our knowledge, not yet been observed or proven (https://observablehq.com/@spond/linkage-disequilibirum-in-sars-cov-2). The virus has been subject to constant antigenic drift with around 22.8 substitutions per year. To better understand the molecular viral epidemiology and clonality in Thuringia, the 9,642 SARS-CoV-2 isolates were visualized on the basis of their divergence from the Wuhan isolate (Fig. 2). We detected over 110 different lineages in the 1,917 German samples, and highlighted the frequent lineages of “wave 2” (winter 2020/21; Fig. 2).

The most divergent genomes sequenced in Thuringia represent lineages B.1.177, B.1.221, B.1.258, and B.1.160, covering most of the diversity observed in German samples, with a few exceptions (B.1.1.7, B.1.367, and B.1.1.70). Both B.1.177 (>72,000 sequences worldwide) and B.1.160 (around 10,000 sequences worldwide) increased rapidly in their frequency over the summer of 2020 and are now frequent and mainly identified in European countries (2020-08 and ongoing; https://virological.org/t/preliminary-genomic-characterisation-of-an-emergent-sars-cov-2-lineage-in-the-uk-defined-by-a-novel-set-of-spike-mutations). Both lineages make up a large part of the EU clades 20A.EU1 for B.1.177 and 20A.EU2 for B.1.160. Various emerging sublineages have been identified (e.g., B.1.160.1 to B.1.160.8; B.1.177.1 to B.1.177.27), reflecting their high prevalence. Similarly, B.1.258 (around 6,601 sequences worldwide) and B.1.221 (around 3,500 sequences worldwide) increased their frequency in Europe around October-December 2020 and are part of clade 20A.

## Discussion

At the time of writing we are in the middle of the COVID-19 pandemic. Although vaccination has recently started, the emergence of VOCs diminishes our hope that the pandemic is nearly over. Molecular surveillance is invaluable in understanding a pathogen’s spread and evolution and can help inform public health decisions. We’ve presented a relatively sparse genomic overview of the Thuringian virus diversity during the pandemic so far, which was possible thanks to employing a low-cost nanopore-based SARS-CoV-2 sequencing approach, together with bioinformatic workflows to automate all subsequent analyses reproducibly.

In Thuringia, a region without an international airport and with a population density below the German average, we detected many of the currently highly frequent lineages in Europe. The current data suggest that any highly frequent lineage spreading throughout Europe will reach Thuringia at some point. While currently over 100 described lineages are identified in Germany (data from GISAID; 21.01.2021), we only detected a subset (9) of these lineages, which may be attributed to undersampling. Denser genomic coverage would probably reveal higher diversity, which in turn would increase the probability to detect novel variants early. While it is imperative to implement systematic genomic surveillance throughout Germany and Europe, care needs to be taken in interpreting the results of genomic investigations. Viral transmission dynamics depend on a range of factors, such as social interactions, regional COVID measures, workplace (location, number of coworkers, space…), etc.. Regional differences can lead to considerable fluctuations in the frequency of mutations. Therefore, solid estimates on transmissibility require clinical field studies and respective modeling, e.g. through estimation of secondary attack rates within households. Indeed, more within-household transmission for the new variant VOC B.1.1.7 have been reported (https://www.ons.gov.uk/peoplepopulationandcommunity/healthandsocialcare/conditionsanddiseases/adhocs/12714newvariantclusteringinhouseholdsanalysis).

Further SARS-CoV-2 VOCs are likely to emerge in different regions of the world. Since most VOCs cannot be discriminated against by regular PCR-tests, continuous genomic surveillance by sequencing is required. Although no lineages that circumvent the efficiency of currently used vaccines have yet been observed, vaccination induces additional selective pressure on the virus. Systematic and dense whole-genome sequencing will enable us to detect concerning new lineages early to quickly limit their spread and, if needed, adjust the vaccines and/or the routine diagnostic standards.

## Data Availability

All data is available on GISAID.

## Acknowledgment

We thank Andreas Sachse, Grit Schubert, and Ariane Düx from the RKI P3 (Epidemiology of highly pathogenic microorganisms) and Thorsten Wolff from the RKI FG17 (Influenza and other respiratory viruses) for the additional Thuringian genome sequences generated and uploaded over Christmas 2020.

We gratefully acknowledge the following Authors from the Originating laboratories responsible for obtaining the specimens, as well as the Submitting laboratories where the genome data were generated and shared via GISAID, on which this research is based. All Submitters of data may be contacted directly via www.gisaid.org. All the Submitting and Originating laboratories and Authors are acknowledged in the Supplementary Table S1.

## Notes

### Competing Interest Statement

The authors have declared no competing interest.

### Funding Statement

The work is funded by the German Ministry of Education and Research (BMBF), grant number 01KX2021, and the Thuringian Region Government, grant number TZUZI82094.

## Literature

1. Wang C, Horby PW, Hayden FG, Gao GF: A novel coronavirus outbreak of global health concern. The Lancet 2020; 395: 470–3.

2. Domingo E, Holland JJ: Rna virus mutations and fitness for survival. Annu Rev Microbiol 1997; 51: 151–78.

3. Domingo E: Viruses at the Edge of Adaptation. Virology 2000; 270: 251–3.

4. Robson F, Khan KS, L. TK, et al.: Coronavirus RNA Proofreading: Molecular Basis and Therapeutic Targeting. Mol Cell 2020; 79: 710–27.

5. Pachetti M, Marini B, Benedetti F, et al.: Emerging SARS-CoV-2 mutation hot spots include a novel RNA-dependent-RNA polymerase variant. J Transl Med 2020; 18: 179.

6. Nobusawa E, Sato K: Comparison of the Mutation Rates of Human Influenza A and B Viruses. J Virol 2006; 80: 3675–8.

7. Zhao Z, Li H, Wu X, et al.: Moderate mutation rate in the SARS coronavirus genome and its implications. BMC Evol Biol 2004; 4: 21.

8. Rambaut A, Holmes EC, O’TooleÁ, et al.: A dynamic nomenclature proposal for SARS-CoV-2 lineages to assist genomic epidemiology. Nat Microbiol 2020; 5: 1403–7.

9. Alm E, Broberg EK, Connor T, et al.: Geographical and temporal distribution of SARS-CoV-2 clades in the WHO European Region, January to June 2020. Eurosurveillance 2020; 25: 2001410.

10. Hadfield J, Megill C, Bell SM, et al.: Nextstrain: real-time tracking of pathogen evolution. Bioinformatics 2018; 34: 4121–3.

11. Plessis L du, McCrone JT, Zarebski AE, et al.: Establishment and lineage dynamics of the SARS-CoV-2 epidemic in the UK. Science 2021;.

12. Wise J: Covid-19: New coronavirus variant is identified in UK. BMJ 2020; 371: m4857.

13. Adam DC, Wu P, Wong JY, et al.: Clustering and superspreading potential of SARS-CoV-2 infections in Hong Kong. Nat Med 2020; 26: 1714–9.

14. Endo A, Centre for the Mathematical Modelling of Infectious Diseases COVID-19 Working Group, Abbott S, Kucharski AJ, Funk S: Estimating the overdispersion in COVID-19 transmission using outbreak sizes outside China. Wellcome Open Res 2020; 5: 67.

15. Bull RA, Adikari TN, Ferguson JM, et al.: Analytical validity of nanopore sequencing for rapid SARS-CoV-2 genome analysis. Nat Commun 2020; 11: 6272.

16. Hufsky F, Lamkiewicz K, Almeida A, et al.: Computational Strategies to Combat COVID-19: Useful Tools to Accelerate SARS-CoV-2 and Coronavirus Research. 2020;.

17. Sagulenko P, Puller V, Neher RA: TreeTime: Maximum-likelihood phylodynamic analysis. Virus Evol 2018; 4.

18. Andersen KG, Rambaut A, Lipkin WI, Holmes EC, Garry RF: The proximal origin of SARS-CoV-2. Nat Med 2020; 26: 450–2.

19. Hu B, Guo H, Zhou P, Shi Z-L: Characteristics of SARS-CoV-2 and COVID-19. Nat Rev Microbiol 2020;: 1–14.

