## Supplementary Table S1 for "Molecular epidemiology of SARS-CoV-2 - a regional to global perspective"

We gratefully acknowledge the following Authors from the Originating laboratories responsible for obtaining the specimens and the
Submitting laboratories where genetic sequence data were generated and shared via the GISAID Initiative, on which this research is based.

| Accession ID | Originating Laboratory | Submitting Laboratory | Authors |
|---|---|---|---|
| EPI_ISL_707792; EPI_ISL_763067; EPI_ISL_635060; EPI_ISL_707793; EPI_ISL_763065; EPI_ISL_699656; EPI_ISL_632310; EPI_ISL_634977; EPI_ISL_707791; EPI_ISL_733500; EPI_ISL_635061; EPI_ISL_733499 | 1-Laboratory of Microbiology, National Reference Lab, Charles Nicolle Hospital; 2-University of Tunis ElManar, Faculty of Medicine of Tunis, LR99ES09, Tunis, Tunisia | 1-Clinical and Experimental Pharmacology Lab, LR16SP02, National Center of Pharmacovigilance, University of Tunis El Manar, Tunis, Tunisia. 2-Neurodegenerative diseases and psychiatric troubles, LR18SP03, Razi Hospital, University of Tunis El Manar, Tunis, Tunisia. 3- Ministry of Health, National Observatory of New and Emerging Diseases, 1006, Tunis, Tunisia | Ilhem Boutiba-Ben Boubaker et al |
| EPI_ISL_451978; EPI_ISL_451979 | 1. ViroGenetics - BSL3 Laboratory of Virology, Małopolska Centre of Biotechnology, Jagiellonian University; 2. II Department of Internal Medicine, Faculty of Medicine, Jagiellonian University Medical College; 3. DIAGNOSTYKA Ltd. | 1. ViroGenetics - BSL3 Laboratory of Virology, Małopolska Centre of Biotechnology, Jagiellonian University; 2. II Department of Internal Medicine, Faculty of Medicine, Jagiellonian University Medical College. | Marek Sanak et al |
| EPI_ISL_455452; EPI_ISL_492067; EPI_ISL_455442; EPI_ISL_455449; EPI_ISL_455440; EPI_ISL_455445; EPI_ISL_455446; EPI_ISL_455444; EPI_ISL_455453 | 1. ViroGenetics - BSL3 Laboratory of Virology, Małopolska Centre of Biotechnology, Jagiellonian University; 2. II Department of Internal Medicine, Faculty of Medicine, Jagiellonian University Medical College; 3. Narodowy Instytut Zdrowia Publicznego – Państwowy Zakład Higieny (NIZP-PZH) | 1. ViroGenetics - BSL3 Laboratory of Virology, Małopolska Centre of Biotechnology, Jagiellonian University; 2. II Department of Internal Medicine, Faculty of Medicine, Jagiellonian University Medical College; 3. Narodowy Instytut Zdrowia Publicznego – Państwowy Zakład Higieny (NIZP-PZH). | Katarzyna Pancer et al |
| EPI_ISL_729589; EPI_ISL_729461; EPI_ISL_729536; EPI_ISL_729716; EPI_ISL_729717; EPI_ISL_729572; EPI_ISL_729471; EPI_ISL_729472; EPI_ISL_729419; EPI_ISL_729627; EPI_ISL_729594; EPI_ISL_729655; EPI_ISL_729635; EPI_ISL_729338; EPI_ISL_729682; EPI_ISL_729720; EPI_ISL_729583; EPI_ISL_729668; EPI_ISL_729699; EPI_ISL_729700; EPI_ISL_729732; EPI_ISL_729715; EPI_ISL_729705; EPI_ISL_729347; EPI_ISL_729703; EPI_ISL_729701; EPI_ISL_729702; EPI_ISL_729707; EPI_ISL_729591; EPI_ISL_729621; EPI_ISL_729501; EPI_ISL_729725; EPI_ISL_729620; EPI_ISL_729638; EPI_ISL_729724; EPI_ISL_729710; EPI_ISL_729394; EPI_ISL_729376; EPI_ISL_729613; EPI_ISL_729374; EPI_ISL_729389; EPI_ISL_729341; EPI_ISL_729435; EPI_ISL_729430; EPI_ISL_729428; EPI_ISL_729434; EPI_ISL_729426; EPI_ISL_729581; EPI_ISL_729470; EPI_ISL_729469; EPI_ISL_729562; EPI_ISL_729563; EPI_ISL_729608; EPI_ISL_729666; EPI_ISL_729628; EPI_ISL_729663; EPI_ISL_729662; EPI_ISL_729631; EPI_ISL_729610; EPI_ISL_729354; EPI_ISL_729361; EPI_ISL_729363; EPI_ISL_729629; EPI_ISL_729694; EPI_ISL_729584; EPI_ISL_729654; EPI_ISL_729568; EPI_ISL_729570; EPI_ISL_729569; EPI_ISL_729343; EPI_ISL_729571; EPI_ISL_729573; EPI_ISL_729367; EPI_ISL_729403; EPI_ISL_729407; EPI_ISL_729340; EPI_ISL_729576; EPI_ISL_729574; EPI_ISL_729577; EPI_ISL_729575; EPI_ISL_729580; EPI_ISL_729381; EPI_ISL_729359; EPI_ISL_729590; EPI_ISL_729399; EPI_ISL_729592; EPI_ISL_729593; EPI_ISL_729386; EPI_ISL_729414; EPI_ISL_729542; EPI_ISL_729586; EPI_ISL_729587; EPI_ISL_729344; EPI_ISL_729637; EPI_ISL_729369; EPI_ISL_729462; EPI_ISL_729686; EPI_ISL_729660; EPI_ISL_729351; EPI_ISL_729375; EPI_ISL_729401; EPI_ISL_729438; EPI_ISL_729439; EPI_ISL_729429; EPI_ISL_729427; EPI_ISL_729349; EPI_ISL_729383; EPI_ISL_729544; EPI_ISL_729611; EPI_ISL_729523; EPI_ISL_729543; EPI_ISL_729526; EPI_ISL_729541; EPI_ISL_729545; EPI_ISL_729588; EPI_ISL_729550; EPI_ISL_729551; EPI_ISL_729502; EPI_ISL_729528; EPI_ISL_729524; EPI_ISL_729380; EPI_ISL_729522; EPI_ISL_729540; EPI_ISL_729465; EPI_ISL_729539; EPI_ISL_729538; EPI_ISL_729508; EPI_ISL_729510; EPI_ISL_729503; EPI_ISL_729505; EPI_ISL_729506; EPI_ISL_729504; EPI_ISL_729498; EPI_ISL_729517; EPI_ISL_729525; EPI_ISL_729547; EPI_ISL_729546; EPI_ISL_729350; EPI_ISL_729337; EPI_ISL_729397; EPI_ISL_729513; EPI_ISL_729512; EPI_ISL_729466; EPI_ISL_729362; EPI_ISL_729609; EPI_ISL_729616; EPI_ISL_729619; EPI_ISL_729615; EPI_ISL_729379; EPI_ISL_729390; EPI_ISL_729618; EPI_ISL_729617; EPI_ISL_729674; EPI_ISL_729521; EPI_ISL_729520; EPI_ISL_729552; EPI_ISL_729537; EPI_ISL_729500; EPI_ISL_729515; EPI_ISL_729527; EPI_ISL_729549; EPI_ISL_729516; EPI_ISL_729519; EPI_ISL_729535; EPI_ISL_729684; EPI_ISL_729579; EPI_ISL_729578; EPI_ISL_729582; EPI_ISL_729729; EPI_ISL_729691; EPI_ISL_729657; EPI_ISL_729622; EPI_ISL_729727; EPI_ISL_729709; EPI_ISL_729726; EPI_ISL_729719; EPI_ISL_729649; EPI_ISL_729595; EPI_ISL_729396; EPI_ISL_729625; EPI_ISL_729669; EPI_ISL_729339; EPI_ISL_729395; EPI_ISL_729623; EPI_ISL_729633; EPI_ISL_729464; EPI_ISL_729373; EPI_ISL_729585; EPI_ISL_729695; EPI_ISL_729687; EPI_ISL_729731; EPI_ISL_729734; EPI_ISL_729688; EPI_ISL_729670; EPI_ISL_729451; EPI_ISL_729453; EPI_ISL_729457; EPI_ISL_729458; EPI_ISL_729422; EPI_ISL_729423; EPI_ISL_729441; EPI_ISL_729424; EPI_ISL_729432; EPI_ISL_729425; EPI_ISL_729358; EPI_ISL_729433; EPI_ISL_729437; EPI_ISL_729436; EPI_ISL_729421; EPI_ISL_729447; EPI_ISL_729460; EPI_ISL_729456; EPI_ISL_729443; EPI_ISL_729392; EPI_ISL_729454; EPI_ISL_729449; EPI_ISL_729450; EPI_ISL_729342; EPI_ISL_729455; EPI_ISL_729431; EPI_ISL_729440; EPI_ISL_729445; EPI_ISL_729446; EPI_ISL_729452; EPI_ISL_729442; EPI_ISL_729352; EPI_ISL_729448; EPI_ISL_729444; EPI_ISL_729364; EPI_ISL_729411; EPI_ISL_729641; EPI_ISL_729509; EPI_ISL_729507; EPI_ISL_729677; EPI_ISL_729566; EPI_ISL_729565; EPI_ISL_729564; EPI_ISL_729567; EPI_ISL_729672; EPI_ISL_729634; EPI_ISL_729681; EPI_ISL_729463; EPI_ISL_729624; EPI_ISL_729652; EPI_ISL_729645; EPI_ISL_729676; EPI_ISL_729671; EPI_ISL_729518; EPI_ISL_729511; EPI_ISL_729417; EPI_ISL_729418; EPI_ISL_729345; EPI_ISL_729722; EPI_ISL_729698; EPI_ISL_729733; EPI_ISL_729626; EPI_ISL_729712; EPI_ISL_729689; EPI_ISL_729664; EPI_ISL_729612; EPI_ISL_729614; EPI_ISL_729640; EPI_ISL_729673; EPI_ISL_729636; EPI_ISL_729653; EPI_ISL_729704; EPI_ISL_729658; EPI_ISL_729685; EPI_ISL_729696; EPI_ISL_729706; EPI_ISL_729721; EPI_ISL_729697; EPI_ISL_729384; EPI_ISL_729693; EPI_ISL_729730; EPI_ISL_729368; EPI_ISL_729723; EPI_ISL_729643; EPI_ISL_729714; EPI_ISL_729644; EPI_ISL_729708; EPI_ISL_729692; EPI_ISL_729690; EPI_ISL_729630; EPI_ISL_729661; EPI_ISL_729420; EPI_ISL_729459; EPI_ISL_729372; EPI_ISL_729659; EPI_ISL_729651; EPI_ISL_729647; EPI_ISL_729371; EPI_ISL_729632; EPI_ISL_729675; EPI_ISL_729679; EPI_ISL_729650; EPI_ISL_729648; EPI_ISL_729646; EPI_ISL_729665; EPI_ISL_729391; EPI_ISL_729711; EPI_ISL_729642; EPI_ISL_729656; EPI_ISL_729639; EPI_ISL_729467; EPI_ISL_729416; EPI_ISL_729385; EPI_ISL_729667; EPI_ISL_729728; EPI_ISL_729678; EPI_ISL_729713; EPI_ISL_729680 | A. Krumbholz, Labor Dr. Krause und Kollegen MVZ GmbH, Kiel | Charité Universitätsmedizin Berlin, Institut für Virologie | Victor M Corman et al A |
| EPI_ISL_467463; EPI_ISL_467433; EPI_ISL_467451; EPI_ISL_467468; EPI_ISL_467434; EPI_ISL_467471 | AMPATH-DBN | KRISP, KZN Research Innovation and Sequencing Platform | Giandhari J et al |
| EPI_ISL_458150 | ANOUAL | ANOUAL | Jouali Farah et al (https://dx.doi.org/10.1128/MRA.00690-20) |
| EPI_ISL_682235 | AREA DE SALUD ALAJUELA NORTE - CLINICA DR. MARCIAL RODRIGUEZ | Inciensa, Instituto Costarricense de Investigación y Enseñanza en Nutrición y Salud | Francisco Duarte et al |
| EPI_ISL_682237 | AREA DE SALUD CORREDORES | Inciensa, Instituto Costarricense de Investigación y Enseñanza en Nutrición y Salud | Francisco Duarte et al |
| EPI_ISL_682238 | AREA DE SALUD LA CRUZ | Inciensa, Instituto Costarricense de Investigación y Enseñanza en Nutrición y Salud | Francisco Duarte et al |
| EPI_ISL_418001 | ARS Algarve - Laboratório Laura Ayres | Instituto Nacional de Saude (INSA) | Guiomar et al et al |
| EPI_ISL_542227; EPI_ISL_542443; EPI_ISL_542401; EPI_ISL_542406; EPI_ISL_542241 | ASST GOM Niguarda | Dep. Of Oncology and Hemato-Oncology University of Milan | Claudia Alteri et al |
| EPI_ISL_735501 | Abdul Malek Ukil Medical College, Noakhali | Central Biological Research Laboratory and Department of Biochemistry and Molecular Biology | H. M. Abdullah Al Masud et al |
| EPI_ISL_517635; EPI_ISL_518806; EPI_ISL_517658; EPI_ISL_518808; EPI_ISL_518810; EPI_ISL_517653; EPI_ISL_517662; EPI_ISL_517616; EPI_ISL_517648; EPI_ISL_517652; EPI_ISL_518803; EPI_ISL_517661; EPI_ISL_518801; EPI_ISL_517621; EPI_ISL_517626; EPI_ISL_517622; EPI_ISL_517659; EPI_ISL_517660; EPI_ISL_518809; EPI_ISL_517651; EPI_ISL_517650; EPI_ISL_518800; EPI_ISL_517647 | Academic Hospital Paramaribo | Erasmus Medical Center | Bas Oude Munnink et al |
| EPI_ISL_458083 | Adi Husada Undaan Hospital | Institute of Tropical Disease, Universitas Airlangga | Rima R Prasetya et al |
| EPI_ISL_528947 | Agenzia di Tutela della Salute di Bergamo | Istituto Zooprofilattico Sperimentale dell'Abruzzo e Molise "G.Caporale" | Lorusso A et al |
| EPI_ISL_549170; EPI_ISL_500794; EPI_ISL_590892; EPI_ISL_500796; EPI_ISL_590890; EPI_ISL_420139; EPI_ISL_420134; EPI_ISL_549091 | Akershus University Hospital, Department for Microbiology and Infectious Disease Control | Norwegian Institute of Public Health, Department of Virology | Kathrine Stene-Johansen et al |
| EPI_ISL_661272 | Al-Quds Nutrition and Health Research Institute, Al-Quds University | Al-Quds Nutrition and Health Research Institute, Al-Quds University | Ereqat et al |
| EPI_ISL_649153; EPI_ISL_739659; EPI_ISL_739660; EPI_ISL_739661 | Al-Quds Nutrition and Health Research Institute, Al-Quds University | Al-Quds Nutrition and Health Research Institute, Al-Quds University | Nasereddin et al |
| EPI_ISL_528694; EPI_ISL_528711; EPI_ISL_528707 | Alsafar - Khalifa University Abu Dhabi | Alsafar - Khalifa University Abu Dhabi | Andreas Henschel et al |
| EPI_ISL_569882; EPI_ISL_569870; EPI_ISL_569878; EPI_ISL_569883; EPI_ISL_569865; EPI_ISL_569872 | Amedeo di savoia | Crosetto lab, Karolinska Institutet, SciLifeLab | Michele Simonetti et al |
| EPI_ISL_450818 | Aneby VC | The Public Health Agency of Sweden | Ken Granath et al |
| EPI_ISL_717714; EPI_ISL_717715; EPI_ISL_717717; EPI_ISL_717716 | Animal Health Centre, British Columbia Ministry of Agriculture | National Centre for Foreign Animal Disease, Canadian Food Inspection Agency | Oliver Lung et al |
| EPI_ISL_512669; EPI_ISL_527740; EPI_ISL_512664 | Area De Salud Alajuela Norte - Clinica Dr. Marcial Rodriguez | Inciensa, Instituto Costarricense de Investigación y Enseñanza en Nutrición y Salud | Francisco Duarte et al |
| EPI_ISL_527756 | Area De Salud Aserri | Inciensa, Instituto Costarricense de Investigación y Enseñanza en Nutrición y Salud | Francisco Duarte et al |
| EPI_ISL_512668; EPI_ISL_527751 | Area De Salud Corredores | Inciensa, Instituto Costarricense de Investigación y Enseñanza en Nutrición y Salud | Francisco Duarte et al |
| EPI_ISL_512662; EPI_ISL_512666; EPI_ISL_512667; EPI_ISL_527747 | Area De Salud La Cruz | Inciensa, Instituto Costarricense de Investigación y Enseñanza en Nutrición y Salud | Francisco Duarte et al |
| EPI_ISL_512661 | Area De Salud Los Chiles | Inciensa, Instituto Costarricense de Investigación y Enseñanza en Nutrición y Salud | Francisco Duarte et al |
| EPI_ISL_500706; EPI_ISL_527047; EPI_ISL_527046; EPI_ISL_527028; EPI_ISL_527029; EPI_ISL_509510; EPI_ISL_500704; EPI_ISL_527014 | Area of Virology, Serology and Virology Division (SAViD), New South Wales Health Pathology Randwick | Area of Virology, Serology and Virology Division (SAViD), New South Wales Health Pathology Randwick | Rawlinson et al |
| EPI_ISL_717707; EPI_ISL_678351; EPI_ISL_717701; EPI_ISL_717702; EPI_ISL_717711; EPI_ISL_678362; EPI_ISL_707898; EPI_ISL_678310; EPI_ISL_717705; EPI_ISL_678323; EPI_ISL_678370; EPI_ISL_678377; EPI_ISL_678376; EPI_ISL_678375 | Area of Virology, Serology and Virology Division (SAViD), New South Wales Health Pathology Randwick | Virology Research Laboratory; Area of Virology, Serology and Virology Division (SAViD), New South Wales Health Pathology Randwick | Foster et al |
| EPI_ISL_406223 | Arizona Department of Health Services | Pathogen Discovery, Respiratory Viruses Branch, Division of Viral Diseases, Centers for Disease Control and Prevention | Ying Tao et al B (https://dx.doi.org/10.1101/2020.03.09.20032896) |
| EPI_ISL_457826 | Army Medical Center - Scientific Department | Army Medical and Veterinary Research Center | Silvia Fillo et al |
| EPI_ISL_717978 | Army Medical Center, Scientific Department, Virology Laboratory | Army Medical Center, Scientific Department, Virology Laboratory | Silvia Fillo et al |
| EPI_ISL_539812; EPI_ISL_539811; EPI_ISL_539821 | Asiaworld Expo Command Post | Hong Kong Department of Health | Alan K.L. Tsang et al |
| EPI_ISL_413490 | Auckland Hospital | Institute of Environmental Science and Research (ESR) | Matt Storey et al A (https://dx.doi.org/10.1101/2020.03.15.992818) |
| EPI_ISL_583885; EPI_ISL_475836; EPI_ISL_583890; EPI_ISL_583889; EPI_ISL_583891; EPI_ISL_583888; EPI_ISL_583887; EPI_ISL_583892; EPI_ISL_475842; EPI_ISL_583674; EPI_ISL_583886; EPI_ISL_475849; EPI_ISL_583666; EPI_ISL_475835; EPI_ISL_583644; EPI_ISL_583683; EPI_ISL_583630; EPI_ISL_583636; EPI_ISL_583645; EPI_ISL_583675; EPI_ISL_583682; EPI_ISL_583883; EPI_ISL_583884; EPI_ISL_583662 | Austrian Agency for Health and Food Safety (AGES) | Bergthaler laboratory, CeMM Research Center for Molecular Medicine of the Austrian Academy of Sciences | Alexandra Popa et al A |
| EPI_ISL_728266; EPI_ISL_728267 | B.J. Govt. Medical College | National Centre For Cell Science | Dhiraj Paul et al |
| EPI_ISL_444481 | B.J. Medical College and Civil hospital | Gujarat Biotechnology Research Centre | Kairavi Joshi et al (https://www.biorxiv.org/content/10.1101/2020.05.28.122143v1) |
| EPI_ISL_461498 | B.J. Medical College and Civil hospital | Gujarat Biotechnology Research Centre | Pooja P Doshi et al |
| EPI_ISL_586518 | B.J. Medical College and Civil hospital, Ahmedabad | Gujarat Biotechnology Research Centre | Dinesh Kumar et al |
| EPI_ISL_747244 | BBMP Urban PHC | Department of Neurovirology, National Institute of Mental Health and Neurosciences (NIMHANS) | Chitra Pattabiraman et al |
| EPI_ISL_415580; EPI_ISL_415578; EPI_ISL_415577 | BCCDC Public Health Laboratory | BCCDC Public Health Laboratory | Harrigan et al A |
| EPI_ISL_412965 | BCCDC Public Health Laboratory | BCCDC Public Health Laboratory | Harrigan et al B (https://dx.doi.org/10.1002/jmv.25773) |
| EPI_ISL_574619 | BTKLPP Kelas I Manado | Eijkman Institute for Molecular Biology, Ministry of Research and Technology/National Agency for Research and Innovation | Hidayat Trimarsanto et al |
| EPI_ISL_538506; EPI_ISL_538507 | Balai Penelitian dan Pengembangan Biomedis Papua | National Institute of Health Research and Development | Pawestri et al |

| | | | |
|---|---|---|---|
| EPI_ISL_403962; EPI_ISL_403963 | Bamrasnaradura Hospital | 1. Department of Medical Sciences, Ministry of Public Health, Thailand 2. Thai Red Cross Emerging Infectious Diseases - Health Science Centre 3. Department of Disease Control, Ministry of Public Health, Thailand | Pilailuk et al B (https://dx.doi.org/10.2807/1560-7917.ES.2020.25.8.2000097) |
| EPI_ISL_434692; EPI_ISL_434693; EPI_ISL_434694 | Bamrasnaradura hospital | National Institute of Health. Department of medical Sciences, Ministry of Public Health, Thailand | Pilailuk et al A |
| EPI_ISL_450341; EPI_ISL_450345 | Bangladesh Institute of Tropical & Infectious Diseases, COVID-19 Testing Laboratory | Basic and Applied Research on Jute Project | Md. Sabbir Hossain et al (https://dx.doi.org/10.1101/2020.06.07.138800) |
| EPI_ISL_450342; EPI_ISL_450339; EPI_ISL_450340; EPI_ISL_450344 | Bangladesh Institute of Tropical & Infectious Diseases, COVID-19 Testing Laboratory | Basic and Applied Research on Jute Project | Rasel Ahmed et al (https://dx.doi.org/10.1101/2020.06.07.138800) |
| EPI_ISL_710613; EPI_ISL_732657 | Barnakuten | The Public Health Agency of Sweden | Department of Microbiology et al |
| EPI_ISL_529217; EPI_ISL_529216; EPI_ISL_529214 | Beijing Institute of Microbiology and Epidemiology | Beijing Institute of Microbiology and Epidemiology | Fan et al |
| EPI_ISL_509711; EPI_ISL_509712; EPI_ISL_509714; EPI_ISL_509713 | Belize Ministry of Health | Pathogen Discovery, Respiratory Viruses Branch, Division of Viral Diseases, Centers for Disease Control and Prevention | Jing Zhang et al B |
| EPI_ISL_430843 | Bethany Hospital | Research Institute for Tropical Medicine | Medado et al |
| EPI_ISL_735495 | Bhashabir M A Wadud RT-PCR Lab, Chandpur | Central Biological Research Laboratory and Department of Biochemistry and Molecular Biology | Md. Imranul Hoq et al |
| EPI_ISL_429998; EPI_ISL_450188; EPI_ISL_635777; EPI_ISL_430008; EPI_ISL_430015; EPI_ISL_450189; EPI_ISL_450186; EPI_ISL_430014; EPI_ISL_429997; EPI_ISL_430011; EPI_ISL_429995; EPI_ISL_430000; EPI_ISL_434516; EPI_ISL_635778; EPI_ISL_635782; EPI_ISL_450187; EPI_ISL_635781; EPI_ISL_730520; EPI_ISL_730549; EPI_ISL_730287; EPI_ISL_730544; EPI_ISL_730550; EPI_ISL_730431; EPI_ISL_730266; EPI_ISL_730524; EPI_ISL_730243; EPI_ISL_730265; EPI_ISL_730385; EPI_ISL_730234; EPI_ISL_730556; EPI_ISL_730559; EPI_ISL_730434; EPI_ISL_730533; EPI_ISL_730529; EPI_ISL_730507; EPI_ISL_730506; EPI_ISL_755255; EPI_ISL_755233; EPI_ISL_755232; EPI_ISL_755234; EPI_ISL_755240; EPI_ISL_755252; EPI_ISL_755141; EPI_ISL_755268; EPI_ISL_755126; EPI_ISL_755254; EPI_ISL_755256; EPI_ISL_755129; EPI_ISL_755258; EPI_ISL_755265; EPI_ISL_755127; EPI_ISL_755266; EPI_ISL_755267; EPI_ISL_730558; EPI_ISL_430003; EPI_ISL_430013; EPI_ISL_430009; EPI_ISL_755257; EPI_ISL_755263; EPI_ISL_635780; EPI_ISL_635779; EPI_ISL_755145 | Biolab Diagnostic Laboratories | Andersen lab at Scripps Research | Issa Abu-Dayyeh et al |
| EPI_ISL_527007 | Biological Prevention, Army | Biological Prevention, Army | Seadawy et al |
| EPI_ISL_510526; EPI_ISL_510532; EPI_ISL_526975; EPI_ISL_526984; EPI_ISL_526998; EPI_ISL_526981; EPI_ISL_526986; EPI_ISL_526990; EPI_ISL_527003; EPI_ISL_527000; EPI_ISL_527002 | Biological prevention, army | Biological prevention, army | Seadawy et al |
| EPI_ISL_582030 | Biology Department, College of Science, Al-Muthanna University | International Centre for Genetic Engineering and Biotechnology (ICGEB) and ARGO Open Lab Platform | Nihad Al-Rashedi et al |
| EPI_ISL_516088; EPI_ISL_516087; EPI_ISL_516084; EPI_ISL_516085; EPI_ISL_516083 | Biomedical Sciences and Public Health, Polytechnic University of Marche | Biomedical Sciences and Public Health, Polytechnic University of Marche | Bagnarelli et al |
| EPI_ISL_747238 | Bogor Public Health | West Java Health Laboratory; School of Life Sciences and Technology, Institut Teknologi Bandung | Azzania Fibriani et al |
| EPI_ISL_596453 | Booali laboratory, Qom, Iran. Department of Virology, School of Public Health, Tehran University of Medical Sciences, Tehran, Iran. | Genetics Research Center, University of Social Welfare and Rehabilitation Sciences | Zohreh Fattahi et al |
| EPI_ISL_718167; EPI_ISL_718176 | Borneo Medical Centre | Institute of Health and Community Medicine | David Perera et al |
| EPI_ISL_765605; EPI_ISL_765589 | Brigham and Womens Hospital | Infectious Disease Program, Broad Institute of Harvard and MIT | Lemieux et al |
| EPI_ISL_732560; EPI_ISL_732557; EPI_ISL_732556; EPI_ISL_732533; EPI_ISL_732558; EPI_ISL_732559; EPI_ISL_732539; EPI_ISL_732538; EPI_ISL_732540; EPI_ISL_732537; EPI_ISL_732535; EPI_ISL_732536; EPI_ISL_732534; EPI_ISL_732658; EPI_ISL_732531; EPI_ISL_732532 | Bundeswehr Institute of Microbiology | Bundeswehr Institute of Microbiology | Elham Khatamzas et al |
| EPI_ISL_755639 | Bundeswehr Institute of Microbiology | Bundeswehr Institute of Microbiology | Malena Bestehorn-Willmann et al |
| EPI_ISL_732527; EPI_ISL_732530; EPI_ISL_732564; EPI_ISL_732542; EPI_ISL_732526; EPI_ISL_732543; EPI_ISL_732525; EPI_ISL_732524; EPI_ISL_732546; EPI_ISL_732541; EPI_ISL_732528; EPI_ISL_732544; EPI_ISL_732555; EPI_ISL_732563; EPI_ISL_732545; EPI_ISL_732548; EPI_ISL_732561; EPI_ISL_732562; EPI_ISL_732552; EPI_ISL_732549; EPI_ISL_732550; EPI_ISL_732547; EPI_ISL_732553; EPI_ISL_732554; EPI_ISL_732551 | Bundeswehr Institute of Microbiology | Bundeswehr Institute of Microbiology | Markus Antwerpen et al |
| EPI_ISL_414521; EPI_ISL_414520 | Bundeswehr Institute of Microbiology | Bundeswehr Institute of Microbiology | Mathias C Walter et al |
| EPI_ISL_605780 | CEIRS Data Processing and Coordinating Center, St. Jude Center of Excellence for Influenza Research and Surveillance (CEIRS) | CEIRS Data Processing and Coordinating Center, St. Jude Center of Excellence for Influenza Research and Surveillance (CEIRS) | Roshdy et al |
| EPI_ISL_750175 | CENUR Este-Sede Rocha-UdelaR | Institut Pasteur de Montevideo | Daiana Mir et al |
| EPI_ISL_751187; EPI_ISL_751188; EPI_ISL_751184 | CENUR Litoral Norte – UdelaR, Salto, Uruguay | Institut Pasteur de Montevideo | Daiana Mir et al |
| EPI_ISL_644253; EPI_ISL_644252; EPI_ISL_644251; EPI_ISL_644258 | CEPHR / Mater Hospital | Irish Coronavirus Sequencing Consortium - National Virus Reference Laboratory | Michael Carr et al |
| EPI_ISL_644284; EPI_ISL_644341; EPI_ISL_644279; EPI_ISL_644338; EPI_ISL_644224; EPI_ISL_644295; EPI_ISL_644307; EPI_ISL_644267; EPI_ISL_644308; EPI_ISL_644236; EPI_ISL_644211; EPI_ISL_644285; EPI_ISL_644301; EPI_ISL_644302 | CEPHR / Vincent's Hospital | Irish Coronavirus Sequencing Consortium - National Virus Reference Laboratory | Michael Carr et al |
| EPI_ISL_733569 | CHAI WAN FAMILIES CLINIC | Hong Kong Department of Health | Alan K.L. Tsang et al |
| EPI_ISL_613558; EPI_ISL_613556; EPI_ISL_613557 | CHRU Pontchaillou - Laboratoire de Virologie 2, rue Henri Le Guilloux | National Reference Center for Viruses of Respiratory Infections, Institut Pasteur, Paris | Marion Barbet et al B |
| EPI_ISL_418219 | CHU - Hôpital Cavale Blanche - Labo. de Virologie | National Reference Center for Viruses of Respiratory Infections, Institut Pasteur, Paris | Mélanie Albert et al A (https://dx.doi.org/10.1101/2020.04.24.059576) |
| EPI_ISL_443265 | CHU - Hôpital Cavale Blanche - Labo. de Virologie | National Reference Center for Viruses of Respiratory Infections, Institut Pasteur, Paris | Mélanie Albert et al C |
| EPI_ISL_660359; EPI_ISL_745314; EPI_ISL_645188 | CHU Clermont-Ferrand | CNR Virus des Infections Respiratoires - France SUD | Antonin Bal et al |
| EPI_ISL_644681; EPI_ISL_660684; EPI_ISL_660698; EPI_ISL_660686 | CHU Montpellier | CNR Virus des Infections Respiratoires - France SUD | Antonin Bal et al |
| EPI_ISL_663270 | CHU Nantes | CNR Virus des Infections Respiratoires - France SUD | Antonin Bal et al |
| EPI_ISL_663235; EPI_ISL_645216 | CHU Poitiers | CNR Virus des Infections Respiratoires - France SUD | Antonin Bal et al |
| EPI_ISL_751454; EPI_ISL_593873; EPI_ISL_593874; EPI_ISL_671943; EPI_ISL_593867; EPI_ISL_593869; EPI_ISL_593868; EPI_ISL_593871; EPI_ISL_593872; EPI_ISL_593875; EPI_ISL_593876; EPI_ISL_593878; EPI_ISL_603217; EPI_ISL_603216; EPI_ISL_671942; EPI_ISL_751495; EPI_ISL_671945; EPI_ISL_751464; EPI_ISL_754138; EPI_ISL_671967; EPI_ISL_751480; EPI_ISL_754137; EPI_ISL_751473; EPI_ISL_751467; EPI_ISL_751475; EPI_ISL_751470; EPI_ISL_593895; EPI_ISL_751490; EPI_ISL_593879; EPI_ISL_671975; EPI_ISL_751452; EPI_ISL_751498; EPI_ISL_751451; EPI_ISL_593855; EPI_ISL_591542; EPI_ISL_591099; EPI_ISL_591545; EPI_ISL_591547; EPI_ISL_591543; EPI_ISL_591546; EPI_ISL_593857; EPI_ISL_593856; EPI_ISL_593862; EPI_ISL_593864; EPI_ISL_593860; EPI_ISL_593861; EPI_ISL_593891; EPI_ISL_593863; EPI_ISL_593858; EPI_ISL_593859 | CHU Purpan - Laboratoire de Virologie - Institut Fédératif de Biologie | CHU Purpan - Laboratoire de Virologie - Institut Fédératif de Biologie | Latour J. et al |
| EPI_ISL_482879; EPI_ISL_482883; EPI_ISL_482882 | CHU Purpan - Laboratoire de Virologie - Institut Fédératif de Biologie | Laboratoire de virologie - École Nationale Vétérinaire de Toulouse | Guillaume Croville et al |
| EPI_ISL_660670 | CHU Toulouse | CNR Virus des Infections Respiratoires - France SUD | Antonin Bal et al |
| EPI_ISL_735391 | CHU Tours | CNR Virus des Infections Respiratoires - France SUD | Antonin Bal et al |
| EPI_ISL_666713 | CHU de Limoges | CNR Virus des Infections Respiratoires - France SUD | Antonin Bal et al |
| EPI_ISL_641534 | CHU de Nice - Hôpital Archet 16 | CNR Virus des Infections Respiratoires - France SUD | Antonin Bal et al |
| EPI_ISL_693387; EPI_ISL_693388 | CHU de Nice - Hôpital Archet 2 | CNR Virus des Infections Respiratoires - France SUD | Antonin Bal et al |
| EPI_ISL_418013; EPI_ISL_417995 | CHULC - H Curry Cabral | Instituto Nacional de Saude (INSA) | Guiomar et al et al |
| EPI_ISL_417992 | CHULC - H D Estefania | Instituto Nacional de Saude (INSA) | Guiomar et al et al |
| EPI_ISL_745260 | CHUM-Site Glen-LAB Microbiologie | Laboratoire de santé publique du Québec | Sandrine Moreira et al |
| EPI_ISL_683835 | CICM | Malaria Research and Training Center (MRTC-Parasito) | Antoine Dara et al |
| EPI_ISL_487463; EPI_ISL_487451; EPI_ISL_487448; EPI_ISL_487460; EPI_ISL_487465; EPI_ISL_487456; EPI_ISL_487457; EPI_ISL_487452; EPI_ISL_487447; EPI_ISL_487450 | CICM-Mali | Bundeswehr Institut of Microbiology | Kouriba et al |
| EPI_ISL_445282 | CLINICA MAGALLANES S.A. | Instituto de Salud Publica de Chile | Andrés E Castillo et al |
| EPI_ISL_445315 | CLINICA UC SAN CARLOS DE APOQUINDO | Instituto de Salud Publica de Chile | Andrés E Castillo et al |
| EPI_ISL_745380; EPI_ISL_639977; EPI_ISL_639997; EPI_ISL_683378; EPI_ISL_623100; EPI_ISL_582111; EPI_ISL_639987; EPI_ISL_582112; EPI_ISL_683382; EPI_ISL_683394; EPI_ISL_692764; EPI_ISL_692737; EPI_ISL_692738; EPI_ISL_692762; EPI_ISL_683388; EPI_ISL_745363; EPI_ISL_640014; EPI_ISL_745383; EPI_ISL_745317; EPI_ISL_728539; EPI_ISL_728542; EPI_ISL_745379; EPI_ISL_745378; EPI_ISL_745337; EPI_ISL_683336; EPI_ISL_732679; EPI_ISL_745373; EPI_ISL_728316; EPI_ISL_745323; EPI_ISL_683398; EPI_ISL_683373; EPI_ISL_683380; EPI_ISL_693493; EPI_ISL_639989; EPI_ISL_728534; EPI_ISL_745385; EPI_ISL_745392; EPI_ISL_728310; EPI_ISL_745307; EPI_ISL_508933; EPI_ISL_508939; EPI_ISL_676558; EPI_ISL_683353; EPI_ISL_508981; EPI_ISL_508930; EPI_ISL_508922; EPI_ISL_692736; EPI_ISL_525541; EPI_ISL_683360; EPI_ISL_683354; EPI_ISL_683361; EPI_ISL_678539; EPI_ISL_678546; EPI_ISL_683356; EPI_ISL_678544; EPI_ISL_678547; EPI_ISL_683359; EPI_ISL_678543; EPI_ISL_683358; EPI_ISL_683352; EPI_ISL_683374; EPI_ISL_745324; EPI_ISL_745325; EPI_ISL_678496; EPI_ISL_508969; EPI_ISL_508993; EPI_ISL_676563; EPI_ISL_681263 | CNR Virus des Infections Respiratoires - France SUD | CNR Virus des Infections Respiratoires - France SUD | Antonin Bal et al |
| EPI_ISL_410486 | CNR Virus des Infections Respiratoires - France SUD | CNR Virus des Infections Respiratoires - France SUD | Bal et al (https://dx.doi.org/10.1016/j.cmi.2020.03.020) |
| EPI_ISL_636980 | CS Xai Xai | KRISP, KZN Research Innovation and Sequencing Platform | Ismael N et al |
| EPI_ISL_495226 | CSIR-Centre for Cellular and Molecular Biology | CSIR-Centre for Cellular and Molecular Biology | M Soujanya Reddy et al |
| EPI_ISL_539659 | CSIR-Centre for Cellular and Molecular Biology | CSIR-Centre for Cellular and Molecular Biology | Payel Mukherjee et al |
| EPI_ISL_458080 | CSIR-Centre for Cellular and Molecular Biology | CSIR-Centre for Cellular and Molecular Biology | Sakshi Shambhavi et al |
| EPI_ISL_539668 | CSIR-Centre for Cellular and Molecular Biology | CSIR-Centre for Cellular and Molecular Biology | Shagufta Khan et al |

| | | | |
|---|---|---|---|
| EPI_ISL_661307; EPI_ISL_581497; EPI_ISL_578175; EPI_ISL_578168; EPI_ISL_661305; EPI_ISL_661308; EPI_ISL_661311; EPI_ISL_661310; EPI_ISL_661309; EPI_ISL_661304; EPI_ISL_581494; EPI_ISL_569861; EPI_ISL_581451 | CSIR-Indian Institute of Chemical Biology, MEDICA Supercpecialty Hospital Kolkata | CSIR-Indian Institute of Chemical Biology, MEDICA Supercpecialty Hospital Kolkata | Sujay Krishna Maity et al |
| EPI_ISL_445303 | CTRO.DE SALUD FAMILIAR DR. RAUL YAZIGI | Instituto de Salud Publica de Chile | Andrés E Castillo et al |
| EPI_ISL_452140 | CUB Hopital Erasme Laboratoire d'Anatomie Pathologique | CUB Hopital Erasme Laboratoire d'Anatomie Pathologique | Isabelle Salmon et al |
| EPI_ISL_451935 | CUB Hopital Erasme Laboratoire d'Anatomie Pathologique | CUB Hopital Erasme Laboratoire d'Anatomie Pathologique | Prof. Isabelle Salmon et al |
| EPI_ISL_756370 | CUMC - CHI Bergan Mercy | Creighton University School of Medicine, Departments of Medical Microbiology and Pharmacology and Neuroscience | Michael Belshan et al |
| EPI_ISL_582477; EPI_ISL_582376; EPI_ISL_582391; EPI_ISL_582397; EPI_ISL_582374; EPI_ISL_582476; EPI_ISL_582468; EPI_ISL_582368; EPI_ISL_582375; EPI_ISL_582390; EPI_ISL_582503; EPI_ISL_582462; EPI_ISL_582422; EPI_ISL_582488; EPI_ISL_582362; EPI_ISL_582407; EPI_ISL_582312 | Cadham Provincial Laboratory | National Microbiology Laboratory (NML) | Anna Majer et al |
| EPI_ISL_408008; EPI_ISL_408009 | California Department of Health | Pathogen Discovery, Respiratory Viruses Branch, Division of Viral Diseases, Centers for Disease Control and Prevention | Krista Queen et al B (https://dx.doi.org/10.1101/2020.03.09.20032896) |
| EPI_ISL_408010 | California Department of Health | Pathogen Discovery, Respiratory Viruses Branch, Division of Viral Diseases, Centers for Disease Control and Prevention | Ying Tao et al B (https://dx.doi.org/10.1101/2020.03.09.20032896) |
| EPI_ISL_406036 | California Department of Public Health | Pathogen Discovery, Respiratory Viruses Branch, Division of Viral Diseases, Centers for Disease Control and Prevention | Anna Uehara et al (https://dx.doi.org/10.1101/2020.03.09.20032896) |
| EPI_ISL_410044 | California Department of Public Health | Pathogen Discovery, Respiratory Viruses Branch, Division of Viral Diseases, Centers for Disease Control and Prevention | Jing Zhang et al A (https://dx.doi.org/10.1101/2020.03.09.20032896) |
| EPI_ISL_411954; EPI_ISL_412862; EPI_ISL_411955 | California Department of Public Health | Pathogen Discovery, Respiratory Viruses Branch, Division of Viral Diseases, Centers for Disease Control and Prevention | Krista Queen et al A |
| EPI_ISL_419554 | California Department of Public Health | Pathogen Discovery, Respiratory Viruses Branch, Division of Viral Diseases, Centers for Disease Control and Prevention | Ying Tao et al A |
| EPI_ISL_447594 | Caloundra Hospital | Public Health Virology Laboratory | Bixing Huang et al |
| EPI_ISL_576373; EPI_ISL_576371; EPI_ISL_576372 | Cancer Biology Department, National Cancer Institute | Cancer Biology Department, National Cancer Institute | Zekri et al |
| EPI_ISL_447054 | Cantacuzino National Military-Medical Institute for Research and Development | Cantacuzino Institute | M.Lazar et al (https://dx.doi.org/10.1101/2020.06.28.175802) |
| EPI_ISL_456373; EPI_ISL_456360; EPI_ISL_456353 | Canterbury Health Laboratories | Institute of Environmental Science and Research (ESR) | Matt Storey et al B |
| EPI_ISL_579224; EPI_ISL_548138; EPI_ISL_548139; EPI_ISL_548140; EPI_ISL_755620; EPI_ISL_755622; EPI_ISL_755621; EPI_ISL_732963; EPI_ISL_579426; EPI_ISL_579496; EPI_ISL_536792 | Canterbury Health Laboratories | Institute of Environmental Science and Research (ESR) | Xiaoyun Ren et al |
| EPI_ISL_756311 | Caribbean Regional Health Authority | Carrington Lab, Department of PreClinical Sciences, Faculty of Medical Sciences, The University of the West Indies | Nikita S. D. Sahadeo et al |
| EPI_ISL_756357; EPI_ISL_756367 | Caribbean Regional Public Health Authority | Carrington Lab, Department of PreClinical Sciences, Faculty of Medical Sciences, The University of the West Indies | Nikita S. D. Sahadeo et al |
| EPI_ISL_450413 | Center for Diagnostics, Institute of Medical Microbiology, Virology and Hygiene | University Medical Center Hamburg-Eppendorf | Huang et al (https://dx.doi.org/10.1128/MRA.00520-20) |
| EPI_ISL_468760 | Center for Genome Regulation (CRG) | Center for Mathematical Modeling and Center for Genome Regulation. Santiago, Chile | Gaete A et al |
| EPI_ISL_516784; EPI_ISL_516785; EPI_ISL_522441; EPI_ISL_522440; EPI_ISL_522445; EPI_ISL_522444; EPI_ISL_522450; EPI_ISL_522464; EPI_ISL_522460; EPI_ISL_522443; EPI_ISL_522447 | Center for Laboratory Control of Infectious Diseases, Korea Centers for Diseases Control and Prevention | Center for Laboratory Control of Infectious Diseases, Korea Centers for Diseases Control and Prevention | Junyoung Kim et al |
| EPI_ISL_493137 | Center for Research and Innovation, Faculty of Medical Technology, Mahidol University | Center for Research and Innovation, Faculty of Medical Technology, Mahidol University | Kantima Sangsiriwut et al |
| EPI_ISL_583563; EPI_ISL_583708; EPI_ISL_583712; EPI_ISL_583713; EPI_ISL_583710; EPI_ISL_583711; EPI_ISL_583709; EPI_ISL_583715; EPI_ISL_583693; EPI_ISL_583692; EPI_ISL_583695; EPI_ISL_583723 | Center for Virology, Medical University of Vienna | Bergthaler laboratory, CeMM Research Center for Molecular Medicine of the Austrian Academy of Sciences | Alexandra Popa et al A |
| EPI_ISL_475795 | Center for Virology, Medical University of Vienna | Bergthaler laboratory, CeMM Research Center for Molecular Medicine of the Austrian Academy of Sciences | Alexandra Popa et al B (https://dx.doi.org/10.1101/2020.07.15.204339 v1) |
| EPI_ISL_419655; EPI_ISL_437993; EPI_ISL_437994; EPI_ISL_419656; EPI_ISL_437995; EPI_ISL_438077; EPI_ISL_438045; EPI_ISL_438046; EPI_ISL_438082; EPI_ISL_438086 | Center for Virology, Medical University of Vienna | Bergthaler laboratory, CeMM Research Center for Molecular Medicine of the Austrian Academy of Sciences | Alexandra Popa et al B (https://www.biorxiv.org/content/10.1101/2020.07.15.204339v1) |
| EPI_ISL_677708; EPI_ISL_516418; EPI_ISL_677706; EPI_ISL_516421; EPI_ISL_516415; EPI_ISL_516414; EPI_ISL_516416; EPI_ISL_678249; EPI_ISL_678248; EPI_ISL_516419 | Center for public health - Skopje | Research Center for Genetic Engineering and Biotechnology "Georgi D. Efremov" , Macedonian Academy of Sciences and Arts | RCGEB - MASA et al |
| EPI_ISL_755572 | Center of Advanced Studies and Technology, CAST | Center of Advanced Studies and Technology, CAST | Ferrante et al |
| EPI_ISL_523950 | Center of Medical Microbiology, Virology, and Hospital Hygiene, University of Duesseldorf | Center of Medical Microbiology, Virology, and Hospital Hygiene, Heinrich Heine University Düsseldorf | Maximilian Damagnez et al |
| EPI_ISL_602547; EPI_ISL_602544; EPI_ISL_523941; EPI_ISL_523931; EPI_ISL_602517; EPI_ISL_523929; EPI_ISL_602552; EPI_ISL_523937; EPI_ISL_602542; EPI_ISL_523936; EPI_ISL_523943; EPI_ISL_602510; EPI_ISL_602513; EPI_ISL_602533; EPI_ISL_523938; EPI_ISL_602534; EPI_ISL_523946; EPI_ISL_523944; EPI_ISL_602512; EPI_ISL_602515; EPI_ISL_602539; EPI_ISL_602516; EPI_ISL_523933; EPI_ISL_523928; EPI_ISL_602550; EPI_ISL_523939; EPI_ISL_602546; EPI_ISL_602511; EPI_ISL_602538; EPI_ISL_523927; EPI_ISL_523932; EPI_ISL_602537; EPI_ISL_523940; EPI_ISL_523935; EPI_ISL_602535; EPI_ISL_523934; EPI_ISL_523947; EPI_ISL_602541; EPI_ISL_602536; EPI_ISL_602551; EPI_ISL_523942; EPI_ISL_602543; EPI_ISL_602549; EPI_ISL_602514; EPI_ISL_602548; EPI_ISL_523945; EPI_ISL_602545; EPI_ISL_602540; EPI_ISL_523948; EPI_ISL_523949 | Center of Medical Microbiology, Virology, and Hospital Hygiene, University of Duesseldorf | Center of Medical Microbiology, Virology, and Hospital Hygiene, University of Duesseldorf | Maximilian Damagnez et al |
| EPI_ISL_425123; EPI_ISL_452180; EPI_ISL_417460; EPI_ISL_414509; EPI_ISL_414574; EPI_ISL_425134; EPI_ISL_414506; EPI_ISL_414499; EPI_ISL_414508; EPI_ISL_413488; EPI_ISL_414497; EPI_ISL_414498; EPI_ISL_414505; EPI_ISL_414504; EPI_ISL_417459; EPI_ISL_417462; EPI_ISL_417461; EPI_ISL_425130; EPI_ISL_419552; EPI_ISL_419548; EPI_ISL_425132; EPI_ISL_425129; EPI_ISL_417458; EPI_ISL_425125; EPI_ISL_417464; EPI_ISL_419542; EPI_ISL_425136; EPI_ISL_425122; EPI_ISL_425138; EPI_ISL_417457; EPI_ISL_419549; EPI_ISL_425133; EPI_ISL_419543; EPI_ISL_417467; EPI_ISL_419545; EPI_ISL_419546; EPI_ISL_425124; EPI_ISL_425131; EPI_ISL_417466; EPI_ISL_417465; EPI_ISL_419541; EPI_ISL_419544; EPI_ISL_425137; EPI_ISL_417468; EPI_ISL_425121; EPI_ISL_417463; EPI_ISL_425126; EPI_ISL_419550; EPI_ISL_419551; EPI_ISL_425120; EPI_ISL_425139; EPI_ISL_425135; EPI_ISL_419547; EPI_ISL_425140 | Center of Medical Microbiology, Virology, and Hospital Hygiene, University of Duesseldorf | Center of Medical Microbiology, Virology, and Hospital Hygiene, University of Duesseldorf | Ortwin Adams et al A (https://dx.doi.org/10.1101/2020.04.25.20079517) |
| EPI_ISL_414507; EPI_ISL_425127; EPI_ISL_425128 | Center of Medical Microbiology, Virology, and Hospital Hygiene, University of Duesseldorf | Center of Medical Microbiology, Virology, and Hospital Hygiene, University of Duesseldorf | Ortwin Adams et al B |
| EPI_ISL_523930 | Center of Medical Microbiology, Virology, and Hospital Hygiene, University of Duesseldorf | Universitätstr.1 40225 Düsseldorf Germany | Maximilian Damagnez et al |
| EPI_ISL_430820 | Center of Scientific Excellence for Influenza Viruses, National Research Centre (NRC), Egypt. | Center of Scientific Excellence for Influenza Viruses, National Research Centre (NRC), Egypt. | Mohamed Ahmed Ali et al (https://dx.doi.org/10.3389/fmicb.2020.01316) |
| EPI_ISL_430819 | Center of Scientific Excellence for Influenza Viruses,National Research Centre (NRC), Egypt. | Center of Scientific Excellence for Influenza Viruses,National Research Centre (NRC), Egypt. | Mohamed Ahmed Ali et al (https://dx.doi.org/10.3389/fmicb.2020.01316) |
| EPI_ISL_428488; EPI_ISL_428489; EPI_ISL_428490 | Centers for Disease Control, R.O.C. (Taiwan) | Centers for Disease Control, R.O.C. (Taiwan) | Ji-Rong Yang et al B |
| EPI_ISL_406031 | Centers for Disease Control, R.O.C. (Taiwan) | Centers for Disease Control, R.O.C. (Taiwan) | Ji-Rong Yang et al C (https://dx.doi.org/10.1099/acmi.0.000133) |
| EPI_ISL_459964; EPI_ISL_459963; EPI_ISL_459962; EPI_ISL_457750 | Centogene AG | Centogene AG | Prof. Dr. Peter Bauer et al |
| EPI_ISL_529032 | Central Molecular Microbiology Laboratory and Next Generation Sequencing Reference Laboratory, Clinical and Chemical Pathology Department, Faculty of Medicine, CAIRO UNIVERSITY | Next Generation Sequencing Reference Laboratory, Faculty of Medicine, CAIRO UNIVERSITY and The Center for Genome and Microbiome Research, Faculty of Pharmacy, CAIRO UNIVERSITY | May Sherif Soliman et al |
| EPI_ISL_693478; EPI_ISL_693475; EPI_ISL_693473; EPI_ISL_693474; EPI_ISL_693472; EPI_ISL_693471; EPI_ISL_693479; EPI_ISL_693480; EPI_ISL_693483; EPI_ISL_693477; EPI_ISL_693476; EPI_ISL_693481; EPI_ISL_693482 | Central Public Health Laboratory | National Public Health Laboratory, National Centre for Infectious Diseases | Tze Minn Mak et al |
| EPI_ISL_419211 | Central Virology Laboratory | Israel Institute for Biological Research | Inbar Cohen-Gihon et al (https://dx.doi.org/10.1101/2020.05.21.20104521) |
| EPI_ISL_737203; EPI_ISL_733498 | Central Virology Laboratory, Israel Ministry of Health | Central Virology Laboratory, Israel Ministry of Health | Neta S. Zuckerman et al |
| EPI_ISL_486417; EPI_ISL_486410 | Centrala laboratorija | Latvian Biomedical Research and Study Centre | Ivars Silamiķelis et al |
| EPI_ISL_548256 | Centralsjukhuset | The Public Health Agency of Sweden | Anna-Malin Linde et al |
| EPI_ISL_458000 | Centre For Biotechnology Research and Development | Centre For Biotechnology Research and Development | Matoke-Muhia et al |
| EPI_ISL_416494; EPI_ISL_414624 | Centre Hositalier Universitaire de Rouen Laboratoire de Virologie | National Reference Center for Viruses of Respiratory Infections, Institut Pasteur, Paris | Mélnie Albert et al (https://dx.doi.org/10.1101/2020.04.24.059576) |
| EPI_ISL_429968; EPI_ISL_418220 | Centre Hospitalier Compiègne Laboratoire de Biologie | National Reference Center for Viruses of Respiratory Infections, Institut Pasteur, Paris | Mélanie Albert et al A (https://dx.doi.org/10.1101/2020.04.24.059576) |
| EPI_ISL_418218 | Centre Hospitalier Compiègne Laboratoire de Biologie | National Reference Center for Viruses of Respiratory Infections, Institut Pasteur, Paris | Mélanie Albert et al C |
| EPI_ISL_636478; EPI_ISL_636480; EPI_ISL_636483; EPI_ISL_636482; EPI_ISL_636486 | Centre Hospitalier Pierre Oudot | CNR Virus des Infections Respiratoires - France SUD | Antonin Bal et al |
| EPI_ISL_419168 | Centre Hospitalier de Valence | CNR Virus des Infections Respiratoires - France SUD | Antonin Bal et al |
| EPI_ISL_660442 | Centre Muraz | Project group Epidemology of Highly Pathogenic Microorganisms, Robert Koch Institut | Soumeya Ouangraoua et al |
| EPI_ISL_539575; EPI_ISL_539573; EPI_ISL_539576; EPI_ISL_539574 | Centre de Recherches Medicales de Lambarene (CERMEL) | Department of Emerging Infectious Diseases, Institute of Tropical Medicine, Nagasaki University | Haruka Abe et al |
| EPI_ISL_471459; EPI_ISL_471457; EPI_ISL_471458; EPI_ISL_471460 | Centre de Virologie des Maladies Tropicales | Functional Genomic Platform/Service Analyses Biologique/UATRS/ Centre National Pour la Recherche Scientifique Et Technique (CNRST) | Hicham ANNAZ et al |
| EPI_ISL_428673; EPI_ISL_428670; EPI_ISL_428671; EPI_ISL_428672 | Centre for Dengue Research | Centre for Dengue Research | Chandima Jeewandara et al A (https://dx.doi.org/10.21203/rs.3.rs-44134/v1) |
| EPI_ISL_525481; EPI_ISL_525479; EPI_ISL_525478; EPI_ISL_525488; EPI_ISL_525486; EPI_ISL_525489; EPI_ISL_525476 | Centre for Dengue Research | Centre for Dengue Research | Chandima Jeewandara et al B |
| EPI_ISL_525474 | Centre for Dengue Research | Centre for Dengue Research, USJ, SL | Chandima Jeewandara et al B |

| | | | |
|---|---|---|---|
| EPI_ISL_602564 | Centre for Dengue Research, Department of Immunology and Molecular Medicine | Centre for Dengue Research | Chandima Jeewandara et al B |
| EPI_ISL_668449; EPI_ISL_668451; EPI_ISL_668453; EPI_ISL_602576; EPI_ISL_602566; EPI_ISL_602574; EPI_ISL_668450; EPI_ISL_668454; EPI_ISL_668447; EPI_ISL_668452; EPI_ISL_668448; EPI_ISL_668455 | Centre for Dengue Research, Department of Immunology and Molecular Medicine | Centre for Dengue Research, Department of Immunology and Molecular Medicine | Chandima Jeewandara et al B |
| EPI_ISL_461996 | Centre for Enzyme Innovation, University of Portsmouth / Translational Research Laboratory, Portsmouth Hospitals NHS Trust | COVID-19 Genomics UK (COG-UK) Consortium | Angela Beckett et al |
| EPI_ISL_733568 | Centre for Health Protection | Hong Kong Department of Health | Alan K.L. Tsang et al |
| EPI_ISL_413550 | Centre for Human and Zoonotic Virology (CHAZVY), College of Medicine University of Lagos/Lagos University Teaching Hospital (LUTH), part of the Laboratory Network of the Nigeria Centre for Disease Control (NCDC) | African Centre of Excellence for Genomics of Infectious Diseases (ACEGID), Redeemer's University, Ede, Osun State, Nigeria | Oluniyi P.E. et al C (https://virological.org/t/first-african-sars-cov-2-genome-sequence-from-nigerian-covid-19-case/421) |
| EPI_ISL_417030; EPI_ISL_413214; EPI_ISL_413213; EPI_ISL_412975 | Centre for Infectious Diseases and Microbiology Laboratory Services | NSW Health Pathology - Institute of Clinical Pathology and Medical Research; Westmead Hospital; University of Sydney | Eden J-S et al (https://www.nature.com/articles/s41591-020-1000-7) |
| EPI_ISL_413595 | Centre for Infectious Diseases and Microbiology Laboratory Services | NSW Health Pathology - Institute of Clinical Pathology and Medical Research; Westmead Hospital; University of Sydney | Rockett R et al (https://www.nature.com/articles/s41591-020-1000-7) |
| EPI_ISL_700389; EPI_ISL_700379; EPI_ISL_700384; EPI_ISL_508934; EPI_ISL_700375 | Centre hospitalier Métropole Savoie | CNR Virus des Infections Respiratoires - France SUD | Antonin Bal et al |
| EPI_ISL_536115 | Centre hospitalier régional du Grand Portage | Laboratoire de santé publique du Québec | Sandrine Moreira et al |
| EPI_ISL_639739 | Centre of Nanotechnologies, INCD IMT-Bucuresti (National Institute for Research and Development in Microtechnologies - Bucharest) | Centre of Nanotechnologies, INCD IMT-Bucuresti (National Institute for Research and Development in Microtechnologies - Bucharest) | Salceanu et al |
| EPI_ISL_413647 | Centro Hospital do Porto, E.P.E. - H. Geral de Santo Antonio | Instituto Nacional de Saude (INSA) | Raquel Guiomar et al |
| EPI_ISL_512670; EPI_ISL_527755 | Centro Nacional De Rehabilitacion Humberto Araya Rojas (Cenare) | Inciensa, Instituto Costarricense de Investigación y Enseñanza en Nutrición y Salud | Francisco Duarte et al |
| EPI_ISL_629024 | Centro de Biotecnología Vegetal, Universidad Andrés Bello, Center for Genome Regulation | Center for Mathematical Modeling and Center for Genome Regulation. Santiago, Chile | Bastias M et al |
| EPI_ISL_510536; EPI_ISL_509431; EPI_ISL_509435; EPI_ISL_529140; EPI_ISL_509432; EPI_ISL_509433; EPI_ISL_509430; EPI_ISL_529139 | Centro de Desenvolvimento Tecnologico em Saude, Fundacao Oswaldo Cruz | Centro de Desenvolvimento Tecnologico em Saude, Fundacao Oswaldo Cruz | Souza et al |
| EPI_ISL_635480; EPI_ISL_635506; EPI_ISL_635510; EPI_ISL_635564; EPI_ISL_635517; EPI_ISL_635561; EPI_ISL_635563; EPI_ISL_635495; EPI_ISL_635500; EPI_ISL_635528; EPI_ISL_635492; EPI_ISL_635545; EPI_ISL_635486; EPI_ISL_635515; EPI_ISL_635491; EPI_ISL_635560; EPI_ISL_635566; EPI_ISL_635521; EPI_ISL_635572; EPI_ISL_635540; EPI_ISL_635569 | Centro de Diagn√≥stico COVID-19 UABC Tijuana | Andersen lab at Scripps Research | SEARCH Alliance San Diego with Idanya Rubí Serafín Higuera et al |
| EPI_ISL_491934; EPI_ISL_491935; EPI_ISL_697797; EPI_ISL_491933; EPI_ISL_491941 | Centro de Investigaciones, Universidad de Especialidades Espíritu Santo | Institute of Microbiology, Universidad San Francisco de Quito | Derly Andrade et al |
| EPI_ISL_500429 | Centro de Investigación Biomédica de La Rioja - Hospital San Pedro Logroño | SeqCOVID-SPAIN consortium/IBV(CSIC) | María de Toro et al |
| EPI_ISL_639657; EPI_ISL_450524; EPI_ISL_639648; EPI_ISL_639678; EPI_ISL_639663; EPI_ISL_639655; EPI_ISL_639656; EPI_ISL_639683; EPI_ISL_639676; EPI_ISL_639672; EPI_ISL_450522; EPI_ISL_450520; EPI_ISL_450521; EPI_ISL_639640; EPI_ISL_639651; EPI_ISL_450523; EPI_ISL_639654; EPI_ISL_639679 | Centrālā Laboratorija | Latvian Biomedical Research and Study Centre | Ivars Silamiķelis et al |
| EPI_ISL_486415; EPI_ISL_501808; EPI_ISL_492989; EPI_ISL_492991; EPI_ISL_492990; EPI_ISL_492992; EPI_ISL_501288; EPI_ISL_515196; EPI_ISL_534202; EPI_ISL_534200; EPI_ISL_534208; EPI_ISL_501915; EPI_ISL_501287; EPI_ISL_501922; EPI_ISL_501896; EPI_ISL_534201; EPI_ISL_534222; EPI_ISL_486414; EPI_ISL_486412; EPI_ISL_486416; EPI_ISL_492988; EPI_ISL_534206; EPI_ISL_486437; EPI_ISL_486418; EPI_ISL_486390; EPI_ISL_486419; EPI_ISL_486391; EPI_ISL_486421; EPI_ISL_486420; EPI_ISL_486413; EPI_ISL_486411; EPI_ISL_501286 | Centrālā laboratorija | Latvian Biomedical Research and Study Centre | Ivars Silamiķelis et al |
| EPI_ISL_754174; EPI_ISL_754175 | Charité Universitätsmedizin Berlin, Institut für Virologie | Charité Universitätsmedizin Berlin, Institut für Virologie | Victor M Corman et al A |
| EPI_ISL_753843; EPI_ISL_753933; EPI_ISL_753732; EPI_ISL_753799; EPI_ISL_753722; EPI_ISL_753822; EPI_ISL_729497; EPI_ISL_729496; EPI_ISL_729478; EPI_ISL_729494; EPI_ISL_729495; EPI_ISL_753814; EPI_ISL_729484; EPI_ISL_729473; EPI_ISL_729474; EPI_ISL_753746; EPI_ISL_729499; EPI_ISL_729482; EPI_ISL_753725; EPI_ISL_729548; EPI_ISL_729404; EPI_ISL_753888; EPI_ISL_754186; EPI_ISL_729370; EPI_ISL_729408; EPI_ISL_754187; EPI_ISL_754193; EPI_ISL_754189; EPI_ISL_754184; EPI_ISL_754188; EPI_ISL_754190; EPI_ISL_753743; EPI_ISL_754006; EPI_ISL_729377; EPI_ISL_753949; EPI_ISL_753775; EPI_ISL_729493; EPI_ISL_754041; EPI_ISL_754039; EPI_ISL_753768; EPI_ISL_754003; EPI_ISL_753821; EPI_ISL_753887; EPI_ISL_753820; EPI_ISL_753758; EPI_ISL_729600; EPI_ISL_753877; EPI_ISL_753986; EPI_ISL_753954; EPI_ISL_753985; EPI_ISL_753815; EPI_ISL_754012; EPI_ISL_754015; EPI_ISL_729365; EPI_ISL_729409; EPI_ISL_729607; EPI_ISL_753865; EPI_ISL_753767; EPI_ISL_729475; EPI_ISL_729488; EPI_ISL_753919; EPI_ISL_753704; EPI_ISL_754026; EPI_ISL_753880; EPI_ISL_753879; EPI_ISL_753862; EPI_ISL_729413; EPI_ISL_753977; EPI_ISL_753978; EPI_ISL_753994; EPI_ISL_753777; EPI_ISL_753904; EPI_ISL_729355; EPI_ISL_729388; EPI_ISL_754044; EPI_ISL_753864; EPI_ISL_753874; EPI_ISL_754054; EPI_ISL_754010; EPI_ISL_754009; EPI_ISL_754014; EPI_ISL_754045; EPI_ISL_753825; EPI_ISL_753810; EPI_ISL_753778; EPI_ISL_753726; EPI_ISL_753857; EPI_ISL_753990; EPI_ISL_753855; EPI_ISL_753744; EPI_ISL_753960; EPI_ISL_753733; EPI_ISL_753962; EPI_ISL_753984; EPI_ISL_753991; EPI_ISL_753728; EPI_ISL_753771; EPI_ISL_753756; EPI_ISL_753856; EPI_ISL_753720; EPI_ISL_753965; EPI_ISL_753835; EPI_ISL_753854; EPI_ISL_754023; EPI_ISL_753963; EPI_ISL_753853; EPI_ISL_753964; EPI_ISL_753706; EPI_ISL_753972; EPI_ISL_753966; EPI_ISL_753981; EPI_ISL_753852; EPI_ISL_753955; EPI_ISL_754011; EPI_ISL_753872; EPI_ISL_753947; EPI_ISL_753774; EPI_ISL_753944; EPI_ISL_753988; EPI_ISL_754002; EPI_ISL_753937; EPI_ISL_754040; EPI_ISL_754001; EPI_ISL_753873; EPI_ISL_753902; EPI_ISL_753708; EPI_ISL_753999; EPI_ISL_753871; EPI_ISL_753992; EPI_ISL_754021; EPI_ISL_753757; EPI_ISL_753900; EPI_ISL_753763; EPI_ISL_753996; EPI_ISL_729346; EPI_ISL_753773; EPI_ISL_753868; EPI_ISL_753914; EPI_ISL_753976; EPI_ISL_753875; EPI_ISL_753910; EPI_ISL_753703; EPI_ISL_753974; EPI_ISL_753997; EPI_ISL_729530; EPI_ISL_729356; EPI_ISL_753998; EPI_ISL_753907; EPI_ISL_754000; EPI_ISL_753759; EPI_ISL_753941; EPI_ISL_753858; EPI_ISL_753995; EPI_ISL_729382; EPI_ISL_729602; EPI_ISL_753752; EPI_ISL_753982; EPI_ISL_753710; EPI_ISL_753737; EPI_ISL_754050; EPI_ISL_754053; EPI_ISL_754051; EPI_ISL_754049; EPI_ISL_754048; EPI_ISL_753836; EPI_ISL_753797; EPI_ISL_753798; EPI_ISL_753928; EPI_ISL_754192; EPI_ISL_753860; EPI_ISL_754022; EPI_ISL_753730; EPI_ISL_753967; EPI_ISL_753819; EPI_ISL_753761; EPI_ISL_753841; EPI_ISL_753812; EPI_ISL_754185; EPI_ISL_753715; EPI_ISL_754194; EPI_ISL_753869; EPI_ISL_753897; EPI_ISL_753712; EPI_ISL_753824; EPI_ISL_729556; EPI_ISL_753742; EPI_ISL_729601; EPI_ISL_753750; EPI_ISL_754031; EPI_ISL_753805; EPI_ISL_753806; EPI_ISL_753764; EPI_ISL_753808; EPI_ISL_753807; EPI_ISL_753809; EPI_ISL_753881; EPI_ISL_754017; EPI_ISL_753882; EPI_ISL_753876; EPI_ISL_753878; EPI_ISL_753892; EPI_ISL_753889; EPI_ISL_753911; EPI_ISL_753883; EPI_ISL_753859; EPI_ISL_753736; EPI_ISL_729555; EPI_ISL_753968; EPI_ISL_753755; EPI_ISL_729557; EPI_ISL_753826; EPI_ISL_753739; EPI_ISL_753848; EPI_ISL_753713; EPI_ISL_753748; EPI_ISL_753958; EPI_ISL_753983; EPI_ISL_753766; EPI_ISL_753942; EPI_ISL_753971; EPI_ISL_753956; EPI_ISL_753961; EPI_ISL_753957; EPI_ISL_753765; EPI_ISL_729406; EPI_ISL_753724; EPI_ISL_753970; EPI_ISL_753969; EPI_ISL_753823; EPI_ISL_753760; EPI_ISL_753723; EPI_ISL_753709; EPI_ISL_753923; EPI_ISL_753811; EPI_ISL_753935; EPI_ISL_753721; EPI_ISL_753929; EPI_ISL_753989; EPI_ISL_753993; EPI_ISL_753849; EPI_ISL_754046; EPI_ISL_753938; EPI_ISL_753705; EPI_ISL_753770; EPI_ISL_753813; EPI_ISL_753837; EPI_ISL_753861; EPI_ISL_753834; EPI_ISL_753718; EPI_ISL_753927; EPI_ISL_729534; EPI_ISL_753740; EPI_ISL_754057; EPI_ISL_754052; EPI_ISL_754025; EPI_ISL_754030; EPI_ISL_754036; EPI_ISL_754032; EPI_ISL_754027; EPI_ISL_754037; EPI_ISL_754028; EPI_ISL_754029; EPI_ISL_754034; EPI_ISL_754024; EPI_ISL_754033; EPI_ISL_754035; EPI_ISL_754038; EPI_ISL_753839; EPI_ISL_729477; EPI_ISL_729514; EPI_ISL_753946; EPI_ISL_754058; EPI_ISL_753747; EPI_ISL_753959; EPI_ISL_753754; EPI_ISL_753940; EPI_ISL_754008; EPI_ISL_754004; EPI_ISL_753867; EPI_ISL_753817; EPI_ISL_753776; EPI_ISL_753913; EPI_ISL_753915; EPI_ISL_753870; EPI_ISL_754019; EPI_ISL_753714; EPI_ISL_729605; EPI_ISL_753741; EPI_ISL_753829; EPI_ISL_753831; EPI_ISL_753828; EPI_ISL_729604; EPI_ISL_753833; EPI_ISL_753769; EPI_ISL_729387; EPI_ISL_753735; EPI_ISL_753830; EPI_ISL_753832; EPI_ISL_753827; EPI_ISL_754043; EPI_ISL_753749; EPI_ISL_753979; EPI_ISL_753702; EPI_ISL_753973; EPI_ISL_753948; EPI_ISL_753762; EPI_ISL_753803; EPI_ISL_753802; EPI_ISL_753801; EPI_ISL_753745; EPI_ISL_753804; EPI_ISL_729489; EPI_ISL_753772; EPI_ISL_753917; EPI_ISL_729533; EPI_ISL_753920; EPI_ISL_753925; EPI_ISL_753922; EPI_ISL_753921; EPI_ISL_753952; EPI_ISL_753738; EPI_ISL_753951; EPI_ISL_753953; EPI_ISL_753838; EPI_ISL_753753; EPI_ISL_753931; EPI_ISL_753934; EPI_ISL_753975; EPI_ISL_753734; EPI_ISL_754007; EPI_ISL_754055; EPI_ISL_729487; EPI_ISL_753846; EPI_ISL_753926; EPI_ISL_729492; EPI_ISL_754056; EPI_ISL_753932; EPI_ISL_753840; EPI_ISL_753844; EPI_ISL_753729; EPI_ISL_753847; EPI_ISL_729485; EPI_ISL_729480; EPI_ISL_729486; EPI_ISL_754042; EPI_ISL_753727; EPI_ISL_753918; EPI_ISL_754047; EPI_ISL_753987; EPI_ISL_753716; EPI_ISL_753980; EPI_ISL_754013; EPI_ISL_753731; EPI_ISL_753818; EPI_ISL_753905; EPI_ISL_754005; EPI_ISL_753886; EPI_ISL_753866; EPI_ISL_753890; EPI_ISL_753903; EPI_ISL_753896; EPI_ISL_753891; EPI_ISL_753898; EPI_ISL_729360; EPI_ISL_729479; EPI_ISL_729476; EPI_ISL_729483; EPI_ISL_753950; EPI_ISL_729481; EPI_ISL_753850; EPI_ISL_753851; EPI_ISL_729529; EPI_ISL_753936; EPI_ISL_753845; EPI_ISL_753945; EPI_ISL_753939; EPI_ISL_753800; EPI_ISL_729491; EPI_ISL_729490; EPI_ISL_753930; EPI_ISL_729531; EPI_ISL_753943; EPI_ISL_753924; EPI_ISL_754016; EPI_ISL_753717; EPI_ISL_754020; EPI_ISL_753912; EPI_ISL_753884; EPI_ISL_753909; EPI_ISL_753901; EPI_ISL_753816; EPI_ISL_753906; EPI_ISL_753908; EPI_ISL_753895; EPI_ISL_729400; EPI_ISL_753894; EPI_ISL_753885; EPI_ISL_729598; EPI_ISL_729410; EPI_ISL_753751; EPI_ISL_753899; EPI_ISL_753916; EPI_ISL_753707; EPI_ISL_753863; EPI_ISL_729398; EPI_ISL_729357; EPI_ISL_729603; EPI_ISL_753785; EPI_ISL_753786; EPI_ISL_753792; EPI_ISL_753788; EPI_ISL_753790; EPI_ISL_753784; EPI_ISL_753787; EPI_ISL_753796; EPI_ISL_753781; EPI_ISL_753719; EPI_ISL_753794; EPI_ISL_753783; EPI_ISL_753782; EPI_ISL_753789; EPI_ISL_753711; EPI_ISL_753791; EPI_ISL_753780; EPI_ISL_753779; EPI_ISL_753795; EPI_ISL_754191; EPI_ISL_753793; EPI_ISL_753842; EPI_ISL_729348; EPI_ISL_729599; EPI_ISL_753893; EPI_ISL_729606; EPI_ISL_729366; EPI_ISL_754018 | Charité Universitätsmedizin Berlin, Institut für Virologie/Labor Berlin | Charité Universitätsmedizin Berlin, Institut für Virologie | Victor M Corman et al A |
| EPI_ISL_516645; EPI_ISL_516644; EPI_ISL_516639; EPI_ISL_516643; EPI_ISL_516633; EPI_ISL_516641; EPI_ISL_516634; EPI_ISL_516642; EPI_ISL_516630; EPI_ISL_516635; EPI_ISL_516632; EPI_ISL_516631; EPI_ISL_516636; EPI_ISL_516629; EPI_ISL_516640; EPI_ISL_516638; EPI_ISL_516637 | Charité Universitätsmedizin Berlin, Institut für Virologie/Labor Berlin | Charité Universitätsmedizin Berlin, Institut für Virologie/Labor Berlin | Victor M Corman et al A |
| EPI_ISL_406862 | Charité Universitätsmedizin Berlin, Institute of Virology; Institut für Mikrobiologie der Bundeswehr, Munich | Charité Universitätsmedizin Berlin, Institute of Virology | Victor M Corman et al B (https://dx.doi.org/10.2807/1560-7917.ES.2020.25.13.2000305) |
| EPI_ISL_735496 | Chattogram Veterinary and Animal Sciences University | Central Biological Research Laboratory and Department of Biochemistry and Molecular Biology | Md. Imranul Hoq et al |
| EPI_ISL_735497 | Chattogram Veterinary and Animal Sciences University | Central Biological Research Laboratory and Department of Biochemistry and Molecular Biology | Mohammad Omar Faruque et al |
| EPI_ISL_480014; EPI_ISL_480012; EPI_ISL_480005 | Chiba Prefectural Institute of Public Health | Pathogen Genomics Center, National Institute of Infectious Diseases | Tsuyoshi Sekizuka et al C (https://www.medrxiv.org/content/10.1101/2020.07.01.20143958v1) |
| EPI_ISL_477134; EPI_ISL_700343; EPI_ISL_700330; EPI_ISL_625472; EPI_ISL_700332; EPI_ISL_700345; EPI_ISL_477125; EPI_ISL_700335; EPI_ISL_700344; EPI_ISL_700336; EPI_ISL_700331; EPI_ISL_700342; EPI_ISL_700329; EPI_ISL_700338; EPI_ISL_700337; EPI_ISL_700334; EPI_ISL_700328; EPI_ISL_700346; EPI_ISL_700339; EPI_ISL_700347; EPI_ISL_625468; EPI_ISL_477128; EPI_ISL_492028; EPI_ISL_468078; EPI_ISL_468070 | Child Health Research Foundation | Child Health Research Foundation | Senjuti Saha et al |
| EPI_ISL_491115 | Cicin-Sain Lab | Cicin-Sain Lab | M. Zeeshan Chaudhry et al |
| EPI_ISL_636981 | City of Chimoio | KRISP, KZN Research Innovation and Sequencing Platform | Ismael N et al |
| EPI_ISL_561337 | Civil Hospital, Panchkula | CSIR-Institute of Microbial Technology | Kanika Bansal et al |
| EPI_ISL_462450; EPI_ISL_462470; EPI_ISL_462476; EPI_ISL_462471; EPI_ISL_462462; EPI_ISL_462465; EPI_ISL_462451; EPI_ISL_462466; EPI_ISL_462468; EPI_ISL_462475; EPI_ISL_462473; EPI_ISL_462472; EPI_ISL_462461; EPI_ISL_462458; EPI_ISL_462452; EPI_ISL_462460; EPI_ISL_462454; EPI_ISL_462453; EPI_ISL_462455; EPI_ISL_462456; EPI_ISL_462457; EPI_ISL_462459; EPI_ISL_462469; EPI_ISL_462474 | Clinical Center, University of Sarajevo | Charite Universitatsmedizin Berlin, Institute of Virology | Victor M Corman et al A |
| EPI_ISL_677718; EPI_ISL_677715 | Clinical Hospital - Bitola | Research Center for Genetic Engineering and Biotechnology "Georgi D. Efremov" , Macedonian Academy of Sciences and Arts | RCGEB - MASA et al |
| EPI_ISL_516430; EPI_ISL_677717; EPI_ISL_516427; EPI_ISL_516431; EPI_ISL_678259; EPI_ISL_677720; EPI_ISL_677721; EPI_ISL_677724; EPI_ISL_516429; EPI_ISL_516428; EPI_ISL_677716; EPI_ISL_678250 | Clinical Hospital - Shtip | Research Center for Genetic Engineering and Biotechnology "Georgi D. Efremov" , Macedonian Academy of Sciences and Arts | RCGEB - MASA et al |
| EPI_ISL_747194 | Clinical Microbiology Laboratory, Faculty of Medicine, Universitas Indonesia | Clinical Microbiology Laboratory, Faculty of Medicine, Universitas Indonesia | Fera Ibrahim et al |
| EPI_ISL_447448; EPI_ISL_447464; EPI_ISL_447469; EPI_ISL_447455; EPI_ISL_447454 | Clinical Microbiology Laboratory, Sheba Medical Center | Stern Lab | Stern Lab et al (https://dx.doi.org/10.1101/2020.05.21.20104521) |
| EPI_ISL_483572; EPI_ISL_490204 | Clinical Microbiology Laboratory- Basurto University Hospital | Biocruces-Bizkaia | Mikel J. Urrutikoetxea-Gutierrez et al |
| EPI_ISL_447328 | Clinical Virology Laboratory, Soroka Medical Center and the Faculty of Health Sciences, Ben-Gurion University of the Negev | Stern Lab | Stern Lab et al (https://dx.doi.org/10.1101/2020.05.21.20104521) |
| EPI_ISL_447411; EPI_ISL_447381; EPI_ISL_447356; EPI_ISL_447358 | Clinical Virology Unit, Hadassah Hebrew University Medical Center | Stern Lab | Stern Lab et al (https://dx.doi.org/10.1101/2020.05.21.20104521) |
| EPI_ISL_753526; EPI_ISL_753618 | Clinical virology Laboratory, Children's Hospital Los Angeles | Center for Personalized Medicine, Children's Hospital Los Angeles | Gai et al et al |
| EPI_ISL_591070 | College of Veterinary Medicine, Chungnam National University | College of Veterinary Medicine, Chungnam National University | Seo et al |
| EPI_ISL_539823 | Communicable Disease Branch | Hong Kong Department of Health | Alan K.L. Tsang et al |
| EPI_ISL_510528 | Communicable Disease Laboratory, Public Health Directorate | Communicable Disease Laboratory, Public Health Directorate | Al Wasti et al |
| EPI_ISL_632906; EPI_ISL_632907 | Communicable Disease Laboratory, Public Health Directorate | Communicable Disease Laboratory, Public Health Directorate | AlAbbas et al |
| EPI_ISL_632260 | Communicable Disease Laboratory, Public Health Directorate | Communicable Disease Laboratory, Public Health Directorate | AlTaif et al |
| EPI_ISL_632267; EPI_ISL_632266; EPI_ISL_632285; EPI_ISL_632262; EPI_ISL_632261; EPI_ISL_632263; EPI_ISL_632284; EPI_ISL_632278; EPI_ISL_632280; EPI_ISL_632251; EPI_ISL_632282; EPI_ISL_632283; EPI_ISL_632270; EPI_ISL_632264; EPI_ISL_632271; EPI_ISL_632265 | Communicable Disease Laboratory, Public Health Directorate | Communicable Disease Laboratory, Public Health Directorate | AlWasti et al |
| EPI_ISL_684030; EPI_ISL_681301; EPI_ISL_681309; EPI_ISL_681308; EPI_ISL_684028; EPI_ISL_682300; EPI_ISL_682302; EPI_ISL_684033; EPI_ISL_678270; EPI_ISL_682320; EPI_ISL_678269; EPI_ISL_681315; EPI_ISL_682303; EPI_ISL_681319 | Communicable Disease Laboratory, Public Health Directorate | Communicable Disease Laboratory, Public Health Directorate | Alwasti et al |

| | | | |
|---|---|---|---|
| EPI_ISL_510531 | Communicable Disease Laboratory, Public Health Directorate | Communicable Disease Laboratory, Public Health Directorate | Shehab et al |
| EPI_ISL_485401 | Communicable Disease Laboratory, Public Health Directorate | Communicable Disease Laboratory, Public Health Directorate | Zaed et al |
| EPI_ISL_537380 | Complejo Hospitalario Universitario de Vigo | SeqCOVID-SPAIN consortium/IBV(CSIC) | Benito Regueiro et al |
| EPI_ISL_735494 | Cox's Bazar Medical College | Central Biological Research Laboratory and Department of Biochemistry and Molecular Biology | Md. Imranul Hoq et al |
| EPI_ISL_735493 | Cox's Bazar Medicila College | Central Biological Research Laboratory and Department of Biochemistry and Molecular Biology Central Biological Research Laboratory and Department of Biochemistry and Molecular Biology | Robiul Hasan Bhuiyan et al |
| EPI_ISL_476178 | DB Diagnósticos do Brasil | Instituto de Medicina Tropical da Univesidade de São Paulo | Samples: Nelson Gaburo Jr et al (https://dx.doi.org/10.1126/science.abd2161) |
| EPI_ISL_515932 | DH | Department of Neurovirology, National Institute of Mental Health and Neuroscience (NIMHANS) | Chitra Pattabiraman et al |
| EPI_ISL_747283 | DNA SOLUTION LTD. | DNA SOLUTION LTD. | Md. Imran Khan et al A |
| EPI_ISL_445216 | DNA Solution Ltd. | DNA Solution Ltd. | Md. Imran Khan et al B (https://dx.doi.org/10.1101/2020.06.07.138800) |
| EPI_ISL_745420 | DOHMH Crown Heights | New York City Public Health Laboratory | Jade Wang et al |
| EPI_ISL_683951 | DOHMH Jamaica | New York City Public Health Laboratory | Jade Wang et al |
| EPI_ISL_671688 | DOHMH Morrisania | New York City Public Health Laboratory | Jade Wang et al |
| EPI_ISL_416543; EPI_ISL_416542; EPI_ISL_421652 | Dasman Diabetes Institute | Dasman Diabetes Institute | Fahd Al-Mulla et al |
| EPI_ISL_416541 | Dasman Diabetes Institute and Virology Laboratory Ministry of Health | Dasman Diabetes Institute | Fahd Al-Mulla et al |
| EPI_ISL_708414; EPI_ISL_708388; EPI_ISL_576150 | Delaware Public Health Lab | Delaware Public Health Lab | Gregory Hovan et al |
| EPI_ISL_430808; EPI_ISL_430797; EPI_ISL_430802; EPI_ISL_430796 | Departamento de Biología y genética molecular, IACA Laboratorios. | Área de Secuenciación del Laboratorio de Virología del Hospital de Niños Dr. Ricardo Gutierrez | Nabaes Jodar et al A (https://dx.doi.org/10.1101/2020.05.23.20111443) |
| EPI_ISL_430806 | Departamento de Biología y genética molecular, IACA Laboratorios. | Área de Secuenciación del Laboratorio de Virología del Hospital de Niños Dr. Ricardo Gutierrez | Nabaes Jodar et al B |
| EPI_ISL_444493 | Departamento de Laboratorios de Salud Publica (DLSP, Division Epidemiologia, Ministerio de Salud Publica) | Facultad de Ciencias (Sección Genética Evolutiva, Sección Virología). | Panzera et al |
| EPI_ISL_508663; EPI_ISL_508667; EPI_ISL_508675 | Departamento de Microbiología, CDB, Hospital Clínic, Barcelona | SeqCOVID-SPAIN consortium/IBV(CSIC) | Andrea Vergara et al |
| EPI_ISL_516923; EPI_ISL_516931; EPI_ISL_516932; EPI_ISL_516927; EPI_ISL_516926; EPI_ISL_516925; EPI_ISL_516933; EPI_ISL_516930; EPI_ISL_516928; EPI_ISL_516929; EPI_ISL_516922; EPI_ISL_516924 | Department for Molecular Diagnostics, Centre for Medical Microbiology, Institute of Public Health of Montenegro | Charité Universitätsmedizin Berlin, Institut für Virologie | Victor M Corman et al A |
| EPI_ISL_481380 | Department for Virology, Molecular Biology and Genome Research, R. G. Lugar Center for Public Health Research, National Center for Disease Control and Public Health (NCDC) of Georgia. | Department for Virology, Molecular Biology and Genome Research, R. G. Lugar Center for Public Health Research, National Center for Disease Control and Public Health (NCDC) of Georgia. | Ana Papkiauri et al |
| EPI_ISL_763062; EPI_ISL_470876 | Department for Virology, Molecular Biology and Genome Research, R. G. Lugar Center for Public Health Research, National Center for Disease Control and Public Health (NCDC) of Georgia. | Department for Virology, Molecular Biology and Genome Research, R. G. Lugar Center for Public Health Research, National Center for Disease Control and Public Health (NCDC) of Georgia. | Giorgi Tomashvili et al |
| EPI_ISL_470877; EPI_ISL_447056 | Department for Virology, Molecular Biology and Genome Research, R. G. Lugar Center for Public Health Research, National Center for Disease Control and Public Health (NCDC) of Georgia. | Department for Virology, Molecular Biology and Genome Research, R. G. Lugar Center for Public Health Research, National Center for Disease Control and Public Health (NCDC) of Georgia. | Gvantsa Brachveli et al |
| EPI_ISL_420144 | Department for Virology, Molecular Biology and Genome Research, R. G. Lugar Center for Public Health Research, National Center for Disease Control and Public Health (NCDC) of Georgia. | Department for Virology, Molecular Biology and Genome Research, R. G. Lugar Center for Public Health Research, National Center for Disease Control and Public Health (NCDC) of Georgia. | Gvantsa Chanturia et al |
| EPI_ISL_420142 | Department for Virology, Molecular Biology and Genome Research, R. G. Lugar Center for Public Health Research, National Center for Disease Control and Public Health (NCDC) of Georgia. | Department for Virology, Molecular Biology and Genome Research, R. G. Lugar Center for Public Health Research, National Center for Disease Control and Public Health (NCDC) of Georgia. | Marine Murtskhvaladze et al |
| EPI_ISL_754181; EPI_ISL_471529; EPI_ISL_447055 | Department for Virology, Molecular Biology and Genome Research, R. G. Lugar Center for Public Health Research, National Center for Disease Control and Public Health (NCDC) of Georgia. | Department for Virology, Molecular Biology and Genome Research, R. G. Lugar Center for Public Health Research, National Center for Disease Control and Public Health (NCDC) of Georgia. | Meri Pantsulaia et al |
| EPI_ISL_420140 | Department for Virology, Molecular Biology and Genome Research, R. G. Lugar Center for Public Health Research, National Center for Disease Control and Public Health (NCDC) of Georgia. | Department for Virology, Molecular Biology and Genome Research, R. G. Lugar Center for Public Health Research, National Center for Disease Control and Public Health (NCDC) of Georgia. | Nato Kotaria et al |
| EPI_ISL_481483 | Department for Virology, Molecular Biology and Genome Research, R. G. Lugar Center for Public Health Research, National Center for Disease Control and Public Health (NCDC) of Georgia. | Department for Virology, Molecular Biology and Genome Research, R. G. Lugar Center for Public Health Research, National Center for Disease Control and Public Health (NCDC) of Georgia. | Nino Berishvili et al |
| EPI_ISL_754180 | Department for Virology, Molecular Biology and Genome Research, R. G. Lugar Center for Public Health Research, National Center for Disease Control and Public Health (NCDC) of Georgia. | Department for Virology, Molecular Biology and Genome Research, R. G. Lugar Center for Public Health Research, National Center for Disease Control and Public Health (NCDC) of Georgia. | Salome Javashvili et al |
| EPI_ISL_477169 | Department for Virology, Molecular Biology and Genome Research, R. G. Lugar Center for Public Health Research, National Center for Disease Control and Public Health (NCDC) of Georgia. | Department for Virology, Molecular Biology and Genome Research, R. G. Lugar Center for Public Health Research, National Center for Disease Control and Public Health (NCDC) of Georgia. | Tata Imnadze et al |
| EPI_ISL_632935 | Department of Acute Infectious Diseases Control and Prevention, Yunnan Provincial Center for Disease Control and Prevention | Department of Acute Infectious Diseases Control and Prevention, Yunnan Provincial Center for Disease Control and Prevention | Meiling Zhang et al |
| EPI_ISL_632934 | Department of Acute Infectious Diseases Control and Prevention, Yunnan Provincial Center for Disease Control and Prevention | Department of Acute Infectious Diseases Control and Prevention,Yunnan Provincial Center for Disease Control and Prevention | Meiling Zhang et al |
| EPI_ISL_515086; EPI_ISL_515109; EPI_ISL_515105; EPI_ISL_515098; EPI_ISL_515106; EPI_ISL_515093; EPI_ISL_515082; EPI_ISL_515103; EPI_ISL_515107; EPI_ISL_515111; EPI_ISL_515097; EPI_ISL_515083; EPI_ISL_515092 | Department of Biochemistry, Cell and Molecular Biology | WACCBIP, University of Ghana | Ngoi et al |
| EPI_ISL_447121; EPI_ISL_678413; EPI_ISL_447148; EPI_ISL_498138; EPI_ISL_498129; EPI_ISL_455962; EPI_ISL_455968; EPI_ISL_455958; EPI_ISL_455967; EPI_ISL_498132; EPI_ISL_484700; EPI_ISL_484699; EPI_ISL_498629; EPI_ISL_498628; EPI_ISL_540564; EPI_ISL_626249; EPI_ISL_737325; EPI_ISL_641558; EPI_ISL_641557; EPI_ISL_707765; EPI_ISL_661227; EPI_ISL_722953; EPI_ISL_455974; EPI_ISL_455971; EPI_ISL_455976; EPI_ISL_484695; EPI_ISL_455978; EPI_ISL_471430; EPI_ISL_737326; EPI_ISL_540506; EPI_ISL_540505; EPI_ISL_540503; EPI_ISL_540560; EPI_ISL_515080; EPI_ISL_540474; EPI_ISL_540490; EPI_ISL_540501; EPI_ISL_540523; EPI_ISL_540551; EPI_ISL_540565; EPI_ISL_515056; EPI_ISL_515068; EPI_ISL_661217; EPI_ISL_737350; EPI_ISL_737349; EPI_ISL_737369; EPI_ISL_581626; EPI_ISL_626243; EPI_ISL_678468; EPI_ISL_722962; EPI_ISL_737351; EPI_ISL_678464; EPI_ISL_708459; EPI_ISL_678435 | Department of Clinical Microbiology | GIGA Medical Genomics | Keith Durkin et al A |
| EPI_ISL_418627; EPI_ISL_424653; EPI_ISL_427371 | Department of Clinical Microbiology | GIGA Medical Genomics | Keith Durkin et al B (https://dx.doi.org/10.1101/2020.05.05.078758) |
| EPI_ISL_452035 | Department of Clinical Microbiology, Copenhagen University Hospital, Hvidovre, Kettegaard Alle 30, 2650 Hvidovre. | Albertsen lab, Department of Chemistry and Bioscience, Aalborg University, Denmark | Rasmus Kirkegaard et al |
| EPI_ISL_419224; EPI_ISL_419228; EPI_ISL_419227 | Department of Clinical Pathology, Pamela Youde Nethersole Eastern Hospital | Department of Health Technology and Informatics, Faculty of Health and Social Science, The Hong Kong Polytechnic University | Kenneth Siu-Sing LEUNG et al (https://dx.doi.org/10.1101/2020.03.30.20045740) |
| EPI_ISL_419242 | Department of Clinical Pathology, Tuen Mun Hospital | Department of Health Technology and Informatics, Faculty of Health and Social Science, The Hong Kong Polytechnic University | Kenneth Siu-Sing LEUNG et al (https://dx.doi.org/10.1101/2020.03.30.20045740) |
| EPI_ISL_419231 | Department of Clinical Pathology, Tuen Mun Hospital, 23 Tsing Chung Koon Road, Tuen Mun, N.T. | Department of Health Technology and Informatics, Faculty of Health and Social Science, The Hong Kong Polytechnic University | Kenneth Siu-Sing LEUNG et al (https://dx.doi.org/10.1101/2020.03.30.20045740) |
| EPI_ISL_481259; EPI_ISL_481261; EPI_ISL_481255 | Department of Emerging Infectious Diseases, Institute of Tropical Medicine, Nagasaki University | Department of Emerging Infectious Diseases, Institute of Tropical Medicine, Nagasaki University | Jiro Yasuda et al |
| EPI_ISL_507214 | Department of Experimental Modeling and Pathogenesis of Infectious Diseases | WHO National Influenza Centre Russian Federation | Andrey Komissarov et al A |
| EPI_ISL_610164; EPI_ISL_610172; EPI_ISL_610202; EPI_ISL_610197; EPI_ISL_610216; EPI_ISL_610193; EPI_ISL_610223; EPI_ISL_610215; EPI_ISL_610203; EPI_ISL_610224; EPI_ISL_610225; EPI_ISL_610198; EPI_ISL_610206; EPI_ISL_610207; EPI_ISL_610211; EPI_ISL_610200; EPI_ISL_610210 | Department of Health Technology and Informatics, The Hong Kong Polytechnic University | Department of Health Technology and Informatics, The Hong Kong Polytechnic University | Siu et al |
| EPI_ISL_417444 | Department of Healthcare Biotechnology, National University of Sciences and Technology (NUST) | Department of Healthcare Biotechnology, National University of Sciences and Technology (NUST) | Javed et al (http://biorxiv.org/lookup/doi/10.1101/2020.04.01.019463) |
| EPI_ISL_522407; EPI_ISL_522408 | Department of Infection Prevention and Infectious Diseases, University Hospital Regensburg | Department of Infection Prevention and Infectious Diseases, University Hospital Regensburg | Fritsch et al |
| EPI_ISL_513307; EPI_ISL_513303; EPI_ISL_513302; EPI_ISL_513304; EPI_ISL_513299; EPI_ISL_513306; EPI_ISL_513305; EPI_ISL_513298; EPI_ISL_513300; EPI_ISL_513301 | Department of Infection Prevention and Infectious Diseases, University Hospital Regensburg | University Hospital Regensburg | Fritsch et al |
| EPI_ISL_605929 | Department of Infectious Disease Prevention and Control, Henan Provincial Center for Disease Control and Prevention | Department of Infectious Disease Prevention and Control, Henan Provincial Center for Disease Control and Prevention | Li et al |
| EPI_ISL_636993; EPI_ISL_636994; EPI_ISL_637000; EPI_ISL_637009; EPI_ISL_637020; EPI_ISL_637016; EPI_ISL_637017; EPI_ISL_637002; EPI_ISL_568570; EPI_ISL_636995; EPI_ISL_637005; EPI_ISL_568573; EPI_ISL_568575; EPI_ISL_637021; EPI_ISL_637013; EPI_ISL_568576; EPI_ISL_637015; EPI_ISL_637018; EPI_ISL_636997; EPI_ISL_568564; EPI_ISL_637019; EPI_ISL_637075 | Department of Infectious Diseases and Immunology, National Hospital Organization Nagoya Medical Center | Clinical Research Center, National Hospital Organization Nagoya Medical Center | Yoshihiro Nakata et al |
| EPI_ISL_457749 | Department of Infectious Diseases, Istituto Superiore di Sanità, Roma , Italy | Army Medical and Veterinary Research Center | Paola Stefanelli et al B |
| EPI_ISL_412973 | Department of Infectious Diseases, Istituto Superiore di Sanità, Roma , Italy | Virology Laboratory, Scientific Department, Army Medical Center | Paola Stefanelli et al A (https://dx.doi.org/10.2807/1560-7917.ES.2020.25.13.2000305) |
| EPI_ISL_412974 | Department of Infectious Diseases, Istituto Superiore di Sanità, Rome, Italy | Virology Laboratory, Scientific Department, Army Medical Center | Paola Stefanelli et al A (https://dx.doi.org/10.2807/1560-7917.ES.2020.25.13.2000305) |
| EPI_ISL_644946; EPI_ISL_644947; EPI_ISL_644952; EPI_ISL_644953; EPI_ISL_644956; EPI_ISL_644954; EPI_ISL_644977; EPI_ISL_644945; EPI_ISL_644948; EPI_ISL_644949; EPI_ISL_644951; EPI_ISL_644950; EPI_ISL_644990; EPI_ISL_644997; EPI_ISL_644988; EPI_ISL_644970; EPI_ISL_644972; EPI_ISL_644982 | Department of Infectious Diseases, Keio University School of Medicine, Tokyo, Japan | Center for Medical Genetics, Keio University School of Medicine, Tokyo, Japan | Kenjiro Kosaki et al A |
| EPI_ISL_479868; EPI_ISL_480100; EPI_ISL_480092 | Department of Infectious Diseases, Kobe Institute of Health | Pathogen Genomics Center, National Institute of Infectious Diseases | Tsuyoshi Sekizuka et al C (https://www.medrxiv.org/content/10.1101/2020.07.01.20143958v1) |

| | | | |
|---|---|---|---|
| EPI_ISL_411219; EPI_ISL_411220; EPI_ISL_411218 | Department of Infectious and Tropical Diseases, Bichat Claude Bernard Hospital, Paris | Laboratoire Virpath, CIRI U111, UCBL1, INSERM, CNRS, ENS Lyon | Olivier Terrier et al (https://dx.doi.org/10.2807/1560-7917.ES.2020.25.13.2000305) |
| EPI_ISL_406597; EPI_ISL_406596; EPI_ISL_408430 | Department of Infectious and Tropical Diseases, Bichat Claude Bernard Hospital, Paris | National Reference Center for Viruses of Respiratory Infections, Institut Pasteur, Paris | Mélanie Albert et al A (https://dx.doi.org/10.1101/2020.04.24.059576) |
| EPI_ISL_410720; EPI_ISL_410984 | Department of Infectious and Tropical Diseases, Bichat Claude Bernard Hospital, Paris | National Reference Center for Viruses of Respiratory Infections, Institut Pasteur, Paris | Mélanie Albert et al B (https://dx.doi.org/10.2807/1560-7917.ES.2020.25.13.2000305) |
| EPI_ISL_413019 | Department of Internal Medicine, Triemli Hospital | Institute of Medical Virology, University of Zurich | Stefan Schmutz et al A |
| EPI_ISL_413020 | Department of Internal Medicine, Triemli Hospital | Institute of Medical Virology, University of Zurich | Stefan Schmutz et al C (https://virological.org/t/phylodynamic-analyses-based-on-11-genomes-from-the-italian-outbreak/426) |
| EPI_ISL_477180 | Department of Laboratory Medicine Tan Tock Seng Hospital | Department of Laboratory Medicine Tan Tock Seng Hospital | Chen YYC et al |
| EPI_ISL_408489; EPI_ISL_463007; EPI_ISL_447621; EPI_ISL_447622; EPI_ISL_422409; EPI_ISL_740547; EPI_ISL_738064; EPI_ISL_693302; EPI_ISL_693305; EPI_ISL_693304; EPI_ISL_534336; EPI_ISL_738065; EPI_ISL_693303; EPI_ISL_693306; EPI_ISL_422414 | Department of Laboratory Medicine, National Taiwan University Hospital | Microbial Genomics Core Lab, National Taiwan University Centers of Genomic and Precision Medicine | Shiou-Hwei Yeh et al A |
| EPI_ISL_410218 | Department of Laboratory Medicine, National Taiwan University Hospital | Microbial Genomics Core Lab, National Taiwan University Centers of Genomic and Precision Medicine | Shiou-Hwei Yeh et al B (http://biorxiv.org/lookup/doi/10.1101/2020.04.01.019463) |
| EPI_ISL_538435; EPI_ISL_648043; EPI_ISL_648071; EPI_ISL_648100; EPI_ISL_648101; EPI_ISL_648093; EPI_ISL_648066; EPI_ISL_648797; EPI_ISL_648753; EPI_ISL_648748; EPI_ISL_648667; EPI_ISL_648758; EPI_ISL_648739; EPI_ISL_476817; EPI_ISL_538481; EPI_ISL_507004; EPI_ISL_648065 | Department of Laboratory Medicine, Tan Tock Seng Hospital | Department of Laboratory Medicine, Tan Tock Seng Hospital | Chen YYC et al |
| EPI_ISL_477170 | Department of Laboratory, Medicine Tan Tock Seng Hospital | Department of Laboratory Medicine Tan Tock Seng Hospital | Chen YYC et al |
| EPI_ISL_710483; EPI_ISL_710484 | Department of Medical Laboratory Sciences, Arab American University | Department of Medical Laboratory Sciences, Arab American University | Al-Jawabreh et al |
| EPI_ISL_591012 | Department of Medical Microbiology - section Molde, Molde Hospital | Norwegian Institute of Public Health, Department of Virology | Kathrine Stene-Johansen et al |
| EPI_ISL_708144; EPI_ISL_708106; EPI_ISL_635126; EPI_ISL_635135; EPI_ISL_708143; EPI_ISL_591014; EPI_ISL_635096; EPI_ISL_759970; EPI_ISL_759972; EPI_ISL_759971; EPI_ISL_668420; EPI_ISL_708142; EPI_ISL_635165; EPI_ISL_668404 | Department of Medical Microbiology, St. Olavs hospital | Norwegian Institute of Public Health, Department of Virology | Kathrine Stene-Johansen et al |
| EPI_ISL_501220; EPI_ISL_501181; EPI_ISL_501221; EPI_ISL_501223; EPI_ISL_501179; EPI_ISL_501226; EPI_ISL_501228; EPI_ISL_501200 | Department of Medical Microbiology, University Malaya Medical Centre | Department of Medical Microbiology, Faculty of Medicine, University of Malaya | Yoong Min CHONG et al |
| EPI_ISL_492886; EPI_ISL_492910 | Department of Medical Microbiology, Western Sussex Hospitals NHS Foundation Trust, St Richard's Hospital | Wellcome Sanger Institute for the COVID-19 Genomics UK (COG-UK) consortium | Manasa Mutingwende et al |
| EPI_ISL_512844 | Department of Medical Research | DMR_Myanmar | Myat Htut Nyunt et al |
| EPI_ISL_476869 | Department of MicroBiology, Government Medical College, Surat | Gujarat Biotechnology Research Centre | Nikha Trivedi et al (https://www.biorxiv.org/content/10.1101/2020.07.10.197095v1) |
| EPI_ISL_495077 | Department of MicroBiology, Government Medical College, Surat | Gujarat Biotechnology Research Centre | Raghawendra Kumar et al |
| EPI_ISL_483850 | Department of Microbiology, Government Medical College, Surat | Gujarat Biotechnology Research Centre | Nitin Savaliya et al (https://www.biorxiv.org/content/10.1101/2020.07.10.197095v1) |
| EPI_ISL_497797; EPI_ISL_497827; EPI_ISL_497820; EPI_ISL_497818; EPI_ISL_497808; EPI_ISL_497784; EPI_ISL_497851; EPI_ISL_498270; EPI_ISL_497796; EPI_ISL_497782; EPI_ISL_497843; EPI_ISL_497803; EPI_ISL_497841; EPI_ISL_510538; EPI_ISL_497823; EPI_ISL_497865; EPI_ISL_497838; EPI_ISL_497770; EPI_ISL_498271; EPI_ISL_497802; EPI_ISL_497840; EPI_ISL_497842; EPI_ISL_516799 | Department of Microbiology, The University of Hong Kong | Department of Microbiology, The University of Hong Kong | Kelvin K.W. To et al |
| EPI_ISL_693299 | Department of Microbiology, Yokohama City University School of Medicine | Department of Microbiology, Yokohama City University School of Medicine | Kei Miyakawa et al |
| EPI_ISL_463742; EPI_ISL_463744; EPI_ISL_463743; EPI_ISL_463741; EPI_ISL_463748; EPI_ISL_463746; EPI_ISL_463745; EPI_ISL_463747 | Department of Molecular Virology, Cyprus Institute of Neurology and Genetics | Department of Molecular Virology, Cyprus Institute of Neurology and Genetics | Jan Richter et al |
| EPI_ISL_596451; EPI_ISL_594185; EPI_ISL_594188; EPI_ISL_594186; EPI_ISL_594187; EPI_ISL_596452; EPI_ISL_596455 | Department of Pathology, School of Medicine, Imam Khomeini Hospital, Tehran University of Medical Sciences | Genetics Research Center, University of Social Welfare and Rehabilitation Sciences | Zohreh Fattahi et al |
| EPI_ISL_582033 | Department of Pathology, School of Medicine, Imam Khomeini Hospital, Tehran University of Medical Sciences | Genetics Research Center. University Of Social Welfare And Rehabilitation Sciences | Zohreh Fattahi et al |
| EPI_ISL_417185; EPI_ISL_417180 | Department of Pathology, United Christian Hospital | Department of Health Technology and Informatics, Faculty of Health and Social Science, The Hong Kong Polytechnic University | Kenneth Siu-Sing LEUNG et al (https://dx.doi.org/10.1101/2020.03.30.20045740) |
| EPI_ISL_425358; EPI_ISL_433689 | Department of Pathology, University of Cambridge | COVID-19 Genomics UK (COG-UK) Consortium | Luke W Meredith et al A (https://dx.doi.org/10.1016/S1473-3099(20)30562-4) |
| EPI_ISL_441710; EPI_ISL_441592; EPI_ISL_440543; EPI_ISL_440535; EPI_ISL_443432; EPI_ISL_440823; EPI_ISL_438301 | Department of Pathology, University of Cambridge | Wellcome Sanger Institute for the COVID-19 Genomics UK (COG-UK) consortium | Luke W Meredith et al B (https://virological.org/t/preliminary-analysis-of-sars-cov-2-importation-establishment-of-uk-transmission-lineages/507) |
| EPI_ISL_576147; EPI_ISL_576146; EPI_ISL_576149; EPI_ISL_576148 | Department of Respiratory & Other Viral Infections of L.V. Gromashevsky Institute of Epidemiology & Infectious Diseases NAMS of Ukraine | Department of Respiratory & Other Viral Infections of L.V. Gromashevsky Institute of Epidemiology & Infectious Diseases NAMS of Ukraine, JSC "Farmak" | Alla Mironenko et al |
| EPI_ISL_582509 | Department of Respiratory and other Viral Infections of L.V. Gromashevsky Institute of Epidemiology & Infectious Diseases NAMS of Ukraine | Department of Respiratory and other Viral Infections of L.V.Gromashevsky Institute of Epidemiology & Infectious Diseases NAMS of Ukraine, JSC "Farmak" | Alla Mironenko et al |
| EPI_ISL_582510 | Department of Respiratory and other Viral Infections of L.V. Gromashevsky Institute of Epidemiology & Infectious Diseases NAMS of Ukraine | Department of Respiratory and other Viral Infections of L.V.Gromashevsky Institute of Epidemiology & Infectious Diseases NAMS of Ukraine, JSC "Farmak" | Alla Mironenko et al |
| EPI_ISL_654818; EPI_ISL_582512; EPI_ISL_582511; EPI_ISL_582513; EPI_ISL_654820; EPI_ISL_654819 | Department of Respiratory and other Viral Infections of L.V. Gromashevsky Institute of Epidemiology & Infectious Diseases NAMS of Ukraine | Department of Respiratory and other Viral Infections of L.V.Gromashevsky Institute of Epidemiology & Infectious Diseases NAMS of Ukraine, JSC "Farmak" | Alla Mironenko et al |
| EPI_ISL_732949; EPI_ISL_732948; EPI_ISL_732950; EPI_ISL_732951; EPI_ISL_732954; EPI_ISL_732953; EPI_ISL_732959; EPI_ISL_732955; EPI_ISL_732956; EPI_ISL_732952; EPI_ISL_732957; EPI_ISL_732958 | Department of Tropical Parasitology | Laboratory of Recombinant Vaccines | Lukasz Rabalski et al |
| EPI_ISL_450199; EPI_ISL_450203; EPI_ISL_450204; EPI_ISL_450208; EPI_ISL_450207; EPI_ISL_450198; EPI_ISL_450209; EPI_ISL_450200 | Department of Virology | Department of Virology | Boehmer et al |
| EPI_ISL_407084 | Department of Virology III, National Institute of Infectious Diseases | Pathogen Genomics Center, National Institute of Infectious Diseases | Tsuyoshi Sekizuka et al A |
| EPI_ISL_418411; EPI_ISL_418405; EPI_ISL_414640; EPI_ISL_737210; EPI_ISL_418399; EPI_ISL_755969; EPI_ISL_756044; EPI_ISL_756110; EPI_ISL_737034; EPI_ISL_759899; EPI_ISL_756165; EPI_ISL_759810; EPI_ISL_756076; EPI_ISL_756057; EPI_ISL_755996; EPI_ISL_756005; EPI_ISL_756031; EPI_ISL_755976; EPI_ISL_755998; EPI_ISL_755997; EPI_ISL_756062; EPI_ISL_737235; EPI_ISL_756183; EPI_ISL_756181; EPI_ISL_737068; EPI_ISL_737064; EPI_ISL_737067; EPI_ISL_732652; EPI_ISL_757394; EPI_ISL_757389; EPI_ISL_737066; EPI_ISL_737065; EPI_ISL_757380; EPI_ISL_759924; EPI_ISL_757384; EPI_ISL_413604; EPI_ISL_414644; EPI_ISL_414643; EPI_ISL_757325; EPI_ISL_732567; EPI_ISL_737036; EPI_ISL_756166; EPI_ISL_756022; EPI_ISL_756119; EPI_ISL_759946; EPI_ISL_737071; EPI_ISL_756102; EPI_ISL_759808; EPI_ISL_757347; EPI_ISL_757348; EPI_ISL_756095; EPI_ISL_756064; EPI_ISL_755951; EPI_ISL_756080; EPI_ISL_759814; EPI_ISL_759811; EPI_ISL_757383; EPI_ISL_756248; EPI_ISL_732633; EPI_ISL_732611; EPI_ISL_732632; EPI_ISL_757294; EPI_ISL_732580; EPI_ISL_732583; EPI_ISL_732570; EPI_ISL_757311; EPI_ISL_732617; EPI_ISL_757315; EPI_ISL_757314; EPI_ISL_732597; EPI_ISL_757300; EPI_ISL_757340; EPI_ISL_732620; EPI_ISL_732600; EPI_ISL_732577; EPI_ISL_757334; EPI_ISL_732619; EPI_ISL_757308; EPI_ISL_757301; EPI_ISL_757310; EPI_ISL_757309; EPI_ISL_732576; EPI_ISL_732610; EPI_ISL_732616; EPI_ISL_732604; EPI_ISL_757332; EPI_ISL_757339; EPI_ISL_732593; EPI_ISL_732578; EPI_ISL_732598; EPI_ISL_757293; EPI_ISL_732608; EPI_ISL_732574; EPI_ISL_757298; EPI_ISL_757319; EPI_ISL_757299; EPI_ISL_757322; EPI_ISL_759830; EPI_ISL_756000; EPI_ISL_759927; EPI_ISL_756282; EPI_ISL_759812; EPI_ISL_757377; EPI_ISL_481550; EPI_ISL_481720; EPI_ISL_481657; EPI_ISL_481678; EPI_ISL_481641; EPI_ISL_481663; EPI_ISL_481700; EPI_ISL_481675; EPI_ISL_481677; EPI_ISL_481719; EPI_ISL_481727; EPI_ISL_481643; EPI_ISL_481679; EPI_ISL_481721; EPI_ISL_481695; EPI_ISL_481715; EPI_ISL_481537; EPI_ISL_756247; EPI_ISL_757379; EPI_ISL_737211; EPI_ISL_757378; EPI_ISL_481656; EPI_ISL_481645; EPI_ISL_481672; EPI_ISL_481664; EPI_ISL_481667; EPI_ISL_481733; EPI_ISL_481543; EPI_ISL_481562; EPI_ISL_481565; EPI_ISL_481693; EPI_ISL_481522; EPI_ISL_481563; EPI_ISL_481539; EPI_ISL_481583; EPI_ISL_481577; EPI_ISL_481607; EPI_ISL_481608; EPI_ISL_481617; EPI_ISL_481614; EPI_ISL_481620; EPI_ISL_481618; EPI_ISL_481624; EPI_ISL_481674; EPI_ISL_481703; EPI_ISL_481696; EPI_ISL_481697; EPI_ISL_481729; EPI_ISL_481536; EPI_ISL_414646; EPI_ISL_737069; EPI_ISL_418391; EPI_ISL_481692; EPI_ISL_481623; EPI_ISL_757329; EPI_ISL_759805; EPI_ISL_756251; EPI_ISL_759817; EPI_ISL_481658; EPI_ISL_757373; EPI_ISL_759772 | Department of Virology and Immunology, University of Helsinki and Helsinki University Hospital, Huslab Finland | Department of Virology, Faculty of Medicine, University of Helsinki, Helsinki, Finland | Teemu Smura et al A |
| EPI_ISL_757285; EPI_ISL_757286 | Department of Virology, Public Health Laboratories Division | Department of Virology, Public Health Laboratories Division | Massab Umair et al |
| EPI_ISL_468163; EPI_ISL_468161 | Department of Virology, Public Health Laboratories Division, National Institute of Health | Department of Virology, Public Health Laboratories Division, National Institute of Health | Massab Umair et al |
| EPI_ISL_669035; EPI_ISL_670220; EPI_ISL_670250; EPI_ISL_671112; EPI_ISL_714191; EPI_ISL_669500; EPI_ISL_670967; EPI_ISL_748638; EPI_ISL_749055; EPI_ISL_670798; EPI_ISL_669297; EPI_ISL_711430; EPI_ISL_711741; EPI_ISL_682358; EPI_ISL_713619; EPI_ISL_714267; EPI_ISL_714266; EPI_ISL_714265; EPI_ISL_682380; EPI_ISL_669549; EPI_ISL_670529; EPI_ISL_749312; EPI_ISL_749318; EPI_ISL_749327; EPI_ISL_748298; EPI_ISL_748306; EPI_ISL_750310; EPI_ISL_712261; EPI_ISL_757916; EPI_ISL_758910; EPI_ISL_669555; EPI_ISL_759107; EPI_ISL_759590; EPI_ISL_749572; EPI_ISL_759258; EPI_ISL_750781; EPI_ISL_749112; EPI_ISL_749039; EPI_ISL_748796; EPI_ISL_711938; EPI_ISL_759673; EPI_ISL_750537 | Department of Virus and Microbiological Special Diagnostics, Statens Serum Institut, Copenhagen, Denmark | Albertsen Lab, Department of Chemistry and Bioscience, Aalborg University, Denmark | Danish Covid-19 Genome Consortium et al |
| EPI_ISL_429440; EPI_ISL_429409; EPI_ISL_429516; EPI_ISL_437657; EPI_ISL_429375; EPI_ISL_429376; EPI_ISL_444818; EPI_ISL_429431; EPI_ISL_437013; EPI_ISL_437633; EPI_ISL_436979; EPI_ISL_437642 | Department of Virus and Microbiological Special Diagnostics, Statens Serum Institut, Copenhagen, Denmark, Artillerivej 5, 2300 Copenahgen S | Albertsen lab, Department of Chemistry and Bioscience, Aalborg University, Denmark | Rasmus Kirkegaard et al |
| EPI_ISL_618102; EPI_ISL_614793; EPI_ISL_614826; EPI_ISL_614807; EPI_ISL_614802; EPI_ISL_614829; EPI_ISL_614812; EPI_ISL_617982; EPI_ISL_618403; EPI_ISL_618404; EPI_ISL_618304; EPI_ISL_618287; EPI_ISL_622608; EPI_ISL_616365; EPI_ISL_615542; EPI_ISL_615732; EPI_ISL_616841; EPI_ISL_616495; EPI_ISL_616811; EPI_ISL_616812; EPI_ISL_616698; EPI_ISL_616832; EPI_ISL_616589; EPI_ISL_618341; EPI_ISL_617726; EPI_ISL_616604; EPI_ISL_616591; EPI_ISL_616653; EPI_ISL_616631; EPI_ISL_616825; EPI_ISL_615342; EPI_ISL_615343; EPI_ISL_622068; EPI_ISL_615319; EPI_ISL_615357; EPI_ISL_621689; EPI_ISL_621226; EPI_ISL_618269; EPI_ISL_618338; EPI_ISL_616409; EPI_ISL_618402; EPI_ISL_617672; EPI_ISL_622342; EPI_ISL_614489; EPI_ISL_614496; EPI_ISL_622585; EPI_ISL_617949; EPI_ISL_622364; EPI_ISL_622615; EPI_ISL_622617; EPI_ISL_622431; EPI_ISL_617973; EPI_ISL_618133; EPI_ISL_622472; EPI_ISL_614452; EPI_ISL_614510; EPI_ISL_615616; EPI_ISL_618319; EPI_ISL_618693; EPI_ISL_621877; EPI_ISL_621295; EPI_ISL_618096; EPI_ISL_618185; EPI_ISL_618240; EPI_ISL_618184; EPI_ISL_618241; EPI_ISL_618223; EPI_ISL_618230; EPI_ISL_618200; EPI_ISL_617972; EPI_ISL_617996; EPI_ISL_618097; EPI_ISL_622419; EPI_ISL_618127; EPI_ISL_614458; EPI_ISL_614463; EPI_ISL_614850; EPI_ISL_618112; EPI_ISL_618095; EPI_ISL_618109; EPI_ISL_614457; EPI_ISL_622414; EPI_ISL_622556; EPI_ISL_618660; EPI_ISL_618093; EPI_ISL_617284; EPI_ISL_619978; EPI_ISL_617107; EPI_ISL_620138; EPI_ISL_614425; EPI_ISL_614801; EPI_ISL_614828; EPI_ISL_614719; EPI_ISL_614732; EPI_ISL_614730; EPI_ISL_614724; EPI_ISL_614721; EPI_ISL_614722; EPI_ISL_616416; EPI_ISL_618335; EPI_ISL_614775; EPI_ISL_614405; EPI_ISL_614609; EPI_ISL_614446; EPI_ISL_619389; EPI_ISL_619384; EPI_ISL_618626; EPI_ISL_618201; EPI_ISL_618316; EPI_ISL_618225; EPI_ISL_621090; EPI_ISL_617045; EPI_ISL_615424; EPI_ISL_616119; EPI_ISL_616156; EPI_ISL_616320; EPI_ISL_619592; EPI_ISL_618690; EPI_ISL_618839; EPI_ISL_615873; EPI_ISL_615885; EPI_ISL_621901; EPI_ISL_615828; EPI_ISL_614448; EPI_ISL_614776; EPI_ISL_621707; EPI_ISL_618017; EPI_ISL_618018; EPI_ISL_619359; EPI_ISL_619322; EPI_ISL_619820; EPI_ISL_619603; EPI_ISL_617205; EPI_ISL_617377; EPI_ISL_617376; EPI_ISL_618427; EPI_ISL_619400; EPI_ISL_621468; EPI_ISL_619160; EPI_ISL_621760; EPI_ISL_618959; EPI_ISL_618958; EPI_ISL_618897; EPI_ISL_625737 | Department of Virus and Microbiological Special Diagnostics, Statens Serum Institut, Denmark | Albertsen lab, Department of Chemistry and Bioscience, Aalborg University, Denmark | Danish Covid-19 Genome Consortia et al |
| EPI_ISL_416142; EPI_ISL_416140 | Department of Virus and Microbiological Special diagnostics, Statens Serum Institut, Copenhagen, Denmark. | Statens Serum Institute | Morten Rasmussen et al |
| EPI_ISL_416143 | Department of Virus and Microbiological Special diagnostics, Statens Serum Institut, Copenhagen, Denmark. | ViFU | Morten Rasmussen et al |
| EPI_ISL_635166; EPI_ISL_708134; EPI_ISL_759988; EPI_ISL_708135; EPI_ISL_708131 | Dept. of Medical Microbiology, Stavanger University Hospital, Helse Stavanger HF | Norwegian Institute of Public Health, Department of Virology | Kathrine Stene-Johansen et al |
| EPI_ISL_447837 | Dept. of Medical Microbiology, Stavanger University Hospital, Helse Stavanger HF, | Norwegian Institute of Public Health, Department of Virology | Kathrine Stene-Johansen et al |
| EPI_ISL_666604; EPI_ISL_666601; EPI_ISL_666568; EPI_ISL_665255; EPI_ISL_666611; EPI_ISL_665254; EPI_ISL_665259; EPI_ISL_666593; EPI_ISL_666594; EPI_ISL_666615; EPI_ISL_666603; EPI_ISL_665258; EPI_ISL_666614; EPI_ISL_666607; EPI_ISL_664106; EPI_ISL_665256; EPI_ISL_666608; EPI_ISL_666610; EPI_ISL_666598; EPI_ISL_666599; EPI_ISL_666596; EPI_ISL_666595 | Dept. of Microbiology and Infection Control, Akershus University Hospital HF | Dept. of Microbiology and Infection Control, Akershus University Hospital HF | Hege Vangstein Aamot et al |
| EPI_ISL_410531 | Dept. of Pathology, National Institute of Infectious Diseases | Pathogen Genomics Center, National Institute of Infectious Diseases | Tsuyoshi Sekizuka et al A |
| EPI_ISL_410532 | Dept. of Pathology, National Institute of Infectious Diseases | Pathogen Genomics Center, National Institute of Infectious Diseases | Tsuyoshi Sekizuka et al B (https://dx.doi.org/10.2807/1560-7917.ES.2020.25.13.2000305) |

| | | | |
|---|---|---|---|
| EPI_ISL_408666; EPI_ISL_408667; EPI_ISL_408669 | Dept. of Virology III, National Institute of Infectious Diseases | Pathogen Genomics Center, National Institute of Infectious Diseases | Tsuyoshi Sekizuka et al A |
| EPI_ISL_408665 | Dept. of Virology III, National Institute of Infectious Diseases | Pathogen Genomics Center, National Institute of Infectious Diseases | Tsuyoshi Sekizuka et al B (https://dx.doi.org/10.2807/1560-7917.ES.2020.25.13.2000305) |
| EPI_ISL_459913 | Devki Devi Foundation, a unit of Max Healthcare | CSIR-IGIB/Max | Rajesh Pandey# et al |
| EPI_ISL_437197; EPI_ISL_437198; EPI_ISL_437201 | Diagnostic- and Research Institute of Pathology, Medical University of Graz | Diagnostic- and Research Institute of Pathology, Medical University of Graz | Karl Kashofer et al (https://www.biorxiv.org/content/10.1101/2020.07.15.204339v1) |
| EPI_ISL_763075; EPI_ISL_763074 | Diagnosticos da America - DASA | Instituto Adolfo Lutz, Interdiciplinary Procedures Center, Strategic Laboratory | Claudio Tavares Sacchi et al B |
| EPI_ISL_754236 | Diagnósticos da América - DASA | Instituto de Medicina Tropical Universidade de São Paulo | Brazil-UK Centre for Arbovirus Discovery Diagnosis Genomics et al |
| EPI_ISL_528753 | Dinkes Kota Bogor | School of Life Sciences and Technology & School of Pharmacy-Institut Teknologi Bandung; Molecular Genetics Laboratory-Faculty of Medicine-Universitas Padjadjaran; Laboratorium Kesehatan Provinsi Jawa Barat | Azzania Fibriani et al |
| EPI_ISL_528748 | Dinkes Provinsi Jawa Barat | School of Life Sciences and Technology & School of Pharmacy-Institut Teknologi Bandung; Molecular Genetics Laboratory-Faculty of Medicine-Universitas Padjadjaran; Laboratorium Kesehatan Provinsi Jawa Barat | Azzania Fibriani et al |
| EPI_ISL_467374 | Dinkes Samarinda | Eijkman Institute for Molecular Biology, Ministry of Research and Technology/National Agency for Research and Innovation | Edison Johar et al |
| EPI_ISL_591329; EPI_ISL_591330; EPI_ISL_591334; EPI_ISL_591338 | Dipartimento di Biotecnologie Mediche, University of Siena | Dipartimento di Biotecnologie Mediche, University of Siena | Cusi et al |
| EPI_ISL_722896; EPI_ISL_722872; EPI_ISL_722858; EPI_ISL_722856; EPI_ISL_722857; EPI_ISL_722855 | Dipartimento di Scienze Biomediche e Oncologia Umana - Azienda Ospedaliero Universitaria Consorziale Policlinico | Istituto Zooprofilattico Sperimentale della Puglia e della Basilicata | Parisi A. et al |
| EPI_ISL_760127; EPI_ISL_760126; EPI_ISL_747271; EPI_ISL_747343; EPI_ISL_747340; EPI_ISL_747345; EPI_ISL_760153; EPI_ISL_747420; EPI_ISL_747421; EPI_ISL_747336; EPI_ISL_760133; EPI_ISL_760134; EPI_ISL_760064; EPI_ISL_760135; EPI_ISL_760131; EPI_ISL_747342; EPI_ISL_747261; EPI_ISL_760190; EPI_ISL_760018; EPI_ISL_760007; EPI_ISL_760020; EPI_ISL_760012; EPI_ISL_747262; EPI_ISL_760195; EPI_ISL_747278; EPI_ISL_760212; EPI_ISL_747452; EPI_ISL_760193; EPI_ISL_760226; EPI_ISL_760145; EPI_ISL_760148; EPI_ISL_760149; EPI_ISL_760156; EPI_ISL_762992; EPI_ISL_760157; EPI_ISL_760139; EPI_ISL_760142; EPI_ISL_760137; EPI_ISL_760143; EPI_ISL_760141; EPI_ISL_760144; EPI_ISL_760140; EPI_ISL_760138 | Division of Emerging Infectious Diseases, Bureau of Infectious Diseases Diagnosis Control, Korea Disease Control and Prevention Agency | Division of Emerging Infectious Diseases, Bureau of Infectious Diseases Diagnosis Control, Korea Disease Control and Prevention Agency | Ae Kyung Park et al |
| EPI_ISL_738141; EPI_ISL_738139; EPI_ISL_738142 | Division of Emerging Infectious Diseases, Bureau of Infectious Diseases Diagnosis Control, Korea Disease Control and Prevention Agency | Division of Emerging Infectious Diseases, Bureau of Infectious Diseases Diagnosis Control, Korea Disease Control and Prevention Agency | Jeong-Min Kim et al A |
| EPI_ISL_509222; EPI_ISL_508867 | Division of Infectious Diseases and Hospital Epidemiology, University Hospital Zürich | Institute of Medical Virology, University of Zurich | Maryam Zaheri et al |
| EPI_ISL_413024; EPI_ISL_413022 | Division of Infectious Diseases, University Hospital Zurich | Institute of Medical Virology, University of Zurich | Stefan Schmutz et al B (https://dx.doi.org/10.1002/jmv.25773) |
| EPI_ISL_497962; EPI_ISL_425118; EPI_ISL_497963; EPI_ISL_412871; EPI_ISL_412870; EPI_ISL_510627; EPI_ISL_510636; EPI_ISL_522490; EPI_ISL_522500; EPI_ISL_522501; EPI_ISL_425117; EPI_ISL_426169; EPI_ISL_426166; EPI_ISL_412872; EPI_ISL_426164; EPI_ISL_471438; EPI_ISL_497960; EPI_ISL_471449; EPI_ISL_497990; EPI_ISL_471442; EPI_ISL_498033; EPI_ISL_506991; EPI_ISL_498030; EPI_ISL_498031; EPI_ISL_498035; EPI_ISL_498034; EPI_ISL_506972; EPI_ISL_506980; EPI_ISL_506994; EPI_ISL_506992; EPI_ISL_506993; EPI_ISL_514916; EPI_ISL_514942; EPI_ISL_522539; EPI_ISL_522541; EPI_ISL_522527; EPI_ISL_514939; EPI_ISL_514928; EPI_ISL_522454; EPI_ISL_514938; EPI_ISL_514940; EPI_ISL_514923; EPI_ISL_506995; EPI_ISL_498053; EPI_ISL_506966; EPI_ISL_498026; EPI_ISL_506988; EPI_ISL_506989 | Division of Viral Diseases, Center for Laboratory Control of Infectious Diseases, Korea Centers for Diseases Control and Prevention | Division of Viral Diseases, Center for Laboratory Control of Infectious Diseases, Korea Centers for Diseases Control and Prevention | Jeong-Min Kim et al A |
| EPI_ISL_412869 | Division of Viral Diseases, Center for Laboratory Control of Infectious Diseases, Korea Centers for Diseases Control and Prevention | Division of Viral Diseases, Center for Laboratory Control of Infectious Diseases, Korea Centers for Diseases Control and Prevention | Jeong-Min Kim et al B (https://dx.doi.org/10.2807/1560-7917.ES.2020.25.13.2000305) |
| EPI_ISL_722209 | Dom Zdravlja Sarajevo | Alea Genetic Center | Salihefendic L. et al |
| EPI_ISL_700517; EPI_ISL_700582; EPI_ISL_700592 | Dr Abdurahman CDC wc DAC | NHLS/UCT | Arash Iranzadeh et al |
| EPI_ISL_745139; EPI_ISL_745138 | Dr Van Niekerk Hospital | National Health Laboratory Service (NHLS), Tygerberg | Susan Engelbrecht et al |
| EPI_ISL_479627 | Dr. Georges-L.-Dumont University Hospital Centre | National Microbiology Laboratory | Anna Majer et al |
| EPI_ISL_583763; EPI_ISL_583739; EPI_ISL_583740; EPI_ISL_583752; EPI_ISL_583747; EPI_ISL_583848; EPI_ISL_583764 | Dr. Gernot Walder GmbH | Bergthaler laboratory, CeMM Research Center for Molecular Medicine of the Austrian Academy of Sciences | Alexandra Popa et al A |
| EPI_ISL_528752 | Dr. H. A. Rotinsulu Lung Hospital | School of Pharmacy & School of Life Sciences and Technology - Institut Teknologi Bandung; Molecular Genetics Laboratory-Faculty of Medicine-Universitas Padjadjaran; Laboratorium Kesehatan Provinsi Jawa Barat | Catur Riani et al |
| EPI_ISL_461101; EPI_ISL_632688; EPI_ISL_722748; EPI_ISL_577861; EPI_ISL_722593; EPI_ISL_722497; EPI_ISL_722766; EPI_ISL_722783; EPI_ISL_523549; EPI_ISL_455147; EPI_ISL_455143; EPI_ISL_455167; EPI_ISL_523606; EPI_ISL_523611; EPI_ISL_523662; EPI_ISL_523409; EPI_ISL_523737; EPI_ISL_523748; EPI_ISL_523740; EPI_ISL_523744; EPI_ISL_523745; EPI_ISL_523735; EPI_ISL_523741; EPI_ISL_523738; EPI_ISL_523739; EPI_ISL_523742; EPI_ISL_523743; EPI_ISL_523736; EPI_ISL_523723; EPI_ISL_523541; EPI_ISL_523454; EPI_ISL_574780; EPI_ISL_461114; EPI_ISL_523272; EPI_ISL_763099; EPI_ISL_523199; EPI_ISL_523625; EPI_ISL_523616; EPI_ISL_523618; EPI_ISL_523280; EPI_ISL_632543; EPI_ISL_577892; EPI_ISL_577886; EPI_ISL_577857; EPI_ISL_632579; EPI_ISL_632580; EPI_ISL_632577; EPI_ISL_632388; EPI_ISL_632583; EPI_ISL_632581; EPI_ISL_523598; EPI_ISL_523633; EPI_ISL_523622; EPI_ISL_523630; EPI_ISL_523629; EPI_ISL_523631; EPI_ISL_523628; EPI_ISL_523640; EPI_ISL_523636; EPI_ISL_523637; EPI_ISL_523638; EPI_ISL_523597; EPI_ISL_523649; EPI_ISL_523647; EPI_ISL_523648; EPI_ISL_523785; EPI_ISL_461247; EPI_ISL_422924; EPI_ISL_460965; EPI_ISL_460972; EPI_ISL_460968; EPI_ISL_460885; EPI_ISL_460998; EPI_ISL_460919; EPI_ISL_461258; EPI_ISL_523182; EPI_ISL_523258; EPI_ISL_422579; EPI_ISL_461279; EPI_ISL_455159; EPI_ISL_455289; EPI_ISL_461119; EPI_ISL_460915; EPI_ISL_523179; EPI_ISL_460944; EPI_ISL_455177; EPI_ISL_455166; EPI_ISL_461072; EPI_ISL_523177; EPI_ISL_523466; EPI_ISL_461323; EPI_ISL_523196; EPI_ISL_523212; EPI_ISL_523210; EPI_ISL_523508; EPI_ISL_523211; EPI_ISL_523440; EPI_ISL_461209; EPI_ISL_461011; EPI_ISL_461274; EPI_ISL_461187; EPI_ISL_523548; EPI_ISL_577900; EPI_ISL_577898; EPI_ISL_461310; EPI_ISL_461368; EPI_ISL_523460; EPI_ISL_523438; EPI_ISL_763217; EPI_ISL_763252; EPI_ISL_722800; EPI_ISL_722548; EPI_ISL_632697; EPI_ISL_632690; EPI_ISL_523775; EPI_ISL_577943; EPI_ISL_722714; EPI_ISL_578064; EPI_ISL_578056; EPI_ISL_632648; EPI_ISL_632651; EPI_ISL_632652; EPI_ISL_763227; EPI_ISL_722348; EPI_ISL_577856; EPI_ISL_578047; EPI_ISL_523773; EPI_ISL_763308; EPI_ISL_763245; EPI_ISL_577902; EPI_ISL_632503; EPI_ISL_763119; EPI_ISL_763103; EPI_ISL_763110; EPI_ISL_763118; EPI_ISL_763116; EPI_ISL_763120; EPI_ISL_763101; EPI_ISL_763112; EPI_ISL_763121; EPI_ISL_763102; EPI_ISL_632553; EPI_ISL_578015; EPI_ISL_577979; EPI_ISL_632387; EPI_ISL_632501; EPI_ISL_577997; EPI_ISL_577996; EPI_ISL_577998; EPI_ISL_632605; EPI_ISL_632604; EPI_ISL_632606; EPI_ISL_763109; EPI_ISL_722717; EPI_ISL_722628; EPI_ISL_577926; EPI_ISL_574786; EPI_ISL_722581; EPI_ISL_763266 | Dutch COVID-19 response team | Erasmus Medical Center | Bas Oude Munnink et al |
| EPI_ISL_414426; EPI_ISL_414429; EPI_ISL_415498; EPI_ISL_415487; EPI_ISL_415460; EPI_ISL_415466; EPI_ISL_415515; EPI_ISL_415465; EPI_ISL_415469; EPI_ISL_415535 | Dutch COVID-19 response team | Erasmus Medical Center | David Nieuwenhuijse et al A (https://dx.doi.org/10.1038/s41591-020-0997-y) |
| EPI_ISL_547453; EPI_ISL_454752; EPI_ISL_636504; EPI_ISL_454794; EPI_ISL_636493; EPI_ISL_636492; EPI_ISL_547451; EPI_ISL_547450; EPI_ISL_547452; EPI_ISL_636541; EPI_ISL_547445; EPI_ISL_547449; EPI_ISL_723372; EPI_ISL_636550; EPI_ISL_723416; EPI_ISL_723408; EPI_ISL_454754; EPI_ISL_547499; EPI_ISL_454777; EPI_ISL_454751; EPI_ISL_454750; EPI_ISL_454762; EPI_ISL_723385; EPI_ISL_728741; EPI_ISL_547520; EPI_ISL_454753; EPI_ISL_636517; EPI_ISL_547518; EPI_ISL_454778; EPI_ISL_547448; EPI_ISL_547446; EPI_ISL_547447; EPI_ISL_636526; EPI_ISL_636516; EPI_ISL_636515; EPI_ISL_636520; EPI_ISL_636519; EPI_ISL_636514; EPI_ISL_636518; EPI_ISL_636521; EPI_ISL_636513; EPI_ISL_728668; EPI_ISL_636578; EPI_ISL_723240; EPI_ISL_636534; EPI_ISL_636535; EPI_ISL_728586; EPI_ISL_636568; EPI_ISL_723253; EPI_ISL_728648; EPI_ISL_728621; EPI_ISL_728645; EPI_ISL_728584; EPI_ISL_723238; EPI_ISL_728577; EPI_ISL_723229; EPI_ISL_728635; EPI_ISL_728692; EPI_ISL_728691; EPI_ISL_636537 | Dutch COVID-19 response team | National Institute for Public Health and the Environment (RIVM) | Adam Meijer et al |
| EPI_ISL_421656; EPI_ISL_426285; EPI_ISL_492997; EPI_ISL_501839; EPI_ISL_492998; EPI_ISL_512646; EPI_ISL_501936; EPI_ISL_501833; EPI_ISL_534203; EPI_ISL_515188; EPI_ISL_515194; EPI_ISL_515193; EPI_ISL_515187; EPI_ISL_515195; EPI_ISL_515192; EPI_ISL_515191; EPI_ISL_534199; EPI_ISL_515189; EPI_ISL_534221; EPI_ISL_450518; EPI_ISL_512313; EPI_ISL_639667; EPI_ISL_639674; EPI_ISL_639653; EPI_ISL_639666; EPI_ISL_492996; EPI_ISL_639690; EPI_ISL_501829; EPI_ISL_501849; EPI_ISL_426286; EPI_ISL_639693; EPI_ISL_639687; EPI_ISL_639669; EPI_ISL_639692; EPI_ISL_639694; EPI_ISL_639671; EPI_ISL_639644; EPI_ISL_639646; EPI_ISL_639668; EPI_ISL_639682; EPI_ISL_639688; EPI_ISL_639643; EPI_ISL_426289; EPI_ISL_639637; EPI_ISL_639664; EPI_ISL_639689; EPI_ISL_639681; EPI_ISL_639684; EPI_ISL_639691; EPI_ISL_639685; EPI_ISL_421655; EPI_ISL_639649; EPI_ISL_421653; EPI_ISL_426288; EPI_ISL_450519; EPI_ISL_639686 | E. Gulbja Laboratorija | Latvian Biomedical Research and Study Centre | Ivars Silamiķelis et al |
| EPI_ISL_486438 | E. Gulbja laboratorija | Latvian Biomedical Research and Study Centre | Ivars Silamiķelis et al |
| EPI_ISL_434540 | EBAIS Concepción Norte | Inciensa, Instituto Costarricense de Investigación y Enseñanza en Nutrición y Salud | Duarte et al |
| EPI_ISL_745198 | ESI Hospital | CSIR-Centre for Cellular and Molecular Biology | Dr.V.Sudha Rani et al |
| EPI_ISL_593517; EPI_ISL_593491; EPI_ISL_593538; EPI_ISL_593540; EPI_ISL_593532; EPI_ISL_593515 | Eastern Ontario Regional Laboratory Association | McMaster University | Leanne Mortimer et al |
| EPI_ISL_745137 | Edameni Clinic | National Health Laboratory Service (NHLS), Tygerberg | Susan Engelbrecht et al |
| EPI_ISL_529142; EPI_ISL_478672; EPI_ISL_524426; EPI_ISL_529144; EPI_ISL_529143; EPI_ISL_524427; EPI_ISL_529141; EPI_ISL_477161; EPI_ISL_475723; EPI_ISL_475724; EPI_ISL_468053; EPI_ISL_468045; EPI_ISL_468060; EPI_ISL_468056; EPI_ISL_529145 | Egyptian National Cancer Institute (ENCI) | Egyptian National Cancer Institute (ENCI) | Zekri et al |
| EPI_ISL_469275 | Egyptian National Cancer Institute (ENCI) | Human Genome Center | Zekri et al |
| EPI_ISL_710611 | Ektorps Vardcentral | The Public Health Agency of Sweden | Department of Microbiology et al |
| EPI_ISL_745110 | Elsies River CHC wc ERP | National Health Laboratory Service (NHLS), Tygerberg | Susan Engelbrecht et al |
| EPI_ISL_486854 | Emergency County Hospital Suceava | Stefan cel Mare, University Metagenomics lab | Lobiuc Andrei et al. et al |
| EPI_ISL_486855 | Emergency county Hospital Suceava | Stefan cel Mare University Metagenomics Lab | Lobiuc Andrei et al. et al |
| EPI_ISL_648168; EPI_ISL_648166; EPI_ISL_648167 | Eskilstuna | The Public Health Agency of Sweden | Anna-Malin Linde et al |
| EPI_ISL_602302; EPI_ISL_602288; EPI_ISL_602289; EPI_ISL_602284; EPI_ISL_602282; EPI_ISL_602300; EPI_ISL_602297; EPI_ISL_602293; EPI_ISL_602294; EPI_ISL_602298; EPI_ISL_602286; EPI_ISL_602295; EPI_ISL_602296; EPI_ISL_602303; EPI_ISL_602287; EPI_ISL_602290; EPI_ISL_602292; EPI_ISL_602285; EPI_ISL_602301; EPI_ISL_602283; EPI_ISL_602299; EPI_ISL_602291 | Evangelisches Klinikum Bethel, Institut für Laboratoriumsmedizin, Mikrobiologie und Hygiene | Bielefeld University | David Brandt et al |
| EPI_ISL_548968 | Expo2020 Emergency Center | Agiomix | Walaa Allam et al |
| EPI_ISL_428851 | FSBSI "Chumakov Federal Scientific Center for Research and Development of Immune-and-Biological Products of Russian Academy of Sciences" | FSBSI "Chumakov Federal Scientific Center for Research and Development of Immune-and-Biological Products of Russian Academy of Sciences" & NRC "Kurchatov institute" | Liubov Kozlovskaya et al |
| EPI_ISL_468751 | Facultad de Medicina UC | Center for Mathematical Modeling and Center for Genome Regulation. Santiago, Chile | Gaete A et al |
| EPI_ISL_752605 | Faculty of Medicine, Al-Quds University | Faculty of Medicine, Al-Quds University | Ereqat et al |
| EPI_ISL_451970; EPI_ISL_451966 | Federal Budget Institution of Science, State Research Center for Applied Microbiology & Biotechnology | Federal Budget Institution of Science, State Research Center for Applied Microbiology & Biotechnology | Dyatlov I et al |
| EPI_ISL_526583; EPI_ISL_613892; EPI_ISL_653263 | Florida Bureau of Public Health Laboratories | Florida Bureau of Public Health Laboratories | Sarah Schmedes et al |
| EPI_ISL_541264 | Florida Bureau of Public Health Laboratories, Florida Department of Health | Florida Bureau of Public Health Laboratories, Florida Department of Health | Schmedes et al |
| EPI_ISL_738313; EPI_ISL_470882; EPI_ISL_708177; EPI_ISL_635065 | Foerde Hospital, Department of Microbiology | Norwegian Institute of Public Health, Department of Virology | Kathrine Stene-Johansen et al |
| EPI_ISL_476139 | Folkhalsomyndigheten | The Public Health Agency of Sweden | Oskar Karlsson Lindsjo et al |
| EPI_ISL_581455; EPI_ISL_581492; EPI_ISL_581493; EPI_ISL_581489; EPI_ISL_581491; EPI_ISL_581487; EPI_ISL_581490 | Fondation Congolaise pour la recherche medicale (FCRM) | NGS Competence Center Tübingen, Institut für Medizinische Mikrobiologie und Hygiene, Universitätsklinikum Tübingen | Angel Angelov et al |
| EPI_ISL_420145 | Forde Hospital Department of Microbiology | Norwegian Institute of Public Health, Department of Virology | Kathrine Stene-Johansen et al |

| | | | |
|---|---|---|---|
| EPI_ISL_413586 | Foundation Elisabeth-Tweesteden Ziekenhuis | Erasmus Medical Center | David Nieuwenhuijse et al A (https://dx.doi.org/10.1038/s41591-020-0997-y) |
| EPI_ISL_413565 | Foundation Pamm | Erasmus Medical Center | David Nieuwenhuijse et al A (https://dx.doi.org/10.1038/s41591-020-0997-y) |
| EPI_ISL_431779 | Fujian Center for Disease Control and Prevention | Fujian Center for Disease Control and Prevention | Lin Qi et al |
| EPI_ISL_707928 | Fujita Health University Hospital | Fujita Health University School of Medicine, Department of Microbiology | Masahiro Suzuki et al |
| EPI_ISL_728000 | Fujita Health University Okazaki Medical Center | Department of Virology and Parasitology, Fujita Health University School of Medicin | Takayuki Murata et al |
| EPI_ISL_718273 | Fujita Health University Okazaki Medical Center | Department of Virology and Parasitology, Fujita Health University School of Medicine | Takayuki Murata et al |
| EPI_ISL_728208 | Fujita Health University Okazaki Medical Center | Fujita health University, Department of Microbiology | Takayuki Murata et al |
| EPI_ISL_549132; EPI_ISL_590903; EPI_ISL_759985; EPI_ISL_759984; EPI_ISL_549054; EPI_ISL_549055; EPI_ISL_549053; EPI_ISL_708059; EPI_ISL_549088; EPI_ISL_549048; EPI_ISL_549168; EPI_ISL_449788; EPI_ISL_449787; EPI_ISL_549078; EPI_ISL_549120; EPI_ISL_549077; EPI_ISL_549079; EPI_ISL_549060; EPI_ISL_549112; EPI_ISL_549058; EPI_ISL_549082; EPI_ISL_493361; EPI_ISL_493362; EPI_ISL_420313; EPI_ISL_420146; EPI_ISL_493375; EPI_ISL_493371; EPI_ISL_493367; EPI_ISL_493359; EPI_ISL_493358; EPI_ISL_493369; EPI_ISL_549124; EPI_ISL_549069; EPI_ISL_549027 | Furst Medical Laboratory | Norwegian Institute of Public Health, Department of Virology | Kathrine Stene-Johansen et al |
| EPI_ISL_524755 | GMERS Medical College and Hospital, Dharpur, Patan | Gujarat Biotechnology Research Centre | Nidhi Patel et al |
| EPI_ISL_648140; EPI_ISL_648163; EPI_ISL_582834; EPI_ISL_648141 | Gavle klinisk mikrobiologi | The Public Health Agency of Sweden | Anna-Malin Linde et al |
| EPI_ISL_475110 | Gavle klinisk mikrobiologi | The Public Health Agency of Sweden | Oskar Karlsson Lindsjo et al |
| EPI_ISL_730577; EPI_ISL_730570; EPI_ISL_730573; EPI_ISL_730567; EPI_ISL_730568; EPI_ISL_730620; EPI_ISL_730574; EPI_ISL_730575; EPI_ISL_730569; EPI_ISL_730571; EPI_ISL_730572 | Gazi University Faculty of Medicine, Medical Virology Laboratory | Gazi University Faculty of Medicine, Medical Virology Laboratory | Erdem Şahin et al |
| EPI_ISL_467300 | General Hospital "Abdulah Nakas" | Alea Genetic Center | Rijad Konjhodzic et al (https://dx.doi.org/10.1101/2020.06.19.160606) |
| EPI_ISL_516413; EPI_ISL_677707; EPI_ISL_677705; EPI_ISL_678252 | General Hospital - Kumanovo | Research Center for Genetic Engineering and Biotechnology "Georgi D. Efremov" , Macedonian Academy of Sciences and Arts | RCGEB - MASA et al |
| EPI_ISL_678256; EPI_ISL_677722; EPI_ISL_677726; EPI_ISL_678255; EPI_ISL_677725; EPI_ISL_677704; EPI_ISL_677709 | General Hospital - Ohrid | Research Center for Genetic Engineering and Biotechnology "Georgi D. Efremov" , Macedonian Academy of Sciences and Arts | RCGEB - MASA et al |
| EPI_ISL_514354; EPI_ISL_678258; EPI_ISL_677703; EPI_ISL_677674 | General Hospital - Prilep | Research Center for Genetic Engineering and Biotechnology "Georgi D. Efremov" , Macedonian Academy of Sciences and Arts | RCGEB - MASA et al |
| EPI_ISL_678260; EPI_ISL_677719 | General Hospital - Struga | Research Center for Genetic Engineering and Biotechnology "Georgi D. Efremov" , Macedonian Academy of Sciences and Arts | RCGEB - MASA et al |
| EPI_ISL_678254 | General Hospital - Strumica | Research Center for Genetic Engineering and Biotechnology "Georgi D. Efremov" , Macedonian Academy of Sciences and Arts | RCGEB - MASA et al |
| EPI_ISL_677675 | General Hospital - Veles | Research Center for Genetic Engineering and Biotechnology "Georgi D. Efremov" , Macedonian Academy of Sciences and Arts | RCGEB - MASA et al |
| EPI_ISL_406798 | General Hospital of Central Theater Command of People's Liberation Army of China | BGI & Institute of Microbiology, Chinese Academy of Sciences & Shandong First Medical University & Shandong Academy of Medical Sciences & General Hospital of Central Theater Command of People's Liberation Army of China | Weijun Chen et al (https://dx.doi.org/10.1016/S0140-6736(20)30251-8) |
| EPI_ISL_586547 | General Hospital,Godhra | Gujarat Biotechnology Research Centre | Pinal Gandhi et al |
| EPI_ISL_614283; EPI_ISL_614285 | General practitioner | National Reference Center for Viruses of Respiratory Infections, Institut Pasteur, Paris | Marion Barbet et al B |
| EPI_ISL_754392 | Genetica Molecular and Subdepartamento de Virologia ISP Chile | Insituto de Salud Publica de Chile | Javier Tognarelli et al |
| EPI_ISL_746663; EPI_ISL_746568; EPI_ISL_746632; EPI_ISL_746810; EPI_ISL_746487; EPI_ISL_746488; EPI_ISL_746794; EPI_ISL_746822; EPI_ISL_746535; EPI_ISL_746534; EPI_ISL_746805; EPI_ISL_746814; EPI_ISL_746775; EPI_ISL_746791; EPI_ISL_746788; EPI_ISL_746494; EPI_ISL_746678; EPI_ISL_746712; EPI_ISL_746741; EPI_ISL_746608; EPI_ISL_746552; EPI_ISL_746617; EPI_ISL_746566; EPI_ISL_746630; EPI_ISL_746547; EPI_ISL_746611; EPI_ISL_746575; EPI_ISL_746637; EPI_ISL_746763; EPI_ISL_746715; EPI_ISL_746716; EPI_ISL_746717; EPI_ISL_746486; EPI_ISL_746591; EPI_ISL_746677; EPI_ISL_746719; EPI_ISL_746764; EPI_ISL_746576; EPI_ISL_746638; EPI_ISL_746679; EPI_ISL_746718; EPI_ISL_746720; EPI_ISL_746582; EPI_ISL_746643; EPI_ISL_746564; EPI_ISL_746626; EPI_ISL_746766; EPI_ISL_746770; EPI_ISL_746536; EPI_ISL_746504; EPI_ISL_746786; EPI_ISL_746606; EPI_ISL_746773 | Genetica Molecular and Subdepartamento de Virologia ISP Chile | Instituto de Salud Publica de Chile | Javier Tognarelli et al |
| EPI_ISL_605783 | Genome Center | Genome Center | Md. Shazid Hasan et al |
| EPI_ISL_480256; EPI_ISL_480246; EPI_ISL_735275; EPI_ISL_480276; EPI_ISL_735344; EPI_ISL_735289; EPI_ISL_735301; EPI_ISL_735320; EPI_ISL_480252; EPI_ISL_735352; EPI_ISL_480228; EPI_ISL_480235; EPI_ISL_735368; EPI_ISL_480247; EPI_ISL_735273; EPI_ISL_480231; EPI_ISL_735372; EPI_ISL_480265; EPI_ISL_735332; EPI_ISL_735324; EPI_ISL_735263; EPI_ISL_735279; EPI_ISL_735330; EPI_ISL_735303; EPI_ISL_735306; EPI_ISL_735346; EPI_ISL_735347; EPI_ISL_735284; EPI_ISL_480277; EPI_ISL_735366; EPI_ISL_735367; EPI_ISL_735373; EPI_ISL_480245; EPI_ISL_735342; EPI_ISL_735350; EPI_ISL_735351; EPI_ISL_735362; EPI_ISL_735359; EPI_ISL_735355; EPI_ISL_735360; EPI_ISL_735364; EPI_ISL_735361; EPI_ISL_735357; EPI_ISL_735365 | Genomic Laboratory (GLAB) (Conjoint lab of Health Directorate of Istanbul and Istanbul Technical University) | Genomic Laboratory (GLAB), Istanbul Technical University | Ilker Karacan et al A |
| EPI_ISL_427391; EPI_ISL_428346 | Genomic Laboratory (GLAB) (Conjoint lab of Health Directorate of Istanbul and Istanbul Technical University) | Genomic Laboratory (GLAB), Istanbul Technical University | Ilker Karacan et al B (https://dx.doi.org/10.1101/2020.05.15.095794) |
| EPI_ISL_632908 | Genomic Sciences, Rehman Medical Institute | Genomic Sciences, Rehman Medical Institute | Ali et al |
| EPI_ISL_730226; EPI_ISL_730216; EPI_ISL_730218; EPI_ISL_730207; EPI_ISL_730212 | Genomica Lab Molecular, M√©xico | Andersen lab at Scripps Research | SEARCH Alliance San Diego with Jonathan Gonzalez Garcia et al |
| EPI_ISL_539557 | Gerencia del área de salud de Badajoz, Llerena y Zafra | Instituto de Salud Carlos III | Iglesias-Caballero et al A |
| EPI_ISL_452218; EPI_ISL_452221; EPI_ISL_452220; EPI_ISL_452222; EPI_ISL_452219; EPI_ISL_452223 | Goethe University Hospital Frankfurt | Institute for Medical Virology, Goethe University Hospital Frankfurt | Tuna Toptan et al |
| EPI_ISL_496605; EPI_ISL_496606; EPI_ISL_496736; EPI_ISL_496863; EPI_ISL_496800; EPI_ISL_496848; EPI_ISL_496851; EPI_ISL_496906; EPI_ISL_496861; EPI_ISL_496854; EPI_ISL_496860; EPI_ISL_496609; EPI_ISL_496607; EPI_ISL_496641; EPI_ISL_496829; EPI_ISL_496636; EPI_ISL_496632; EPI_ISL_496670; EPI_ISL_496902; EPI_ISL_496658; EPI_ISL_496612; EPI_ISL_496801; EPI_ISL_496809; EPI_ISL_496645; EPI_ISL_496644; EPI_ISL_496853; EPI_ISL_496724; EPI_ISL_496844; EPI_ISL_496617; EPI_ISL_496628; EPI_ISL_496862 | Gorgas Memorial Laboratory of Health Studies | Gorgas Memorial Laboratory of Health Studies | Danilo Franco et al (https://www.medrxiv.org/content/10.1101/2020.07.31.20160929v1) |
| EPI_ISL_524736 | Government General Hospital, Jam Khambhaliya, Devbhoomi Dwarka | Gujarat Biotechnology Research Centre | Janvi Raval et al |
| EPI_ISL_508217; EPI_ISL_508262 | Government Medical College | National Institute of Biomedical Genomics | Arindam Maitra et al |
| EPI_ISL_640054; EPI_ISL_640040; EPI_ISL_640051; EPI_ISL_640060 | Groote Schuur Hospital wc GSH | NHLS/UCT | Arash Iranzadeh et al |
| EPI_ISL_699137; EPI_ISL_698131; EPI_ISL_699126; EPI_ISL_699132; EPI_ISL_698118; EPI_ISL_698217; EPI_ISL_698305; EPI_ISL_698694; EPI_ISL_699122; EPI_ISL_698121; EPI_ISL_698716; EPI_ISL_698272; EPI_ISL_698476; EPI_ISL_698860; EPI_ISL_698788; EPI_ISL_698425; EPI_ISL_698647; EPI_ISL_699081 | Group 42 (G42) Healthcare, Abu Dhabi, United Arab Emirates; Department of Health, The United Arab Emirates | G42 Healthcare | Rong Liu et al |
| EPI_ISL_403932; EPI_ISL_403933 | Guangdong Provincial Center for Diseases Control and Prevention; Guangdong Provincial Public Health | Department of Microbiology, Guangdong Provincial Center for Diseases Control and Prevention | Min Kang et al A |
| EPI_ISL_403935 | Guangdong Provincial Center for Diseases Control and Prevention; Guangdong Provincial Public Health | Department of Microbiology, Guangdong Provincial Center for Diseases Control and Prevention | Min Kang et al B (https://dx.doi.org/10.2807/1560-7917.ES.2020.25.13.2000305) |
| EPI_ISL_444969 | Guangzhou Eighth People's Hospital (Jiahe Sector) | Institute of Human Virology, Zhongshan School of Medicine, Sun Yat-sen University | Junsong Zhang et al |
| EPI_ISL_509710 | Guatemala Ministry of Public Health | Pathogen Discovery, Respiratory Viruses Branch, Division of Viral Diseases, Centers for Disease Control and Prevention | Jing Zhang et al B |
| EPI_ISL_509701; EPI_ISL_509695; EPI_ISL_509696; EPI_ISL_509699; EPI_ISL_509700; EPI_ISL_509702; EPI_ISL_509703; EPI_ISL_509698; EPI_ISL_509697 | Guatemala Ministry of Public Health | Pathogen Discovery, Respiratory Viruses Branch, Division of Viral Diseases, Centers for Disease Control and Prevention | Ying Tao et al A |
| EPI_ISL_700481; EPI_ISL_700512 | Guguletu CHC wc GDH | NHLS/UCT | Arash Iranzadeh et al |
| EPI_ISL_547627; EPI_ISL_547754 | Gundersen Molecular Diagnostics Laboratory | Kabara Cancer Research Institute | Craig S. Richmond et al |
| EPI_ISL_479898 | Gunma Prefectural Institute of Public Health and Environmental Sciences | Pathogen Genomics Center, National Institute of Infectious Diseases | Tsuyoshi Sekizuka et al C (https://www.medrxiv.org/content/10.1101/2020.07.01.20143958v1) |
| EPI_ISL_745201 | Guntur General Hospital | CSIR-Centre for Cellular and Molecular Biology | Dr.V.Sudha Rani et al |
| EPI_ISL_418021; EPI_ISL_418020 | H Braga | Instituto Nacional de Saude (INSA) | Guiomar et al et al |
| EPI_ISL_421446 | H Guimaraes | Instituto Nacional de Saude (INSA) | Guiomar et al et al |
| EPI_ISL_596366; EPI_ISL_596285; EPI_ISL_596377; EPI_ISL_596277 | HELIX LCC | WHO National Influenza Centre Russian Federation | Andrey Komissarov et al A |
| EPI_ISL_733332; EPI_ISL_733078; EPI_ISL_733451; EPI_ISL_733432; EPI_ISL_733050; EPI_ISL_733127; EPI_ISL_733109; EPI_ISL_733046; EPI_ISL_733051; EPI_ISL_733077; EPI_ISL_733120; EPI_ISL_733139; EPI_ISL_733358; EPI_ISL_602397; EPI_ISL_733111; EPI_ISL_602452; EPI_ISL_733098; EPI_ISL_733439; EPI_ISL_733307; EPI_ISL_733346; EPI_ISL_733354; EPI_ISL_733342; EPI_ISL_733126; EPI_ISL_733420; EPI_ISL_733440; EPI_ISL_733349; EPI_ISL_639953; EPI_ISL_639957; EPI_ISL_602445; EPI_ISL_733452; EPI_ISL_733151; EPI_ISL_733131; EPI_ISL_733421; EPI_ISL_733054; EPI_ISL_733433; EPI_ISL_733052; EPI_ISL_733056; EPI_ISL_602340; EPI_ISL_733060; EPI_ISL_733442; EPI_ISL_733146; EPI_ISL_733361 | HELIX LLC | WHO National Influenza Centre Russian Federation | Andrey Komissarov et al A |
| EPI_ISL_445359 | HOSP. SANTIAGO ORIENTE DR. LUIS TISNE B. | Instituto de Salud Publica de Chile | Andrés E Castillo et al |
| EPI_ISL_682239 | HOSPITAL CIUDAD NEILY | Inciensa, Instituto Costarricense de Investigación y Enseñanza en Nutrición y Salud | Francisco Duarte et al |
| EPI_ISL_445281 | HOSPITAL CLINICO DEL SUR | Instituto de Salud Publica de Chile | Andrés E Castillo et al |
| EPI_ISL_445378 | HOSPITAL DE BULNES | Instituto de Salud Publica de Chile | Andrés E Castillo et al |
| EPI_ISL_418247 | HOSPITAL GENERAL DE SEGOVIA | Instituto de Salud Carlos III | Iglesias-Caballero et al B (https://dx.doi.org/10.1101/2020.04.20.050039) |
| EPI_ISL_445331 | HOSPITAL HERMINDA MARTIN CHILLAN | Instituto de Salud Publica de Chile | Andrés E Castillo et al |
| EPI_ISL_593776; EPI_ISL_593772 | HOSPITAL REGIONAL LAMBAYEQUE | GENOMA MAYOR | Franklin R. Aguilar-Gamboa et al |
| EPI_ISL_682271; EPI_ISL_682266; EPI_ISL_682263; EPI_ISL_682270; EPI_ISL_682272; EPI_ISL_682267; EPI_ISL_682273; EPI_ISL_682262 | HOSPITAL SAN JUAN DE DIOS | Inciensa, Instituto Costarricense de Investigación y Enseñanza en Nutrición y Salud | Francisco Duarte et al |

| | | | |
|---|---|---|---|
| EPI_ISL_445351; EPI_ISL_445350; EPI_ISL_445375; EPI_ISL_445374 | HOSPITAL SAN JUAN DE DIOS | Instituto de Salud Publica de Chile | Andrés E Castillo et al |
| EPI_ISL_418251 | HOSPITAL UNIVERSITARIO LA PAZ | Instituto de Salud Carlos III | Iglesias-Caballero et al B (https://dx.doi.org/10.1101/2020.04.20.050039) |
| EPI_ISL_418243 | HOSPITAL UNIVERSITARIO VIRGEN DE LAS NIEVES | Instituto de Salud Carlos III | Iglesias-Caballero et al B (https://dx.doi.org/10.1101/2020.04.20.050039) |
| EPI_ISL_636978; EPI_ISL_636979; EPI_ISL_636976; EPI_ISL_636977; EPI_ISL_636975 | HP Pemba | KRISP, KZN Research Innovation and Sequencing Platform | Ismael N et al |
| EPI_ISL_430847 | HS mikrobiologi virus | The Public Health Agency of Sweden | Zhibing Yun et al |
| EPI_ISL_421463 | HSE Ilha Terceira - Angra do Heroismo | Instituto Nacional de Saude (INSA) | Guiomar et al et al |
| EPI_ISL_412971 | HUS Diagnostiikkakeskus, Hallinto | Department of Virology Faculty of Medicine, Medicum University of Helsinki | Teemu Smura et al B (https://dx.doi.org/10.1002/jmv.25773) |
| EPI_ISL_615105; EPI_ISL_582805; EPI_ISL_454436 | Halmstad klinisk mikrobiologi | The Public Health Agency of Sweden | Anna-Malin Linde et al |
| EPI_ISL_475548; EPI_ISL_475557; EPI_ISL_469074; EPI_ISL_475158; EPI_ISL_475156 | Halmstad klinisk mikrobiologi | The Public Health Agency of Sweden | Oskar Karlsson Lindsjo et al |
| EPI_ISL_710607; EPI_ISL_710606 | Halsomedicinskt Center | The Public Health Agency of Sweden | Department of Microbiology et al |
| EPI_ISL_482579; EPI_ISL_482581 | Hangzhou Center for Diseases Control and Prevention | Hangzhou Center for Diseases Control and Prevention | Jun Li et al |
| EPI_ISL_700516; EPI_ISL_700480; EPI_ISL_700566 | Hanover Park CHC wc HPH | NHLS/UCT | Arash Iranzadeh et al |
| EPI_ISL_700508 | Heideveld CDC wc HVP | NHLS/UCT | Arash Iranzadeh et al |
| EPI_ISL_700584 | Heideveld Emergency Centre | NHLS/UCT | Arash Iranzadeh et al |
| EPI_ISL_755592; EPI_ISL_755590 | Helix/Illumina | Genomics and Discovery, Respiratory Viruses Branch, Division of Viral Diseases, Centers for Disease Control and Prevention | Peter W. Cook et al |
| EPI_ISL_501231; EPI_ISL_501248; EPI_ISL_501250; EPI_ISL_487377; EPI_ISL_501233; EPI_ISL_487379; EPI_ISL_501235; EPI_ISL_501236; EPI_ISL_487381; EPI_ISL_501251; EPI_ISL_487370; EPI_ISL_501249 | Hellenic Pasteur Institute, National Influenza Reference laboratory of Southern Greece & Unit of Bioinformatics and Applied Genomics | Hellenic Pasteur Institute, National Influenza Reference laboratory of Southern Greece & Unit of Bioinformatics and Applied Genomics | Vasiliki Pogka et al |
| EPI_ISL_501230 | Hellenic Pasteur Institute, Public Health Laboratories | Hellenic Pasteur Institute, National Influenza Reference laboratory of Southern Greece & Unit of Bioinformatics and Applied Genomics | Vasiliki Pogka et al |
| EPI_ISL_430469 | Hellenic Pasteur Institute, Public Health Laboratories | Hellenic Pasteur Institute, Public Health Laboratories, Unit of Bioinformatics and Applied Genomics | Vasiliki Pogka et al |
| EPI_ISL_450529; EPI_ISL_450526; EPI_ISL_512297; EPI_ISL_512296; EPI_ISL_512299; EPI_ISL_512300; EPI_ISL_512302; EPI_ISL_450525 | Hematology Laboratory, Section of Molecular Diagnostics, University Clinical Centre, Medical University of Gdansk | Department of Virology, Faculty of Medicine, University of Helsinki, Helsinki, Finland | Maciej Grzybek et al |
| EPI_ISL_428236; EPI_ISL_428232; EPI_ISL_428233 | Hematology Laboratory, Section of Molecular Diagnostics, University Clinical Centre, Medical University of Gdansk | Department of Virology, Faculty of Medicine, University of Helsinki, Helsinki, Finland | Marlena Robakowska et al |
| EPI_ISL_451654; EPI_ISL_451649; EPI_ISL_451650; EPI_ISL_451648 | Hematology Laboratory, Section of Molecular Diagnostics, University Clinical Centre, Medical University of Gdansk | Laboratory of Recombinant Vaccines | Lukasz Rabalski et al |
| EPI_ISL_700274; EPI_ISL_700289; EPI_ISL_700306; EPI_ISL_699969; EPI_ISL_723067; EPI_ISL_699899; EPI_ISL_700065 | Hematopathology Laboratory, ACTREC, TMC | Hematopathology Laboratory, ACTREC, TMC | Hematopathology Laboratory et al |
| EPI_ISL_470575 | Hermes Pardini | Bioinformatics Laboratory / LNCC | Alexandra Gerber et al (https://dx.doi.org/10.1126/science.abd2161) |
| EPI_ISL_479815 | Hokkaido Institute of Public Health | Pathogen Genomics Center, National Institute of Infectious Diseases | Tsuyoshi Sekizuka et al C (https://www.medrxiv.org/content/10.1101/2020.07.01.20143958v1) |
| EPI_ISL_412030 | Hong Kong Department of Health | School of Public Health, The University of Hon g Kong | Dominic N.C. Tsang et al |
| EPI_ISL_760034; EPI_ISL_760033; EPI_ISL_760036; EPI_ISL_760035; EPI_ISL_760055; EPI_ISL_760039; EPI_ISL_760037; EPI_ISL_760038; EPI_ISL_760041 | Hong Kong Department of Health | School of Public Health, The University of Hong Kong | Daniel Chu et al |
| EPI_ISL_718246; EPI_ISL_718234; EPI_ISL_718249; EPI_ISL_718240 | Hospital | National Reference Center for Viruses of Respiratory Infections, Institut Pasteur, Paris | Marion Barbet et al B |
| EPI_ISL_560605; EPI_ISL_560601; EPI_ISL_560604; EPI_ISL_560599; EPI_ISL_560600; EPI_ISL_560598 | Hospital | National Reference Center for Viruses of Respiratory Infections, Institut Pasteur, Paris | Sylvie Behillil et al |
| EPI_ISL_434380 | Hospital AZ Rivierenland | Institute of Tropical Medicine | Philippe Selhorst et al |
| EPI_ISL_491455 | Hospital Clinica Biblica | Inciensa, Instituto Costarricense de Investigación y Enseñanza en Nutrición y Salud | Francisco Duarte et al |
| EPI_ISL_480318; EPI_ISL_480320; EPI_ISL_480316; EPI_ISL_480319 | Hospital Clínica Bíblica | Charité Virology-University of Costa Rica | Andres Moreira-Soto et al |
| EPI_ISL_691718; EPI_ISL_691702; EPI_ISL_691715 | Hospital Clínico San Carlos | Instituto de Salud Carlos III | Iglesias-Caballero et al A |
| EPI_ISL_683595; EPI_ISL_671800 | Hospital Clínico Universitario Lozano Blesa de Zaragoza (España) | SeqCOVID-SPAIN consortium/IBV(CSIC) | Rafael Benito et al |
| EPI_ISL_510284; EPI_ISL_510283 | Hospital Clínico Universitario de Santiago de Compostela | SeqCOVID-SPAIN consortium/IBV(CSIC) | José Javier Costa Alcalde et al |
| EPI_ISL_541046 | Hospital Clínico Universitario de Santiago de Compostela | SeqCOVID-SPAIN consortium/Institute of Biomedicine of Valencia, IBV-CSIC | José Javier Costa Alcalde et al |
| EPI_ISL_512674; EPI_ISL_512672 | Hospital De Niños Dr. Carlos Saenz Herrera [San Jose/San Jose] | Inciensa, Instituto Costarricense de Investigación y Enseñanza en Nutrición y Salud | Francisco Duarte et al |
| EPI_ISL_476561 | Hospital Garrahan | Héritas | Roberta Crespo et al |
| EPI_ISL_578186 | Hospital General Juan Ramón Jiménez | Instituto de Salud Carlos III | Iglesias-Caballero et al A |
| EPI_ISL_756271; EPI_ISL_756273 | Hospital General Universitario Gregorio Marañón | Hospital General Universitario Gregorio Marañón | Sergio Buenestado Serrano et al |
| EPI_ISL_654190; EPI_ISL_654032; EPI_ISL_654280; EPI_ISL_654223; EPI_ISL_654147; EPI_ISL_654088; EPI_ISL_654118; EPI_ISL_654121; EPI_ISL_654199 | Hospital General Universitario Gregorio Marañón | SeqCOVID-SPAIN consortium/IBV(CSIC) | Darío García de Viedma et al |
| EPI_ISL_467246; EPI_ISL_510115; EPI_ISL_481045; EPI_ISL_481041; EPI_ISL_481068; EPI_ISL_510104; EPI_ISL_481107; EPI_ISL_541897; EPI_ISL_541914; EPI_ISL_541895; EPI_ISL_541919; EPI_ISL_481072 | Hospital General Universitario Gregorio Marañón | SeqCOVID-SPAIN consortium/IBV(CSIC) | Laura Pérez-Lago et al |
| EPI_ISL_756272 | Hospital General Universitario La Princesa | Hospital General Universitario Gregorio Marañón | Sergio Buenestado Serrano et al |
| EPI_ISL_574431; EPI_ISL_481247; EPI_ISL_481244; EPI_ISL_481245 | Hospital IESS Babahoyo | Institute of Microbiology, Universidad San Francisco de Quito | Belén Prado-Vivar et al |
| EPI_ISL_471267 | Hospital IESS Babahoyo | Institute of Microbiology, Universidad San Francisco de Quito | Sully Márquez et al |
| EPI_ISL_412964 | Hospital Israelita Albert Einstein | Instituto Adolfo Lutz Interdisciplinary Procedures Center Strategic Laboratory | Jaqueline Goes de Jesus et al (https://virological.org/t/first-cases-of-coronavirus-disease-covid-19-in-brazil-south-america-2-genomes-3rd-march-2020/409) |
| EPI_ISL_416033 | Hospital Israelita Albert Einstein | Instituto Adolfo Lutz, Interdiciplinary Procedures Center, Strategic Laboratory | Claudio Tavares Sacchi et al A (https://dx.doi.org/10.1101/2020.05.23.20111443) |
| EPI_ISL_414014 | Hospital Israelita Albert Einstein | Instituto Adolfo Lutz, Interdiciplinary Procedures Center, Strategic Laboratory | Claudio Tavares Sacchi et al C (https://dx.doi.org/10.1590/s1678-9946202062030) |
| EPI_ISL_413016 | Hospital Israelita Albert Einstein | Instituto Adolfo Lutz, Interdisciplinary Procedures Center, Strategic Laboratory | Jaqueline Goes de Jesus et al (https://virological.org/t/first-cases-of-coronavirus-disease-covid-19-in-brazil-south-america-2-genomes-3rd-march-2020/409) |
| EPI_ISL_480313; EPI_ISL_480314; EPI_ISL_480312 | Hospital Mexico | Charité Virology-University of Costa Rica | Andres Moreira-Soto et al |
| EPI_ISL_480326; EPI_ISL_480325; EPI_ISL_480327 | Hospital Nacional de Niños | Charité Virology-University of Costa Rica | Andres Moreira-Soto et al |
| EPI_ISL_539496 | Hospital Nostra Senyora de Meritxell | Instituto de Salud Carlos III | Iglesias-Caballero et al A |
| EPI_ISL_471271; EPI_ISL_471269; EPI_ISL_471270 | Hospital Oncológico Solca Núcleo de Quito | Institute of Microbiology, Universidad San Francisco de Quito | Sully Márquez et al |
| EPI_ISL_419386 | Hospital Prof. Doutor Fernando Fonseca, EPE | Instituto Gulbenkian de Ciência | João Costa et al |
| EPI_ISL_455336 | Hospital San Pedro | Instituto de Salud Carlos III | Iglesias-Caballero et al A |
| EPI_ISL_539558 | Hospital San Pedro de Alcántara | Instituto de Salud Carlos III | Iglesias-Caballero et al A |
| EPI_ISL_510308; EPI_ISL_510306 | Hospital San Pedro de Alcántara (Cáceres) | SeqCOVID-SPAIN consortium/IBV(CSIC) | Cristina Muñoz Cuevas et al |
| EPI_ISL_527753 | Hospital San Vicente De Paul | Inciensa, Instituto Costarricense de Investigación y Enseñanza en Nutrición y Salud | Francisco Duarte et al |
| EPI_ISL_414017; EPI_ISL_414016; EPI_ISL_414015 | Hospital São Joaquim Beneficencia Portuguesa | Instituto Adolfo Lutz, Interdiciplinary Procedures Center, Strategic Laboratory | Claudio Tavares Sacchi et al C (https://dx.doi.org/10.1590/s1678-9946202062030) |

| | | | |
|---|---|---|---|
| EPI_ISL_455351 | Hospital Txagorritxu | Instituto de Salud Carlos III | Iglesias-Caballero et al A |
| EPI_ISL_737930 | Hospital Universitario 12 de Octubre | Hospital Universitario 12 de Octubre | Raul Recio et al |
| EPI_ISL_467134; EPI_ISL_467130; EPI_ISL_467159; EPI_ISL_467175; EPI_ISL_467142; EPI_ISL_467124 | Hospital Universitario Araba. Vitoria-Gasteiz | SeqCOVID-SPAIN consortium/IBV(CSIC) | Silvia Hernáez Crespo et al |
| EPI_ISL_452772; EPI_ISL_452692; EPI_ISL_452722 | Hospital Universitario Araba. Vitoria-Gasteiz, | SeqCOVID-SPAIN consortium/IBV(CSIC) | Silvia Hernáez Crespo et al |
| EPI_ISL_417975 | Hospital Universitario La Paz | Hospital Universitario La Paz | Elias Dahdouh et al (https://dx.doi.org/10.1101/2020.04.20.050039) |
| EPI_ISL_530072; EPI_ISL_530028; EPI_ISL_530042 | Hospital Universitario La Paz | Hospital Universitario La Paz | María Rodríguez et al |
| EPI_ISL_582101 | Hospital Universitario Marqués de Valdecilla - IDIVAL (Santander, Cantabria) | SeqCOVID-SPAIN consortium/IBV(CSIC) | María Eliecer Cano García et al |
| EPI_ISL_452467 | Hospital Universitario Puerta del Mar de Cádiz - INIBICA | SeqCOVID-SPAIN consortium/IBV(CSIC) | Salud Rodríguez-Pallares et al |
| EPI_ISL_530117 | Hospital Universitario Ramón y Cajal | Hospital Universitario La Paz | María Rodríguez et al |
| EPI_ISL_691730 | Hospital Universitario Severo Ochoa | Instituto de Salud Carlos III | Iglesias-Caballero et al A |
| EPI_ISL_534314 | Hospital Universitario da USP de SP | Instituto Adolfo Lutz, Interdiciplinary Procedures Center, Strategic Laboratory | Claudio Tavares Sacchi et al B |
| EPI_ISL_691697; EPI_ISL_691686 | Hospital Universitario de Ceuta | Instituto de Salud Carlos III | Iglesias-Caballero et al A |
| EPI_ISL_693657; EPI_ISL_693639; EPI_ISL_693642; EPI_ISL_693640; EPI_ISL_693638; EPI_ISL_693641; EPI_ISL_693535; EPI_ISL_693652; EPI_ISL_693653; EPI_ISL_693651; EPI_ISL_693654; EPI_ISL_693636; EPI_ISL_693520; EPI_ISL_693631; EPI_ISL_693634; EPI_ISL_693630; EPI_ISL_693632; EPI_ISL_693633; EPI_ISL_693523; EPI_ISL_693635; EPI_ISL_693637 | Hospital Vila Franca de Xira | Instituto Nacional de Saude (INSA) | Borges et al et al |
| EPI_ISL_455316; EPI_ISL_578188; EPI_ISL_578187; EPI_ISL_539497 | Hospital Virgen de las Nieves | Instituto de Salud Carlos III | Iglesias-Caballero et al A |
| EPI_ISL_455314 | Hospital Virgen del Rocio | Instituto de Salud Carlos III | Iglesias-Caballero et al A |
| EPI_ISL_721989; EPI_ISL_722003 | Hospital das Clínicas Universidade de São Paulo Medical School | Laboratório de Parasitologia Médica - Instituto de Medicina Tropical - Universidade de São Paulo | Brazil-UK Centre for Arbovirus Discovery Diagnosis Genomics et al |
| EPI_ISL_476565 | Hospital de Pediatría "Prof. Dr. Juan P Garrahan" | Héritas | Andrea Mangano et al |
| EPI_ISL_615121; EPI_ISL_476568 | Hospital de Pediatría "Prof. Dr. Juan P Garrahan" | Héritas | Cristian Rohr et al |
| EPI_ISL_476573; EPI_ISL_476567; EPI_ISL_476563; EPI_ISL_476571 | Hospital de Pediatría "Prof. Dr. Juan P Garrahan" | Héritas | Dalmacio Pereyra et al |
| EPI_ISL_414578 | Hospital de Talca, Chile | Instituto de Salud Publica de Chile | Andrés E. Castillo et al (https://dx.doi.org/10.1002/jmv.25797) |
| EPI_ISL_671817; EPI_ISL_671808; EPI_ISL_671833; EPI_ISL_671825; EPI_ISL_671809; EPI_ISL_671819; EPI_ISL_671823; EPI_ISL_671816 | Hospital de la Santa Creu i Sant Pau. Servicio de Microbiología | SeqCOVID-SPAIN consortium/IBV(CSIC) | Ferran Navarro et al |
| EPI_ISL_541078; EPI_ISL_541081 | Hospital de la Santa Creu i Sant Pau. Servicio de Microbiología | SeqCOVID-SPAIN consortium/Institute of Biomedicine of Valencia, IBV-CSIC | Ferran Navarro et al |
| EPI_ISL_707710; EPI_ISL_707709; EPI_ISL_458026; EPI_ISL_458027; EPI_ISL_498259; EPI_ISL_498257 | Hospital for Tropical Diseases | COVID-19 Network Investigations (CONI) Alliance | Elizabeth Batty et al |
| EPI_ISL_590915; EPI_ISL_500797; EPI_ISL_590936; EPI_ISL_549036; EPI_ISL_471173; EPI_ISL_471176; EPI_ISL_420147; EPI_ISL_500799; EPI_ISL_668428 | Hospital of Southern Norway - Kristiansand, Department of Medical Microbiology | Norwegian Institute of Public Health, Department of Virology | Kathrine Stene-Johansen et al |
| EPI_ISL_593916 | Hospital, Sarreguemines | National Reference Center for Viruses of Respiratory Infections, Institut Pasteur, Paris | Sylvie Behillil et al |
| EPI_ISL_543834; EPI_ISL_545523; EPI_ISL_544136; EPI_ISL_546491; EPI_ISL_543262; EPI_ISL_543560 | Houston Methodist Hospital | Houston Methodist Hospital | S. Wesley Long et al |
| EPI_ISL_452237 | Huddinge VC | The Public Health Agency of Sweden | Anders Johansson et al |
| EPI_ISL_445088 | Human Genetic Research Center, Kawsar Biotech Company | Human Genetic Research Center, Kawsar Biotech Company | Abbasalipour Bashash et al |
| EPI_ISL_645011; EPI_ISL_645012; EPI_ISL_645042; EPI_ISL_645006; EPI_ISL_645005; EPI_ISL_645030; EPI_ISL_645004; EPI_ISL_645036; EPI_ISL_645037; EPI_ISL_645077; EPI_ISL_645007; EPI_ISL_645009; EPI_ISL_645008; EPI_ISL_645065; EPI_ISL_645063; EPI_ISL_645055; EPI_ISL_645109; EPI_ISL_645059; EPI_ISL_645080; EPI_ISL_645078; EPI_ISL_645082; EPI_ISL_645081; EPI_ISL_645079; EPI_ISL_645021; EPI_ISL_645026; EPI_ISL_645025; EPI_ISL_645043; EPI_ISL_645089; EPI_ISL_645097; EPI_ISL_645061; EPI_ISL_645088; EPI_ISL_645069; EPI_ISL_645087; EPI_ISL_645064; EPI_ISL_645060; EPI_ISL_645062; EPI_ISL_645056; EPI_ISL_645057 | Human Genome Variation Research Group, Malopolska Centre of Biotechnology | Human Genome Variation Research Group, Malopolska Centre of Biotechnology | Kowalski et al |
| EPI_ISL_526217; EPI_ISL_526222; EPI_ISL_526230; EPI_ISL_526232; EPI_ISL_526219; EPI_ISL_526220; EPI_ISL_526221; EPI_ISL_526236; EPI_ISL_526223; EPI_ISL_526224; EPI_ISL_526225 | Hungarian Defence Forces Military Medical Centre | National Laboratory of Virology, Szentágothai Research Centre | Endre Gábor Tóth et al |
| EPI_ISL_759956 | Husada Utama Hospital | Institute of Tropical Disease, Universitas Airlangga | Jezzy R Dewantari et al |
| EPI_ISL_529961 | Husada Utama Hospital | Institute of Tropical Disease, Universitas Airlangga | Krisnoadi Rahardjo et al |
| EPI_ISL_536297; EPI_ISL_536094; EPI_ISL_536350 | Hôpital Charles-LeMoyne | Laboratoire de santé publique du Québec | Sandrine Moreira et al |
| EPI_ISL_710540; EPI_ISL_710534; EPI_ISL_712060; EPI_ISL_710532; EPI_ISL_710541; EPI_ISL_710537; EPI_ISL_710575; EPI_ISL_711057 | Hôpital Fattouma-Bourguiba de Monastir | Laboratoire des Procédés de Criblage Moléculaire et Cellulaire-Centre de Biotechnologie de Sfax | Souissi et al |
| EPI_ISL_536168 | Hôpital de Hull | Laboratoire de santé publique du Québec | Sandrine Moreira et al |
| EPI_ISL_536227 | Hôpital du Suroît | Laboratoire de santé publique du Québec | Sandrine Moreira et al |
| EPI_ISL_535875 | Hôpital général du Lakeshore | Laboratoire de santé publique du Québec | Sandrine Moreira et al |
| EPI_ISL_535821 | Hôtel-Dieu de Lévis | Laboratoire de santé publique du Québec | Sandrine Moreira et al |
| EPI_ISL_653778; EPI_ISL_653777; EPI_ISL_653770; EPI_ISL_653783; EPI_ISL_653814; EPI_ISL_722865; EPI_ISL_722868; EPI_ISL_722863; EPI_ISL_722864; EPI_ISL_722900; EPI_ISL_722851; EPI_ISL_722860; EPI_ISL_722859; EPI_ISL_722861; EPI_ISL_722867; EPI_ISL_722853; EPI_ISL_722895; EPI_ISL_653805; EPI_ISL_722852; EPI_ISL_722871; EPI_ISL_653780; EPI_ISL_653773; EPI_ISL_653774; EPI_ISL_653768; EPI_ISL_653806; EPI_ISL_653804 | I.R.C.C.S. "S. De Bellis" - Ente Ospedaliero | Istituto Zooprofilattico Sperimentale della Puglia e della Basilicata | Parisi A. et al |
| EPI_ISL_455645 | ICMR-National Institute of Cholera and Enteric Diseases | National Institute of Biomedical Genomics | Arindam Maitra et al |
| EPI_ISL_404253 | IL Department of Public Health Chicago Laboratory | Pathogen Discovery, Respiratory Viruses Branch, Division of Viral Diseases, Centers for Disease Control and Prevention | Ying Tao et al B (https://dx.doi.org/10.1101/2020.03.09.20032896) |
| EPI_ISL_445379 | IMALAB- HOSPITAL FACH | Instituto de Salud Publica de Chile | Andrés E Castillo et al |
| EPI_ISL_648209; EPI_ISL_648210; EPI_ISL_648214; EPI_ISL_648211; EPI_ISL_648215; EPI_ISL_648216; EPI_ISL_648212; EPI_ISL_648213; EPI_ISL_648677 | INBIRS-UBA | Laboratorio Mixto de Biotecnología Acuática (LMBA) | Joaquín Ezpeleta et al |
| EPI_ISL_603173; EPI_ISL_603158; EPI_ISL_590694; EPI_ISL_590698 | INMI Lazzaro Spallanzani IRCCS | INMI Lazzaro Spallanzani IRCCS | Barbara Bartolini et al |
| EPI_ISL_609989 | INMI Lazzaro Spallanzani IRCCS | INMI Lazzaro Spallanzani IRCCS | C.E.M Gruber et al |
| EPI_ISL_603161; EPI_ISL_603144; EPI_ISL_603146; EPI_ISL_603147; EPI_ISL_590695 | INMI Lazzaro Spallanzani IRCCS | INMI Lazzaro Spallanzani IRCCS | Cesare E.M. Gruber et al B |
| EPI_ISL_603157; EPI_ISL_603149 | INMI Lazzaro Spallanzani IRCCS | INMI Lazzaro Spallanzani IRCCS | Francesco Messina et al |
| EPI_ISL_590697; EPI_ISL_603154; EPI_ISL_603155; EPI_ISL_590693 | INMI Lazzaro Spallanzani IRCCS | INMI Lazzaro Spallanzani IRCCS | Martina Rueca et al |
| EPI_ISL_410545 | INMI Lazzaro Spallanzani IRCCS | Laboratory of Virology, INMI Lazzaro Spallanzani IRCCS | Maria R. Capobianchi et al A (https://dx.doi.org/10.2807/1560-7917.ES.2020.25.13.2000305) |
| EPI_ISL_410546 | INMI Lazzaro Spallanzani IRCCS | Laboratory of Virology, INMI Lazzaro Spallanzani IRCCS | Maria R. Capobianchi et al B (https://dx.doi.org/10.1002/jmv.25699) |
| EPI_ISL_751349; EPI_ISL_751323; EPI_ISL_751324; EPI_ISL_751388; EPI_ISL_751346; EPI_ISL_751432; EPI_ISL_751443; EPI_ISL_751444; EPI_ISL_492986; EPI_ISL_751343 | IRCCS Sacro Cuore Don Calabria Hospital, Department of Infectious, Tropical Diseases & Microbiology | University of Verona, Department of Biotechnology | Antonio Mori et al |
| EPI_ISL_584072; EPI_ISL_572322; EPI_ISL_572323; EPI_ISL_572320; EPI_ISL_572321 | IZSM | IZSM | Maurizio Viscardi et al |
| EPI_ISL_479938; EPI_ISL_480184; EPI_ISL_480029 | Ibaraki Prefectural Institute of Public Health | Pathogen Genomics Center, National Institute of Infectious Diseases | Tsuyoshi Sekizuka et al C (https://www.medrxiv.org/content/10.1101/2020.07.01.20143958v1) |
| EPI_ISL_481145; EPI_ISL_481159 | Immunogenomics lab, Institute of Life Sciences, Bhubaneswar | Immunogenomics lab, Institute of Life Sciences, Bhubaneswar | Sunil Raghav et al |
| EPI_ISL_476704; EPI_ISL_476702 | Incubadora Venezolana de Ciencia, Venezuela | Incubadora Venezolana de Ciencia, Venezuela / Instituto Nacional de Salud, Bogotá, Colombia / Grupo de Investigaciones Microbiológicas-UR (GIMUR), Departamento de Biología, Facultad de Ciencias Naturales, Universidad del Rosario, Bogotá, Colombia / Icahn School of Medicine at Mount Sinai, New York, USA | Alberto Paniz-Mondolfi et al |
| EPI_ISL_413522 | Indian Council of Medical Research - National Institute of Virology | National Influenza Center, Indian Council of Medical Research - National Institute of Virology | Potdar V et al A (https://dx.doi.org/10.4103/ijmr.IJMR_663_20) |
| EPI_ISL_413523 | Indian Council of Medical Research-National Institute of Virology | National Influenza Center, Indian Council of Medical Research-National Institute of Virology | Potdar V et al A (https://dx.doi.org/10.4103/ijmr.IJMR_663_20) |
| EPI_ISL_637099; EPI_ISL_637102 | Indian Council of Medical Research-National Institute of Virology, Microbial Containment Complex | Indian Council of Medical Research-National Institute of Virology, Microbial Containment Complex | Pragya D. Yadav et al A |

| | | | |
|---|---|---|---|
| EPI_ISL_596454 | Infectious Disease and Tropical Medicine Research Center, Resistant Tuberculosis Institute, Zahedan University of Medical Sciences, Zahedan, Iran. | Genetics Research Center, University of Social Welfare and Rehabilitation Sciences | Zohreh Fattahi et al |
| EPI_ISL_496371; EPI_ISL_496364; EPI_ISL_496366; EPI_ISL_496359; EPI_ISL_496373; EPI_ISL_496367; EPI_ISL_496365; EPI_ISL_496348; EPI_ISL_496341; EPI_ISL_496354; EPI_ISL_496340; EPI_ISL_496374 | Infectolab | Andersen lab at Scripps Research | SEARCH Alliance San Diego with Samuel Navarro Alvarez et al |
| EPI_ISL_418216; EPI_ISL_420078; EPI_ISL_480789; EPI_ISL_420077; EPI_ISL_420069; EPI_ISL_477142; EPI_ISL_477151; EPI_ISL_486859; EPI_ISL_486872; EPI_ISL_482878; EPI_ISL_485710; EPI_ISL_476493; EPI_ISL_486865; EPI_ISL_486868; EPI_ISL_481234; EPI_ISL_420076; EPI_ISL_418206; EPI_ISL_485716; EPI_ISL_420070; EPI_ISL_477152; EPI_ISL_477141; EPI_ISL_420074; EPI_ISL_485715; EPI_ISL_481235; EPI_ISL_486871; EPI_ISL_476574; EPI_ISL_480783; EPI_ISL_418210; EPI_ISL_418217; EPI_ISL_486863 | Institut Pasteur Dakar | Institut Pasteur de Dakar | Ndongo Dia et al |
| EPI_ISL_498238; EPI_ISL_498239; EPI_ISL_498252; EPI_ISL_498231 | Institut Pasteur de Dakar | Institut Pasteur de Dakar | Ndongo Dia et al |
| EPI_ISL_613433; EPI_ISL_613443; EPI_ISL_613432; EPI_ISL_613438; EPI_ISL_613446; EPI_ISL_613421; EPI_ISL_613422; EPI_ISL_613450; EPI_ISL_613424; EPI_ISL_613431; EPI_ISL_613452; EPI_ISL_613429; EPI_ISL_613423; EPI_ISL_613419; EPI_ISL_613447; EPI_ISL_613455; EPI_ISL_613454; EPI_ISL_613453; EPI_ISL_613451; EPI_ISL_613448; EPI_ISL_613449; EPI_ISL_613456 | Institut Pasteur de la Guadeloupe | Institut Pasteur de la Guadeloupe | Marion Barbet et al B |
| EPI_ISL_459969; EPI_ISL_459977; EPI_ISL_459965; EPI_ISL_459978; EPI_ISL_459983; EPI_ISL_459984; EPI_ISL_459976; EPI_ISL_459967; EPI_ISL_459974; EPI_ISL_459972; EPI_ISL_459968; EPI_ISL_459970 | Institut Pasteur du Maroc | Institut Pasteur du Maroc | Marion Barbet et al A (https://dx.doi.org/10.1101/2020.06.23.165902) |
| EPI_ISL_417336; EPI_ISL_508873; EPI_ISL_508896 | Institut des Agents Infectieux (IAI), Hospices Civils de Lyon | CNR Virus des Infections Respiratoires - France SUD | Antonin Bal et al |
| EPI_ISL_583610 | Institut für Virologie am Department für Hygiene, Mikrobiologie und Public Health | Bergthaler laboratory, CeMM Research Center for Molecular Medicine of the Austrian Academy of Sciences | Alexandra Popa et al A |
| EPI_ISL_475827 | Institut für Virologie am Department für Hygiene, Mikrobiologie und Public Health | Bergthaler laboratory, CeMM Research Center for Molecular Medicine of the Austrian Academy of Sciences | Alexandra Popa et al B (https://dx.doi.org/10.1101/2020.07.15.204339 v1) |
| EPI_ISL_437990; EPI_ISL_437932; EPI_ISL_437915; EPI_ISL_437991; EPI_ISL_437986 | Institut für Virologie am Department für Hygiene, Mikrobiologie und Public Health | Bergthaler laboratory, CeMM Research Center for Molecular Medicine of the Austrian Academy of Sciences | Alexandra Popa et al B (https://www.biorxiv.org/content/10.1101/2020. 07.15.204339v1) |
| EPI_ISL_508688; EPI_ISL_508687; EPI_ISL_508699; EPI_ISL_508704; EPI_ISL_508701; EPI_ISL_508700; EPI_ISL_508694; EPI_ISL_508703; EPI_ISL_508697; EPI_ISL_508693; EPI_ISL_508692; EPI_ISL_508702; EPI_ISL_508698; EPI_ISL_508695; EPI_ISL_508690; EPI_ISL_508689; EPI_ISL_508691; EPI_ISL_508696 | Institut für Virologie und Epidemiologie der Viruskrankheiten, Universitätsklinikum Tübingen | NGS Competence Center Tübingen, Institut für Medizinische Mikrobiologie und Hygiene, Universitätsklinikum Tübingen | Angel Angelov et al |
| EPI_ISL_485809 | Institut für Virologie und Epidemiologie der Viruskrankheiten, Universitätsklinikum Tübingen | NGS Competence Center Tübingen, Institut für Medizinische Mikrobiologie und Hygiene, Universitätsklinikum Tübingen | Angelov at al. et al |
| EPI_ISL_485811; EPI_ISL_485813; EPI_ISL_485812; EPI_ISL_485810 | Institut für Virologie und Epidemiologie der Viruskrankheiten, Universitätsklinikum Tübingen | NGS Competence Center Tübingen, Institut für Medizinische Mikrobiologie und Hygiene, Universitätsklinikum Tübingen | Angelov et al. et al |
| EPI_ISL_574790 | Institute for Infectious Diseases, University of Bern | Institute for Infectious Diseases, University of Bern | Michel C Koch et al |
| EPI_ISL_583865; EPI_ISL_583856 | Institute for Laboratory Diagnostics and Microbiology, Klinikum Klagenfurt am Worthersee | Bergthaler laboratory, CeMM Research Center for Molecular Medicine of the Austrian Academy of Sciences | Alexandra Popa et al A |
| EPI_ISL_677723; EPI_ISL_678253; EPI_ISL_678251 | Institute for Lung Diseases in Children - Skopje | Research Center for Genetic Engineering and Biotechnology "Georgi D. Efremov" , Macedonian Academy of Sciences and Arts | RCGEB - MASA et al |
| EPI_ISL_430439 | Institute for Medical Research, Infectious Disease Research Centre, National Institutes of Health, Ministry of Health Malaysia | Institute for Medical Research Infectious Disease Research Centre, National Institutes of Health, Ministry of Health Malaysia | Suppiah.J et al |
| EPI_ISL_490103; EPI_ISL_728165; EPI_ISL_718308; EPI_ISL_718307; EPI_ISL_718309; EPI_ISL_728170; EPI_ISL_728244; EPI_ISL_728167; EPI_ISL_718314; EPI_ISL_718283; EPI_ISL_718284; EPI_ISL_490089; EPI_ISL_490101; EPI_ISL_718278; EPI_ISL_718270; EPI_ISL_718268; EPI_ISL_718267; EPI_ISL_596449; EPI_ISL_718269; EPI_ISL_718279; EPI_ISL_718275; EPI_ISL_718274; EPI_ISL_718276; EPI_ISL_718271; EPI_ISL_718272; EPI_ISL_718277; EPI_ISL_718280; EPI_ISL_718281; EPI_ISL_718290; EPI_ISL_718299; EPI_ISL_718295; EPI_ISL_718292; EPI_ISL_718302; EPI_ISL_718303; EPI_ISL_718293; EPI_ISL_718298; EPI_ISL_718288; EPI_ISL_718294; EPI_ISL_728247; EPI_ISL_728209; EPI_ISL_728253; EPI_ISL_728158 | Institute for Medical Research, Infectious Disease Research Centre, National Institutes of Health, Ministry of Health Malaysia | Institute for Medical Research, Infectious Disease Research Centre, National Institutes of Health, Ministry of Health Malaysia | Suppiah J et al |
| EPI_ISL_430443; EPI_ISL_430444; EPI_ISL_430442; EPI_ISL_430441; EPI_ISL_430440 | Institute for Medical Research, Infectious Disease Research Centre, National Institutes of Health, Ministry of Health Malaysia | Institute for Medical Research, Infectious Disease Research Centre, National Institutes of Health, Ministry of Health Malaysia | Suppiah.J et al |
| EPI_ISL_455790; EPI_ISL_455792; EPI_ISL_455791; EPI_ISL_455793 | Institute for Medical Research, Infectious Disease Research Centre, National Institutes of Health, Ministry of Health Malaysia | Malaysia Genome Institute | Mohd Noor Mat Isa et al |
| EPI_ISL_455567; EPI_ISL_454605; EPI_ISL_454606; EPI_ISL_455566; EPI_ISL_468591; EPI_ISL_468656 | Institute for Public Health | Laboratory for advanced genomics | Filip Rokić et al |
| EPI_ISL_492076; EPI_ISL_492083 | Institute for Public Health of the Republic of North Macedonia | Charite Universitatsmedizin Berlin, Institute of Virology | Victor M Corman et al A |
| EPI_ISL_733542; EPI_ISL_733556; EPI_ISL_733548; EPI_ISL_733549; EPI_ISL_733535; EPI_ISL_733566; EPI_ISL_733533; EPI_ISL_733561; EPI_ISL_707707 | Institute for Urban Disease Control and Prevention | COVID-19 Network Investigations (CONI) Alliance | Kamolthip Atsawawaranunt et al |
| EPI_ISL_572330; EPI_ISL_572331; EPI_ISL_572333; EPI_ISL_572397; EPI_ISL_574259; EPI_ISL_573152 | Institute for Virology, University Hospital Duesseldorf, Medical Faculty, Heinrich-Heine-University Duesseldorf | Institute for Virology, University Hospital Duesseldorf, Medical Faculty, Heinrich-Heine-University Duesseldorf | Maximilian Damagnez et al |
| EPI_ISL_626228; EPI_ISL_602519; EPI_ISL_602528; EPI_ISL_602520; EPI_ISL_602527; EPI_ISL_626224; EPI_ISL_602522; EPI_ISL_602523; EPI_ISL_602521; EPI_ISL_602525; EPI_ISL_602526; EPI_ISL_602481; EPI_ISL_602505; EPI_ISL_602492; EPI_ISL_602468; EPI_ISL_602518; EPI_ISL_602501; EPI_ISL_602503; EPI_ISL_602508; EPI_ISL_602477; EPI_ISL_626223; EPI_ISL_602484; EPI_ISL_602479; EPI_ISL_602482; EPI_ISL_602506; EPI_ISL_602509; EPI_ISL_626216; EPI_ISL_602485; EPI_ISL_602466; EPI_ISL_602490; EPI_ISL_602524; EPI_ISL_626217; EPI_ISL_602500; EPI_ISL_602465; EPI_ISL_602464; EPI_ISL_602488; EPI_ISL_602489; EPI_ISL_602476; EPI_ISL_602474; EPI_ISL_602498; EPI_ISL_602499; EPI_ISL_602475; EPI_ISL_626226; EPI_ISL_602529; EPI_ISL_602478; EPI_ISL_602502; EPI_ISL_626214; EPI_ISL_626220; EPI_ISL_626211; EPI_ISL_626212; EPI_ISL_602496; EPI_ISL_602494; EPI_ISL_602495; EPI_ISL_602504; EPI_ISL_602470; EPI_ISL_602471; EPI_ISL_602472; EPI_ISL_626215; EPI_ISL_602480; EPI_ISL_626225; EPI_ISL_626219; EPI_ISL_602530; EPI_ISL_602531; EPI_ISL_602493; EPI_ISL_602469; EPI_ISL_602532; EPI_ISL_626218; EPI_ISL_626227; EPI_ISL_602463; EPI_ISL_602487; EPI_ISL_602467; EPI_ISL_602491; EPI_ISL_626221; EPI_ISL_626222; EPI_ISL_626229; EPI_ISL_626230; EPI_ISL_602483; EPI_ISL_602497; EPI_ISL_602507; EPI_ISL_602473; EPI_ISL_626213 | Institute for Virology, University Hospital Essen | Center of Medical Microbiology, Virology, and Hospital Hygiene, University of Duesseldorf | Olympia E. Anastasiou et al |
| EPI_ISL_728346 | Institute for medical Microbiology and clinical Hygene | Institute for medical Microbiology and clinical Hygene | Aljoscha Tersteegen et al |
| EPI_ISL_728357 | Institute for medical Microbiology and hospital Hygene | Insitute for medical Microbiology and hospital Hygene | Prof. Dr. Achim Kaasch et al |
| EPI_ISL_728348 | Institute for medical Microbiology and hospital Hygene | Institute for medical Microbiology and hospital Hygene | Aljsocha Tersteegen et al |
| EPI_ISL_728356; EPI_ISL_728351; EPI_ISL_728354 | Institute for medical Microbiology and hospital Hygene | Institute for medical Microbiology and hospital Hygene | Prof. Dr. Achim Kaasch et al |
| EPI_ISL_525472; EPI_ISL_525473 | Institute of Clinical Microbiology and Hygiene, University Hospital Regensburg | Institute of Clinical Microbiology and Hygiene, University Hospital Regensburg | Hiergeist et al |
| EPI_ISL_600524; EPI_ISL_600562 | Institute of Epidemiology Disease Control And Research | Institute for Developing Science and Health Initiatives | Lauren Cowley et al |
| EPI_ISL_582032; EPI_ISL_582031 | Institute of Human Genetics, Polish Academy of Sciences | Institute of Human Genetics, Polish Academy of Sciences | Szymon Hryhorowicz et al |
| EPI_ISL_485400 | Institute of Human Genetics, Polish Academy of Sciences | Institute of Human Genetics, Polish Academy of Sciences, | Szymon Hryhorowicz et al |
| EPI_ISL_463029; EPI_ISL_463058 | Institute of Life Sciences, Bhubaneswar | Immunogenomics lab, Institute of Life Sciences, Bhubaneswar | Sunil Raghav et al |
| EPI_ISL_723090; EPI_ISL_723084; EPI_ISL_723087; EPI_ISL_723099; EPI_ISL_723089; EPI_ISL_723101; EPI_ISL_723095; EPI_ISL_723096; EPI_ISL_723088; EPI_ISL_723116; EPI_ISL_723085; EPI_ISL_723114; EPI_ISL_723097; EPI_ISL_723103; EPI_ISL_723104; EPI_ISL_723079; EPI_ISL_723107; EPI_ISL_723111; EPI_ISL_723112; EPI_ISL_723083; EPI_ISL_723093; EPI_ISL_723115; EPI_ISL_723113; EPI_ISL_723098; EPI_ISL_723109; EPI_ISL_723094; EPI_ISL_723108; EPI_ISL_723117; EPI_ISL_723110; EPI_ISL_723081; EPI_ISL_723105; EPI_ISL_723106; EPI_ISL_723102; EPI_ISL_723091; EPI_ISL_723092; EPI_ISL_723100; EPI_ISL_723086; EPI_ISL_723077; EPI_ISL_723082 | Institute of Medical Genetics and Applied Genomics | Institute of Medical Genetics and Applied Genomics | Caspar Gross et al |
| EPI_ISL_590825; EPI_ISL_590831; EPI_ISL_590833; EPI_ISL_590836; EPI_ISL_590835 | Institute of Medical Virology, University of Zurich | Institute of Medical Virology, University of Zurich | Marie O. Pohl et al |
| EPI_ISL_422563 | Institute of Microbiology Universidad San Francisco de Quito | Institute of Microbiology Universidad San Francisco de Quito | Belen Prado-Vivar et al A (https://dx.doi.org/10.1101/2020. 06.11.20128330) |
| EPI_ISL_635200; EPI_ISL_635206; EPI_ISL_635203; EPI_ISL_635248; EPI_ISL_635225; EPI_ISL_635221; EPI_ISL_635230; EPI_ISL_635233; EPI_ISL_635237; EPI_ISL_635239; EPI_ISL_635209; EPI_ISL_635211; EPI_ISL_635212; EPI_ISL_635213; EPI_ISL_420294 | Institute of Microbiology and Immunology, Faculty of Medicine, University of Ljubljana | Institute of Microbiology and Immunology, Faculty of Medicine, University of Ljubljana | Samo Zakotnik et al |
| EPI_ISL_635293; EPI_ISL_635283; EPI_ISL_635272; EPI_ISL_635297; EPI_ISL_635284; EPI_ISL_635286; EPI_ISL_635275; EPI_ISL_635256; EPI_ISL_635289; EPI_ISL_635285; EPI_ISL_635281; EPI_ISL_635296; EPI_ISL_635279; EPI_ISL_635250; EPI_ISL_635267; EPI_ISL_635295; EPI_ISL_635299; EPI_ISL_635282; EPI_ISL_635277; EPI_ISL_636461; EPI_ISL_635271; EPI_ISL_635298; EPI_ISL_635292; EPI_ISL_635273 | Institute of Microbiology and Immunology, Faculty of Medicine, University of Ljubljana | Institute of Microbiology and Immunology, Faculty of Medicine, University of Ljubljana | Tomaž Mark Zorec et al |
| EPI_ISL_635204 | Institute of Microbiology and Immunology, Faculty of Medicine, University of Ljubljana | The National Laboratory of Health, Environment and Food - Centre for Medical Microbiology Maribor | Samo Zakotnik et al |
| EPI_ISL_697794; EPI_ISL_539785; EPI_ISL_697787; EPI_ISL_697795 | Institute of Microbiology, Universidad San Francisco de Quito | Institute of Microbiology, Universidad San Francisco de Quito | Andrea Macias et al |
| EPI_ISL_477014 | Institute of Microbiology, Universidad San Francisco de Quito | Institute of Microbiology, Universidad San Francisco de Quito | Belen Prado-Vivar et al B |
| EPI_ISL_527810; EPI_ISL_527811; EPI_ISL_539789; EPI_ISL_697790; EPI_ISL_697792; EPI_ISL_728205; EPI_ISL_697791; EPI_ISL_486843; EPI_ISL_486842; EPI_ISL_486844; EPI_ISL_697793; EPI_ISL_491937; EPI_ISL_491938; EPI_ISL_491936; EPI_ISL_491939; EPI_ISL_491940; EPI_ISL_486848; EPI_ISL_527814; EPI_ISL_527817; EPI_ISL_539792; EPI_ISL_486847; EPI_ISL_486845; EPI_ISL_697789 | Institute of Microbiology, Universidad San Francisco de Quito | Institute of Microbiology, Universidad San Francisco de Quito | Belén Prado-Vivar et al |
| EPI_ISL_525430; EPI_ISL_516649; EPI_ISL_477016; EPI_ISL_525435; EPI_ISL_525431; EPI_ISL_525437 | Institute of Microbiology, Universidad San Francisco de Quito | Institute of Microbiology, Universidad San Francisco de Quito | Juan José Guadalupe et al |
| EPI_ISL_516650; EPI_ISL_516651; EPI_ISL_516652 | Institute of Microbiology, Universidad San Francisco de Quito | Institute of Microbiology, Universidad San Francisco de Quito | Prado-Vivar et al |
| EPI_ISL_728204; EPI_ISL_417482; EPI_ISL_660530; EPI_ISL_660537; EPI_ISL_728202; EPI_ISL_728203; EPI_ISL_660529; EPI_ISL_477015; EPI_ISL_660534; EPI_ISL_660538; EPI_ISL_660536; EPI_ISL_660535; EPI_ISL_660532; EPI_ISL_660531; EPI_ISL_660533 | Institute of Microbiology, Universidad San Francisco de Quito | Institute of Microbiology, Universidad San Francisco de Quito | Sully Márquez et al |
| EPI_ISL_548945; EPI_ISL_548944; EPI_ISL_548942; EPI_ISL_548943; EPI_ISL_548946 | Institute of Microbiology, University of Veterinary and Animal sciences | Institute of Microbiology, University of Veterinary and Animal sciences | Yaqub et al |
| EPI_ISL_463008 | Institute of Molecular Virology, University Münster | Institute of Molecular Virology, University Münster | Angeles Mecate Zambrano et al |
| EPI_ISL_403929 | Institute of Pathogen Biology, Chinese Academy of Medical Sciences & Peking Union Medical College | Institute of Pathogen Biology, Chinese Academy of Medical Sciences & Peking Union Medical College | Lili Ren et al (https://dx.doi.org/10.1093/cid/ciaa203/5780800) |
| EPI_ISL_508385; EPI_ISL_508356; EPI_ISL_508368 | Institute of Post Graduate Medical Education & Research | National Institute of Biomedical Genomics | Arindam Maitra et al |
| EPI_ISL_511901 | Institute of Post Graduate Medical Education & Research | National Institute of Biomedical Genomics - DBT's PAN-INDIA 1000 SARS--CoV-2 RNA Genome Sequencing Consortium | Arindam Maitra et al |
| EPI_ISL_718256; EPI_ISL_718255; EPI_ISL_577740; EPI_ISL_583487; EPI_ISL_717975; EPI_ISL_718259; EPI_ISL_718257; EPI_ISL_577742; EPI_ISL_583488; EPI_ISL_577741 | Institute of Virology, Biomedical Research Center of the Slovak Academy of Sciences, Bratislava | Faculty of Natural Sciences, Comenius University, Bratislava | Broňa Brejová et al |
| EPI_ISL_718250; EPI_ISL_583484; EPI_ISL_583482; EPI_ISL_718254; EPI_ISL_577735; EPI_ISL_577737; EPI_ISL_577739; EPI_ISL_583486; EPI_ISL_718252 | Institute of Virology, Biomedical Research Center of the Slovak Academy of Sciences, Bratislava | Faculty of Natural Sciences, Comenius University, Bratislava | Kristína Boršová et al |
| EPI_ISL_583483; EPI_ISL_583485; EPI_ISL_577738; EPI_ISL_577736; EPI_ISL_577734; EPI_ISL_583489; EPI_ISL_583481; EPI_ISL_718260; EPI_ISL_718258; EPI_ISL_718251; EPI_ISL_718253 | Institute of Virology, Biomedical Research Center of the Slovak Academy of Sciences, Bratislava | Faculty of Natural Sciences, Comenius University, Bratislava | Viktória Hodorová et al |

| | | | |
|---|---|---|---|
| EPI_ISL_417880; EPI_ISL_417878; EPI_ISL_417879; EPI_ISL_417877 | Institute of Virology, Biomedical Research Center of the Slovak Academy of Sciences, Bratislava; Public Health Authority of the Slovak Republic, Bratislava | Institute of Virology, Biomedical Research Center of the Slovak Academy of Sciences, Bratislava; Comenius University Science Park, Bratislava | Monika Slávikova et al |
| EPI_ISL_755643; EPI_ISL_755651 | Instituto Adolfo Lutz - Central | Instituto Adolfo Lutz, Interdiciplinary Procedures Center, Strategic Laboratory | Claudio Tavares Sacchi et al B |
| EPI_ISL_755641; EPI_ISL_755647; EPI_ISL_755646 | Instituto Adolfo Lutz - Regional de Santo Andre | Instituto Adolfo Lutz, Interdiciplinary Procedures Center, Strategic Laboratory | Claudio Tavares Sacchi et al B |
| EPI_ISL_491203; EPI_ISL_491224; EPI_ISL_491230; EPI_ISL_491204 | Instituto Gulbenkian de Ciência | Instituto Gulbenkian de Ciência | Cathy Paulino et al |
| EPI_ISL_491246; EPI_ISL_491248; EPI_ISL_491250; EPI_ISL_491242; EPI_ISL_491231; EPI_ISL_491259; EPI_ISL_491262; EPI_ISL_491233; EPI_ISL_491234 | Instituto Gulbenkian de Ciência | Instituto Gulbenkian de Ciência | Joao Sobral et al |
| EPI_ISL_491185; EPI_ISL_491189; EPI_ISL_491190; EPI_ISL_491191 | Instituto Gulbenkian de Ciência | Instituto Gulbenkian de Ciência | João Costa et al |
| EPI_ISL_491297; EPI_ISL_491275; EPI_ISL_491288; EPI_ISL_491268; EPI_ISL_491280; EPI_ISL_491277; EPI_ISL_491281; EPI_ISL_491294 | Instituto Gulbenkian de Ciência | Instituto Gulbenkian de Ciência | Susana Ladeiro et al |
| EPI_ISL_426362 | Instituto Nacional de Ciencias Medicas y Nutricion Salvador Zubiran | Instituto Nacional de Ciencias Medicas y Nutricion Salvador Zubiran | Guillermo M. Ruiz-Palacios et al (https://dx.doi.org/10.1101/2020.05.27.120402) |
| EPI_ISL_412972 | Instituto Nacional de Enfermedades Respiratorias | Instituto de Diagnostico y Referencia Epidemiologicos (INDRE) | Ramirez-Gonzalez Ernesto et al B (https://dx.doi.org/10.1007/s00705-020-04695-3) |
| EPI_ISL_574294 | Instituto Nacional de Investigacion en Salud Pública | Instituto Nacional de Investigación en Salud Pública | Leandro Patino Patino et al |
| EPI_ISL_574293 | Instituto Nacional de Investigación en Salud Pública | Instituto Nacional de Investigación en Salud Publica | Leandro Patino Patino et al |
| EPI_ISL_574292 | Instituto Nacional de Investigación en Salud Pública | Instituto Nacional de Investigación en Salud Pública | Leandro Patino Patino et al |
| EPI_ISL_491953; EPI_ISL_491950; EPI_ISL_491949; EPI_ISL_491947; EPI_ISL_491945; EPI_ISL_491944; EPI_ISL_491946; EPI_ISL_491948 | Instituto Nacional de Investigación en Salud Pública - INSPI | INSPI - Charité | Alfredo Bruno Caicedo et al |
| EPI_ISL_522879 | Instituto Nacional de Medicina Genómica | Instituto Nacional de Medicina Genómica | Hidalgo-Miranda A et al |
| EPI_ISL_536545; EPI_ISL_536559; EPI_ISL_536554; EPI_ISL_536508; EPI_ISL_536550; EPI_ISL_536534; EPI_ISL_536563; EPI_ISL_536527; EPI_ISL_536487; EPI_ISL_536507; EPI_ISL_536505; EPI_ISL_536509; EPI_ISL_536520; EPI_ISL_536519; EPI_ISL_536481; EPI_ISL_536496; EPI_ISL_536495; EPI_ISL_536491; EPI_ISL_536477; EPI_ISL_536498; EPI_ISL_536490; EPI_ISL_536506; EPI_ISL_536512; EPI_ISL_536500; EPI_ISL_536511; EPI_ISL_536489; EPI_ISL_536521 | Instituto Nacional de Salud | Laboratorio de Infecciones Respiratorias Agudas | Eduardo Juscamayta Lopez et al |
| EPI_ISL_456117; EPI_ISL_456119; EPI_ISL_456124; EPI_ISL_456153; EPI_ISL_456144; EPI_ISL_456151 | Instituto Nacional de Salud - Unidad de Secuenciación y Análisis Genómico | Instituto Nacional de Salud, Universidad Cooperativa de Colombia, Instituto Alexander von Humboldt, Imperial College-London, London School of Hygiene & Tropical Medicine | Katherine Laiton-Donato et al B (https://dx.doi.org/10.1101/2020.06.02.20120782) |
| EPI_ISL_447800; EPI_ISL_447812; EPI_ISL_447777; EPI_ISL_447816 | Instituto Nacional de Salud, Bogotá, Colombia | Grupo de Investigaciones Microbiológicas-UR (GIMUR), Departamento de Biología, Facultad de Ciencias Naturales, Universidad del Rosario, Bogotá, Colombia Instituto Nacional de Salud, Bogotá, Colombia Icahn School of Medicine at Mount Sinai, New York, USA | Juan David Ramírez et al (https://www.medrxiv.org/content/10.1101/2020.06.11.20125799v1) |
| EPI_ISL_653749; EPI_ISL_653751; EPI_ISL_653761; EPI_ISL_526933; EPI_ISL_526932; EPI_ISL_653759; EPI_ISL_526971; EPI_ISL_653756; EPI_ISL_653758; EPI_ISL_739676; EPI_ISL_739675; EPI_ISL_739674; EPI_ISL_739677; EPI_ISL_653762; EPI_ISL_739672; EPI_ISL_498164; EPI_ISL_498155; EPI_ISL_653757; EPI_ISL_526967; EPI_ISL_653745; EPI_ISL_739673; EPI_ISL_653754; EPI_ISL_653760; EPI_ISL_526958; EPI_ISL_526974; EPI_ISL_526969; EPI_ISL_653755; EPI_ISL_653746; EPI_ISL_653747; EPI_ISL_739680; EPI_ISL_739671; EPI_ISL_497744; EPI_ISL_498158; EPI_ISL_498167; EPI_ISL_526949; EPI_ISL_526950; EPI_ISL_498159; EPI_ISL_498163 | Instituto Nacional de Salud, Bogotá, Colombia | Instituto Nacional de Salud, Bogotá, Colombia | Katherine Laiton-Donato et al A |
| EPI_ISL_511142; EPI_ISL_510930; EPI_ISL_511727; EPI_ISL_511105; EPI_ISL_511189; EPI_ISL_511468; EPI_ISL_738089; EPI_ISL_731990; EPI_ISL_510953; EPI_ISL_511573; EPI_ISL_759723; EPI_ISL_738110; EPI_ISL_759708; EPI_ISL_759709; EPI_ISL_759721; EPI_ISL_738103; EPI_ISL_738105; EPI_ISL_738108; EPI_ISL_738096; EPI_ISL_759761; EPI_ISL_738093; EPI_ISL_738099; EPI_ISL_738101; EPI_ISL_693574; EPI_ISL_693527; EPI_ISL_693597; EPI_ISL_693571; EPI_ISL_693594; EPI_ISL_511030; EPI_ISL_693560; EPI_ISL_693589; EPI_ISL_693598; EPI_ISL_693551; EPI_ISL_693622; EPI_ISL_693620; EPI_ISL_693615; EPI_ISL_693604; EPI_ISL_693627; EPI_ISL_693628; EPI_ISL_511683; EPI_ISL_693626; EPI_ISL_693610; EPI_ISL_693605; EPI_ISL_693592; EPI_ISL_693525; EPI_ISL_693606; EPI_ISL_693552; EPI_ISL_693553; EPI_ISL_693540; EPI_ISL_693568; EPI_ISL_693611; EPI_ISL_693567; EPI_ISL_693618; EPI_ISL_693596; EPI_ISL_693619; EPI_ISL_511060; EPI_ISL_511028; EPI_ISL_511033; EPI_ISL_738112; EPI_ISL_738113; EPI_ISL_738090; EPI_ISL_738114; EPI_ISL_759718; EPI_ISL_731902; EPI_ISL_731961; EPI_ISL_759720; EPI_ISL_732115; EPI_ISL_732034; EPI_ISL_732048; EPI_ISL_732054; EPI_ISL_731901; EPI_ISL_732015; EPI_ISL_759719 | Instituto Nacional de Saude (INSA) | Instituto Nacional de Saude (INSA) | Borges et al et al |
| EPI_ISL_511487; EPI_ISL_511508; EPI_ISL_511752; EPI_ISL_511507 | Instituto Nacional de Saude (INSA) | Instituto Nacional de Saude (INSA) and Instituto Gulbenkian de Ciencia (IGC) | Borges et al et al |
| EPI_ISL_731897; EPI_ISL_732162; EPI_ISL_732285; EPI_ISL_732195; EPI_ISL_732234; EPI_ISL_732237; EPI_ISL_732123; EPI_ISL_732213; EPI_ISL_732219; EPI_ISL_732139 | Instituto Nacional de Saude (INSA) and Instituto Gulbenkian de Ciencia (IGC) | Instituto Nacional de Saude (INSA) and Instituto Gulbenkian de Ciencia (IGC) | Borges et al et al |
| EPI_ISL_492048 | Instituto de Biologia do Exército | Laboratório Metabolismo Macromolecular FirminoTorres de Castro, Instituto de Biofísica Carlos Chagas Filho, Universidade Federal do Rio de Janeiro | Bianca Catarina Azevedo Cabral et al |
| EPI_ISL_658886; EPI_ISL_658901; EPI_ISL_576270; EPI_ISL_576269; EPI_ISL_658883; EPI_ISL_658878; EPI_ISL_658891; EPI_ISL_658880; EPI_ISL_658888; EPI_ISL_576273; EPI_ISL_493346; EPI_ISL_493345 | Instituto de Diagnostico y Referencia Epidemiologicos (INDRE) | Instituto de Diagnostico y Referencia Epidemiologicos (INDRE) | Ernesto Ramirez-Gonzalez et al |
| EPI_ISL_455435 | Instituto de Diagnostico y Referencia Epidemiologicos (INDRE) | Instituto de Diagnostico y Referencia Epidemiologicos (INDRE) | Garces-Ayala Fabiola. Taboada Ramírez Blanca. Ramirez-Gonzalez Ernesto et al |
| EPI_ISL_576261; EPI_ISL_658875; EPI_ISL_658870; EPI_ISL_658865; EPI_ISL_516611; EPI_ISL_493338 | Instituto de Diagnostico y Referencia Epidemiologicos (INDRE) | Instituto de Diagnostico y Referencia Epidemiologicos (INDRE) | Gisela Barrera-Badillo et al |
| EPI_ISL_455456; EPI_ISL_455455 | Instituto de Diagnostico y Referencia Epidemiologicos (INDRE) | Instituto de Diagnostico y Referencia Epidemiologicos (INDRE) | Rodriguez-Maldonado Abril et al |
| EPI_ISL_452141; EPI_ISL_452139 | Instituto de Diagnostico y Referencia Epidemiologicos (INDRE) | Instituto de diagnóstico y Referencia Epidemiologicos (INDRE) | Ramirez-Gonzalez Ernesto et al A |
| EPI_ISL_660068 | Instituto de Diagnostico y Referencia Epidemiologicos (INDRE) | Instituto de diagnóstico y Referencia Epidemiologicos (INDRE) Departamento de Virología | Gisela Barrera-Badillo et al |
| EPI_ISL_748142; EPI_ISL_748144; EPI_ISL_748140; EPI_ISL_748145 | Instituto de Investigaciones Biológicas Clemente Estable | Institut Pasteur de Montevideo | Daiana Mir et al |
| EPI_ISL_510075; EPI_ISL_510068 | Instituto de Investigaciones Biomédicas de Barcelona (CSIC), Hospital Clinic i Provincial de Barcelona, Instituto de Biomedicina de Valencia (CSIC), Hospital de Sant Pau | SeqCOVID-SPAIN consortium/IBV(CSIC) | Anna M. Planas et al |
| EPI_ISL_729911; EPI_ISL_729871; EPI_ISL_729904; EPI_ISL_729907; EPI_ISL_729916; EPI_ISL_729909; EPI_ISL_729908; EPI_ISL_729915; EPI_ISL_729906; EPI_ISL_729869 | Instituto de Medicina Tropical, Universidad Nacional Toribio Rodríguez de Mendoza de Amazonas | Laboratorio de Genómica Microbiana, Universidad Peruana Cayetano Heredia | Pablo Tsukayama et al |
| EPI_ISL_649087; EPI_ISL_483725; EPI_ISL_649102; EPI_ISL_745051; EPI_ISL_649094; EPI_ISL_745050; EPI_ISL_745062; EPI_ISL_745047; EPI_ISL_474965; EPI_ISL_649071; EPI_ISL_649070; EPI_ISL_475003; EPI_ISL_649075; EPI_ISL_475007; EPI_ISL_474982; EPI_ISL_514286; EPI_ISL_516902; EPI_ISL_514299; EPI_ISL_514297; EPI_ISL_514278; EPI_ISL_514282; EPI_ISL_514294; EPI_ISL_514270; EPI_ISL_514288; EPI_ISL_516903; EPI_ISL_516886; EPI_ISL_516894; EPI_ISL_516893; EPI_ISL_516887; EPI_ISL_516884; EPI_ISL_516885; EPI_ISL_516913; EPI_ISL_575334; EPI_ISL_514274; EPI_ISL_514302; EPI_ISL_745056; EPI_ISL_575333; EPI_ISL_745070; EPI_ISL_575332; EPI_ISL_649083; EPI_ISL_649072; EPI_ISL_649089; EPI_ISL_745040; EPI_ISL_745073 | Israel Central Virology laboratory | Israel Central Virology laboratory | Neta Zuckerman et al |
| EPI_ISL_594157 | Israel Institute for Biological Research | Israel Institute for Biological Research | Galia Zaide et al |
| EPI_ISL_477193 | Istituto Zooprofilattico Sperimentale Puglia e Basilicata; | Beaconlab (Bioinformatics, Evolution and Comparative Genomics lab), Dept of Biosciences, University on Mila | Parisi A. et al |
| EPI_ISL_477195; EPI_ISL_477201 | Istituto Zooprofilattico Sperimentale Puglia e Basilicata; | Beaconlab (Bioinformatics, Evolution and Comparative Genomics lab), Dept of Biosciences, University on Milan | Parisi A. et al |
| EPI_ISL_525574; EPI_ISL_527380; EPI_ISL_525565; EPI_ISL_525567; EPI_ISL_525563; EPI_ISL_525564; EPI_ISL_525554 | Istituto Zooprofilattico Sperimentale Puglia e Basilicata; Dipartimento di Bioscienze, Biotecnologie e Biofarmaceutica dell'Università degli Studi di Bari "A.Moro"; Istituto di Biomembrane. Bioenergetica e Biotecnologie Molecolari del Consiglio Nazionale delle Ricerche di Bari | Beaconlab (Bioinformatics, Evolution and Comparative Genomics lab), Dept of Biosciences, University on Milan | Parisi A. et al |
| EPI_ISL_736887; EPI_ISL_736857; EPI_ISL_736850; EPI_ISL_736854; EPI_ISL_736853; EPI_ISL_736790; EPI_ISL_736827; EPI_ISL_736847; EPI_ISL_736858; EPI_ISL_736828; EPI_ISL_736833 | Istituto Zooprofilattico Sperimentale del Mezzogiorno | TIGEM | in alphabetical order - Patrizia Annunziata et al |
| EPI_ISL_653784; EPI_ISL_653785; EPI_ISL_649191; EPI_ISL_722916; EPI_ISL_722878; EPI_ISL_722915; EPI_ISL_722904; EPI_ISL_722879; EPI_ISL_722886; EPI_ISL_722888; EPI_ISL_722907; EPI_ISL_653786; EPI_ISL_722901; EPI_ISL_722909; EPI_ISL_722911; EPI_ISL_722894 | Istituto Zooprofilattico Sperimentale della Puglia e della Basilicata | Istituto Zooprofilattico Sperimentale della Puglia e della Basilicata | Parisi A. et al |
| EPI_ISL_422427; EPI_ISL_422426 | JABER AL AHMAD AL SABAH HOSPITAL – KUWAIT CITY | Dasman Diabetes Institute | Fahd Al-Mulla et al |
| EPI_ISL_422424 | Jaber Al Ahmad Al Sabah Hospital | Dasman diabetes Institute | Fahd Al-Mulla et al |
| EPI_ISL_631767 | Jamaica Hospital Medical Center | New York City Public Health Laboratory | Jade Wang et al |
| EPI_ISL_450792; EPI_ISL_450798 | Jamaica Ministry of Health and Wellness | Pathogen Discovery, Respiratory Viruses Branch, Division of Viral Diseases, Centers for Disease Control and Prevention | Krista Queen et al A |
| EPI_ISL_450794; EPI_ISL_450793; EPI_ISL_450796; EPI_ISL_450795; EPI_ISL_450799; EPI_ISL_450797 | Jamaica Ministry of Health and Wellness | Pathogen Discovery, Respiratory Viruses Branch, Division of Viral Diseases, Centers for Disease Control and Prevention | Yan Li et al |
| EPI_ISL_451958 | Jamil-ur-Rahman Center for Genome Research, Dr. Panjwani Center for Molecular Medicine and Drug Research, International Center for Chemical and Biological Sciences, University of Karachi | Jamil-ur-Rahman Center for Genome Research, Dr. Panjwani Center for Molecular Medicine and Drug Research, International Center for Chemical and Biological Sciences, University of Karachi | Shakeel et al |
| EPI_ISL_416629; EPI_ISL_416632; EPI_ISL_416631; EPI_ISL_416569; EPI_ISL_416580 | Japanese Quarantine Stations | Pathogen Genomics Center, National Institute of Infectious Diseases | Tsuyoshi Sekizuka et al D (https://dx.doi.org/10.1101/2020.03.23.20041970) |
| EPI_ISL_728288 | Jena University Hospital, Institute for Infectious Diseases and Infection Control | Institute for infectious medicine & hospital hygiene, CaSe-Group | Spott et al |
| EPI_ISL_763088; EPI_ISL_728345; EPI_ISL_728324; EPI_ISL_728337; EPI_ISL_728338; EPI_ISL_768725; EPI_ISL_728331; EPI_ISL_728333; EPI_ISL_728321; EPI_ISL_728293; EPI_ISL_728303; EPI_ISL_728302; EPI_ISL_763087; EPI_ISL_763093; EPI_ISL_763089; EPI_ISL_763090; EPI_ISL_763092; EPI_ISL_763086; EPI_ISL_763091; EPI_ISL_728296; EPI_ISL_728298; EPI_ISL_763077; EPI_ISL_728292; EPI_ISL_728323; EPI_ISL_728294; EPI_ISL_728335; EPI_ISL_728341; EPI_ISL_768633 | Jena University Hospital, Institute for Infectious Diseases and Infection Control | Institute of infectious medicine & hospital hygiene, CaSe-Group | Spott et al |
| EPI_ISL_632938 | Jena University Hospital; Institute for Infectious Diseases and Infection Control | Institute of infectious medicine & hospital hygiene, CaSe-Group | Spott et al |
| EPI_ISL_455460 | Jiangxi Province Center for Disease Control and Prevention | Jiangxi Province Center for Disease Control and Prevention | JianXiong Li et al |
| EPI_ISL_457871; EPI_ISL_457875; EPI_ISL_457874; EPI_ISL_457916; EPI_ISL_457876; EPI_ISL_457853; EPI_ISL_457847; EPI_ISL_457845; EPI_ISL_457848; EPI_ISL_457856; EPI_ISL_457867 | KEMRI-CGMR-C | KEMRI-Wellcome Trust Research Programme/KEMRI-CGMR-C Kilifi | Githinji G. et al 2020 et al (https://virological.org/t/introduction-and-local-transmission-of-sars-cov-2-cases-in-kenya/497) |
| EPI_ISL_457932; EPI_ISL_457933; EPI_ISL_457934 | KEMRI-Centre for Virus Research | KEMRI-Wellcome Trust Research Programme/KEMRI-CGMR-C Kilifi | Githinji G. et al 2020 et al (https://virological.org/t/introduction-and-local-transmission-of-sars-cov-2-cases-in-kenya/497) |
| EPI_ISL_568702; EPI_ISL_568804; EPI_ISL_568815; EPI_ISL_568805; EPI_ISL_568843; EPI_ISL_568847; EPI_ISL_568819; EPI_ISL_568866; EPI_ISL_568867; EPI_ISL_568734; EPI_ISL_568860; EPI_ISL_568872; EPI_ISL_568708; EPI_ISL_568725; EPI_ISL_568723 | KEMRI-Wellcome Trust Research Programme/KEMRI-CGMR-C Kilifi | KEMRI-Wellcome Trust Research Programme/KEMRI-CGMR-C Kilifi | Githinji et al 2020 et al |
| EPI_ISL_660221; EPI_ISL_660229; EPI_ISL_660231; EPI_ISL_660230; EPI_ISL_660228; EPI_ISL_660258; EPI_ISL_660257 | KRISP, KZN Research Innovation and Sequencing Platform | KRISP, KZN Research Innovation and Sequencing Platform | Giandhari J et al |

| | | | |
|---|---|---|---|
| EPI_ISL_407976; EPI_ISL_415159; EPI_ISL_418797 | KU Leuven, Clinical and Epidemiological Virology | KU Leuven, Clinical and Epidemiological Virology | Bert Vanmechelen et al (https://dx.doi.org/10.1101/2020.05.05.078758) |
| EPI_ISL_417429 | KU Leuven, Clinical and Epidemiological Virology | KU Leuven, Clinical and Epidemiological Virology | Joan Marti-Carreras et al (https://dx.doi.org/10.1101/2020.05.05.078758) |
| EPI_ISL_420339; EPI_ISL_420356; EPI_ISL_420374; EPI_ISL_420426; EPI_ISL_420414 | KU Leuven, Clinical and Epidemiological Virology | KU Leuven, Clinical and Epidemiological Virology | Joan Marti-Carreras et al (https://dx.doi.org/10.1101/2020.05.05.078758) |
| EPI_ISL_464075; EPI_ISL_462193; EPI_ISL_462224; EPI_ISL_476986; EPI_ISL_476958; EPI_ISL_476955; EPI_ISL_476990; EPI_ISL_462274; EPI_ISL_462227; EPI_ISL_476993; EPI_ISL_462216 | KU Leuven, Rega Institute, Clinical and Epidemiological Virology | KU Leuven, Rega Institute, Clinical and Epidemiological Virology | Tony Wawina-Bokalanga et al |
| EPI_ISL_495448; EPI_ISL_495445; EPI_ISL_495428; EPI_ISL_495443; EPI_ISL_495429; EPI_ISL_495414; EPI_ISL_495426; EPI_ISL_495419; EPI_ISL_495427; EPI_ISL_495440 | Kafkas University, Faculty of Medicine, Department of Medical Microbiology | Kafkas University, Faculty of Medicine, Department of Medical Microbiology | Murat Karamese et al |
| EPI_ISL_745197 | Kakatiya Medical College | CSIR-Centre for Cellular and Molecular Biology | Dr.V.Sudha Rani et al |
| EPI_ISL_648164 | Kalmar klinisk mikrobiologi | The Public Health Agency of Sweden | Anna-Malin Linde et al |
| EPI_ISL_510832; EPI_ISL_510865 | Kalmar klinisk mikrobiologi | The Public Health Agency of Sweden | Oskar Karlsson Lindsjo et al |
| EPI_ISL_455871; EPI_ISL_455893; EPI_ISL_455894; EPI_ISL_454878; EPI_ISL_455899; EPI_ISL_615086 | Karolinska Universitetslaboratoriet | The Public Health Agency of Sweden | Anna-Malin Linde et al |
| EPI_ISL_475558; EPI_ISL_475544; EPI_ISL_475546; EPI_ISL_475545; EPI_ISL_475547; EPI_ISL_469077; EPI_ISL_475095; EPI_ISL_475086; EPI_ISL_510861 | Karolinska Universitetslaboratoriet | The Public Health Agency of Sweden | Oskar Karlsson Lindsjo et al |
| EPI_ISL_560984; EPI_ISL_534235; EPI_ISL_560983 | Karolinska universitetslaboratoriet SOLNA | The Public Health Agency of Sweden | Anna-Malin Linde et al |
| EPI_ISL_754062; EPI_ISL_754065; EPI_ISL_754064; EPI_ISL_754063; EPI_ISL_754067 | Kathmandu University | Nepal Health Research Council | Pradip Gyanwali et al |
| EPI_ISL_442044 | Kawsar Human Genetic Research Center | Kawsar Human Genetic Research Center | Mohammad Ali Khosravi et al |
| EPI_ISL_469294; EPI_ISL_469287; EPI_ISL_469288; EPI_ISL_469290; EPI_ISL_469292; EPI_ISL_469295; EPI_ISL_469291 | Keio University Hospital | Keio University Hospital | Kenjiro Kosaki et al A |
| EPI_ISL_438969 | Keio University School of Medicine | Keio University School of Medicine | Kenjiro Kosaki et al B (https://dx.doi.org/10.1101/2020.05.21.20107599) |
| EPI_ISL_512816; EPI_ISL_512821; EPI_ISL_512820; EPI_ISL_512819; EPI_ISL_512817; EPI_ISL_512814; EPI_ISL_512815; EPI_ISL_512812; EPI_ISL_512811; EPI_ISL_512813; EPI_ISL_512818 | Kenema Government Hospital, Ministry of Health and Sanitation | Kenema Government Hospital, Ministry of Health and Sanitation | Goba et al |
| EPI_ISL_745111 | Khayelitsha (Site B) CHC wc KHC | National Health Laboratory Service (NHLS), Tygerberg | Susan Engelbrecht et al |
| EPI_ISL_745115 | Khayelitsha Hospital wc KHA | National Health Laboratory Service (NHLS), Tygerberg | Susan Engelbrecht et al |
| EPI_ISL_489998; EPI_ISL_490011; EPI_ISL_490005; EPI_ISL_489997; EPI_ISL_490009; EPI_ISL_490010; EPI_ISL_490007; EPI_ISL_490008; EPI_ISL_490001; EPI_ISL_490006 | King Fahad Medical City | King Fahad Medical City | Alosaimi et al |
| EPI_ISL_483553; EPI_ISL_483638; EPI_ISL_483542; EPI_ISL_483551; EPI_ISL_483547; EPI_ISL_483548; EPI_ISL_483544; EPI_ISL_483543; EPI_ISL_483556; EPI_ISL_483639; EPI_ISL_483565; EPI_ISL_483559; EPI_ISL_483557; EPI_ISL_483558; EPI_ISL_483546; EPI_ISL_483549; EPI_ISL_483562; EPI_ISL_483560; EPI_ISL_483554; EPI_ISL_483563 | Kingdom of Bahrein Ministry of Health | Erasmus Medical Center | Bas Oude Munnink et al |
| EPI_ISL_538321; EPI_ISL_538351; EPI_ISL_538386; EPI_ISL_538376 | Kingston Health Sciences Centre / Queen's University | Ontario Institute for Cancer Research | Prameet M. Sheth et al |
| EPI_ISL_413021 | Klinik Hirslanden Zurich | Institute of Medical Virology, University of Zurich | Stefan Schmutz et al B (https://dx.doi.org/10.1002/jmv.25773) |
| EPI_ISL_654954 | Klinisk Mikrobiologi | The Public Health Agency of Sweden | Anna-Malin Linde et al |
| EPI_ISL_676500; EPI_ISL_676501; EPI_ISL_676492; EPI_ISL_676498 | Klinisk Mikrobiologi | The Public Health Agency of Sweden | Department of Microbiology et al |
| EPI_ISL_654893; EPI_ISL_660417; EPI_ISL_654950; EPI_ISL_654948; EPI_ISL_654949; EPI_ISL_660420; EPI_ISL_660419 | Klinisk mikrobiologi | The Public Health Agency of Sweden | Anna-Malin Linde et al |
| EPI_ISL_710602; EPI_ISL_710597; EPI_ISL_661276; EPI_ISL_661277; EPI_ISL_676502; EPI_ISL_676508; EPI_ISL_676520; EPI_ISL_661278; EPI_ISL_710603; EPI_ISL_710601; EPI_ISL_676531; EPI_ISL_676532; EPI_ISL_710599; EPI_ISL_710600; EPI_ISL_661274; EPI_ISL_732654; EPI_ISL_676507 | Klinisk mikrobiologi | The Public Health Agency of Sweden | Department of Microbiology et al |
| EPI_ISL_623092 | Klinisk mikrobiologi Lanssjukhuset Ryhov, Jonkoping | The Public Health Agency of Sweden | Anna-Malin Linde et al |
| EPI_ISL_648173; EPI_ISL_648169 | Klinisk mikrobiologi NAL Trollhattan | The Public Health Agency of Sweden | Anna-Malin Linde et al |
| EPI_ISL_510844; EPI_ISL_510870 | Klinisk mikrobiologi NAL Trollhattan | The Public Health Agency of Sweden | Oskar Karlsson Lindsjo et al |
| EPI_ISL_582806; EPI_ISL_648193; EPI_ISL_582807 | Klinisk mikrobiologi SAS Boras | The Public Health Agency of Sweden | Anna-Malin Linde et al |
| EPI_ISL_654957; EPI_ISL_582837 | Klinisk mikrobiologi Vasternorrland | The Public Health Agency of Sweden | Anna-Malin Linde et al |
| EPI_ISL_710594 | Klinisk mikrobiologi Vasternorrland | The Public Health Agency of Sweden | Department of Microbiology et al |
| EPI_ISL_510858 | Klinisk mikrobiologi Vasternorrland | The Public Health Agency of Sweden | Oskar Karlsson Lindsjo et al |
| EPI_ISL_548252 | Klinisk mikrobiologi centralsjukhuset Karlstad | The Public Health Agency of Sweden | Anna-Malin Linde et al |
| EPI_ISL_510823; EPI_ISL_510822 | Klinisk mikrobiologi centralsjukhuset Karlstad | The Public Health Agency of Sweden | Oskar Karlsson Lindsjo et al |
| EPI_ISL_429124 | Klinisk mikrobiologi och vardhygien Halmstad | The Public Health Agency of Sweden | Arne Kotz et al |
| EPI_ISL_534256 | Klinisk mikrobiologi, Laboratoriemedicin Gavleborg | The Public Health Agency of Sweden | Anna-Malin Linde et al |
| EPI_ISL_623085; EPI_ISL_623081; EPI_ISL_623080 | Klinisk mikrobiologi, Skanes universitetssjukhus, Lund | The Public Health Agency of Sweden | Anna-Malin Linde et al |
| EPI_ISL_455904 | Klinisk mikrobiologi, UAS | The Public Health Agency of Sweden | Anna-Malin Linde et al |
| EPI_ISL_534245 | Kliniskt mikrobiologiska laboratoriet | The Public Health Agency of Sweden | Anna-Malin Linde et al |
| EPI_ISL_615110; EPI_ISL_615118; EPI_ISL_660413; EPI_ISL_582773; EPI_ISL_660387; EPI_ISL_660402; EPI_ISL_594152; EPI_ISL_615117; EPI_ISL_615119 | Klinsisk mikrobiologi Linkoping | The Public Health Agency of Sweden | Anna-Malin Linde et al |
| EPI_ISL_676519; EPI_ISL_676512 | Klinsisk mikrobiologi Linkoping | The Public Health Agency of Sweden | Department of Microbiology et al |
| EPI_ISL_510874; EPI_ISL_510875 | Klinsisk mikrobiologi Linkoping | The Public Health Agency of Sweden | Oskar Karlsson Lindsjo et al |
| EPI_ISL_407193 | Korea Centers for Disease Control & Prevention (KCDC) Center for Laboratory Control of Infectious Diseases Division of Viral Diseases | Korea Centers for Disease Control & Prevention (KCDC) Center for Laboratory Control of Infectious Diseases Division of Viral Diseases | Jeong-Min Kim et al C |
| EPI_ISL_745109 | Kraaifontein CHC wc KFP | National Health Laboratory Service (NHLS), Tygerberg | Susan Engelbrecht et al |
| EPI_ISL_515181; EPI_ISL_515248; EPI_ISL_515183; EPI_ISL_515182 | Kumasi Centre for Collaborative Research in Tropical Medicine, Kumasi. | Institute of Virology, Charité – Universitätsmedizin Berlin | Augustina Sylverken et al |
| EPI_ISL_735492 | Kumilla Medical College | Central Biological Research Laboratory and Department of Biochemistry and Molecular Biology | Robiul Hasan Bhuiyan et al |
| EPI_ISL_434644 | Kungsholmsdoktorn | The Public Health Agency of Sweden | Linus Hammar et al |
| EPI_ISL_475530 | Kungsors VC | The Public Health Agency of Sweden | Oskar Karlsson Lindsjo et al |
| EPI_ISL_508931 | L'Hôpital Nord-Ouest Tarare-Grandris | CNR Virus des Infections Respiratoires - France SUD | Antonin Bal et al |
| EPI_ISL_418240 | LABM GH nord Essonne | National Reference Center for Viruses of Respiratory Infections, Institut Pasteur, Paris | Mélanie Albert et al A (https://dx.doi.org/10.1101/2020.04.24.059576) |
| EPI_ISL_445271 | LABORATORIO CLINICA CHILLAN | Instituto de Salud Publica de Chile | Andrés E Castillo et al |
| EPI_ISL_682276; EPI_ISL_682277 | LABORATORIOS LABIN | Inciensa, Instituto Costarricense de Investigación y Enseñanza en Nutrición y Salud | Francisco Duarte et al |
| EPI_ISL_717917; EPI_ISL_717834; EPI_ISL_717912 | LACEN Dr. Francisco Rimolo Neto | Bioinformatics Laboratory / LNCC | Carolina M Voloch et al |
| EPI_ISL_717903 | LACEN RJ - Noel Nutels | Bioinformatics Laboratory / LNCC | Carolina M Voloch et al |
| EPI_ISL_415128 | LACEN/ES - Laboratório Central de Saúde Pública do Espírito Santo | Instituto Oswaldo Cruz FIOCRUZ - Laboratory of Respiratory Viruses and Measles (LVRS) | Paola Resende et al A (https://dx.doi.org/10.1101/2020.06.17.158006) |
| EPI_ISL_572386; EPI_ISL_572359; EPI_ISL_572381 | LACEN/PE | WallauLab, Aggeu Magalhaes Institute | Marcelo Henrique Santos Paiva et al |
| EPI_ISL_541374 | LACEN/SE | Laboratory of Respiratory Viruses and Measles, Oswaldo Cruz Institute, FIOCRUZ | Paola Resende et al B |
| EPI_ISL_498226 | LIC | LIC | LIC et al |
| EPI_ISL_469049; EPI_ISL_469053; EPI_ISL_469051 | LNR National Reference Laboratory, Mohammed VI University of Health Sciences | Medical Biotechnology Laboratory, Rabat Medical and Pharmacy School, Mohammed The Vth University in Rabat | Meriem LAAMARTI et al A (https://www.biorxiv.org/content/10.1101/2020.06.30.181123v1) |

| | | | |
|---|---|---|---|
| EPI_ISL_482737; EPI_ISL_482740 | LNR National Reference Laboratory, Mohammed VI University of Health Sciences | Medical Biotechnology Laboratory, Rabat Medical and Pharmacy School, Mohammed The Vth University in Rabat | Meriem LAAMARTI et al B |
| EPI_ISL_728236 | LNR National Reference Laboratory, Mohammed VI University of Health Sciences | Medical Biotechnology Laboratory, Rabat Medical and Pharmacy School, Mohammed The Vth University in Rabat | Souad KARTTI et al |
| EPI_ISL_755644; EPI_ISL_755645 | Lab LOC - Itapecerica da Serra | Instituto Adolfo Lutz, Interdiciplinary Procedures Center, Strategic Laboratory | Claudio Tavares Sacchi et al B |
| EPI_ISL_481223; EPI_ISL_634878; EPI_ISL_718011; EPI_ISL_718015; EPI_ISL_718014; EPI_ISL_451164; EPI_ISL_468734; EPI_ISL_434352; EPI_ISL_717590; EPI_ISL_718000 | Lab voor klinische biologie | Onderzoeksgroep Virologie | Laurens Lambrechts et al |
| EPI_ISL_718016; EPI_ISL_451175; EPI_ISL_468740; EPI_ISL_468746; EPI_ISL_434361; EPI_ISL_717983; EPI_ISL_718005 | Lab voor klinische biologie | Onderzoeksgroep Virologie | Nick Vereecke et al A |
| EPI_ISL_425055 | Lab voor klinische biologie | Onderzoeksgroep Virologie | Nick Vereecke et al B (https://dx.doi.org/10.1101/2020.05.05.078758) |
| EPI_ISL_456202; EPI_ISL_456390 | LabPLUS | Institute of Environmental Science and Research (ESR) | Matt Storey et al B |
| EPI_ISL_755631; EPI_ISL_626356; EPI_ISL_682291; EPI_ISL_579114; EPI_ISL_661261; EPI_ISL_548109; EPI_ISL_548068; EPI_ISL_548071; EPI_ISL_548025; EPI_ISL_548049; EPI_ISL_548069; EPI_ISL_548073; EPI_ISL_548077; EPI_ISL_548072; EPI_ISL_622816; EPI_ISL_579425; EPI_ISL_755627; EPI_ISL_755632; EPI_ISL_579403; EPI_ISL_579119; EPI_ISL_622803; EPI_ISL_548108; EPI_ISL_637094; EPI_ISL_755637; EPI_ISL_582017; EPI_ISL_579109; EPI_ISL_732962; EPI_ISL_661256; EPI_ISL_661262; EPI_ISL_732965 | LabPLUS | Institute of Environmental Science and Research (ESR) | Xiaoyun Ren et al |
| EPI_ISL_548105; EPI_ISL_548104; EPI_ISL_579422 | LabTests | Institute of Environmental Science and Research (ESR) | Xiaoyun Ren et al |
| EPI_ISL_593905 | Labo Analyses Med, Sarcelles | National Reference Center for Viruses of Respiratory Infections, Institut Pasteur, Paris | Sylvie Behillil et al |
| EPI_ISL_487427; EPI_ISL_487408; EPI_ISL_487424; EPI_ISL_487411; EPI_ISL_487429; EPI_ISL_487409; EPI_ISL_487399; EPI_ISL_487417; EPI_ISL_487418; EPI_ISL_487400; EPI_ISL_487401; EPI_ISL_487426; EPI_ISL_487431; EPI_ISL_487407; EPI_ISL_487430; EPI_ISL_487404; EPI_ISL_487403; EPI_ISL_487414; EPI_ISL_487405; EPI_ISL_487398; EPI_ISL_476705; EPI_ISL_487406; EPI_ISL_487419; EPI_ISL_487428; EPI_ISL_487416; EPI_ISL_487402; EPI_ISL_487410; EPI_ISL_487412; EPI_ISL_487413; EPI_ISL_487420; EPI_ISL_487423; EPI_ISL_487425; EPI_ISL_487422; EPI_ISL_487421; EPI_ISL_487415 | Labor Kneißler GmbH & Co. KG | Heinrich Pette Institute, Leibniz Institute for Experimental Virology | Thomas Günther et al |
| EPI_ISL_428940; EPI_ISL_429754; EPI_ISL_434500; EPI_ISL_445065; EPI_ISL_445066; EPI_ISL_434495; EPI_ISL_459906; EPI_ISL_459896; EPI_ISL_459900; EPI_ISL_459893; EPI_ISL_459899; EPI_ISL_459901; EPI_ISL_445075; EPI_ISL_459894; EPI_ISL_459903; EPI_ISL_428962; EPI_ISL_434504; EPI_ISL_445067; EPI_ISL_421742; EPI_ISL_428957; EPI_ISL_428947; EPI_ISL_428960; EPI_ISL_428961; EPI_ISL_429759; EPI_ISL_428956; EPI_ISL_428943; EPI_ISL_428958; EPI_ISL_421760; EPI_ISL_434505; EPI_ISL_429769; EPI_ISL_429786; EPI_ISL_428942; EPI_ISL_421738; EPI_ISL_459902; EPI_ISL_459898; EPI_ISL_459905; EPI_ISL_421746; EPI_ISL_459895; EPI_ISL_434493; EPI_ISL_445069; EPI_ISL_421752; EPI_ISL_421753; EPI_ISL_421737 | Laboratoire National de Sante, Microbiology, Virology | Laboratoire National de Sante, Microbiology, Epidemiology and Microbial Genomics | Anke Wienecke-Baldacchino et al |
| EPI_ISL_413593 | Laboratoire National de Santé | Erasmus Medical Center | David Nieuwenhuijse et al B |
| EPI_ISL_419566; EPI_ISL_419581; EPI_ISL_419583; EPI_ISL_419582; EPI_ISL_419588; EPI_ISL_419562 | Laboratoire National de Santé, Microbiology, Virology | Laboratoire National de Santé, Microbiology, Epidemiology and Microbial Genomics | Anke Wienecke-Baldacchino et al |
| EPI_ISL_417528; EPI_ISL_417530 | Laboratoire Nationale de Santé, Microbiology, Virology | Laboratoire Nationale de Santé, Microbiology, Epidemiology and Microbial Genomics | Anke Wienecke-Baldacchino et al |
| EPI_ISL_754147; EPI_ISL_754171; EPI_ISL_754172 | Laboratoire de Microbiologie | National Reference Center for Viruses of Respiratory Infections, Institut Pasteur, Paris | Marion Barbet et al B |
| EPI_ISL_443287 | Laboratoire de Microbiologie - Bât A - CH René Dubois | National Reference Center for Viruses of Respiratory Infections, Institut Pasteur, Paris | Mélanie Albert et al C |
| EPI_ISL_660474; EPI_ISL_660505; EPI_ISL_660473; EPI_ISL_660521; EPI_ISL_660507; EPI_ISL_660468; EPI_ISL_660511; EPI_ISL_660497; EPI_ISL_660519; EPI_ISL_660472; EPI_ISL_660455; EPI_ISL_660514; EPI_ISL_660513; EPI_ISL_660450; EPI_ISL_660451; EPI_ISL_660448; EPI_ISL_660453; EPI_ISL_660449; EPI_ISL_660461; EPI_ISL_660522; EPI_ISL_660475; EPI_ISL_660447; EPI_ISL_660476; EPI_ISL_660508; EPI_ISL_660499; EPI_ISL_660500; EPI_ISL_660509; EPI_ISL_660478; EPI_ISL_660487; EPI_ISL_660479; EPI_ISL_660488; EPI_ISL_660489; EPI_ISL_660512; EPI_ISL_660482; EPI_ISL_660486; EPI_ISL_660480; EPI_ISL_660484; EPI_ISL_660483; EPI_ISL_660477; EPI_ISL_660446; EPI_ISL_660452; EPI_ISL_660445; EPI_ISL_660470; EPI_ISL_660515; EPI_ISL_660516 | Laboratoire de Microbiologie CHU Souro Sanou | Centre Muraz | Abdoul-Salam Ouedraogo et al |
| EPI_ISL_712568; EPI_ISL_712065; EPI_ISL_712066; EPI_ISL_712064; EPI_ISL_712068; EPI_ISL_712067; EPI_ISL_712069 | Laboratoire de Microbiologie- CHU Habib Bourguiba – Sfax adresse | Laboratoire des Procédés de Criblage Moléculaire et Cellulaire-Centre de Biotechnologie de Sfax | Souissi et al |
| EPI_ISL_476025 | Laboratoire de Recherche et d'Analyses Médicales de la Gendarmerie Royale | Laboratoire de Recherche et d'Analyses Médicales de la Gendarmerie Royale | Sanaâ LEMRISS et al |
| EPI_ISL_476024; EPI_ISL_476026 | Laboratoire de Recherche et d'Analyses Médicales de la Gendarmerie Royale | Laboratoire de Recherche et d'Analyses Médicales de la Gendarmerie Royale | Sanaâ Lemriss et al |
| EPI_ISL_754843; EPI_ISL_754845; EPI_ISL_754846; EPI_ISL_754844 | Laboratoire de Virologie Hôpital Robert Debré | National Reference Center for Viruses of Respiratory Infections, Institut Pasteur, Paris | Marion Barbet et al B |
| EPI_ISL_414600; EPI_ISL_414623 | Laboratoire de Virologie Institut de Virologie - INSERM U 1109 Hôpitaux Universitaires de Strasbourg | National Reference Center for Viruses of Respiratory Infections, Institut Pasteur, Paris | Mélnie Albert et al (https://dx.doi.org/10.1101/2020.04.24.059576) |
| EPI_ISL_548212 | Laboratoire de Virologie, HUG | Swiss National Reference Centre for Influenza | LAUBSCHER F. et al |
| EPI_ISL_413997; EPI_ISL_414021; EPI_ISL_414022; EPI_ISL_413999; EPI_ISL_413996; EPI_ISL_414020; EPI_ISL_414019 | Laboratoire de Virologie, HUG | Swiss National Reference Centre for Influenza | LAUBSCHER Florian et al. et al |
| EPI_ISL_765479 | Laboratoire de Virologie, Hôpital Robert Debré | National Reference Center for Viruses of Respiratory Infections, Institut Pasteur, Paris | Marion Barbet et al B |
| EPI_ISL_515204; EPI_ISL_515221 | Laboratoire de microbiologie, Hopital de Verdun | Smith Laboratory, Centre de Recherche CHU Sainte-Justine | Martin Smith et al |
| EPI_ISL_476833; EPI_ISL_476829; EPI_ISL_476830; EPI_ISL_476831; EPI_ISL_476823; EPI_ISL_476828; EPI_ISL_476834; EPI_ISL_476824; EPI_ISL_476825; EPI_ISL_476826; EPI_ISL_476822; EPI_ISL_476827 | Laboratoire des Fièvres Hémorragiques Virales du Benin | Charité-Universitätsmedizin Berlin | Yadouleton et al (https://www.medrxiv.org/content/10.1101/2020.06.29.20140749v1) |
| EPI_ISL_629081; EPI_ISL_629084; EPI_ISL_629082; EPI_ISL_629083; EPI_ISL_629085 | Laboratoire du Centre Hospitalier Annecy Genevois | CNR Virus des Infections Respiratoires - France SUD | Antonin Bal et al |
| EPI_ISL_739700; EPI_ISL_740129; EPI_ISL_744295; EPI_ISL_744819 | Laboratoire national de santé, Microbiology, Virology | Laboratoire national de santé, Microbiology, Epidemiology and Microbial Genomics | Anke Wienecke-Baldacchino et al |
| EPI_ISL_740028; EPI_ISL_744139; EPI_ISL_740006; EPI_ISL_739933; EPI_ISL_744528; EPI_ISL_744291; EPI_ISL_744418; EPI_ISL_744812; EPI_ISL_745023; EPI_ISL_744655; EPI_ISL_744294; EPI_ISL_744336; EPI_ISL_739866; EPI_ISL_740189; EPI_ISL_740027; EPI_ISL_739795; EPI_ISL_740183; EPI_ISL_744456; EPI_ISL_744809; EPI_ISL_739971; EPI_ISL_740267; EPI_ISL_744894; EPI_ISL_740307; EPI_ISL_744886; EPI_ISL_744165; EPI_ISL_740226; EPI_ISL_740343; EPI_ISL_739937; EPI_ISL_740259; EPI_ISL_744766; EPI_ISL_739835; EPI_ISL_744480; EPI_ISL_744245; EPI_ISL_744958; EPI_ISL_744577; EPI_ISL_739934; EPI_ISL_744846; EPI_ISL_755589; EPI_ISL_744187; EPI_ISL_740430; EPI_ISL_740051; EPI_ISL_739978; EPI_ISL_744631; EPI_ISL_739870; EPI_ISL_740486; EPI_ISL_740005; EPI_ISL_740063; EPI_ISL_744221; EPI_ISL_740229; EPI_ISL_740471; EPI_ISL_739773; EPI_ISL_740146; EPI_ISL_740227; EPI_ISL_744681; EPI_ISL_744311; EPI_ISL_744968; EPI_ISL_739848; EPI_ISL_744720; EPI_ISL_744486; EPI_ISL_744444; EPI_ISL_740079; EPI_ISL_739834; EPI_ISL_744954; EPI_ISL_739833; EPI_ISL_744367; EPI_ISL_744500; EPI_ISL_739753; EPI_ISL_740360; EPI_ISL_744586; EPI_ISL_744724; EPI_ISL_744711; EPI_ISL_740456; EPI_ISL_740438; EPI_ISL_744598; EPI_ISL_744434; EPI_ISL_740062; EPI_ISL_740127; EPI_ISL_739733; EPI_ISL_744688; EPI_ISL_740176; EPI_ISL_740398; EPI_ISL_744246; EPI_ISL_740092; EPI_ISL_744496; EPI_ISL_744149; EPI_ISL_740222; EPI_ISL_740289; EPI_ISL_740405; EPI_ISL_739897; EPI_ISL_740039; EPI_ISL_740061; EPI_ISL_744994; EPI_ISL_740182; EPI_ISL_740441; EPI_ISL_744687; EPI_ISL_740537; EPI_ISL_739839; EPI_ISL_740527; EPI_ISL_744625; EPI_ISL_744397; EPI_ISL_739780; EPI_ISL_739840; EPI_ISL_739774; EPI_ISL_744712; EPI_ISL_739969; EPI_ISL_744691; EPI_ISL_745000; EPI_ISL_744470; EPI_ISL_739945; EPI_ISL_744178; EPI_ISL_744146; EPI_ISL_744606; EPI_ISL_744280; EPI_ISL_744435; EPI_ISL_744521; EPI_ISL_740021; EPI_ISL_740193; EPI_ISL_745017; EPI_ISL_740292; EPI_ISL_744263; EPI_ISL_744861; EPI_ISL_739698; EPI_ISL_744471; EPI_ISL_740023; EPI_ISL_744489; EPI_ISL_744515; EPI_ISL_740331; EPI_ISL_739828; EPI_ISL_740319; EPI_ISL_740025; EPI_ISL_740203; EPI_ISL_744676; EPI_ISL_739943; EPI_ISL_744400; EPI_ISL_740022; EPI_ISL_744734; EPI_ISL_740546 | Laboratoire national de santé, Microbiology, Virology | Laboratoire national de santé, Microbiology, Microbial Genomics Platform | Anke Wienecke-Baldacchino et al |
| EPI_ISL_429133 | Laboratoriemedicin | The Public Health Agency of Sweden | Olov Svartstrom et al |
| EPI_ISL_676538 | Laboratoriemedicin, Klinisk mikrobiologi | The Public Health Agency of Sweden | Department of Microbiology et al |
| EPI_ISL_613955; EPI_ISL_613706; EPI_ISL_614889 | Laboratorio Biologia Molecolare Sars Cov2 - UOC Laboratorio Analisi - Servizio Medicina di Laboratorio, Ospedale "San Francesco" - ATS-ASSL Nuoro | Laboratorio specialistico UOC Ematologia - Ospedale "San Francesco" - ATS-ASSL Nuoro | Piras Giovanna et al |
| EPI_ISL_729858; EPI_ISL_729860; EPI_ISL_729855; EPI_ISL_729852; EPI_ISL_729861; EPI_ISL_729815; EPI_ISL_729847 | Laboratorio Central de Saude Publica do Estado do Rio Grande do Sul (LACEN-RS) | Laboratory of Respiratory Viruses and Measles, Oswaldo Cruz Institute, FIOCRUZ | Paola Resende et al B |
| EPI_ISL_480321 | Laboratorio Clínico San José | Charité Virology-University of Costa Rica | Andres Moreira-Soto et al |
| EPI_ISL_751201 | Laboratorio DILAVE/MGAP-INIA-UdelaR -Tacuarembó | Institut Pasteur de Montevideo | Daiana Mir et al |
| EPI_ISL_424667 | Laboratorio Estatal de Salud Publica del Estado de México | Instituto de Diagnóstico y Referencia Epidemiológicos | Irma López Martínez et al (https://dx.doi.org/10.1101/2020.05.27.120402) |
| EPI_ISL_424670 | Laboratorio Estatal de Salud Publica del Estado de Queretaro | Instituto de Diagnóstico y Referencia Epidemiologicos | Gisela Barrera Badillo et al (https://dx.doi.org/10.1101/2020.05.27.120402) |
| EPI_ISL_480328 | Laboratorio LABIN | Charité Virology-University of Costa Rica | Andres Moreira-Soto et al |
| EPI_ISL_457965; EPI_ISL_457958; EPI_ISL_457956; EPI_ISL_457942; EPI_ISL_457952; EPI_ISL_457951; EPI_ISL_457969; EPI_ISL_480431; EPI_ISL_480429; EPI_ISL_480435; EPI_ISL_480434; EPI_ISL_480436; EPI_ISL_480437; EPI_ISL_457967; EPI_ISL_457968; EPI_ISL_480432 | Laboratorio de Biología Molecular Asociación Española Primera en Salud | Departments of Pathology and Medicine, New York University School of Medicine | Maria Victoria Elizondo et al |
| EPI_ISL_648597 | Laboratorio de Infectología Servicio de Infectología Hospital Universitario Dr. José Eleuterio González - Universidad Autónoma de Nuevo León | Laboratorio de Infectología Molecular Departamento de Bioquímica y Medicina Molecular Facultad de Medicina - Universidad Autónoma de Nuevo León | Kame A. Galán-Huerta et al |
| EPI_ISL_648318; EPI_ISL_649155; EPI_ISL_648314; EPI_ISL_648335; EPI_ISL_648333; EPI_ISL_648326; EPI_ISL_648325; EPI_ISL_648337; EPI_ISL_648367; EPI_ISL_648373; EPI_ISL_648376; EPI_ISL_648324; EPI_ISL_648338; EPI_ISL_648317; EPI_ISL_649166; EPI_ISL_649172; EPI_ISL_648359; EPI_ISL_648375; EPI_ISL_648307; EPI_ISL_648331; EPI_ISL_648341; EPI_ISL_648327; EPI_ISL_648339; EPI_ISL_648379; EPI_ISL_648345; EPI_ISL_648328; EPI_ISL_648374; EPI_ISL_648309; EPI_ISL_648352; EPI_ISL_648304; EPI_ISL_648308; EPI_ISL_648343; EPI_ISL_648370; EPI_ISL_648377; EPI_ISL_649163; EPI_ISL_648358; EPI_ISL_648362; EPI_ISL_648322; EPI_ISL_648365; EPI_ISL_648351; EPI_ISL_648356; EPI_ISL_648315; EPI_ISL_648311; EPI_ISL_648366; EPI_ISL_648372; EPI_ISL_648321 | Laboratorio de Investigaciones de Baney | University Hospital Basel, Clinical Bacteriology | Carlos Cortes et al |
| EPI_ISL_523979; EPI_ISL_527789; EPI_ISL_523954; EPI_ISL_516646 | Laboratorio de Referencia Nacional de Virus Respiratorio. Centro Nacional de Salud Publica. Instituto Nacional de Salud Peru. | Laboratorio de Referencia Nacional de Biotecnología y Biología Molecular. Centro Nacional de Salud Publica. Instituto Nacional de Salud Peru. | Carlos Padilla Rojas et al |
| EPI_ISL_514340; EPI_ISL_514339; EPI_ISL_491428; EPI_ISL_491458; EPI_ISL_491435 | Laboratorio de Referencia Nacional de Virus Respiratorio. Instituto Nacional de Salud Perú | Laboratorio de Referencia Nacional de Biotecnología y Biología Molecular. Instituto Nacional de Salud Perú | Carlos Padilla Rojas et al |
| EPI_ISL_514227 | Laboratorio de Referencia Nacional de Virus Respiratorio. Instituto Nacional de Salud. Peru | Laboratorio de Referencia Nacional de Biotecnologia y Biologia Molecular.Instituto Nacional de Salud.Peru | Carlos Padilla Rojas et al |
| EPI_ISL_540959; EPI_ISL_529067; EPI_ISL_568516; EPI_ISL_729889; EPI_ISL_540950; EPI_ISL_529077; EPI_ISL_540933; EPI_ISL_568533; EPI_ISL_568519; EPI_ISL_568543; EPI_ISL_540969; EPI_ISL_540946; EPI_ISL_568548; EPI_ISL_568554; EPI_ISL_729883; EPI_ISL_729863; EPI_ISL_729891; EPI_ISL_568532; EPI_ISL_729873; EPI_ISL_729877; EPI_ISL_729874; EPI_ISL_568518; EPI_ISL_568552; EPI_ISL_729867; EPI_ISL_729884; EPI_ISL_568555; EPI_ISL_540954; EPI_ISL_729897; EPI_ISL_529079; EPI_ISL_540962; EPI_ISL_568527 | Laboratorio de Referencia Nacional de Virus Respiratorios, Instituto Nacional de Salud Peru | Laboratorio de Genómica Microbiana, Universidad Peruana Cayetano Heredia | Pablo Tsukayama et al |
| EPI_ISL_470622; EPI_ISL_470643 | Laboratorio de Virologia Molecular / UFRJ | Bioinformatics Laboratory / LNCC | Alexandra Gerber et al (https://dx.doi.org/10.1126/science.abd2161) |
| EPI_ISL_623136; EPI_ISL_623137; EPI_ISL_717889; EPI_ISL_717871; EPI_ISL_717872; EPI_ISL_717890; EPI_ISL_717866; EPI_ISL_717883; EPI_ISL_717869; EPI_ISL_717870; EPI_ISL_717882; EPI_ISL_717876; EPI_ISL_717873; EPI_ISL_717881; EPI_ISL_717894; EPI_ISL_717896; EPI_ISL_717864; EPI_ISL_717867; EPI_ISL_717868; EPI_ISL_717893; EPI_ISL_717898; EPI_ISL_717880; EPI_ISL_717888; EPI_ISL_717884; EPI_ISL_717806; EPI_ISL_623131; EPI_ISL_717812; EPI_ISL_717808; EPI_ISL_717821; EPI_ISL_717952; EPI_ISL_717816; EPI_ISL_717817; EPI_ISL_717830; EPI_ISL_717814; EPI_ISL_717825; EPI_ISL_717815; EPI_ISL_717819; EPI_ISL_623123 | Laboratorio de Virologia Molecular / UFRJ | Bioinformatics Laboratory / LNCC | Carolina M Voloch et al |
| EPI_ISL_430814; EPI_ISL_430811; EPI_ISL_430812; EPI_ISL_430809; EPI_ISL_430807; EPI_ISL_430803; EPI_ISL_430799; EPI_ISL_430801; EPI_ISL_430804 | Laboratorio de Virología del Hospital de Niños Dr. Ricardo Gutierrez | Área de Secuenciación del Laboratorio de Virología del Hospital de Niños Dr. Ricardo Gutierrez | Nabaes Jodar et al A (https://dx.doi.org/10.1101/2020.05.23.20111443) |
| EPI_ISL_430813; EPI_ISL_430795; EPI_ISL_430800 | Laboratorio de Virología del Hospital de Niños Dr. Ricardo Gutierrez | Área de Secuenciación del Laboratorio de Virología del Hospital de Niños Dr. Ricardo Gutierrez | Nabaes Jodar et al B |

| | | | |
|---|---|---|---|
| EPI_ISL_671974 | Laboratorio de Virología y Microbiología Molecular, Depto. de Microbiología, Facultad de Medicina, Universidad de El Salvador/INS-laboratorio de Ref. Ministerio de Salud | Laboratorio de Virología y Microbiología Molecular, Depto. de Microbiología, Facultad de Medicina, Universidad de El Salvador/INS-laboratorio de Ref. Ministerio de Salud | Rivera NR et al |
| EPI_ISL_511879 | Laboratorium Kesehatan Provinsi Jawa Barat | Molecular Genetics Laboratory-Faculty of Medicine-Universitas Padjadjaran; School of Life Sciences and Technology & School of Pharmacy-Institut Teknologi Bandung; Laboratorium Kesehatan Provinsi Jawa Barat | Marselina Irasonia Tan et al |
| EPI_ISL_528745 | Laboratorium Kesehatan Provinsi Jawa Barat | School of Life Sciences and Technology & School of Pharmacy-Institut Teknologi Bandung; Molecular Genetics Laboratory-Faculty of Medicine-Universitas Padjadjaran; Laboratorium Kesehatan Provinsi Jawa Barat | Marselina Irasonia Tan et al |
| EPI_ISL_514426; EPI_ISL_541655; EPI_ISL_541651; EPI_ISL_541660; EPI_ISL_445087; EPI_ISL_445086; EPI_ISL_514430; EPI_ISL_514429; EPI_ISL_541650; EPI_ISL_541658; EPI_ISL_514427; EPI_ISL_541653; EPI_ISL_541657; EPI_ISL_541652; EPI_ISL_541659; EPI_ISL_541649; EPI_ISL_541661; EPI_ISL_514428; EPI_ISL_541654 | Laboratory Diagnostic, Veterinary Specialized Institute Kraljevo | Laboratory Diagnostic, Veterinary Specialized Institute Kraljevo | Vidanovic et al |
| EPI_ISL_754907; EPI_ISL_754904; EPI_ISL_754906; EPI_ISL_754912; EPI_ISL_754910; EPI_ISL_754905; EPI_ISL_754902; EPI_ISL_754913; EPI_ISL_754909 | Laboratory Diagnostics and Clinical Immunology of Developmental Age, Medical University of Warsaw | genXone SA, Research & Development Laboratory; The Faculty of Mathematics, Informatics and Mechanics of the University of Warsaw | Maciej Sykulski et al |
| EPI_ISL_464093; EPI_ISL_444274; EPI_ISL_452178; EPI_ISL_452179; EPI_ISL_667809; EPI_ISL_660543; EPI_ISL_660545; EPI_ISL_464094; EPI_ISL_660544 | Laboratory Medicine | Department of Laboratory Medicine, Lin-Kou Chang Gung Memorial Hospital, Taoyuan, Taiwan | Kuo-Chien Tsao et al A |
| EPI_ISL_417518; EPI_ISL_415742; EPI_ISL_415743; EPI_ISL_415741; EPI_ISL_417522; EPI_ISL_424969 | Laboratory Medicine | Department of Laboratory Medicine, Lin-Kou Chang Gung Memorial Hospital, Taoyuan, Taiwan | Kuo-Chien Tsao et al B (https://dx.doi.org/10.1080/22221751.2020.1782271) |
| EPI_ISL_411915 | Laboratory Medicine | Department of Laboratory Medicine, Lin-Kou Chang Gung Memorial Hospital, Taoyuan, Taiwan. | Kuo-Chien Tsao et al B (https://dx.doi.org/10.1080/22221751.2020.1782271) |
| EPI_ISL_445243; EPI_ISL_455477; EPI_ISL_455474; EPI_ISL_455475; EPI_ISL_455470; EPI_ISL_455472; EPI_ISL_455469; EPI_ISL_455468; EPI_ISL_455471; EPI_ISL_455476 | Laboratory for Respiratory Viruses, Cantacuzino National Military-Medical Institute for Research and Development | Cantacuzino Institute | M.Lazar et al (https://dx.doi.org/10.1101/2020.06.28.175802) |
| EPI_ISL_754238 | Laboratory for Respiratory Viruses, Cantacuzino National Military-Medical Institute for Research and Development | Cantacuzino Institute Virology | Luiza Ustea et al |
| EPI_ISL_471418; EPI_ISL_471424; EPI_ISL_471416; EPI_ISL_471417; EPI_ISL_471419; EPI_ISL_471420; EPI_ISL_471422; EPI_ISL_471423; EPI_ISL_471421 | Laboratory for Respiratory Viruses, National Influenza Centre, Cantacuzino National Military-Medical Institute for Research and Development | Cantacuzino Institute | Luiza Ustea et al |
| EPI_ISL_437690 | Laboratory for Urgent Response to Biological Threats | Institut Pasteur CIBU /ERI | V. Caro et al |
| EPI_ISL_435045; EPI_ISL_435047; EPI_ISL_435046 | Laboratory of Applied Genetics | RSE "National Center for Biotechnology" | Shevtsov et al |
| EPI_ISL_735392 | Laboratory of Genetics and Personalized Medicine, Zan Mitrev Clinic | Laboratory of Genetics and Personalized Medicine, Zan Mitrev Clinic | Kungulovski et al. et al |
| EPI_ISL_437625 | Laboratory of Genomics & Bioinformatics, Institute of Immunology and Experimental Therapy, Polish Academy of Sciences Oddzial Mikrobiologii Wojewodzkiej Stacji Sanitarno-Epidemiologicznej. | Laboratory of Genomics & Bioinformatics, Institute of Immunology and Experimental Therapy, Polish Academy of Sciences | Dorota Kujawa et al |
| EPI_ISL_452329; EPI_ISL_452360; EPI_ISL_452347; EPI_ISL_452339; EPI_ISL_452341; EPI_ISL_452343 | Laboratory of Infectious Diseases Center of Beijing Ditan Hospital | Laboratory of Infectious Diseases Center of Beijing Ditan Hospital | Siyuan Yang et al |
| EPI_ISL_417447 | Laboratory of Infectious Diseases, Department of Biomedical and Clinical Sciences L. Sacco, University of Milan | Laboratory of Infectious Diseases, Department of Biomedical and Clinical Sciences L. Sacco, University of Milan | Gianguglielmo Zehender et al (https://onlinelibrary.wiley.com/doi/abs/10.1002/jmv.25794) |
| EPI_ISL_717979 | Laboratory of Microbiology and Infectious Diseases, Faculty of Veterinary Medicine, Aristotle University of Thessaloniki, University Campus, 541 24, Thessaloniki, Greece. | Laboratory of Biology, Department of Medicine, Democritus University of Thrace, Alexandroupolis, Greece | Dovrolis N. et al |
| EPI_ISL_437907; EPI_ISL_434464; EPI_ISL_434471; EPI_ISL_434483; EPI_ISL_434466; EPI_ISL_434481; EPI_ISL_437910; EPI_ISL_437894; EPI_ISL_437879; EPI_ISL_434476; EPI_ISL_434486; EPI_ISL_434477; EPI_ISL_434465; EPI_ISL_434479; EPI_ISL_434484; EPI_ISL_434470; EPI_ISL_434478; EPI_ISL_437911; EPI_ISL_437899 | Laboratory of Microbiology, Medical School, National and Kapodistrian University of Athens | Laboratory of Biology, Department of Medicine, Democritus University of Thrace | Kassela K. et al (https://dx.doi.org/10.1101/2020.06.03.20121236) |
| EPI_ISL_654019; EPI_ISL_654020; EPI_ISL_654018; EPI_ISL_654016; EPI_ISL_654017 | Laboratory of Microbiology, National Reference Lab, Charles Nicolle Hospital; 2-University of Tunis ElManar, Faculty of Medicine of Tunis, LR99ES09, Tunis, Tunisia | 1-Clinical and Experimental Pharmacology Lab, LR16SP02, National Center of Pharmacovigilance, University of Tunis El Manar, Tunis, Tunisia. 2-Neurodegenerative diseases and psychiatric troubles, LR18SP03, Razi Hospital, University of Tunis El Manar, Tunis, Tunisia. 3- Ministry of Health, National Observatory of New and Emerging Diseases, 1006, Tunis, Tunisia | Ilhem Boutiba-Ben Boubaker et al |
| EPI_ISL_428209; EPI_ISL_451646; EPI_ISL_451647; EPI_ISL_451644; EPI_ISL_451645 | Laboratory of Molecular Biology, Diagnostyka sp. z o.o. | Laboratory of Recombinant Vaccines | Lukasz Rabalski et al |
| EPI_ISL_425228 | Laboratory of Molecular Genetics, 2nd Faculty of Medicine, Charles University in Prague, Prague, Czech Republic | Laboratory of Molecular Genetics, 2nd Faculty of Medicine, Charles University in Prague, Prague, Czech Republic | Lenka Kramná et al |
| EPI_ISL_625679; EPI_ISL_625678 | Laboratory of Molecular Medicine, University of Magallanes | Centro Asistencial Docente y de Investigacion, Universidad de Magallanes | Jorge Gonzalez et al |
| EPI_ISL_498563; EPI_ISL_498559; EPI_ISL_498561; EPI_ISL_498560 | Laboratory of Molecular Virology International Center for Genetic Engineering and Biotechnology (ICGEB) | ARGO Open Lab Platform for Genome Sequencing | Licastro D et al A |
| EPI_ISL_417421 | Laboratory of Molecular Virology International Center for Genetic Engineering and Biotechnology (ICGEB) | ARGO Open Lab Platform for Genome sequencing | Licastro D et al B (https://dx.doi.org/10.1128/JVI.00543-20) |
| EPI_ISL_525495; EPI_ISL_525496 | Laboratory of Molecular Virology of the International Centre for Genetic Engineering and Biotechnology (ICGEB) | ARGO Open Lab Platform for Genome Sequencing | Licastro D et al A |
| EPI_ISL_541351 | Laboratory of Respiratory Viruses and Measles, Oswaldo Cruz Institute, FIOCRUZ | Laboratory of Respiratory Viruses and Measles, Oswaldo Cruz Institute, FIOCRUZ | Paola Resende et al B |
| EPI_ISL_451302 | Laboratory of Virology, INMI Lazzaro Spallanzani IRCCS | Laboratory of Virology, INMI Lazzaro Spallanzani IRCCS | Cesare E.M. Gruber et al A (https://dx.doi.org/10.1002/jmv.25699) |
| EPI_ISL_454584; EPI_ISL_454599; EPI_ISL_454597; EPI_ISL_454575; EPI_ISL_454585 | Laboratory of virology, National Center of Expertise | Laboratory of molecular-genetic research, National Center for Expertise, Kazakhstan National Center for Biotechnology, Kazakhstan | Abdaliyev Askar et al |
| EPI_ISL_454577; EPI_ISL_454576; EPI_ISL_454580; EPI_ISL_454579; EPI_ISL_454582; EPI_ISL_454593 | Laboratory of virology, National Center of Expertise | Laboratory of molecular-genetic research, National Center of Expertise, Kazakhstan National Center for Biotechnology, Kazakhstan | Abdaliyev Askar et al |
| EPI_ISL_576123; EPI_ISL_576121; EPI_ISL_576119; EPI_ISL_576120; EPI_ISL_576122; EPI_ISL_576124; EPI_ISL_576125; EPI_ISL_576118; EPI_ISL_576117 | Laboratory, The Bio Arte Limited | Laboratory, The Bio Arte Limited | Biazzo et al |
| EPI_ISL_636834 | Laboratório de Imunofarmacologia - Instituto Oswaldo Cruz | Laboratório de Imunofarmacologia - Instituto Oswaldo Cruz | Souza et al |
| EPI_ISL_434669 | Lakargruppen | The Public Health Agency of Sweden | Boris Klanger et al |
| EPI_ISL_738324; EPI_ISL_738334; EPI_ISL_738329; EPI_ISL_738348; EPI_ISL_738327; EPI_ISL_738349; EPI_ISL_738333; EPI_ISL_738347; EPI_ISL_738332; EPI_ISL_738339; EPI_ISL_738345; EPI_ISL_738346; EPI_ISL_738338; EPI_ISL_738328; EPI_ISL_738319; EPI_ISL_738326; EPI_ISL_738342; EPI_ISL_738344; EPI_ISL_738341; EPI_ISL_738343 | Landstuhl Regional Medical Center | United States Air Force School of Aerospace Medicine | Anthony Fries et al |
| EPI_ISL_534238; EPI_ISL_548255 | Lanssjukhuset Kalmar | The Public Health Agency of Sweden | Anna-Malin Linde et al |
| EPI_ISL_407079 | Lapland Central Hospital | Department of Virology, University of Helsinki and Helsinki University Hospital, Helsinki, Finland | Teemu Smura et al A |
| EPI_ISL_486426; EPI_ISL_486423; EPI_ISL_512650; EPI_ISL_534205; EPI_ISL_515186; EPI_ISL_486431; EPI_ISL_512647; EPI_ISL_486433; EPI_ISL_486435; EPI_ISL_486425; EPI_ISL_437090; EPI_ISL_486424; EPI_ISL_486422; EPI_ISL_486436; EPI_ISL_437089; EPI_ISL_486428; EPI_ISL_512651; EPI_ISL_486430; EPI_ISL_512652 | Latvijas Infektoloģijas centrs | Latvian Biomedical Research and Study Centre | Ivars Silamiķelis et al |
| EPI_ISL_498552; EPI_ISL_498556; EPI_ISL_498554; EPI_ISL_498551 | Lebanese American University | Lebanese American University | Abi Habib et al |
| EPI_ISL_756619; EPI_ISL_731730; EPI_ISL_731581; EPI_ISL_731629; EPI_ISL_675379; EPI_ISL_675342; EPI_ISL_720823; EPI_ISL_730721; EPI_ISL_730779; EPI_ISL_655528; EPI_ISL_656413; EPI_ISL_731480; EPI_ISL_731526; EPI_ISL_645919 | Lighthouse Lab in Alderley Park | Wellcome Sanger Institute for the COVID-19 Genomics UK (COG-UK) Consortium | Jacquelyn Wynn et al |
| EPI_ISL_589287; EPI_ISL_567932; EPI_ISL_567942; EPI_ISL_606128; EPI_ISL_597757; EPI_ISL_597618; EPI_ISL_606203; EPI_ISL_610461 | Lighthouse Lab in Alderley Park | Wellcome Sanger Institute for the COVID-19 Genomics UK (COG-UK) consortium | Jacquelyn Wynn et al |
| EPI_ISL_552776; EPI_ISL_558208; EPI_ISL_557587; EPI_ISL_553731; EPI_ISL_559408; EPI_ISL_554772; EPI_ISL_559427; EPI_ISL_555720; EPI_ISL_554716; EPI_ISL_559311; EPI_ISL_555611; EPI_ISL_554069; EPI_ISL_559556; EPI_ISL_559612; EPI_ISL_555126; EPI_ISL_555015; EPI_ISL_554897; EPI_ISL_555609; EPI_ISL_553654; EPI_ISL_555369; EPI_ISL_552884; EPI_ISL_552971 | Lighthouse Lab in Alderley Park | Wellcome Sanger Institute for the COVID-19 Genomics UK (COG-UK) consortium | The Lighthouse Lab in Alderley Park et al |
| EPI_ISL_647527; EPI_ISL_659537; EPI_ISL_659447; EPI_ISL_645525; EPI_ISL_647482; EPI_ISL_645885 | Lighthouse Lab in Cambridge | Wellcome Sanger Institute for the COVID-19 Genomics UK (COG-UK) Consortium | Rob Howes et al |
| EPI_ISL_555993; EPI_ISL_556041; EPI_ISL_555948; EPI_ISL_556825; EPI_ISL_556848 | Lighthouse Lab in Cambridge | Wellcome Sanger Institute for the COVID-19 Genomics UK (COG-UK) consortium | Rob Howes et al |
| EPI_ISL_735987; EPI_ISL_736321; EPI_ISL_757266 | Lighthouse Lab in Glasgow | Wellcome Sanger Institute for the COVID-19 Genomics UK (COG-UK) Consortium | Harper VanSteenhouse et al |
| EPI_ISL_567508; EPI_ISL_629765; EPI_ISL_540188; EPI_ISL_530926; EPI_ISL_537081; EPI_ISL_532618; EPI_ISL_532975; EPI_ISL_532548; EPI_ISL_540370; EPI_ISL_568338; EPI_ISL_532543; EPI_ISL_598953; EPI_ISL_531046 | Lighthouse Lab in Glasgow | Wellcome Sanger Institute for the COVID-19 Genomics UK (COG-UK) consortium | Harper VanSteenhouse et al |
| EPI_ISL_540831 | Lighthouse Lab in Glasgow / MRC-University of Glasgow Centre for Virus Research | COVID-19 Genomics UK (COG-UK) Consortium | Ana da Silva Filipe et al A |
| EPI_ISL_720901; EPI_ISL_735664; EPI_ISL_735628; EPI_ISL_736256; EPI_ISL_762341 | Lighthouse Lab in Milton Keynes | Wellcome Sanger Institute for the COVID-19 Genomics UK (COG-UK) Consortium | The Lighthouse Lab in Milton Keynes et al |
| EPI_ISL_550145 | Lighthouse Lab in Milton Keynes | Wellcome Sanger Institute for the COVID-19 Genomics UK (COG-UK) consortium | The Lighthouse Lab in Alderley Park et al |
| EPI_ISL_550282; EPI_ISL_551273; EPI_ISL_550500; EPI_ISL_558284; EPI_ISL_550509; EPI_ISL_557239; EPI_ISL_581428; EPI_ISL_581388; EPI_ISL_557191; EPI_ISL_552104; EPI_ISL_551272; EPI_ISL_549938; EPI_ISL_601582; EPI_ISL_598437; EPI_ISL_630991; EPI_ISL_552410; EPI_ISL_606535; EPI_ISL_630025; EPI_ISL_598370 | Lighthouse Lab in Milton Keynes | Wellcome Sanger Institute for the COVID-19 Genomics UK (COG-UK) consortium | The Lighthouse Lab in Milton Keynes et al |
| EPI_ISL_751191; EPI_ISL_730621 | Limmattal Hospital | Institute of Medical Virology, University of Zurich | Stefan Schmutz et al A |
| EPI_ISL_648200 | Linkoping | The Public Health Agency of Sweden | Anna-Malin Linde et al |
| EPI_ISL_541852; EPI_ISL_541849; EPI_ISL_541855; EPI_ISL_541859; EPI_ISL_541860; EPI_ISL_541858; EPI_ISL_541862; EPI_ISL_541863; EPI_ISL_541854 | Lithuanian University of Health Sciences Hospital, Department of Laboratory Medicine | Lithuanian University of Health Sciences, Laboratory of Molecular Cardiology | Lukas Zemaitis et al |
| EPI_ISL_603085; EPI_ISL_603091; EPI_ISL_603096; EPI_ISL_603087; EPI_ISL_603101; EPI_ISL_603104; EPI_ISL_603098; EPI_ISL_603097; EPI_ISL_603088; EPI_ISL_603095; EPI_ISL_603120; EPI_ISL_636886; EPI_ISL_636863; EPI_ISL_636872; EPI_ISL_636866; EPI_ISL_636842; EPI_ISL_636844; EPI_ISL_636853; EPI_ISL_636856; EPI_ISL_636857; EPI_ISL_636843; EPI_ISL_636849; EPI_ISL_636848; EPI_ISL_636869; EPI_ISL_636887; EPI_ISL_636877; EPI_ISL_636878; EPI_ISL_636879; EPI_ISL_636846; EPI_ISL_636850; EPI_ISL_636851; EPI_ISL_636854; EPI_ISL_636852; EPI_ISL_636882; EPI_ISL_636855; EPI_ISL_636604; EPI_ISL_636839; EPI_ISL_636841; EPI_ISL_636867; EPI_ISL_636874; EPI_ISL_636840; EPI_ISL_636894; EPI_ISL_636902; EPI_ISL_636888; EPI_ISL_636890; EPI_ISL_636897; EPI_ISL_636892; EPI_ISL_636899; EPI_ISL_636889; EPI_ISL_636903; EPI_ISL_636893; EPI_ISL_636906; EPI_ISL_636901; EPI_ISL_636900; EPI_ISL_636898; EPI_ISL_636895; EPI_ISL_636905; EPI_ISL_636904 | Lithuanian University of Health Sciences Hospital, Department of Laboratory Medicine | Lithuanian University of Health Sciences, Molecular cardiology lab. | Lukas Zemaitis et al |
| EPI_ISL_500101; EPI_ISL_500055 | Liverpool Clinical Laboratories | COVID-19 Genomics UK (COG-UK) Consortium | Sam Haldenby et al |
| EPI_ISL_633083 | MD PHL | MD PHL | Maryland Department of Health Laboratories Administration et al |

| | | | |
|---|---|---|---|
| EPI_ISL_594137 | MDU-PHL, The Peter Doherty Institute for Infection and Immunity | MDU-PHL, The Peter Doherty Institute for Infection and Immunity | Caly et al B |
| EPI_ISL_569033; EPI_ISL_569173; EPI_ISL_569039; EPI_ISL_569036; EPI_ISL_569132; EPI_ISL_644555; EPI_ISL_569051; EPI_ISL_644547; EPI_ISL_569112; EPI_ISL_644492; EPI_ISL_568903; EPI_ISL_569495; EPI_ISL_569024; EPI_ISL_569056; EPI_ISL_569190; EPI_ISL_569196; EPI_ISL_569084; EPI_ISL_569143; EPI_ISL_569529; EPI_ISL_644403 | MEPHI, Aix Marseille University | MEPHI, Aix Marseille University | Anthony LEVASSEUR et al |
| EPI_ISL_413574 | MHC West-Brabant | Erasmus Medical Center | David Nieuwenhuijse et al A (https://dx.doi.org/10.1038/s41591-020-0997-y) |
| EPI_ISL_710592 | MLF Care AB | The Public Health Agency of Sweden | Department of Microbiology et al |
| EPI_ISL_765491 | MONTEFIORE MEDICAL CENTER LABORATORIES | Wadsworth Center, New York State Department.of Health | Kirsten St. George et al |
| EPI_ISL_428856 | MRCG at LSHTM Genomics Lab | MRCG at LSHTM Genomics lab | Sesay et al et al A (https://dx.doi.org/10.1101/2020.04.30.070771) |
| EPI_ISL_561015; EPI_ISL_561217; EPI_ISL_561022; EPI_ISL_561234; EPI_ISL_561236; EPI_ISL_561283; EPI_ISL_561148; EPI_ISL_561157; EPI_ISL_561167; EPI_ISL_561156; EPI_ISL_561077; EPI_ISL_561076; EPI_ISL_561043; EPI_ISL_561237; EPI_ISL_561238; EPI_ISL_561040; EPI_ISL_561226; EPI_ISL_561207; EPI_ISL_561215; EPI_ISL_561214; EPI_ISL_561213; EPI_ISL_561288; EPI_ISL_561289; EPI_ISL_561292; EPI_ISL_561302; EPI_ISL_561285; EPI_ISL_561094; EPI_ISL_561016; EPI_ISL_561018; EPI_ISL_561216; EPI_ISL_561028; EPI_ISL_561218 | MRCG at LSHTM Genomics lab | MRCG at LSHTM Genomics lab | Abdul Karim sesay et al |
| EPI_ISL_428857 | MRCG at LSHTM Genomics lab | MRCG at LSHTM Genomics lab | Sesay et al et al A (https://dx.doi.org/10.1101/2020.04.30.070771) |
| EPI_ISL_471158; EPI_ISL_471160; EPI_ISL_471159; EPI_ISL_471166; EPI_ISL_471162; EPI_ISL_471167; EPI_ISL_471161; EPI_ISL_471163; EPI_ISL_471164; EPI_ISL_471171; EPI_ISL_471168; EPI_ISL_471165; EPI_ISL_471170; EPI_ISL_471169 | MRCG at LSHTM Genomics lab | MRCG at LSHTM Genomics lab | Sesay et al et al B |
| EPI_ISL_428855 | MRCG at LSHTM Geomics lab | MRCG at LSHTM Genomics lab | Sesay et al et al A (https://dx.doi.org/10.1101/2020.04.30.070771) |
| EPI_ISL_414476 | MSHS Clinical Microbiology Laboratories | MSHS Pathogen Surveillance Program | Gopi Patel et al (https://dx.doi.org/10.1101/2020.04.08.20056929) |
| EPI_ISL_631301; EPI_ISL_631300; EPI_ISL_631302; EPI_ISL_632311; EPI_ISL_631289; EPI_ISL_631288; EPI_ISL_631294; EPI_ISL_631297; EPI_ISL_631277; EPI_ISL_631282; EPI_ISL_631283; EPI_ISL_631284; EPI_ISL_631303; EPI_ISL_631299; EPI_ISL_631298; EPI_ISL_631285; EPI_ISL_631279; EPI_ISL_631296; EPI_ISL_631295; EPI_ISL_631281; EPI_ISL_631293; EPI_ISL_631278; EPI_ISL_631291; EPI_ISL_631287; EPI_ISL_631286; EPI_ISL_631292; EPI_ISL_631290; EPI_ISL_631280 | MVZ DIAMEDIS Diagnostische Medizin Sennestadt GmbH | Bielefeld University | David Brandt et al |
| EPI_ISL_640223; EPI_ISL_640259; EPI_ISL_640239; EPI_ISL_640233; EPI_ISL_640227; EPI_ISL_640236; EPI_ISL_640224; EPI_ISL_640263; EPI_ISL_640243; EPI_ISL_640268; EPI_ISL_640229; EPI_ISL_640255; EPI_ISL_640253; EPI_ISL_640244; EPI_ISL_640271; EPI_ISL_640231; EPI_ISL_640232; EPI_ISL_640249; EPI_ISL_640270; EPI_ISL_640222; EPI_ISL_640258; EPI_ISL_640250; EPI_ISL_640240; EPI_ISL_640260; EPI_ISL_640264; EPI_ISL_640247; EPI_ISL_640238; EPI_ISL_640256; EPI_ISL_640245; EPI_ISL_640257; EPI_ISL_640246; EPI_ISL_640242; EPI_ISL_640266; EPI_ISL_640262; EPI_ISL_640251; EPI_ISL_640252; EPI_ISL_640237; EPI_ISL_640265; EPI_ISL_640230; EPI_ISL_640226; EPI_ISL_640261; EPI_ISL_640267; EPI_ISL_640272; EPI_ISL_640220; EPI_ISL_640219; EPI_ISL_640221; EPI_ISL_640235; EPI_ISL_640241; EPI_ISL_640248; EPI_ISL_640225; EPI_ISL_640234; EPI_ISL_640269; EPI_ISL_640254; EPI_ISL_640228 | MVZ Laborärzte Singen | MVZ Laborärzte Singen | Jonas Schmidt et al |
| EPI_ISL_672098; EPI_ISL_672099 | Madera County Department of Public Health | Chan-Zuckerberg Biohub | CZB Cliahub Consortium et al |
| EPI_ISL_582124; EPI_ISL_528744; EPI_ISL_528743 | Malaysia Genome Institute | Malaysia Genome Institute | Mohd Noor Mat Isa et al |
| EPI_ISL_682063 | Maryland Public Health Laboratory | Maryland Public Health Laboratory | Maryland Department of Health Laboratories Administration et al |
| EPI_ISL_409067 | Massachusetts Department of Public Health | Pathogen Discovery, Respiratory Viruses Branch, Division of Viral Diseases, Centers for Disease Control and Prevention | Clinton R. Paden et al (https://dx.doi.org/10.1101/2020.03.09.20032896) |
| EPI_ISL_765626; EPI_ISL_765617; EPI_ISL_765711; EPI_ISL_765733; EPI_ISL_765778; EPI_ISL_765787; EPI_ISL_765753; EPI_ISL_765802; EPI_ISL_765758 | Massachusetts General Hospital | Infectious Disease Program, Broad Institute of Harvard and MIT | Lemieux et al |
| EPI_ISL_420904; EPI_ISL_420903; EPI_ISL_420902; EPI_ISL_420899; EPI_ISL_420900; EPI_ISL_420898; EPI_ISL_420911; EPI_ISL_420908; EPI_ISL_420909; EPI_ISL_420912; EPI_ISL_420906; EPI_ISL_420905; EPI_ISL_420907; EPI_ISL_420901 | Max von Pettenkofer Institute, Virology, National Reference Center for Retroviruses, LMU Munich | Laboratory for Functional Genome Analysis, Dept. Genomics, Gene Center of the LMU Munich | Max Muenchhoff et al |
| EPI_ISL_451947; EPI_ISL_466895; EPI_ISL_466907; EPI_ISL_452103; EPI_ISL_437245; EPI_ISL_437266; EPI_ISL_466905; EPI_ISL_437274; EPI_ISL_437275; EPI_ISL_437273; EPI_ISL_437243; EPI_ISL_437359; EPI_ISL_437231; EPI_ISL_437254; EPI_ISL_437272; EPI_ISL_451942; EPI_ISL_437283; EPI_ISL_437225; EPI_ISL_437282; EPI_ISL_466906; EPI_ISL_437210; EPI_ISL_466875; EPI_ISL_437294; EPI_ISL_437277; EPI_ISL_437250; EPI_ISL_437248; EPI_ISL_437230; EPI_ISL_437247; EPI_ISL_437204; EPI_ISL_437251; EPI_ISL_437244; EPI_ISL_437246; EPI_ISL_548960; EPI_ISL_437218; EPI_ISL_466894; EPI_ISL_548954; EPI_ISL_437285; EPI_ISL_437212; EPI_ISL_466911; EPI_ISL_466901; EPI_ISL_466908; EPI_ISL_437217; EPI_ISL_466881; EPI_ISL_466896; EPI_ISL_466891; EPI_ISL_437268; EPI_ISL_437284; EPI_ISL_437264; EPI_ISL_437239; EPI_ISL_437267; EPI_ISL_437263; EPI_ISL_466892; EPI_ISL_437280; EPI_ISL_437205; EPI_ISL_451938; EPI_ISL_437256; EPI_ISL_451939; EPI_ISL_437206; EPI_ISL_437289; EPI_ISL_437269; EPI_ISL_466893; EPI_ISL_451945; EPI_ISL_451946; EPI_ISL_466909; EPI_ISL_437262; EPI_ISL_437257; EPI_ISL_548956; EPI_ISL_437236; EPI_ISL_437297; EPI_ISL_437260; EPI_ISL_437249; EPI_ISL_437271; EPI_ISL_437265; EPI_ISL_466877; EPI_ISL_437232; EPI_ISL_466882; EPI_ISL_548950; EPI_ISL_437255; EPI_ISL_437288; EPI_ISL_437220; EPI_ISL_437228; EPI_ISL_451941; EPI_ISL_451943; EPI_ISL_437253; EPI_ISL_437237; EPI_ISL_437229; EPI_ISL_437252; EPI_ISL_437219; EPI_ISL_437223; EPI_ISL_466887; EPI_ISL_437227; EPI_ISL_437296; EPI_ISL_437214; EPI_ISL_437270; EPI_ISL_437213; EPI_ISL_466876; EPI_ISL_466879; EPI_ISL_466878; EPI_ISL_451944; EPI_ISL_437222; EPI_ISL_437215; EPI_ISL_437208; EPI_ISL_466888; EPI_ISL_437209; EPI_ISL_437221; EPI_ISL_466889; EPI_ISL_437286; EPI_ISL_548952; EPI_ISL_466884; EPI_ISL_466874; EPI_ISL_451936; EPI_ISL_549025; EPI_ISL_437234; EPI_ISL_451937; EPI_ISL_437258; EPI_ISL_437279; EPI_ISL_437293; EPI_ISL_466914; EPI_ISL_437292; EPI_ISL_437291; EPI_ISL_437259; EPI_ISL_451940; EPI_ISL_437224; EPI_ISL_437233; EPI_ISL_466900; EPI_ISL_437242; EPI_ISL_466918; EPI_ISL_466912; EPI_ISL_466919; EPI_ISL_466920; EPI_ISL_466925; EPI_ISL_466923; EPI_ISL_466913; EPI_ISL_466915; EPI_ISL_466917; EPI_ISL_466921; EPI_ISL_466922; EPI_ISL_548957; EPI_ISL_548959; EPI_ISL_466916; EPI_ISL_548958; EPI_ISL_466924; EPI_ISL_437211; EPI_ISL_437240; EPI_ISL_466890; EPI_ISL_466904; EPI_ISL_437278; EPI_ISL_437281; EPI_ISL_466883; EPI_ISL_466880; EPI_ISL_437241; EPI_ISL_437295; EPI_ISL_437207; EPI_ISL_466897; EPI_ISL_466886; EPI_ISL_437290; EPI_ISL_466898; EPI_ISL_437238; EPI_ISL_452104; EPI_ISL_437287; EPI_ISL_466902; EPI_ISL_437261; EPI_ISL_548949; EPI_ISL_437235; EPI_ISL_466885; EPI_ISL_437276; EPI_ISL_548953; EPI_ISL_437226; EPI_ISL_437216; EPI_ISL_466910; EPI_ISL_466903; EPI_ISL_466899 | Max von Pettenkofer Institute, Virology, National Reference Center for Retroviruses, LMU München | Laboratory for Functional Genome Analysis, Dept. Genomics, Gene Center of the LMU Munich | Max Muenchhoff et al |
| EPI_ISL_483036; EPI_ISL_483035; EPI_ISL_482768; EPI_ISL_482767; EPI_ISL_482765; EPI_ISL_482773; EPI_ISL_482769; EPI_ISL_482760; EPI_ISL_482759; EPI_ISL_482761 | Medical Ain Shams Research Institute (MASRI), Ain Shams University | Medical Ain Shams Research Institute (MASRI), Ain Shams University | Hesham Elghazaly et al |
| EPI_ISL_476832 | Medical Biology Department, Kocaeli University | Medical Genetics Department, Kocaeli University | Savli H et al |
| EPI_ISL_515800 | Medical Diagnostics Services (MDS) | KRISP, KZN Research Innovation and Sequencing Platform | Giandhari J et al |
| EPI_ISL_635092; EPI_ISL_549039; EPI_ISL_549085; EPI_ISL_549166; EPI_ISL_493383; EPI_ISL_447839; EPI_ISL_590893; EPI_ISL_590932; EPI_ISL_590945; EPI_ISL_493382; EPI_ISL_493381; EPI_ISL_549081 | Medical Microbiology Unit, Department for Laboratory Medicine, Drammen Hospital, Vestre Viken Health Trust, | Norwegian Institute of Public Health, Department of Virology | Kathrine Stene-Johansen et al |
| EPI_ISL_707778 | Medical Research Center, Faculty of Medicine, Syarif Hidayatullah State Islamic University Jakarta | Medical Research Center, Faculty of Medicine, Syarif Hidayatullah State Islamic University Jakarta | Erike A Suwarsono et al |
| EPI_ISL_707696 | Medical Research Center, Faculty of Medicine, Syarif Hidayatullah State Islamic University Jakarta | Medical Research Center, Faculty of Medicine, Syarif Hidayatullah State Islamic University Jakarta | Laifa Hendarmin et al |
| EPI_ISL_707890 | Medical Research Center, Faculty of Medicine, Syarif Hidayatullah State Islamic University Jakarta | Medical Research Center, Faculty of Medicine, Syarif Hidayatullah State Islamic University Jakarta | Zeti Harriyati et al |
| EPI_ISL_581469; EPI_ISL_581480; EPI_ISL_581456; EPI_ISL_581482; EPI_ISL_581470; EPI_ISL_581473; EPI_ISL_581453; EPI_ISL_581454; EPI_ISL_581457; EPI_ISL_581478; EPI_ISL_581464; EPI_ISL_581465; EPI_ISL_581485; EPI_ISL_581483; EPI_ISL_581459; EPI_ISL_581479; EPI_ISL_581466; EPI_ISL_581461; EPI_ISL_581463; EPI_ISL_581481; EPI_ISL_581460; EPI_ISL_581471; EPI_ISL_581484; EPI_ISL_581452; EPI_ISL_581468; EPI_ISL_581458 | Medizinische Klinik Innere Medizin I, Universitätsklinikum Tübingen | NGS Competence Center Tübingen, Institut für Medizinische Mikrobiologie und Hygiene, Universitätsklinikum Tübingen | Angel Angelov et al |
| EPI_ISL_536411 | Medtimes Molecular Laboratory | Medtimes Molecular Laboratory | Eric Chan et al |
| EPI_ISL_507602; EPI_ISL_708603; EPI_ISL_708558; EPI_ISL_447118; EPI_ISL_436827 | Michigan Department of Health and Human Services, Bureau of Laboratories | Michigan Department of Health and Human Services, Bureau of Laboratories | Blankenship HM et al |
| EPI_ISL_426479 | Microbial Genomics Laboratory, Institut Pasteur Montevideo | Microbial Genomics Laboratory, Institut Pasteur Montevideo | Cecilia Salazar et al B (https://dx.doi.org/10.1101/2020.05.09.086223) |
| EPI_ISL_426476; EPI_ISL_426477 | Microbial Genomics Laboratory, Institut Pasteur Montevideo | Microbial Genomics Laboratory, Institut Pasteur Montevideo, Uruguay | Cecilia Salazar et al B (https://dx.doi.org/10.1101/2020.05.09.086223) |
| EPI_ISL_426584 | Microbial Genomics Laboratory, Institut Pasteur Montevideo, Uruguay | Microbial Genomics Laboratory, Institut Pasteur Montevideo, Uruguay | Cecilia Salazar et al B (https://dx.doi.org/10.1101/2020.05.09.086223) |
| EPI_ISL_480345; EPI_ISL_480335; EPI_ISL_480334; EPI_ISL_480332; EPI_ISL_480347 | Microbial Genomics Laboratory, Institut Pasteur de Montevideo | Microbial Genomics Laboratory, Institut Pasteur de Montevideo | Cecilia Salazar et al A |
| EPI_ISL_480558; EPI_ISL_593147; EPI_ISL_518063 | Microbiological Diagnostic Unit - Public Health Laboratory (MDU-PHL) | MDU-PHL | Seemann T. et al A |
| EPI_ISL_561722; EPI_ISL_562386; EPI_ISL_565596; EPI_ISL_564581; EPI_ISL_564694 | Microbiological Diagnostic Unit - Public Health Laboratory (MDU-PHL) | MDU-PHL | Seemann et al |
| EPI_ISL_430657 | Microbiological Diagnostic Unit Public Health Laboratory | Microbiological Diagnostic Unit Public Health Laboratory | Seemann T. et al B (https://dx.doi.org/10.1101/2020.05.12.20099929) |
| EPI_ISL_513925 | Microbiology & Bioinformatics and Biostatistics, Kohat University of Science and Technology (Pakistan) & Shanghai Jiao Tong University (China) | Microbiology & Bioinformatics and Biostatistics, Kohat University of Science and Technology (Pakistan) & Shanghai Jiao Tong University (China) | Khan et al |
| EPI_ISL_418269 | Microbiology and Immunology department, Pasteur institute in Ho Chi Minh city | Microbiology and Immunology department, Pasteur institute in Ho Chi Minh city | Cao et al (http://biorxiv.org/lookup/doi/10.1101/2020.04.01.019463) |
| EPI_ISL_418267 | Microbiology and Immunology department, Pasteur institute in Ho Chi Minh city | Microbiology and Immunology department, Pasteur institute in Ho Chi Minh city | Nguyen et al |
| EPI_ISL_667762; EPI_ISL_667771; EPI_ISL_667767; EPI_ISL_667775; EPI_ISL_667758; EPI_ISL_667770; EPI_ISL_667760; EPI_ISL_667768 | Microbiology, Infectious Diseases and Immunology, Centre de Recherche du Centre Hospitalier de l'Universite de Montreal | Microbiology, Infectious Diseases and Immunology, Centre de Recherche du Centre Hospitalier de l'Universite de Montreal | Benoit et al |
| EPI_ISL_613462 | Microbiology, Koc University | Microbiology, Koc University | Nurtop et al |
| EPI_ISL_486660; EPI_ISL_486657 | Microbiology, Virology and Biemergency Laboratory-ASST FBF Sacco | Microbiology, Virology and Biemergency Laboratory-ASST FBF Sacco | Rimoldi SG et al |
| EPI_ISL_486658 | Microbiology, Virology and Biemergency Laboratory-ASST FBF Sacco | Microbiology, Virology and Biemergency Laboratory-ASST FBF Sacco | Romeri F et al |
| EPI_ISL_456258 | Middlemore Hospital | Institute of Environmental Science and Research (ESR) | Matt Storey et al B |
| EPI_ISL_548130; EPI_ISL_637087; EPI_ISL_622772; EPI_ISL_548002; EPI_ISL_548004; EPI_ISL_548005; EPI_ISL_548011; EPI_ISL_548145; EPI_ISL_548132; EPI_ISL_548133; EPI_ISL_682279; EPI_ISL_649122; EPI_ISL_682278 | Middlemore Hospital | Institute of Environmental Science and Research (ESR) | Xiaoyun Ren et al |
| EPI_ISL_678276; EPI_ISL_678289 | Mikrobiologie, RARI | Mikrobiologie, RARI | Krasnov et al |
| EPI_ISL_718135; EPI_ISL_718133; EPI_ISL_718134; EPI_ISL_718165; EPI_ISL_718143; EPI_ISL_718160; EPI_ISL_718156; EPI_ISL_718132; EPI_ISL_718166; EPI_ISL_718140; EPI_ISL_718163; EPI_ISL_718139; EPI_ISL_718148; EPI_ISL_718146; EPI_ISL_718169; EPI_ISL_718170; EPI_ISL_718168; EPI_ISL_718219; EPI_ISL_718179; EPI_ISL_718181; EPI_ISL_718196; EPI_ISL_718151; EPI_ISL_718155; EPI_ISL_718152; EPI_ISL_718149; EPI_ISL_718161; EPI_ISL_718153 | Ministry of Health Hospitals | Institute of Health and Community Medicine | David Perera et al |
| EPI_ISL_437317; EPI_ISL_437323; EPI_ISL_437324; EPI_ISL_437312; EPI_ISL_437314; EPI_ISL_437318; EPI_ISL_437306; EPI_ISL_437325; EPI_ISL_437327; EPI_ISL_437334; EPI_ISL_437319; EPI_ISL_437320; EPI_ISL_437321; EPI_ISL_437313; EPI_ISL_437315 | Ministry of Health Turkey | Ministry of Health Turkey | Fatma Bayrakdar et al A |
| EPI_ISL_429868; EPI_ISL_429865; EPI_ISL_429872; EPI_ISL_428722 | Ministry of Health Turkey | Ministry of Health Turkey | Fatma Bayrakdar et al B (https://dx.doi.org/10.1101/2020.05.15.095794) |
| EPI_ISL_759964 | Mitra Keluarga Hospital Waru | Institute of Tropical Disease, Universitas Airlangga | Jezzy R Dewantari et al |
| EPI_ISL_435137; EPI_ISL_435134; EPI_ISL_435126; EPI_ISL_435121; EPI_ISL_435120; EPI_ISL_435122; EPI_ISL_435139; EPI_ISL_435131; EPI_ISL_435124; EPI_ISL_435125; EPI_ISL_435129; EPI_ISL_435130; EPI_ISL_435128 | Mohammed Bin Rashid University of Medicine and Health Sciences | Al Jalila Genomics Center | Ahmad Abou Tayoun et al A (https://dx.doi.org/10.1101/2020.05.06.080606) |
| EPI_ISL_520734; EPI_ISL_520729; EPI_ISL_520733; EPI_ISL_520737; EPI_ISL_520744; EPI_ISL_469276; EPI_ISL_469279; EPI_ISL_520678; EPI_ISL_469278; EPI_ISL_520668; EPI_ISL_520671 | Mohammed Bin Rashid University of Medicine and Health Sciences | Al Jalila Genomics Center | Ahmad Abou Tayoun et al B |

| | | | |
|---|---|---|---|
| EPI_ISL_482862; EPI_ISL_467478; EPI_ISL_467431; EPI_ISL_482714; EPI_ISL_467444 | Molecular Diagnostics Services (MDS) | KRISP, KZN Research Innovation and Sequencing Platform | Giandhari J et al |
| EPI_ISL_681696; EPI_ISL_681699; EPI_ISL_681684; EPI_ISL_681681; EPI_ISL_681674 | Molecular Medicine Laboratory, University of Magallanes | Centro Asistencial Docente y de Investigacion, Universidad de Magallanes | Jorge González et al |
| EPI_ISL_451957; EPI_ISL_430063; EPI_ISL_430018 | Molecular Pathology Division, Department of Pathology, Hong Kong Sanatorium & Hospital | Molecular Pathology Division, Department of Pathology, Hong Kong Sanatorium & Hospital | Chun Hang AU et al (https://mra.asm.org/content/9/31/e00697-20) |
| EPI_ISL_460089; EPI_ISL_460091 | Molecular Virology Unit, Fondazione IRCCS Policlinico San Matteo , Pavia | Laboratory of Virology, INMI Lazzaro Spallanzani IRCCS | Antonino Di Caro et al |
| EPI_ISL_451306 | Molecular Virology Unit, Fondazione IRCCS Policlinico San Matteo , Pavia | Laboratory of Virology, INMI Lazzaro Spallanzani IRCCS | Antonio Piralla et al |
| EPI_ISL_460079 | Molecular Virology Unit, Fondazione IRCCS Policlinico San Matteo , Pavia | Laboratory of Virology, INMI Lazzaro Spallanzani IRCCS | Barbara Bartolini et al |
| EPI_ISL_460087 | Molecular Virology Unit, Fondazione IRCCS Policlinico San Matteo , Pavia | Laboratory of Virology, INMI Lazzaro Spallanzani IRCCS | Cesare E.M. Gruber et al B |
| EPI_ISL_451309 | Molecular Virology Unit, Fondazione IRCCS Policlinico San Matteo , Pavia | Laboratory of Virology, INMI Lazzaro Spallanzani IRCCS | Fausto Baldanti et al |
| EPI_ISL_460086 | Molecular Virology Unit, Fondazione IRCCS Policlinico San Matteo , Pavia | Laboratory of Virology, INMI Lazzaro Spallanzani IRCCS | Maria R. Capobianchi et al C |
| EPI_ISL_460083 | Molecular Virology Unit, Fondazione IRCCS Policlinico San Matteo , Pavia | Laboratory of Virology, INMI Lazzaro Spallanzani IRCCS | Martina Rueca et al |
| EPI_ISL_610227; EPI_ISL_653930; EPI_ISL_754196; EPI_ISL_754198; EPI_ISL_754197; EPI_ISL_745253; EPI_ISL_745243; EPI_ISL_754203; EPI_ISL_717973; EPI_ISL_732670; EPI_ISL_610244; EPI_ISL_717965; EPI_ISL_653922 | Molecular diagnostic laboratory of Federal Budget Institution of Science "Central Research Institute of Epidemiology" of The Federal Service on Customers' Rights Protection and Human Well-being Surveillance | Group of Genomics and Postgenomic Technologies of Central Research Institute of Epidemilology | Samoilov AE et al |
| EPI_ISL_486823; EPI_ISL_486827; EPI_ISL_534337; EPI_ISL_534340; EPI_ISL_479623 | Molecular diagnostic laboratory of Federal Budget Institution of Science "Central Research Institute of Epidemiology" of The Federal Service on Customers' Rights Protection and Human Well-being Surveillance | Group of Genomics and Postgenomic Technologies of Central Research Institute of Epidemilology | Speranskaya AS et al |
| EPI_ISL_681839; EPI_ISL_614386; EPI_ISL_614382; EPI_ISL_614393; EPI_ISL_681840; EPI_ISL_614348; EPI_ISL_614349; EPI_ISL_614390; EPI_ISL_614392; EPI_ISL_614358; EPI_ISL_614357; EPI_ISL_614360; EPI_ISL_614362; EPI_ISL_681838; EPI_ISL_681834; EPI_ISL_614365; EPI_ISL_681835; EPI_ISL_614352; EPI_ISL_614375; EPI_ISL_681836; EPI_ISL_614359; EPI_ISL_614384; EPI_ISL_614391; EPI_ISL_614351; EPI_ISL_614376; EPI_ISL_614387; EPI_ISL_614380; EPI_ISL_614350; EPI_ISL_614388; EPI_ISL_681841; EPI_ISL_614385; EPI_ISL_681842; EPI_ISL_614347; EPI_ISL_614378; EPI_ISL_614379; EPI_ISL_614394; EPI_ISL_614395 | Molecular diagnostic unit for viral haemorrhagic fevers and emerging viruses, Bouaké CHU Laboratory | Project group Epidemology of Highly Pathogenic Microorganisms, Robert Koch-Institute | Chantal Akoua-Koffi et al |
| EPI_ISL_406844 | Monash Medical Centre | Collaboration between the University of Melbourne at The Peter Doherty Institute for Infection and Immunity, and the Victorian Infectious Disease Reference Laboratory | Caly et al A (https://dx.doi.org/10.5694/mja2.50569) |
| EPI_ISL_426886; EPI_ISL_426891; EPI_ISL_426885; EPI_ISL_426894; EPI_ISL_426897; EPI_ISL_426896; EPI_ISL_426889; EPI_ISL_426892; EPI_ISL_426884; EPI_ISL_426887; EPI_ISL_426895; EPI_ISL_426893 | Motol University Hospital | Institute of Applied Biotechnologies a.s. | Petr Brož et al |
| EPI_ISL_490206; EPI_ISL_490208; EPI_ISL_490205; EPI_ISL_490207 | München Klinik Schwabing | MGZ Medical Genetics Center | Dieter A. Wolf et al |
| EPI_ISL_605902; EPI_ISL_605887; EPI_ISL_605889; EPI_ISL_605888; EPI_ISL_605891; EPI_ISL_605890; EPI_ISL_605906; EPI_ISL_605903 | NGS Lab, DNA SOLUTION LTD. | NGS Lab, DNA SOLUTION LTD. | Khan et al |
| EPI_ISL_682340; EPI_ISL_682349 | NHLS Universitas Academic | UFS Virology | PA Bester et al |
| EPI_ISL_660166 | NHLS-IALCH | KRISP, KZN Research Innovation and Sequencing Platform | Gazy I et al |
| EPI_ISL_515766; EPI_ISL_535527; EPI_ISL_529781; EPI_ISL_529722; EPI_ISL_529741; EPI_ISL_602702; EPI_ISL_487301; EPI_ISL_487294; EPI_ISL_487292; EPI_ISL_515620; EPI_ISL_509353; EPI_ISL_515731; EPI_ISL_602869; EPI_ISL_495534; EPI_ISL_602880; EPI_ISL_660218; EPI_ISL_602657; EPI_ISL_602739; EPI_ISL_602671; EPI_ISL_602677; EPI_ISL_736984; EPI_ISL_535488; EPI_ISL_602791; EPI_ISL_509274; EPI_ISL_498103; EPI_ISL_529753; EPI_ISL_529761; EPI_ISL_529751; EPI_ISL_660251; EPI_ISL_602772; EPI_ISL_482727; EPI_ISL_487321; EPI_ISL_487288; EPI_ISL_602703; EPI_ISL_602699; EPI_ISL_602864; EPI_ISL_602925; EPI_ISL_736941; EPI_ISL_736969; EPI_ISL_660247; EPI_ISL_467443; EPI_ISL_467441; EPI_ISL_467442; EPI_ISL_467509; EPI_ISL_515698 | NHLS-IALCH | KRISP, KZN Research Innovation and Sequencing Platform | Giandhari J et al |
| EPI_ISL_533262 | NHSGGC West of Scotland Specialist Virology Centre / MRC-University of Glasgow Centre for Virus Research | Wellcome Sanger Institute for the COVID-19 Genomics UK (COG-UK) consortium | Ana da Silva Filipe et al A |
| EPI_ISL_420037; EPI_ISL_418241; EPI_ISL_418242 | NIC Viral Respiratory Unit - Institut Pasteur of Algeria | National Reference Center for Viruses of Respiratory Infections, Institut Pasteur, Paris | Mélanie Albert et al C |
| EPI_ISL_577702; EPI_ISL_479529; EPI_ISL_479518; EPI_ISL_577682 | NIV Influenza | NIV Influenza | Potdar V et al B |
| EPI_ISL_450324 | NIV Pune | CSIR-Centre for Cellular and Molecular Biology | Dr V A Potdar et al (https://dx.doi.org/10.1101/2020.05.31.126136) |
| EPI_ISL_422397; EPI_ISL_422387; EPI_ISL_422403; EPI_ISL_422402 | NMIMR, Department of Virology | WACCBIP, University of Ghana | Joyce M. Ngoi et al |
| EPI_ISL_416742 | NRL for Influenza, Centrum Epidemiology and Microbiology of National Institute of Public Health, Czech Republic | Charite Universitaetsmedizin Berlin, Institute of Virology | Victor M Corman et al A |
| EPI_ISL_514441 | NSTU COVID-19 Diagnostic Center | NSU Genome Research Institute (NGRI), North South University | Dr. Muhammad Maqsud Hossain et al |
| EPI_ISL_489122; EPI_ISL_489114 | NU-OMICS DNA Sequencing research facility, Northumbria University | Wellcome Sanger Institute for the COVID-19 Genomics UK (COG-UK) consortium | Chris Duncan et al |
| EPI_ISL_436057 | NYC Department of Health and Mental Hygiene | Pathogen Discovery, Respiratory Viruses Branch, Division of Viral Diseases, Centers for Disease Control and Prevention | Ying Tao et al A |
| EPI_ISL_419697; EPI_ISL_421724; EPI_ISL_427477; EPI_ISL_424959 | NYU Langone Health | Departments of Pathology and Medicine, New York University School of Medicine | Maria Aguero-Rosenfeld et al A (https://dx.doi.org/10.1101/2020.04.15.20064931) |
| EPI_ISL_444741; EPI_ISL_444767; EPI_ISL_430374; EPI_ISL_428793 | NYU Langone Health | Departments of Pathology and Medicine, New York University School of Medicine | Maria Aguero-Rosenfeld et al B |
| EPI_ISL_452235 | Narhalsan Backa vardcentral | The Public Health Agency of Sweden | Mats Olsson et al |
| EPI_ISL_582840 | Narhalsan Fjallbacka VC | The Public Health Agency of Sweden | Anna-Malin Linde et al |
| EPI_ISL_710591; EPI_ISL_732656 | Narhalsan Fjallbacka VC | The Public Health Agency of Sweden | Department of Microbiology et al |
| EPI_ISL_450829 | Narhalsan Sjobo vardcentral | The Public Health Agency of Sweden | Lovisa Hjerten et al |
| EPI_ISL_469058; EPI_ISL_475539 | Narhalsan Sjobo vardcentral | The Public Health Agency of Sweden | Oskar Karlsson Lindsjo et al |
| EPI_ISL_523962; EPI_ISL_523960; EPI_ISL_523964; EPI_ISL_523968; EPI_ISL_523966 | National Agency for Public Health, Republic of Moldova | Charite Universitatsmedizin Berlin, Institute of Virology | Victor M Corman et al A |
| EPI_ISL_454572 | National Center of Expertise | National Center for Expertise, Kazakhstan National Center for Biotechnology, Kazakhstan | Abdaliyev Askar et al |
| EPI_ISL_454571 | National Center of Expertise | National Center for Expertise, National Center for Biotechnology, Kazakhstan | Abdaliyev Askar et al |
| EPI_ISL_496536 | National Centre For Cell Science | National Centre For Cell Science | Dhiraj Paul et al |
| EPI_ISL_636759; EPI_ISL_636793; EPI_ISL_636796; EPI_ISL_636800 | National Centre for Disease control (NCDC) | NCDC/CSIR-IGIB | Mahesh S. Dhar1* et al |
| EPI_ISL_482498; EPI_ISL_482634 | National Centre for Disease control (NCDC) | NCDC/CSIR-IGIB | Pramod Kumar# et al A |
| EPI_ISL_436458; EPI_ISL_436463 | National Centre for Disease control (NCDC) | NCDC/CSIR-IGIB | Pramod Kumar# et al B (https://dx.doi.org/10.1101/2020.06.04.128751) |
| EPI_ISL_410535; EPI_ISL_407988 | National Centre for Infectious Diseases | Programme in Emerging Infectious Diseases, Duke-NUS Medical School | Danielle E Anderson et al (https://dx.doi.org/doi/10.1101/2020.03.11.987222) |
| EPI_ISL_560388; EPI_ISL_560389; EPI_ISL_560390; EPI_ISL_560387; EPI_ISL_560385 | National Health Laboratory | Botswana Institute for Technology Research and Innovation | Kefentse Arnold Tumedi et al |
| EPI_ISL_560386 | National Health Laboratory | Botswana Institute for Technology Research and innovation | Kefentse Arnold Tumedi et al |
| EPI_ISL_635001; EPI_ISL_635003; EPI_ISL_634985; EPI_ISL_635000 | National Health Laboratory Service - Inkosi Albert Luthuli Central Hospital (NHLS-IALCH) | KRISP, KZN Research Innovation and Sequencing Platform | Giandhari J et al |
| EPI_ISL_456598; EPI_ISL_456605; EPI_ISL_456610; EPI_ISL_456604; EPI_ISL_456611; EPI_ISL_456597; EPI_ISL_456608; EPI_ISL_456606; EPI_ISL_456607; EPI_ISL_456612; EPI_ISL_456600 | National Health Laboratory, Timor-Leste | Microbiological Diagnostic Unit Public Health Laboratory, The Peter Doherty Institute for Infection and Immunity | Soares da Silva et al |
| EPI_ISL_511893; EPI_ISL_455711; EPI_ISL_455718; EPI_ISL_455708; EPI_ISL_435305; EPI_ISL_455710; EPI_ISL_455712; EPI_ISL_511898; EPI_ISL_455707; EPI_ISL_455709; EPI_ISL_455700 | National Hospital of Tropical Diseases | Oxford University Clinical Research Unit, Hanoi, Vietnam | Nguyen Thi Tam et al |
| EPI_ISL_416032; EPI_ISL_416028 | National Influenza Center - Instituto Adolfo Lutz | Instituto Adolfo Lutz, Interdiciplinary Procedures Center, Strategic Laboratory | Claudio Tavares Sacchi et al A (https://dx.doi.org/10.1101/2020.05.23.20111443) |
| EPI_ISL_515525 | National Influenza Center - Instituto Adolfo Lutz | Instituto Adolfo Lutz, Interdiciplinary Procedures Center, Strategic Laboratory | Claudio Tavares Sacchi et al B |
| EPI_ISL_408668 | National Influenza Center - National Institute of Hygiene and Epidemiology (NIHE) | National Influenza Center - National Institute of Hygiene and Epidemiology (NIHE) | Ung Thi Hong Trang et al |
| EPI_ISL_486888 | National Influenza Center, Bahrain | National Influenza Center, Bahrain | AlWasti et al |
| EPI_ISL_486887 | National Influenza Center, Bahrain | National Influenza Center, Bahrain | Zaed et al |
| EPI_ISL_424361 | National Influenza Center, Indian Council of Medical Research - National Institute of Virology | Indian Council of Medical Research-National Institute of Virology, Microbial Containment Complex | Pragya D. Yadav et al B (https://www.biorxiv.org/content/10.1101/2020.05.28.122143v1) |

| | | | |
|---|---|---|---|
| EPI_ISL_421667; EPI_ISL_421671; EPI_ISL_421669; EPI_ISL_421672; EPI_ISL_421666; EPI_ISL_424365; EPI_ISL_424364 | National Influenza Center, Indian Council of Medical Research - National Institute of Virology | Indian Council of Medical Research-National Institute of Virology, Microbial Containment Complex | Pragya D. Yadav et al C (http://www.ijmr.org.in/article.asp?issn=0971-5916;year=2020;volume=151;issue=2;spage=255;epage=260;aulast=Potdar) |
| EPI_ISL_416429; EPI_ISL_660541; EPI_ISL_416427 | National Influenza Center, National Institute of Hygiene and Epidemiology (NIHE) | National Influenza Center, National Institute of Hygiene and Epidemiology (NIHE) | Le Quynh Mai et al |
| EPI_ISL_660542 | National Influenza Center, National Institute of Hygiene and Epidemiology (NIHE) | National Institute of Hygiene and Epidemiology (NIHE) | Le Quynh Mai et al |
| EPI_ISL_467781 | National Influenza Centre Romania | Charite Universitatsmedizin Berlin, Institute of Virology | Victor M Corman et al C (https://dx.doi.org/10.1101/2020.06.28.175802) |
| EPI_ISL_476843; EPI_ISL_476837; EPI_ISL_476841; EPI_ISL_476835; EPI_ISL_476847 | National Influenza Centre for Northern Greece | National Influenza Centre for Northern Greece | Maria Christoforidi et al |
| EPI_ISL_410301 | National Influenza Centre, National Public Health Laboratory, Kathmandu, Nepal | The University of Hong Kong | Ranjit Sah et al (http://biorxiv.org/lookup/doi/10.1101/2020.04.01.019463) |
| EPI_ISL_402125 | National Institute for Communicable Disease Control and Prevention (ICDC) Chinese Center for Disease Control and Prevention (China CDC) | National Institute for Communicable Disease Control and Prevention (ICDC) Chinese Center for Disease Control and Prevention (China CDC) | Zhang et al (https://dx.doi.org/10.1038/s41586-020-2008-3) |
| EPI_ISL_430297; EPI_ISL_470879 | National Institute for Communicable Diseases of the National Health Laboratory Service | National Institute for Communicable Diseases of the National Health Laboratory Service | Allam M et al |
| EPI_ISL_746827; EPI_ISL_728283; EPI_ISL_765569; EPI_ISL_746834 | National Institute for Infectious Diseases, INMI, "L. Spallanzani" IRCCS | National Institute for Infectious Diseases, INMI, "L. Spallanzani" IRCCS | B Bartolini et al |
| EPI_ISL_765572; EPI_ISL_718262 | National Institute for Infectious Diseases, INMI, "L. Spallanzani" IRCCS | National Institute for Infectious Diseases, INMI, "L. Spallanzani" IRCCS | C.E.M Gruber et al |
| EPI_ISL_746832; EPI_ISL_746830 | National Institute for Infectious Diseases, INMI, "L. Spallanzani" IRCCS | National Institute for Infectious Diseases, INMI, "L. Spallanzani" IRCCS | E Giombini et al |
| EPI_ISL_721627; EPI_ISL_746833; EPI_ISL_728280 | National Institute for Infectious Diseases, INMI, "L. Spallanzani" IRCCS | National Institute for Infectious Diseases, INMI, "L. Spallanzani" IRCCS | F Messina et al |
| EPI_ISL_746828; EPI_ISL_746835 | National Institute for Infectious Diseases, INMI, "L. Spallanzani" IRCCS | National Institute for Infectious Diseases, INMI, "L. Spallanzani" IRCCS | M Rueca et al |
| EPI_ISL_728282; EPI_ISL_735510 | National Institute for Infectious Diseases, INMI, "L. Spallanzani" IRCCS | National Institute for Infectious Diseases, INMI, "L. Spallanzani" IRCCS | M. Rueca et al |
| EPI_ISL_765568 | National Institute for Infectious Diseases, INMI, "L. Spallanzani" IRCCS | National Institute for Infectious Diseases, INMI, "L. Spallanzani" IRCCS | O Butera et al |
| EPI_ISL_469254 | National Institute for Viral Disease Control and Prevention, China CDC | Institute of Viral Disease Control and Prevention, China CDC | Wenjie Tan et al B |
| EPI_ISL_469255 | National Institute for Viral Disease Control and Prevention, China CDC | Institute of Viral Disease Control and Prevention, China CDC | Xiang Zhao et al |
| EPI_ISL_591271; EPI_ISL_591275; EPI_ISL_591273; EPI_ISL_591272; EPI_ISL_591278; EPI_ISL_591280; EPI_ISL_591279; EPI_ISL_591277; EPI_ISL_591276 | National Institute for Viral Disease Control and Prevention, China CDC | National Institute for Viral Disease Control and Prevention, China CDC | Huilai Ma et al |
| EPI_ISL_575330 | National Institute for Viral Disease Control and Prevention, China CDC | National Institute for Viral Disease Control and Prevention, China CDC | Rongbao Gao et al |
| EPI_ISL_402119 | National Institute for Viral Disease Control and Prevention, China CDC | National Institute for Viral Disease Control and Prevention, China CDC | Wenjie Tan et al A (https://dx.doi.org/10.1056/NEJMoa2001017) |
| EPI_ISL_498692; EPI_ISL_498694; EPI_ISL_498691; EPI_ISL_498693; EPI_ISL_469256 | National Institute for Viral Disease Control and Prevention, China CDC | National Institute for Viral Disease Control and Prevention, China CDC | Xiang Zhao et al |
| EPI_ISL_458133 | National Institute of Biotechnology | Bioinformatics Division, National Institute of Biotechnology | Mohammad Uzzal Hossain et al |
| EPI_ISL_709044; EPI_ISL_709542; EPI_ISL_708839; EPI_ISL_708840 | National Institute of Blood Diseases (NIBD), Molecular Biology Lab | Genomics Lab NIBD | Samina Naz Mukry et al |
| EPI_ISL_756401 | National Institute of Health Research and Development | National Institute of Health Research and Development | Adam et al |
| EPI_ISL_574433; EPI_ISL_538508 | National Institute of Health Research and Development | National Institute of Health Research and Development | Pawestri et al |
| EPI_ISL_756376 | National Institute of Health Research and Development | National Institute of Health Research and Development | Ramadhany et al |
| EPI_ISL_757287 | National Institute of Health Research and Development | National Institute of Health Research and Development | Rukminiati et al |
| EPI_ISL_747237; EPI_ISL_747233; EPI_ISL_747234 | National Institute of Health Research and Development | National Institute of Health Research and Development | Subangkit et al |
| EPI_ISL_450197 | National Institute of Health. Department of medical Sciences, Ministry of Public Health, Thailand | National Institute of Health. Department of medical Sciences, Ministry of Public Health, Thailand | Pilailuk et al A |
| EPI_ISL_498816; EPI_ISL_483694; EPI_ISL_483703; EPI_ISL_480447; EPI_ISL_466645; EPI_ISL_466644 | National Institute of Laboratory Medicine and Referral Center | Genomic Research Lab, BCSIR | Abu Sayeed Mohammad Mahmud et al |
| EPI_ISL_603242; EPI_ISL_603243; EPI_ISL_487368 | National Institute of Laboratory Medicine and Referral Center | Genomic Research Lab, BCSIR | Barna Goswami et al |
| EPI_ISL_514246; EPI_ISL_603245; EPI_ISL_603244 | National Institute of Laboratory Medicine and Referral Center | Genomic Research Lab, BCSIR | Iffat Jahan et al |
| EPI_ISL_464162; EPI_ISL_480445 | National Institute of Laboratory Medicine and Referral Center | Genomic Research Lab, BCSIR | Md. Ahasan Habib et al |
| EPI_ISL_603249; EPI_ISL_603250 | National Institute of Laboratory Medicine and Referral Center | Genomic Research Lab, BCSIR | Md. Ahashan Habib et al |
| EPI_ISL_487375; EPI_ISL_487376 | National Institute of Laboratory Medicine and Referral Center | Genomic Research Lab, BCSIR | Md. Saddam Hossain et al |
| EPI_ISL_487366; EPI_ISL_603238; EPI_ISL_498227 | National Institute of Laboratory Medicine and Referral Center | Genomic Research Lab, BCSIR | Shahina Akter et al |
| EPI_ISL_603241; EPI_ISL_603240; EPI_ISL_487363; EPI_ISL_487364 | National Institute of Laboratory Medicine and Referral Center | Genomic Research Lab, BCSIR | Tanjina Akhter Banu et al |
| EPI_ISL_539779; EPI_ISL_539778; EPI_ISL_539782; EPI_ISL_539780 | National Institute of Public Health (Czech Republic) | State Veterinary Institute Prague | Nagy et al |
| EPI_ISL_541706; EPI_ISL_541723 | National Institute of Virology, NIV Influenza | National Institute of Virology, NIV Influenza | Potdar V et al B |
| EPI_ISL_434554; EPI_ISL_434557; EPI_ISL_434556; EPI_ISL_434555; EPI_ISL_434558 | National Institutes of Health, University of the Philippines Manila | Philippine Genome Center | Carlo M. Lapid et al |
| EPI_ISL_431833 | National Institutes of Health, University of the Philippines Manila | Philippine Genome Center, University of the Philippines System | Carlo M. Lapid et al |
| EPI_ISL_449799 | National Laboratory for Health, Environment and Food | National Laboratory for Health, Environment and Food | Mahnic A. et al |
| EPI_ISL_512636; EPI_ISL_512633; EPI_ISL_512634; EPI_ISL_512616; EPI_ISL_512639; EPI_ISL_512637; EPI_ISL_512613; EPI_ISL_512640; EPI_ISL_512617; EPI_ISL_512597; EPI_ISL_512644; EPI_ISL_512605; EPI_ISL_512601; EPI_ISL_512620; EPI_ISL_512625; EPI_ISL_512621; EPI_ISL_512606; EPI_ISL_512610; EPI_ISL_512609; EPI_ISL_512608; EPI_ISL_512622; EPI_ISL_512624; EPI_ISL_512607; EPI_ISL_512611; EPI_ISL_512635; EPI_ISL_512598; EPI_ISL_512612; EPI_ISL_512627; EPI_ISL_512626; EPI_ISL_512600; EPI_ISL_512628; EPI_ISL_512599; EPI_ISL_512614; EPI_ISL_512615; EPI_ISL_512630; EPI_ISL_512631; EPI_ISL_512632; EPI_ISL_512629; EPI_ISL_512618 | National Laboratory for Influenza/Virology reference laboratory, Public Health Center of the Ministry of Health of Ukraine | Respiratory Virus Unit, Microbiology Services Colindale, Public Health England | PHE Covid Sequencing Team et al |
| EPI_ISL_671416 | National Laboratory of Virology, Szentágothai Research Centre | National Laboratory of Virology, Szentágothai Research Centre | Endre Gábor Tóth et al |
| EPI_ISL_647980; EPI_ISL_647977; EPI_ISL_647978; EPI_ISL_647971 | National Microbiology Reference Laboratory | Quadram Institute Bioscience | Thanh Le Viet et al |
| EPI_ISL_457834; EPI_ISL_457835; EPI_ISL_457836 | National Public Health Laboratory | KEMRI-Wellcome Trust Research Programme/KEMRI-CGMR-C Kilifi | Githinji G. et al 2020 et al (https://virological.org/t/introduction-and-local-transmission-of-sars-cov-2-cases-in-kenya/497) |
| EPI_ISL_416885; EPI_ISL_416886; EPI_ISL_416829; EPI_ISL_416866; EPI_ISL_416884; EPI_ISL_416907 | National Public Health Laboratory | Malaysia Genome Institute | Mohd Noor Mat Isa et al |
| EPI_ISL_754073; EPI_ISL_754072 | National Public Health Laboratory | Nepal Health Research Council | Pradip Gyanwali et al |
| EPI_ISL_536435; EPI_ISL_418993; EPI_ISL_475997; EPI_ISL_462283; EPI_ISL_462293; EPI_ISL_418996; EPI_ISL_479594; EPI_ISL_422428; EPI_ISL_418994; EPI_ISL_498578; EPI_ISL_462412; EPI_ISL_490078; EPI_ISL_493404; EPI_ISL_527362; EPI_ISL_527369; EPI_ISL_512831; EPI_ISL_490057; EPI_ISL_498573; EPI_ISL_527374; EPI_ISL_527373; EPI_ISL_536421; EPI_ISL_536420; EPI_ISL_536432; EPI_ISL_536430; EPI_ISL_462334; EPI_ISL_462398; EPI_ISL_516823; EPI_ISL_512839; EPI_ISL_512840; EPI_ISL_536442; EPI_ISL_536434; EPI_ISL_498586; EPI_ISL_512836; EPI_ISL_512835; EPI_ISL_516828; EPI_ISL_536419; EPI_ISL_536429; EPI_ISL_479597; EPI_ISL_479596; EPI_ISL_475964; EPI_ISL_428828; EPI_ISL_462413; EPI_ISL_483615 | National Public Health Laboratory, National Centre for Infectious Diseases | National Public Health Laboratory, National Centre for Infectious Diseases | Mak TM et al |
| EPI_ISL_443197; EPI_ISL_443236 | National Public Health Laboratory, National Centre for Infectious Diseases | National Public Health Laboratory, National Centre for Infectious Diseases | Mak Tze Minn et al |
| EPI_ISL_410713 | National Public Health Laboratory, National Centre for Infectious Diseases | National Public Health Laboratory, National Centre for Infectious Diseases | Octavia S et al |
| EPI_ISL_645117; EPI_ISL_693322; EPI_ISL_754096; EPI_ISL_693330; EPI_ISL_728182; EPI_ISL_693331; EPI_ISL_728194; EPI_ISL_596470; EPI_ISL_693313; EPI_ISL_728186; EPI_ISL_728193; EPI_ISL_574503; EPI_ISL_626640; EPI_ISL_596486; EPI_ISL_693310; EPI_ISL_693316; EPI_ISL_693311; EPI_ISL_626646; EPI_ISL_605819; EPI_ISL_754093; EPI_ISL_754105; EPI_ISL_693314; EPI_ISL_754075; EPI_ISL_754076; EPI_ISL_574529; EPI_ISL_728188; EPI_ISL_596483; EPI_ISL_596494; EPI_ISL_574501; EPI_ISL_596477; EPI_ISL_596475; EPI_ISL_596474; EPI_ISL_596469; EPI_ISL_626639; EPI_ISL_605822; EPI_ISL_626637 | National Public Health Laboratory, National Centre for Infectious Diseases | National Public Health Laboratory, National Centre for Infectious Diseases | Tze Minn Mak et al |
| EPI_ISL_480298; EPI_ISL_480297; EPI_ISL_480304; EPI_ISL_480302; EPI_ISL_480301; EPI_ISL_480224; EPI_ISL_480299; EPI_ISL_480305; EPI_ISL_480306; EPI_ISL_480308; EPI_ISL_480303; EPI_ISL_480300; EPI_ISL_480310; EPI_ISL_480309; EPI_ISL_480307 | National Reference Laboratory "Influenza and acute respiratory diseases" | NRL-HIV | Ivan Ivanov et al |
| EPI_ISL_625683 | National Reference Laboratory for COVID-19, Pasteur Institute of Iran | National Reference Laboratory for COVID-19, Pasteur Institute of Iran | Zahra Ahmadi et al |
| EPI_ISL_737200 | National Reference Laboratory, Nigeria Centre for Disease Control. | National Reference Laboratory, Nigeria Centre for Disease Control, Gaduwa,Abuja, Nigeria | Dr Ndodo Nnaemeka et al |
| EPI_ISL_443187; EPI_ISL_435674; EPI_ISL_435675; EPI_ISL_435677; EPI_ISL_435676 | National Virology Reference Laboratory | National Public Health Laboratory, National Centre for Infectious Diseases | Mak Tze Minn et al |
| EPI_ISL_666696 | National Virus Reference Lab (NVRL) | Irish Coronavirus Sequencing Consortium-Teagasc Grange | Matthew McCabe et al |

| | | | |
|---|---|---|---|
| EPI_ISL_605797; EPI_ISL_605075; EPI_ISL_605127; EPI_ISL_605131; EPI_ISL_605117; EPI_ISL_605064; EPI_ISL_605143; EPI_ISL_605145; EPI_ISL_605060; EPI_ISL_605081; EPI_ISL_605101; EPI_ISL_605077; EPI_ISL_605141; EPI_ISL_603266; EPI_ISL_605137; EPI_ISL_605093; EPI_ISL_605098 | National Virus Reference Laboratory | Irish Coronavirus Sequencing Consortium - Helixworks | Sachin Chalapati et al |
| EPI_ISL_671343; EPI_ISL_671351; EPI_ISL_671352; EPI_ISL_671337; EPI_ISL_671346; EPI_ISL_671344; EPI_ISL_671354 | National Virus Reference Laboratory | Irish Coronavirus Sequencing Consortium - Teagasc Moorepark | Calm Walsh et al |
| EPI_ISL_681928; EPI_ISL_681903; EPI_ISL_681889 | National Virus Reference Laboratory | Irish Coronavirus Sequencing Consortium - Teagasc Moorepark | Paul Cotter et al |
| EPI_ISL_752548; EPI_ISL_752557; EPI_ISL_752564; EPI_ISL_671921; EPI_ISL_732464; EPI_ISL_671879; EPI_ISL_671878; EPI_ISL_578216; EPI_ISL_578241; EPI_ISL_596915; EPI_ISL_578319; EPI_ISL_596922; EPI_ISL_525365; EPI_ISL_525367; EPI_ISL_525371; EPI_ISL_501260; EPI_ISL_732514; EPI_ISL_732444; EPI_ISL_735387; EPI_ISL_735385; EPI_ISL_735384; EPI_ISL_735386; EPI_ISL_735389; EPI_ISL_735390; EPI_ISL_735388; EPI_ISL_752527; EPI_ISL_752544; EPI_ISL_501261; EPI_ISL_528447; EPI_ISL_528461; EPI_ISL_578323; EPI_ISL_528458; EPI_ISL_752543; EPI_ISL_752585; EPI_ISL_752594; EPI_ISL_501270; EPI_ISL_525354; EPI_ISL_501272; EPI_ISL_525369; EPI_ISL_578223; EPI_ISL_578291; EPI_ISL_525352; EPI_ISL_525362; EPI_ISL_501273; EPI_ISL_512091; EPI_ISL_512090; EPI_ISL_512103; EPI_ISL_512104; EPI_ISL_525363; EPI_ISL_525360; EPI_ISL_578253; EPI_ISL_671877; EPI_ISL_671871; EPI_ISL_639832; EPI_ISL_671867; EPI_ISL_752511; EPI_ISL_512105; EPI_ISL_512097; EPI_ISL_512099; EPI_ISL_525386; EPI_ISL_525376; EPI_ISL_525377; EPI_ISL_525383; EPI_ISL_525394; EPI_ISL_525391; EPI_ISL_525407; EPI_ISL_528449; EPI_ISL_528479; EPI_ISL_578249; EPI_ISL_639835; EPI_ISL_639848; EPI_ISL_644610; EPI_ISL_671866; EPI_ISL_596919; EPI_ISL_578269; EPI_ISL_578270; EPI_ISL_528467; EPI_ISL_578256; EPI_ISL_578258; EPI_ISL_578232; EPI_ISL_578252; EPI_ISL_578265; EPI_ISL_639845; EPI_ISL_732460; EPI_ISL_752545; EPI_ISL_671876; EPI_ISL_732384; EPI_ISL_732394; EPI_ISL_732451; EPI_ISL_732409; EPI_ISL_752566; EPI_ISL_671912; EPI_ISL_671886; EPI_ISL_752516; EPI_ISL_671898; EPI_ISL_671897; EPI_ISL_671896; EPI_ISL_671895; EPI_ISL_596905; EPI_ISL_512092; EPI_ISL_525358; EPI_ISL_525373; EPI_ISL_671882; EPI_ISL_732459; EPI_ISL_671864; EPI_ISL_732520; EPI_ISL_732426; EPI_ISL_732516; EPI_ISL_732433; EPI_ISL_732381; EPI_ISL_578218; EPI_ISL_639850 | National Virus Reference Laboratory | National Virus Reference Laboratory | Michael Carr et al |
| EPI_ISL_493157 | National Virus Resource Center, Chinese Academy of Sciences, Wuhan 430071, China | Computational Virology Group, Center for Bacteria and Viruses Resources and Bioinformation, Wuhan Institute of Virology, Chinese Academy of Sciences, Wuhan 430071, China | Jianjun Chen et al |
| EPI_ISL_444998; EPI_ISL_445000; EPI_ISL_444999 | Naval Health Research Center | Naval Medical Research Center Biological Defense Research Directorate | Logan Voegtly et al |
| EPI_ISL_754069; EPI_ISL_754071; EPI_ISL_754068; EPI_ISL_754070 | Nepal Korea Friendship Municipality Hospital | Nepal Health Research Council | Pradip Gyanwali et al |
| EPI_ISL_678594 | Netcare | KRISP, KZN Research Innovation and Sequencing Platform | Giandhari J et al |
| EPI_ISL_515410 | Nevada State Public Health Laboratory | Nevada State Public Health Laboratory | Richard Tillett et al |
| EPI_ISL_516938; EPI_ISL_516936; EPI_ISL_516934 | Nicolae Testemitanu State University of Medicine and Pharmacy | International Centre for Genetic Engineering and Biotechnology (ICGEB) and ARGO Open Lab Platform for Genome Sequencing | Ulinici M et al |
| EPI_ISL_455422; EPI_ISL_455426 | Nigeria Centre for Disease Control | African Centre of Excellence for Genomics of Infectious Diseases (ACEGID), Redeemer's University, Ede, Osun State, Nigeria | Oluniyi P.E. et al A (https://virological.org/t/sars-cov-2-genomes-from-nigeria-reveal-community-transmission-multiple-virus-lineages-and-spike-protein-mutation-associated-with-higher-transmission-and-pathogenicity/494) |
| EPI_ISL_455424; EPI_ISL_455415; EPI_ISL_455418; EPI_ISL_455423; EPI_ISL_455413; EPI_ISL_455431 | Nigeria Centre for Disease Control (NCDC) | African Centre of Excellence for Genomics of Infectious Diseases (ACEGID), Redeemer's University, Ede, Osun State, Nigeria | Oluniyi P.E. et al A (https://virological.org/t/sars-cov-2-genomes-from-nigeria-reveal-community-transmission-multiple-virus-lineages-and-spike-protein-mutation-associated-with-higher-transmission-and-pathogenicity/494) |
| EPI_ISL_487107; EPI_ISL_487106; EPI_ISL_487110; EPI_ISL_487109 | Nigeria Centre for Disease Control (NCDC) | African Centre of Excellence for Genomics of Infectious Diseases (ACEGID), Redeemer's University, Ede, Osun State, Nigeria | Oluniyi P.E. et al B (https://virological.org/t/new-sars-cov-2-genomes-from-nigeria-reveals-dominance-of-viruses-with-spike-protein-mutation-d614g-and-additional-virus-lineages-in-circulation/527) |
| EPI_ISL_527877; EPI_ISL_527876; EPI_ISL_729921; EPI_ISL_729995; EPI_ISL_730010; EPI_ISL_729999; EPI_ISL_730000; EPI_ISL_730023; EPI_ISL_730003; EPI_ISL_729959; EPI_ISL_729997; EPI_ISL_730014; EPI_ISL_729946; EPI_ISL_527874; EPI_ISL_729924; EPI_ISL_730048; EPI_ISL_729928; EPI_ISL_729974; EPI_ISL_729970; EPI_ISL_729955; EPI_ISL_729929; EPI_ISL_730017; EPI_ISL_729933; EPI_ISL_729968; EPI_ISL_527914; EPI_ISL_729956; EPI_ISL_527889; EPI_ISL_729985; EPI_ISL_729981; EPI_ISL_729975; EPI_ISL_729984; EPI_ISL_729982; EPI_ISL_729976; EPI_ISL_729977; EPI_ISL_729945; EPI_ISL_527911; EPI_ISL_730021; EPI_ISL_729972; EPI_ISL_527893; EPI_ISL_729979; EPI_ISL_729978; EPI_ISL_729980; EPI_ISL_730031; EPI_ISL_729993; EPI_ISL_729952; EPI_ISL_729987; EPI_ISL_730042; EPI_ISL_527886; EPI_ISL_527885 | Nigeria Centre for Disease Control (NCDC) | African Centre of Excellence for Genomics of Infectious Diseases (ACEGID), Redeemer's University, Ede, Osun State, Nigeria | Oluniyi P.E. et al et al |
| EPI_ISL_745200 | Nizams Institute of Medical Sciences | CSIR-Centre for Cellular and Molecular Biology | Dr.V.Sudha Rani et al |
| EPI_ISL_420151; EPI_ISL_708101; EPI_ISL_708105; EPI_ISL_635142; EPI_ISL_668422; EPI_ISL_708104 | Nordland Hospital - Bodo, Laboratory Department, Molecular Biology Unit | Norwegian Institute of Public Health, Department of Virology | Kathrine Stene-Johansen et al |
| EPI_ISL_534243 | Norra Alvsborgs lanssjukhus | The Public Health Agency of Sweden | Anna-Malin Linde et al |
| EPI_ISL_548067; EPI_ISL_548076; EPI_ISL_579406; EPI_ISL_579407 | North Shore Hospital | Institute of Environmental Science and Research (ESR) | Xiaoyun Ren et al |
| EPI_ISL_478640; EPI_ISL_577003; EPI_ISL_478526; EPI_ISL_576997 | Northumbria University / South Tees Hospitals NHS Foundation Trust / North Cumbria Integrated Care NHS Foundation Trust / North Tees and Hartlepool NHS Foundation Trust / Newcastle Hospitals NHS Foundation Trust | COVID-19 Genomics UK (COG-UK) Consortium | Darren L Smith et al |
| EPI_ISL_708181; EPI_ISL_708051; EPI_ISL_759989 | Norwegian Institute of Public Health, Department of Virology | Norwegian Institute of Public Health, Department of Virology | Kathrine Stene-Johansen et al |
| EPI_ISL_735438 | Nuclei Acid Testing - Rwanda National Reference Laboratory | GIGA Medical Genomics | Yvan Butera et al |
| EPI_ISL_735444; EPI_ISL_735448; EPI_ISL_735437; EPI_ISL_735447; EPI_ISL_735436; EPI_ISL_735446 | Nucleic Acid Testing - Rwanda National Reference Laboratory | GIGA Medical Genomics | Yvan Butera et al |
| EPI_ISL_700486 | Nyanga CDC wc NGC | NHLS/UCT | Arash Iranzadeh et al |
| EPI_ISL_457705 | OMAN-NIC | Department of Microbiology and Immunology- SQUH | Fahad Zadjali et al |
| EPI_ISL_419558 | OR State PHL-Virology/Immunology Section | Pathogen Discovery, Respiratory Viruses Branch, Division of Viral Diseases, Centers for Disease Control and Prevention | Ying Tao et al A |
| EPI_ISL_498182; EPI_ISL_498181 | OUCRU | OUCRU | Nguyen Van Vinh Chau et al |
| EPI_ISL_450739 | OUCRU/HTD | OUCRU/HTD | Nguyen Van Vinh Chau et al |
| EPI_ISL_458116; EPI_ISL_458119; EPI_ISL_458121 | Oman National Influenza Centre | Department of Microbiology and Immunology-SQUH | Fahad Zadjali et al |
| EPI_ISL_492023; EPI_ISL_492026; EPI_ISL_491994; EPI_ISL_491969; EPI_ISL_491998; EPI_ISL_492001; EPI_ISL_492000; EPI_ISL_492022; EPI_ISL_492021; EPI_ISL_492014; EPI_ISL_492006; EPI_ISL_491979; EPI_ISL_492025; EPI_ISL_492019; EPI_ISL_492024 | Oman-NIC | Department of Microbiology and Immunology-SQUH | Fahad Zadjali et al |
| EPI_ISL_457976; EPI_ISL_457993; EPI_ISL_457978; EPI_ISL_457704; EPI_ISL_457937; EPI_ISL_457701; EPI_ISL_457990; EPI_ISL_457992; EPI_ISL_457996; EPI_ISL_457994 | Oman-NIC | Oman-NIC | Samira Al-Maruqi et al |
| EPI_ISL_491154 | Oman-National Influenza Center | Biotechnology & OMICs Laboratory | Abdul Latif Khan et al |
| EPI_ISL_525426; EPI_ISL_525423; EPI_ISL_491161; EPI_ISL_491166 | Oman-National Influenza Center | Biotechnology & OMICs Laboratory | Sajjad Asaf et al |
| EPI_ISL_491133 | Oman-National Influenza Center | Biotechnology & OMICs Laboratory | Samiha Al-Kharusi et al |
| EPI_ISL_525429; EPI_ISL_491128; EPI_ISL_491131; EPI_ISL_491130 | Oman-National Influenza Center | Biotechnology & OMICs Laboratory | Samira Al-Mahruqi et al |
| EPI_ISL_518847 | Oman-National Influenza Center | Biotechnology & OMICs Laboratory, Natural & Medical Sciences Research Center, University of Nizwa | Abdul Latif Khan et al |
| EPI_ISL_518823 | Oman-National Influenza Center | Biotechnology & OMICs Laboratory, Natural & Medical Sciences Research Center, University of Nizwa | Samira Al-Mahruqi et al |
| EPI_ISL_492065 | Oman-National Influenza Center | Department of Microbiology and Immunology-SQUH Department of Microbiology and Immunology, Sultan Qaboos University Hospital, P.O 35, Postal code 123 | Samira Al-Maruqi et al |
| EPI_ISL_569779; EPI_ISL_639913; EPI_ISL_569827; EPI_ISL_569851; EPI_ISL_569804; EPI_ISL_569843; EPI_ISL_569785; EPI_ISL_569764; EPI_ISL_569836; EPI_ISL_569808; EPI_ISL_569841; EPI_ISL_569801; EPI_ISL_639936; EPI_ISL_569848; EPI_ISL_569825; EPI_ISL_569818; EPI_ISL_569855; EPI_ISL_569748; EPI_ISL_569747; EPI_ISL_639931; EPI_ISL_639923; EPI_ISL_569766; EPI_ISL_569835; EPI_ISL_569759; EPI_ISL_569807; EPI_ISL_569816; EPI_ISL_569771; EPI_ISL_569767; EPI_ISL_639934; EPI_ISL_569828; EPI_ISL_569810; EPI_ISL_569784; EPI_ISL_569814; EPI_ISL_569794 | Omsk Research Institute of Natural Focal Infections | WHO National Influenza Centre Russian Federation | Artem Fadeev et al |
| EPI_ISL_475533; EPI_ISL_475534 | Omtanken Grimmered | The Public Health Agency of Sweden | Oskar Karlsson Lindsjo et al |
| EPI_ISL_625639 | Orange County Public Health Lab | Chan-Zuckerberg Biohub | CZB Cliahub Consortium et al |
| EPI_ISL_548637 | Orange County Public Health Laboratory | Chan-Zuckerberg Biohub | CZB Cliahub Consortium et al |
| EPI_ISL_582791; EPI_ISL_582788; EPI_ISL_660382; EPI_ISL_660379; EPI_ISL_615120; EPI_ISL_648205 | Orebro klinisk mikrobiologi | The Public Health Agency of Sweden | Anna-Malin Linde et al |
| EPI_ISL_676513; EPI_ISL_676493 | Orebro klinisk mikrobiologi | The Public Health Agency of Sweden | Department of Microbiology et al |
| EPI_ISL_475135 | Orebro klinisk mikrobiologi | The Public Health Agency of Sweden | Oskar Karlsson Lindsjo et al |
| EPI_ISL_710586 | Orsa VC | The Public Health Agency of Sweden | Department of Microbiology et al |
| EPI_ISL_475519 | Orsa VC | The Public Health Agency of Sweden | Oskar Karlsson Lindsjo et al |
| EPI_ISL_417483 | Oslo University Hospital, Department of Medical Microbiology | Norwegian Institute of Public Health | Kathrine Stene-Johansen et al |
| EPI_ISL_417484; EPI_ISL_417489; EPI_ISL_417488; EPI_ISL_417490; EPI_ISL_420135; EPI_ISL_549034; EPI_ISL_759990; EPI_ISL_471175; EPI_ISL_481218; EPI_ISL_493355; EPI_ISL_590918; EPI_ISL_590900; EPI_ISL_590930; EPI_ISL_481215; EPI_ISL_493354; EPI_ISL_493353; EPI_ISL_493389; EPI_ISL_471177; EPI_ISL_590916; EPI_ISL_493387; EPI_ISL_708175 | Oslo University Hospital, Department of Medical Microbiology | Norwegian Institute of Public Health, Department of Virology | Kathrine Stene-Johansen et al |
| EPI_ISL_745195; EPI_ISL_745194; EPI_ISL_745196 | Osmania Medical College | CSIR-Centre for Cellular and Molecular Biology | Dr.V.Sudha Rani et al |
| EPI_ISL_528924 | Ospedale "Giuseppe Mazzini"-Teramo | Istitituto Zooprofilattico Sperimentale dell'Abruzzo e Molise "G.Caporale" | Lorusso A et al |
| EPI_ISL_529015; EPI_ISL_529014; EPI_ISL_529022 | Ospedale "Ss. Annunziata" | Istitituto Zooprofilattico Sperimentale dell'Abruzzo e Molise "G.Caporale" | Lorusso A et al |
| EPI_ISL_420564 | Ospedale Civile Castel Di Sangro | Istituto Zooprofilattico Sperimentale dell'Abruzzo e Molise "G.Caporale" | Lorusso A et al |

| | | | |
|---|---|---|---|
| EPI_ISL_528992 | Ospedale SS Annunziata-Sulmona | Istituto Zooprofilattico Sperimentale dell'Abruzzo e Molise "G.Caporale" | Lorusso A et al |
| EPI_ISL_481210; EPI_ISL_590986; EPI_ISL_708112; EPI_ISL_481207; EPI_ISL_481213; EPI_ISL_635116; EPI_ISL_549092; EPI_ISL_549103; EPI_ISL_549114; EPI_ISL_549115; EPI_ISL_549118; EPI_ISL_549117; EPI_ISL_591003; EPI_ISL_449786 | Ostfold Hospital Trust - Kalnes, Centre for Laboratory Medicine, Section for gene technology and infection serology | Norwegian Institute of Public Health, Department of Virology | Kathrine Stene-Johansen et al |
| EPI_ISL_420148 | Ostfold Hospital Trust -Kalnes Centre for Laboratory Medicine Section for gene technology and infection serology | Norwegian Institute of Public Health, Department of Virology | Kathrine Stene-Johansen et al |
| EPI_ISL_535021; EPI_ISL_573216; EPI_ISL_573218; EPI_ISL_573637; EPI_ISL_559879; EPI_ISL_573169; EPI_ISL_595771; EPI_ISL_573587; EPI_ISL_535007; EPI_ISL_706132; EPI_ISL_679770; EPI_ISL_479095; EPI_ISL_559985; EPI_ISL_534827; EPI_ISL_560074; EPI_ISL_627256; EPI_ISL_638073; EPI_ISL_679725; EPI_ISL_559787; EPI_ISL_559756; EPI_ISL_573069; EPI_ISL_628575; EPI_ISL_638762; EPI_ISL_628516; EPI_ISL_652128; EPI_ISL_706043; EPI_ISL_706018; EPI_ISL_679656; EPI_ISL_559796; EPI_ISL_559797; EPI_ISL_573193; EPI_ISL_559989; EPI_ISL_573834 | Oxford Viromics, NDM, University of Oxford; Oxford University Hospitals; Basingstoke and North Hampshire Hospital | COVID-19 Genomics UK (COG-UK) Consortium | Tanya Golubchik et al A |
| EPI_ISL_448801 | Oxford Viromics, NDM, University of Oxford; Oxford University Hospitals; Basingstoke and North Hampshire Hospital | COVID-19 Genomics UK (COG-UK) Consortium | Tanya Golubchik et al B (https://virological.org/t/preliminary-analysis-of-sars-cov-2-importation-establishment-of-uk-transmission-lineages/507) |
| EPI_ISL_458372; EPI_ISL_467714; EPI_ISL_492793 | PHE South West Regional Laboratory, National Infection Service | Wellcome Sanger Institute for the COVID-19 Genomics UK (COG-UK) consortium | Stephanie Hutchings et al |
| EPI_ISL_678257 | PHI Institute of Nephrology - Struga | Research Center for Genetic Engineering and Biotechnology "Georgi D. Efremov" , Macedonian Academy of Sciences and Arts | RCGEB - MASA et al |
| EPI_ISL_733570 | POK OI HOSPITAL | Hong Kong Department of Health | Alan K.L. Tsang et al |
| EPI_ISL_596517; EPI_ISL_596527; EPI_ISL_596554; EPI_ISL_596553; EPI_ISL_596548; EPI_ISL_596552; EPI_ISL_596562; EPI_ISL_596549; EPI_ISL_596543; EPI_ISL_596545; EPI_ISL_596556; EPI_ISL_596565; EPI_ISL_596544; EPI_ISL_596501; EPI_ISL_596555; EPI_ISL_596522; EPI_ISL_596559; EPI_ISL_596557; EPI_ISL_596563; EPI_ISL_596520; EPI_ISL_596525; EPI_ISL_596531; EPI_ISL_596526; EPI_ISL_596528; EPI_ISL_596507; EPI_ISL_596514; EPI_ISL_596537; EPI_ISL_596538; EPI_ISL_596523; EPI_ISL_596535; EPI_ISL_596529; EPI_ISL_596503; EPI_ISL_596504; EPI_ISL_596502; EPI_ISL_596505; EPI_ISL_596506; EPI_ISL_596512 | Palestinian Ministry of Health | Molecular Genetics Lab | Nouar Qutob et al |
| EPI_ISL_707939; EPI_ISL_708193; EPI_ISL_708190; EPI_ISL_708195; EPI_ISL_708188; EPI_ISL_708185; EPI_ISL_707937; EPI_ISL_707938; EPI_ISL_708189; EPI_ISL_708194; EPI_ISL_708186; EPI_ISL_708192; EPI_ISL_708184; EPI_ISL_707936; EPI_ISL_708191 | Pamukkale University Hospital | Pamukkale University Department of Medical Genetics | Onur TOKGUN et al. et al |
| EPI_ISL_461482 | Pandit Deendayal Upadhyay Government Medical College, Rajkot | Gujarat Biotechnology Research Centre | Anjali Rajwar et al |
| EPI_ISL_430845 | Pasig City General Hospital | Research Institute for Tropical Medicine | Medado et al |
| EPI_ISL_654866 | Pasteur Institute in Ho Chi Minh city | Department of Microbiology and Immunology - Pasteur Institute in Ho Chi Minh city | Cao Minh Thắng et al |
| EPI_ISL_654889 | Pasteur Institute in Ho Chi Minh city | Department of Microbiology and Immunology - Pasteur Institute in Ho Chi Minh city | Hoàng Quốc Cường et al |
| EPI_ISL_654881; EPI_ISL_654882 | Pasteur Institute in Ho Chi Minh city | Department of Microbiology and Immunology - Pasteur Institute in Ho Chi Minh city | Lương Chấn Quang et al |
| EPI_ISL_654883 | Pasteur Institute in Ho Chi Minh city | Department of Microbiology and Immunology - Pasteur Institute in Ho Chi Minh city | Nguyễn Thanh Long et al |
| EPI_ISL_654887 | Pasteur Institute in Ho Chi Minh city | Department of Microbiology and Immunology - Pasteur Institute in Ho Chi Minh city | Phan Trọng Lân et al |
| EPI_ISL_442523 | Pasteur Institute of Iran | Kawsar Human Genetic Research Company | Sirous Zeinali et al |
| EPI_ISL_463749 | Pasteur Institute of Iran | Rapid Response Team | Mahboobeh Rafigh et al |
| EPI_ISL_456301 | PathLab Bay of Plenty | Institute of Environmental Science and Research (ESR) | Matt Storey et al B |
| EPI_ISL_582018; EPI_ISL_582020; EPI_ISL_582019; EPI_ISL_582021 | PathLab Bay of Plenty | Institute of Environmental Science and Research (ESR) | Xiaoyun Ren et al |
| EPI_ISL_420456; EPI_ISL_470860; EPI_ISL_470861 | PathWest Laboratory Medicine WA | PathWest Laboratory Medicine WA | Chisha Sikazwe et al |
| EPI_ISL_596691; EPI_ISL_596694; EPI_ISL_596766; EPI_ISL_596764; EPI_ISL_596699; EPI_ISL_512715; EPI_ISL_512716; EPI_ISL_512737; EPI_ISL_672637; EPI_ISL_596770; EPI_ISL_596842 | PathWest Laboratory Medicine WA | PathWest Laboratory Medicine WA Microbial Surveillance Unit | PathWest Laboratory Medicine WA Microbial Surveillance Unit et al |
| EPI_ISL_582022; EPI_ISL_582023 | Pathlab Lakes | Institute of Environmental Science and Research (ESR) | Xiaoyun Ren et al |
| EPI_ISL_513020 | Pathogen Genomics Lab King Abdullah University of Science and Technology(KAUST) | Pathogen Genomics Lab King Abdullah University of Science and Technology(KAUST) | Afrah Alsomali et al |
| EPI_ISL_678097; EPI_ISL_678101 | Pathogen Genomics Lab King Abdullah University of Science and Technology(KAUST) | Pathogen Genomics Lab King Abdullah University of Science and Technology(KAUST) | Amanda Ooi et al |
| EPI_ISL_513178; EPI_ISL_512994; EPI_ISL_513177 | Pathogen Genomics Lab King Abdullah University of Science and Technology(KAUST) | Pathogen Genomics Lab King Abdullah University of Science and Technology(KAUST) | Amit Kumar Subudhi et al |
| EPI_ISL_512923; EPI_ISL_512922; EPI_ISL_512920; EPI_ISL_512924; EPI_ISL_512921; EPI_ISL_512926; EPI_ISL_512927; EPI_ISL_512925 | Pathogen Genomics Lab King Abdullah University of Science and Technology(KAUST) | Pathogen Genomics Lab King Abdullah University of Science and Technology(KAUST) | Fadwa Alofi et al |
| EPI_ISL_678066; EPI_ISL_513081; EPI_ISL_636966 | Pathogen Genomics Lab King Abdullah University of Science and Technology(KAUST) | Pathogen Genomics Lab King Abdullah University of Science and Technology(KAUST) | Fathia Ben Rached et al |
| EPI_ISL_678125 | Pathogen Genomics Lab King Abdullah University of Science and Technology(KAUST) | Pathogen Genomics Lab King Abdullah University of Science and Technology(KAUST) | Luke Esau et al |
| EPI_ISL_678090; EPI_ISL_678239 | Pathogen Genomics Lab King Abdullah University of Science and Technology(KAUST) | Pathogen Genomics Lab King Abdullah University of Science and Technology(KAUST) | Muhammad Shuaib et al |
| EPI_ISL_678196; EPI_ISL_512898; EPI_ISL_512890; EPI_ISL_513211; EPI_ISL_512875; EPI_ISL_513210 | Pathogen Genomics Lab King Abdullah University of Science and Technology(KAUST) | Pathogen Genomics Lab King Abdullah University of Science and Technology(KAUST) | Raeece Naeem et al |
| EPI_ISL_513203; EPI_ISL_513035; EPI_ISL_513205; EPI_ISL_636963 | Pathogen Genomics Lab King Abdullah University of Science and Technology(KAUST) | Pathogen Genomics Lab King Abdullah University of Science and Technology(KAUST) | Rahul P Salunke et al |
| EPI_ISL_678205 | Pathogen Genomics Lab King Abdullah University of Science and Technology(KAUST) | Pathogen Genomics Lab King Abdullah University of Science and Technology(KAUST) | Raushan Nugmanova et al |
| EPI_ISL_512948; EPI_ISL_678035; EPI_ISL_437484; EPI_ISL_678224; EPI_ISL_678185; EPI_ISL_677952; EPI_ISL_751230; EPI_ISL_678218; EPI_ISL_678223; EPI_ISL_678240; EPI_ISL_751231; EPI_ISL_437497; EPI_ISL_512980; EPI_ISL_678220; EPI_ISL_513171; EPI_ISL_678221; EPI_ISL_678164; EPI_ISL_678222; EPI_ISL_678005 | Pathogen Genomics Lab King Abdullah University of Science and Technology(KAUST) | Pathogen Genomics Lab King Abdullah University of Science and Technology(KAUST) | Sara Mfarrej et al |
| EPI_ISL_437460; EPI_ISL_437475; EPI_ISL_437459; EPI_ISL_678080; EPI_ISL_678141; EPI_ISL_437757; EPI_ISL_437701; EPI_ISL_678072; EPI_ISL_437753 | Pathogen Genomics Lab King Abdullah University of Science and Technology(KAUST) | Pathogen Genomics Lab King Abdullah University of Science and Technology(KAUST) | Sharif Hala et al |
| EPI_ISL_733175; EPI_ISL_733169; EPI_ISL_566079; EPI_ISL_566080; EPI_ISL_596351; EPI_ISL_596336; EPI_ISL_566083; EPI_ISL_596339; EPI_ISL_733165; EPI_ISL_733196; EPI_ISL_733166; EPI_ISL_733189; EPI_ISL_733209 | Pathogenic Microorganisms Variability Laboratory | WHO National Influenza Centre Russian Federation | Andrey Komissarov et al A |
| EPI_ISL_583625 | Pathologie-Labor Dr. Obrist- Dr. Brunhuber | Bergthaler laboratory, CeMM Research Center for Molecular Medicine of the Austrian Academy of Sciences | Alexandra Popa et al A |
| EPI_ISL_407894; EPI_ISL_410718 | Pathology Queensland | Public Health Virology Laboratory | Ben Huang et al A |
| EPI_ISL_410717 | Pathology Queensland | Public Health Virology Laboratory | Ben Huang et al B (https://dx.doi.org/10.2807/1560-7917.ES.2020.25.13.2000305) |
| EPI_ISL_414414 | Pathology Queensland | Public Health Virology Laboratory | Bixing Huang et al |
| EPI_ISL_539850; EPI_ISL_539851 | Pok Oi Hospital | Hong Kong Department of Health | Alan K.L. Tsang et al |
| EPI_ISL_745187 | Port Nolloth Hospital | National Health Laboratory Service (NHLS), Tygerberg | Susan Engelbrecht et al |
| EPI_ISL_529013 | Presidio Ospedaliero "S.Filippo e Nicola"-Avezzano | Istituto Zooprofilattico Sperimentale dell'Abruzzo e Molise "G.Caporale" | Lorusso A et al |
| EPI_ISL_429226 | Presidio Ospedaliero Santo Spirito | Istituto Zooprofilattico Sperimentale dell'Abruzzo e Molise "G. Caporale" | Lorusso A et al |
| EPI_ISL_759955 | Primasatya Husada Citra Hospital | Institute of Tropical Disease, Universitas Airlangga | Aldise M Nastri et al |
| EPI_ISL_417064 | Prince of Wales Hospital | Hong Kong Department of Health | Alan K.L. Tsang et al |
| EPI_ISL_513534; EPI_ISL_513550; EPI_ISL_513536; EPI_ISL_513559 | Programa de Oncovirologia, Instituto Nacional de Câncer | Programa de Oncovirologia, Instituto Nacional de Câncer | Juliana D. Siqueira et al |
| EPI_ISL_591519; EPI_ISL_591521 | Programa de Virología, Facultad de Medicina UC | Center for Mathematical Modeling and Center for Genome Regulation. Santiago, Chile | Gaggero A et al |
| EPI_ISL_574492; EPI_ISL_574491; EPI_ISL_574480 | Programme in Emerging Infectious Diseases, Duke-NUS Medical School | National Public Health Laboratory, National Centre for Infectious Diseases | Tze Minn Mak et al |
| EPI_ISL_523959 | Pronto Socorro Municipal de Perus | Instituto Adolfo Lutz, Interdiciplinary Procedures Center, Strategic Laboratory | Claudio Tavares Sacchi et al B |
| EPI_ISL_693231 | Pronto Socorro Municipal de Santa Branca | Instituto Adolfo Lutz, Interdiciplinary Procedures Center, Strategic Laboratory | Claudio Tavares Sacchi et al B |
| EPI_ISL_582134 | Protzer Lab | Protzer Lab | Ulrike Protzer et al |
| EPI_ISL_404895 | Providence Regional Medical Center | Division of Viral Diseases, Centers for Disease Control and Prevention | Queen et al (https://dx.doi.org/10.1101/2020.03.02.972935) |
| EPI_ISL_572329 | Public Health Authority of the Slovak Republic, Bratislava | Faculty of Natural Sciences, Comenius University in Bratislava | Dominika Fričová et al |
| EPI_ISL_636973 | Public Health Lab | Public Health Lab | Alwasti et al |
| EPI_ISL_425177 | Public Health Ontario | Public Health Agency of Canada - National Microbiology Laboratory | Amrit S. Boese et al |
| EPI_ISL_413015 | Public Health Ontario Laboratory | National Microbiology Laboratory | Shari Tyson et al |

| | | | |
|---|---|---|---|
| EPI_ISL_513313; EPI_ISL_513312 | Public Health, United States Air Force School of Aerospace Medicine | Public Health, United States Air Force School of Aerospace Medicine | Fries et al |
| EPI_ISL_763066 | Puskesmas Grogol, Sukoharjo | Universitas Sebelas Maret (UNS); Rumah Sakit UNS (RS-UNS); National Institute of Health Research and Development, Indonesian Ministry of Health. | Yulia Sari et al |
| EPI_ISL_632936 | Puskesmas Mlati 1 Sleman | Genetics Working Group (Pokja Genetik) Faculty of Medicine, Public Health and Nursing Universitas Gadjah Mada (FK-KMK UGM); Disease Investigation Center Wates Ministry of Agriculture Indonesia; Department of Microbiology FK-KMK UGM; Laboratorium Diagnostik Yayasan Tahija World Mosquito Program (WMP) Yogyakarta Center for Tropical Medicine FK-KMK UGM; Integrated Research Center FK-KMK UGM; Department of Computer Science and Electronics FMIPA UGM; RSUP Dr. Sardjito | Gunadi et al |
| EPI_ISL_574610 | Puskesmas Tambora | Eijkman Institute for Molecular Biology, Ministry of Research and Technology/National Agency for Research and Innovation | Frilasita A Yudhaputri et al |
| EPI_ISL_518819 | Qadr Hospital, Tangerang, Banten | Biosafety Level-3 Laboratory, Indonesian Institute of Sciences (LIPI) | Anik Budhi Dharmayanthi et al |
| EPI_ISL_457362; EPI_ISL_457365; EPI_ISL_457333 | Quadram Institute Bioscience | COVID-19 Genomics UK (COG-UK) Consortium | Dave J. Baker et al A (https://virological.org/t/preliminary-analysis-of-sars-cov-2-importation-establishment-of-uk-transmission-lineages/507) |
| EPI_ISL_596108; EPI_ISL_664248; EPI_ISL_627080 | Quadram Institute Bioscience | COVID-19 Genomics UK (COG-UK) Consortium | Dave J. Baker et al B |
| EPI_ISL_487435; EPI_ISL_487436; EPI_ISL_487433 | Queen Astrid Military Hospital | Institute of Tropical Medicine | Philippe Selhorst et al |
| EPI_ISL_759896 | Queen Elizabeth Hospital | Hong Kong Department of Health | Alan K.L. Tsang et al |
| EPI_ISL_733571; EPI_ISL_539816; EPI_ISL_539817 | Queen Mary Hospital | Hong Kong Department of Health | Alan K.L. Tsang et al |
| EPI_ISL_499809 | Queens Medical Centre, Clinical Microbiology Department / DeepSeq Nottingham | COVID-19 Genomics UK (COG-UK) Consortium | Gemma Clark et al |
| EPI_ISL_530232; EPI_ISL_530238 | Queensland Health Forensic and Scientific Services, Public Health Virology | Public Health Virology Laboratory, Forensic and Scientific Services, Queensland Health | Son Nguyen et al et al |
| EPI_ISL_569623 | Quick Care Watertown | South Dakota Public Health Laboratory | Matt Plumb et al |
| EPI_ISL_416482 | R. G. Lugar Center for Public Health Research, National Center for Disease Control and Public Health (NCDC) of Georgia. | R. G. Lugar Center for Public Health Research, National Center for Disease Control and Public Health (NCDC) of Georgia. | Adam Kotorashvili et al |
| EPI_ISL_416480 | R. G. Lugar Center for Public Health Research, National Center for Disease Control and Public Health (NCDC) of Georgia. | R. G. Lugar Center for Public Health Research, National Center for Disease Control and Public Health (NCDC) of Georgia. | Ann Machablishvili et al |
| EPI_ISL_416481 | R. G. Lugar Center for Public Health Research, National Center for Disease Control and Public Health (NCDC) of Georgia. | R. G. Lugar Center for Public Health Research, National Center for Disease Control and Public Health (NCDC) of Georgia. | Gvantsa Chanturia et al |
| EPI_ISL_416477; EPI_ISL_416479; EPI_ISL_416478 | R. G. Lugar Center for Public Health Research, National Center for Disease Control and Public Health (NCDC) of Georgia. | R. G. Lugar Center for Public Health Research, National Center for Disease Control and Public Health (NCDC) of Georgia. | Marine Murtskhvaladze et al |
| EPI_ISL_415644; EPI_ISL_415643; EPI_ISL_415641; EPI_ISL_415642 | R. G. Lugar Center for Public Health Research, National Center for Disease Control and Public Health (NCDC) of Georgia. | R. G. Lugar Center for Public Health Research, National Center for Disease Control and Public Health (NCDC) of Georgia. | Nato Kotaria et al |
| EPI_ISL_413581; EPI_ISL_413576; EPI_ISL_413569; EPI_ISL_413570; EPI_ISL_413575 | RIVM | Erasmus Medical Center | David Nieuwenhuijse et al A (https://dx.doi.org/10.1038/s41591-020-0997-y) |
| EPI_ISL_568693 | RS Freeport Tembagapura | Eijkman Institute for Molecular Biology, Ministry of Research and Technology/National Agency for Research and Innovation | Frilasita A Yudhaputri et al |
| EPI_ISL_574613 | RS Harapan Bunda | Eijkman Institute for Molecular Biology, Ministry of Research and Technology/National Agency for Research and Innovation | Frilasita A Yudhaputri et al |
| EPI_ISL_574612 | RS Hermina Mekarsari | Eijkman Institute for Molecular Biology, Ministry of Research and Technology/National Agency for Research and Innovation | Frilasita A Yudhaputri et al |
| EPI_ISL_763064 | RS Kasih Ibu, Surakarta | Universitas Sebelas Maret (UNS); Rumah Sakit UNS (RS-UNS); National Institute of Health Research and Development, Indonesian Ministry of Health. | Betty Suryawati et al |
| EPI_ISL_574618 | RS Kramat 128 | Eijkman Institute for Molecular Biology, Ministry of Research and Technology/National Agency for Research and Innovation | Hidayat Trimarsanto et al |
| EPI_ISL_763063 | RS Panti Waluyo, Surakarta | Universitas Sebelas Maret (UNS); Rumah Sakit UNS (RS-UNS); National Institute of Health Research and Development, Indonesian Ministry of Health. | Betty Suryawati et al |
| EPI_ISL_568688 | RS Pelni | Eijkman Institute for Molecular Biology, Ministry of Research and Technology/National Agency for Research and Innovation | Frilasita A Yudhaputri et al |
| EPI_ISL_435282; EPI_ISL_437190; EPI_ISL_437191 | RS Pondok Indah Hospital – Pondok Indah | Eijkman Institute for Molecular Biology, Ministry of Research and Technology/National Agency for Research and Innovation | Edison Johar et al |
| EPI_ISL_568689 | RS Pondok Indah Puri Indah | Eijkman Institute for Molecular Biology, Ministry of Research and Technology/National Agency for Research and Innovation | Frilasita A Yudhaputri et al |
| EPI_ISL_574611 | RS Premier Bintaro | Eijkman Institute for Molecular Biology, Ministry of Research and Technology/National Agency for Research and Innovation | Frilasita A Yudhaputri et al |
| EPI_ISL_759702 | RS UNS Surakarta | Universitas Sebelas Maret (UNS), Rumah sakit Universitas Sebelas Maret (RS-UNS) Surakarta, National Institute of Health Research and Development, Indonesian Ministry of Health | Betty Suryawati et al |
| EPI_ISL_759701 | RS UNS, Surakarta | UNS/RS-UNS, Faculty of Medicine | Betty Suryawati et al |
| EPI_ISL_529138 | RSAL Dr. Ramelan Surabaya | Institute of Tropical Disease, Universitas Airlangga | Jezzy R Dewantari et al |
| EPI_ISL_454497; EPI_ISL_454510 | RSE "National Center for Biotechnology" | RSE "National Center for Biotechnology" | Alexandr Shevtsov et al |
| EPI_ISL_759968 | RSI Darus Syifa' | Institute of Tropical Disease, Universitas Airlangga | Maria I Lusida et al |
| EPI_ISL_574609 | RSJPD Harapan Kita | Eijkman Institute for Molecular Biology, Ministry of Research and Technology/National Agency for Research and Innovation | Frilasita A Yudhaputri et al |
| EPI_ISL_576383 | RSUD Budi Rahayu Kota Magelang | Genetics Working Group (Pokja Genetik) Faculty of Medicine, Public Health and Nursing Universitas Gadjah Mada (FK-KMK UGM); Disease Investigation Center Wates Ministry of Agriculture Indonesia; Department of Microbiology FK-KMK UGM; Laboratorium Diagnostik Yayasan Tahija World Mosquito Program (WMP) Yogyakarta Center for Tropical Medicine FK-KMK UGM; Integrated Research Center FK-KMK UGM; Department of Computer Science and Electronics FMIPA UGM; Balai Besar Teknik Kesehatan Lingkungan dan Pengendalian Penyakit (BBTKLPP) Yogyakarta | Gunadi et al |
| EPI_ISL_759703 | RSUD Bung Karno | Universitas Sebelas Maret (UNS), Rumah sakit Universitas Sebelas Maret (RS-UNS) Surakarta, National Institute of Health Research and Development, Indonesian Ministry of Health | Betty Suryawati et al |
| EPI_ISL_759705 | RSUD Bung Karno, Surakarta | Universitas Sebelas Maret (UNS); Rumah Sakit Universitas Sebelas Maret (RS-UNS), Surakarta; National Institute of Health Research and Development, Indonesian Ministry of Health , Jakarta). | Betty Suryawati et al |
| EPI_ISL_610161 | RSUD Dr. Tjitrowardojo | Genetics Working Group (Pokja Genetik) Faculty of Medicine, Public Health and Nursing Universitas Gadjah Mada (FK-KMK UGM); Disease Investigation Center Wates Ministry of Agriculture Indonesia; Department of Microbiology FK-KMK UGM; Laboratorium Diagnostik Yayasan Tahija World Mosquito Program (WMP) Yogyakarta Center for Tropical Medicine FK-KMK UGM; Integrated Research Center FK-KMK UGM; Department of Computer Science and Electronics FMIPA UGM; Balai Besar Teknik Kesehatan Lingkungan dan Pengendalian Penyakit (BBTKLPP) Yogyakarta | Gunadi et al |
| EPI_ISL_516829 | RSUD Nyi Ageng Serang | Genetics Working Group (Pokja Genetik) Faculty of Medicine, Public Health and Nursing Universitas Gadjah Mada (FK-KMK UGM), Disease Investigation Center Wates Ministry of Agriculture Indonesia, Department of Microbiology FK-KMK UGM, Laboratorium Diagnostik Yayasan Tahija World Mosquito Program (WMP) Yogyakarta Center for Tropical Medicine FK-KMK UGM, Integrated Research Center FK-KMK UGM, Department of Computer Science and Electronics FMIPA UGM | Gunadi et al |
| EPI_ISL_568686 | RSUD Prof. DR. Margono Soekarjo | Eijkman Institute for Molecular Biology, Ministry of Research and Technology/National Agency for Research and Innovation | Frilasita A Yudhaputri et al |
| EPI_ISL_632937 | RSUD Saptosari Gunung Kidul | Genetics Working Group (Pokja Genetik) Faculty of Medicine, Public Health and Nursing Universitas Gadjah Mada (FK-KMK UGM); Disease Investigation Center Wates Ministry of Agriculture Indonesia; Department of Microbiology FK-KMK UGM; Laboratorium Diagnostik Yayasan Tahija World Mosquito Program (WMP) Yogyakarta Center for Tropical Medicine FK-KMK UGM; Integrated Research Center FK-KMK UGM; Department of Computer Science and Electronics FMIPA UGM; Balai Besar Teknik Kesehatan Lingkungan dan Pengendalian Penyakit (BBTKLPP) Yogyakarta | Gunadi et al |
| EPI_ISL_529967; EPI_ISL_759963 | RSUD Sidoarjo | Institute of Tropical Disease, Universitas Airlangga | Aldise M Nastri et al |
| EPI_ISL_529966 | RSUD Sidoarjo | Institute of Tropical Disease, Universitas Airlangga | Kazufumi Shimizu et al |
| EPI_ISL_759966 | RSUD Sidoarjo | Institute of Tropical Disease, Universitas Airlangga | Krisnoadi Rahardjo et al |
| EPI_ISL_753699 | RSUD Ulin Banjarmasin Kalimantan Selatan | National Institute of Health Research and Development | Subangkit et al |
| EPI_ISL_538510 | RSUD Ulin Banjarmasin South Kalimantan | National Institute of Health Research and Development | Pawestri et al |
| EPI_ISL_576128; EPI_ISL_576130; EPI_ISL_576114 | RSUP Dr. Sardjito | Genetics Working Group (Pokja Genetik) Faculty of Medicine, Public Health and Nursing Universitas Gadjah Mada (FK-KMK UGM); Disease Investigation Center Wates Ministry of Agriculture Indonesia; Department of Microbiology FK-KMK UGM; Laboratorium Diagnostik Yayasan Tahija World Mosquito Program (WMP) Yogyakarta Center for Tropical Medicine FK-KMK UGM; Integrated Research Center FK-KMK UGM; Department of Computer Science and Electronics FMIPA UGM | Gunadi et al |
| EPI_ISL_467376 | RSUP Fatmawati | Eijkman Institute for Molecular Biology, Ministry of Research and Technology/National Agency for Research and Innovation | Edison Johar et al |
| EPI_ISL_568691; EPI_ISL_568687; EPI_ISL_568690 | RSUP Fatmawati | Eijkman Institute for Molecular Biology, Ministry of Research and Technology/National Agency for Research and Innovation | Frilasita A Yudhaputri et al |
| EPI_ISL_637113; EPI_ISL_637112; EPI_ISL_637110 | Rafik Hariri University Hospital | Microbial Pathogenomics Lab | Georgi Merhi et al |
| EPI_ISL_450512; EPI_ISL_450515; EPI_ISL_450508; EPI_ISL_450511; EPI_ISL_450514; EPI_ISL_450516; EPI_ISL_454420; EPI_ISL_450509 | Rafik Hariri University Hospital | Rafik Hariri University Hospital | Rita Feghali et al |

| | | | |
|---|---|---|---|
| EPI_ISL_447026; EPI_ISL_447020; EPI_ISL_447019; EPI_ISL_447025; EPI_ISL_455922; EPI_ISL_447028; EPI_ISL_447009; EPI_ISL_498264; EPI_ISL_568580; EPI_ISL_447022; EPI_ISL_447018; EPI_ISL_447014; EPI_ISL_447027 | Ramathibodi Hospital | COVID-19 Network Investigations (CONI) Alliance | Elizabeth Batty et al |
| EPI_ISL_735490 | Rangamati General Hospital RT-PCR lab, | Central Biological Research Laboratory and Department of Biochemistry and Molecular Biology | Robiul Hasan Bhuiyan et al |
| EPI_ISL_745199 | Rangaraya Medical College | CSIR-Centre for Cellular and Molecular Biology | Dr.V.Sudha Rani et al |
| EPI_ISL_708735 | Regional Medical Sciences Center 11/1 phuket | National Institute of Health, Department of Medical Sciences, Ministry of Public Health, Thailand | Pilailuk Okada et al |
| EPI_ISL_585174 | Regional Virus Laboratory, Belfast Health and Social Care Trust | COVID-19 Genomics UK (COG-UK) Consortium | Conall McCaughey et al |
| EPI_ISL_482042; EPI_ISL_501049; EPI_ISL_501025; EPI_ISL_470022 | Regional Virus Laboratory, Belfast Health and Social Care Trust | Wellcome Sanger Institute for the COVID-19 Genomics UK (COG-UK) consortium | Conall McCaughey et al |
| EPI_ISL_708811; EPI_ISL_708810 | Regional medical sciences center 2 Phitsanulok | National Institute of Health, Department of Medical Sciences, Ministry of Public Health, Thailand | Pilailuk Okada et al |
| EPI_ISL_708808; EPI_ISL_708806; EPI_ISL_708800; EPI_ISL_708802; EPI_ISL_708801; EPI_ISL_708797; EPI_ISL_708737; EPI_ISL_708804; EPI_ISL_708807 | Regional medical sciences center 6 chonburi | National Institute of Health, Department of Medical Sciences, Ministry of Public Health, Thailand | Pilailuk Okada et al |
| EPI_ISL_491474 | Research Institute for Tropical Medicine | Research Institute for Tropical Medicine | Ma. Angelica Tujan et al |
| EPI_ISL_451934 | Research Unit, University Hospital for Infectious Diseases "Dr. Fran Mihaljević" | Cicin Sain lab, Helmholtz Centre for Infection Research | Zeeshan Chaudhry et al |
| EPI_ISL_407071; EPI_ISL_407073; EPI_ISL_414042; EPI_ISL_414041; EPI_ISL_414044; EPI_ISL_414522; EPI_ISL_414009; EPI_ISL_417215; EPI_ISL_414011; EPI_ISL_424103; EPI_ISL_423715; EPI_ISL_423340 | Respiratory Virus Unit, Microbiology Services Colindale, Public Health England | Respiratory Virus Unit, Microbiology Services Colindale, Public Health England | Monica Galiano et al (https://virological.org/t/preliminary-analysis-of-sars-cov-2-importation-establishment-of-uk-transmission-lineages/507) |
| EPI_ISL_464199; EPI_ISL_464210; EPI_ISL_464228; EPI_ISL_464223; EPI_ISL_464247; EPI_ISL_464189; EPI_ISL_464212; EPI_ISL_464242; EPI_ISL_464168; EPI_ISL_464169; EPI_ISL_464178; EPI_ISL_464180; EPI_ISL_464412; EPI_ISL_465709; EPI_ISL_465621; EPI_ISL_466512; EPI_ISL_490249 | Respiratory Virus Unit, Microbiology Services Colindale, Public Health England | Respiratory Virus Unit, Microbiology Services Colindale, Public Health England | PHE Covid Sequencing Team et al |
| EPI_ISL_682298 | Respiratory virus Laboratory, Chinese Academy of Medical Science | Respiratory virus Laboratory, Chinese Academy of Medical Science | Li et al |
| EPI_ISL_430456 | Rizal Medical Center | Research Institute for Tropical Medicine | Medado et al |
| EPI_ISL_481256; EPI_ISL_481262; EPI_ISL_481264; EPI_ISL_481253 | Robert Koch Institute, National Reference center for Influenza, Berlin, Germany | Robert Koch Institute, Bioinformatics MF1, Berlin, Germany | Marianne Wedde et al |
| EPI_ISL_751799 | Robert Koch Institute, National Reference center for Influenza, Berlin, Germany | Robert Koch Institute, Bioinformatics MF1, Berlin, Germany | Stephan Fuchs et al |
| EPI_ISL_483142; EPI_ISL_483149; EPI_ISL_483153; EPI_ISL_483156; EPI_ISL_483154; EPI_ISL_483155; EPI_ISL_483150; EPI_ISL_483143; EPI_ISL_483144; EPI_ISL_483157; EPI_ISL_483152; EPI_ISL_483139; EPI_ISL_483140; EPI_ISL_483145; EPI_ISL_483148; EPI_ISL_483141; EPI_ISL_483151; EPI_ISL_483146; EPI_ISL_483147 | Robert Koch Institute, ZBS1 Highly Pathogenic Viruses, Berlin, Germany | Robert Koch Institute, Bioinformatics MF1, Berlin, Germany | Janine Michel et al |
| EPI_ISL_514134; EPI_ISL_514136 | Rondônia Central Public Health Laboratory (LACEN/RO), vinculated to State Health Secretariat of Rondônia (SESAU/RO) | Molecular Virology Laboratory of Oswaldo Cruz Foundation of Rondônia | Luan Felipo Botelho-Souza et al |
| EPI_ISL_516800 | Rumah Sakit Akademik Universitas Gadjah Mada | Genetics Working Group (Pokja Genetik) Faculty of Medicine, Public Health and Nursing Universitas Gadjah Mada (FK-KMK UGM); Disease Investigation Center Wates Ministry of Agriculture Indonesia; Department of Microbiology FK-KMK UGM; Laboratorium Diagnostik Yayasan Tahija World Mosquito Program (WMP) Yogyakarta Center for Tropical Medicine FK-KMK UGM; Integrated Research center FK-KMK UGM; Department of Computer Science and Electronics FMIPA UGM | Gunadi et al |
| EPI_ISL_516806 | Rumah Sakit PKU Gamping | Genetics Working Group (Pokja Genetik) Faculty of Medicine, Public Health and Nursing Universitas Gadjah Mada (FK-KMK UGM); Disease Investigation Center Wates Ministry of Agriculture Indonesia; Department of Microbiology FK-KMK UGM; Laboratorium Diagnostik Yayasan Tahija World Mosquito Program (WMP) Yogyakarta Center for Tropical Medicine FK-KMK UGM; Integrated Research center FK-KMK UGM; Department of Computer Science and Electronics FMIPA UGM | Gunadi et al |
| EPI_ISL_759842 | Rumah Sakit Universitas Sebelas Maret (RS-UNS) | Universitas Sebelas Maret (UNS); Rumah Sakit Universitas Sebelas Maret (RS-UNS), Surakarta; National Institute of Health Research and Development, Indonesian Ministry of Health, Jakarta. | Betty Suryawati et al |
| EPI_ISL_491298 | Rural Health Unit - Calauan, Laguna | Research Institute for Tropical Medicine | Tujan et al |
| EPI_ISL_470896 | Russian State Collection of Viruses | Pathogenic Microorganisms Variability Laboratory | Alexey Shchetinin et al |
| EPI_ISL_735445 | Rwanda National Reference Laboratory | GIGA Medical Genomics | Yvan Butera et al |
| EPI_ISL_707789 | Rwanda National Reference Laboratory | Rwanda National Reference Laboratory | Enatha Mukantwari et al |
| EPI_ISL_468037; EPI_ISL_468032; EPI_ISL_603124; EPI_ISL_654795; EPI_ISL_654805; EPI_ISL_654808; EPI_ISL_654804; EPI_ISL_654806; EPI_ISL_654799; EPI_ISL_755571; EPI_ISL_603123; EPI_ISL_732961 | SA Pathology | SA Pathology | Lex Leong et al |
| EPI_ISL_455603 | SA Pathology | VPRL | Beard et al |
| EPI_ISL_738044 | SIESP CHIETI - DRIVE IN LANCIANO | Istituto Zooprofilattico Sperimentale dell'Abruzzo e Molise "G. Caporale" | Lorusso A et al |
| EPI_ISL_648217 | SILAB | Laboratorio Mixto de Biotecnología Acuática (LMBA) | Joaquín Ezpeleta et al |
| EPI_ISL_512073 | Saikrishna Hospital, Mehsana | Gujarat Biotechnology Research Centre | Apurvasinh Puvar et al |
| EPI_ISL_512072 | Saikrishna Hospital, Mehsana | Gujarat Biotechnology Research Centre | Harshadbhai Parmar et al |
| EPI_ISL_636144; EPI_ISL_649028; EPI_ISL_494594; EPI_ISL_494621; EPI_ISL_494451; EPI_ISL_636080 | San Diego County Public Health Laboratory | Andersen lab at Scripps Research | SEARCH Alliance San Diego with Tracy Basler et al |
| EPI_ISL_542285; EPI_ISL_542398 | San Matteo Hospital Pavia | Dep. Of Oncology and Hemato-Oncology University of Milan | Claudia Alteri et al |
| EPI_ISL_750178; EPI_ISL_750161; EPI_ISL_750170; EPI_ISL_749151; EPI_ISL_749155; EPI_ISL_749238; EPI_ISL_750169; EPI_ISL_750168; EPI_ISL_749906; EPI_ISL_751011; EPI_ISL_750820; EPI_ISL_750172; EPI_ISL_750171; EPI_ISL_750176; EPI_ISL_750177 | Sanatorio Americano | Institut Pasteur de Montevideo | Daiana Mir et al |
| EPI_ISL_435580; EPI_ISL_435583 | Santa Clara County Public Health Department | Chiu Laboratory, University of California, San Francisco | Xianding Deng et al |
| EPI_ISL_529718 | Santo Borromeus Hospital | Laboratorium Kesehatan Provinsi Jawa Barat; School of Life Sciences and Technology & School of Pharmacy-Institut Teknologi Bandung; Molecular Genetics Laboratory-Faculty of Medicine-Universitas Padjadjaran | Ema Rahmawati et al |
| EPI_ISL_528751 | Santo Borromeus Hospital | Molecular Genetics Laboratory-Faculty of Medicine-Universitas Padjadjaran; School of Life Sciences and Technology & School of Pharmacy-Institut Teknologi Bandung; Laboratorium Kesehatan Provinsi Jawa Barat | Yunia Sribudiani et al |
| EPI_ISL_528749 | Santosa Hospital Bandung Central | School of Life Sciences and Technology & School of Pharmacy-Institut Teknologi Bandung; Molecular Genetics Laboratory-Faculty of Medicine-Universitas Padjadjaran; Laboratorium Kesehatan Provinsi Jawa Barat | Husna Nugrahapraja et al |
| EPI_ISL_754239 | Santosa Hospital Bandung Central | School of Life Sciences and Technology & School of Pharmacy-Institut Teknologi Bandung; Molecular Genetics Laboratory-Faculty of Medicine-Universitas Padjadjaran; Laboratorium Kesehatan Provinsi Jawa Barat | Marselina Irasonia Tan et al |
| EPI_ISL_511878 | Santosa Hospital Bandung Kopo | Laboratorium Kesehatan Provinsi Jawa Barat; Molecular Genetics Laboratory-Faculty of Medicine-Universitas Padjadjaran; School of Life Sciences and Technology & School of Pharmacy-Institut Teknologi Bandung | Ema Rahmawati et al |
| EPI_ISL_479833; EPI_ISL_479835; EPI_ISL_479800; EPI_ISL_479799 | Sapporo City Institute of Public Health | Pathogen Genomics Center, National Institute of Infectious Diseases | Tsuyoshi Sekizuka et al C (https://www.medrxiv.org/content/10.1101/2020.07.01.20143958v1) |
| EPI_ISL_510529 | School of Veterinary Medicine, Disease Control | School of Veterinary Medicine, Disease Control | Simulundu et al |
| EPI_ISL_676576; EPI_ISL_676596; EPI_ISL_676580; EPI_ISL_676592; EPI_ISL_676590; EPI_ISL_661179; EPI_ISL_661195; EPI_ISL_676582; EPI_ISL_661184; EPI_ISL_676584; EPI_ISL_661189; EPI_ISL_676591; EPI_ISL_676602; EPI_ISL_676603; EPI_ISL_661191; EPI_ISL_676601; EPI_ISL_661181; EPI_ISL_661196; EPI_ISL_676578; EPI_ISL_661185; EPI_ISL_676574; EPI_ISL_661180; EPI_ISL_661183; EPI_ISL_676577; EPI_ISL_676586; EPI_ISL_676587; EPI_ISL_676588; EPI_ISL_676579; EPI_ISL_661197; EPI_ISL_676583; EPI_ISL_661188; EPI_ISL_676585; EPI_ISL_676589; EPI_ISL_661192; EPI_ISL_676593 | Scientific Veterinary Institute Novi Sad | Veterinary Specialized Institute "Kraljevo", Serbia | Vidanovic et al |
| EPI_ISL_693238 | Secao Centro de Diagnostico Secedi | Instituto Adolfo Lutz, Interdiciplinary Procedures Center, Strategic Laboratory | Claudio Tavares Sacchi et al B |
| EPI_ISL_708530 | Secretaria Municipal de Saude de Fernandópolis | Instituto Adolfo Lutz, Interdiciplinary Procedures Center, Strategic Laboratory | Claudio Tavares Sacchi et al B |
| EPI_ISL_417924 | Secretaría de Salud Medellín | Instituto Nacional de Salud, Universidad Cooperativa de Colombia, Instituto Alexander von Humboldt, Imperial College-London, London School of Hygiene & Tropical Medicine | Marcela Mercado-Reyes et al (https://dx.doi.org/10.1101/2020.06.02.20120782) |
| EPI_ISL_708529 | Secretária Municipal de Saude de Fernandópolis | Instituto Adolfo Lutz, Interdiciplinary Procedures Center, Strategic Laboratory | Claudio Tavares Sacchi et al B |
| EPI_ISL_593934 | Sentinelles, Chanteloup-En-Brie | National Reference Center for Viruses of Respiratory Infections, Institut Pasteur, Paris | Sylvie Behillil et al |
| EPI_ISL_421512 | Service de Biologie Médicale - BP 125 | National Reference Center for Viruses of Respiratory Infections, Institut Pasteur, Paris | Mélanie Albert et al C |
| EPI_ISL_420599; EPI_ISL_420600; EPI_ISL_420598 | Servicio Virosis Respiratorias-Departamento Virología-INEI | Instituto Nacional Enfermedades Infecciosas C.G.Malbran | Baumeister E. et al (https://dx.doi.org/10.1101/2020.05.23.20111443) |
| EPI_ISL_416485 | Servicio de Microbiologia. Consorcio Hospital General Universitario de Valencia | Sequencing and Bioinformatics Service and Molecular Epidemiology Research Group. FISABIO-Public Health | Griselda De Marco et al A (https://dx.doi.org/10.1101/2020.04.20.050039) |
| EPI_ISL_538163 | Servicio de Microbiología y Parasitología clínica. UCEIMP. Hospital Universitario Virgen del Rocío/IBIS/CSIC/US | SeqCOVID-SPAIN consortium/IBV(CSIC) | Guillermo Martín Gutiérrez et al |
| EPI_ISL_468790; EPI_ISL_500355; EPI_ISL_510366; EPI_ISL_510349; EPI_ISL_510343; EPI_ISL_500339; EPI_ISL_510361; EPI_ISL_722214; EPI_ISL_537734; EPI_ISL_722242; EPI_ISL_722243; EPI_ISL_722268; EPI_ISL_722272; EPI_ISL_722271 | Servicio de Microbiología, Hospital Miguel Servet, Zaragoza | SeqCOVID-SPAIN consortium/IBV(CSIC) | Antonio Rezusta López et al |
| EPI_ISL_468982; EPI_ISL_691623; EPI_ISL_691666; EPI_ISL_541968; EPI_ISL_691659; EPI_ISL_541959; EPI_ISL_541961; EPI_ISL_541967; EPI_ISL_541956; EPI_ISL_541964; EPI_ISL_691643; EPI_ISL_691656 | Servicio de Microbiología, Hospital Universitario Son Espases | SeqCOVID-SPAIN consortium/IBV(CSIC) | Carla López-Causapé et al |
| EPI_ISL_683638; EPI_ISL_660266; EPI_ISL_510518; EPI_ISL_510523; EPI_ISL_660275; EPI_ISL_683626; EPI_ISL_660319; EPI_ISL_683640; EPI_ISL_671862; EPI_ISL_683639; EPI_ISL_660303; EPI_ISL_660309; EPI_ISL_654525; EPI_ISL_654531; EPI_ISL_654532; EPI_ISL_654518; EPI_ISL_654526; EPI_ISL_654522; EPI_ISL_654527; EPI_ISL_654530; EPI_ISL_654536; EPI_ISL_660322; EPI_ISL_683622; EPI_ISL_683613; EPI_ISL_671857; EPI_ISL_660267; EPI_ISL_660268; EPI_ISL_671861; EPI_ISL_660301; EPI_ISL_660289; EPI_ISL_660321; EPI_ISL_660316; EPI_ISL_660310; EPI_ISL_683650; EPI_ISL_683642; EPI_ISL_683647; EPI_ISL_683606; EPI_ISL_683607; EPI_ISL_683608; EPI_ISL_683637; EPI_ISL_683648; EPI_ISL_683636; EPI_ISL_683623; EPI_ISL_683610; EPI_ISL_683641; EPI_ISL_683621 | Servicio de Microbiología, Laboratori Clínic Metropolitana Nord. Hospital Universitari Germans Trias i Pujol. Institut d'Investigació en Ciències de la Salut Germans Trias i Pujol (IGTP) | SeqCOVID-SPAIN consortium/IBV(CSIC) | Elisa Martró et al |

| | | | |
|---|---|---|---|
| EPI_ISL_425178 | Servicio de Microbiología. Consorcio Hospital General Universitario de Valencia | Sequencing and Bioinformatics Service and Molecular Epidemiology Research Group. FISABIO-Public Health | David Navarro et al |
| EPI_ISL_419687 | Servicio de Microbiología. Consorcio Hospital General Universitario de Valencia | Sequencing and Bioinformatics Service and Molecular Epidemiology Research Group. FISABIO-Public Health | Giuseppe D'Auria et al |
| EPI_ISL_436220 | Servicio de Microbiología. Consorcio Hospital General Universitario de Valencia | Sequencing and Bioinformatics Service and Molecular Epidemiology Research Group. FISABIO-Public Health | Griselda De Marco et al B |
| EPI_ISL_416483 | Servicio de Microbiología. Consorcio Hospital General Universitario de Valencia | Sequencing and Bioinformatics Service and Molecular Epidemiology Research Group. FISABIO-Public Health | Maria Alma Bracho et al |
| EPI_ISL_416484 | Servicio de Microbiología. Consorcio Hospital General Universitario de Valencia | Sequencing and Bioinformatics Service and Molecular Epidemiology Research Group. FISABIO-Public Health | Maria Dolores Ocete et al (https://dx.doi.org/10.1101/2020.04.20.050039) |
| EPI_ISL_436223 | Servicio de Microbiología. Consorcio Hospital General Universitario de Valencia | Sequencing and Bioinformatics Service and Molecular Epidemiology Research Group. FISABIO-Public Health | Marta Pla Diaz et al |
| EPI_ISL_436361; EPI_ISL_419689 | Servicio de Microbiología. Consorcio Hospital General Universitario de Valencia | Sequencing and Bioinformatics Service and Molecular Epidemiology Research Group. FISABIO-Public Health | Neris Garcia-Gonzalez et al |
| EPI_ISL_420119 | Servicio de Microbiología. Consorcio Hospital General Universitario de Valencia | Sequencing and Bioinformatics Service and Molecular Epidemiology Research Group. FISABIO-Public Health | Paula Ruiz-Hueso et al |
| EPI_ISL_654487 | Servicio de Microbiología. Hospital Clínico Universitario de Valencia | SeqCOVID-SPAIN consortium/IBV(CSIC) | David Navarro Ortega et al |
| EPI_ISL_654609; EPI_ISL_654570; EPI_ISL_654614; EPI_ISL_654605; EPI_ISL_654589; EPI_ISL_654593; EPI_ISL_654592; EPI_ISL_654588 | Servicio de Microbiología. Hospital Universitario Donostia. OSI Donostialdea. Área de Enfermedades Infecciosas, Grupo de Infección Respiratoria y Resistencia Antimicrobiana. Instituto de Investigación Sanitaria Biodonostia | SeqCOVID-SPAIN consortium/IBV(CSIC) | Gustavo Cilla Eguiluz et al |
| EPI_ISL_510421; EPI_ISL_510477; EPI_ISL_500301; EPI_ISL_582079; EPI_ISL_565920; EPI_ISL_509637; EPI_ISL_509636; EPI_ISL_509638; EPI_ISL_565923; EPI_ISL_509642; EPI_ISL_509644; EPI_ISL_509653; EPI_ISL_509651; EPI_ISL_509649; EPI_ISL_509647; EPI_ISL_510509; EPI_ISL_509635; EPI_ISL_582052; EPI_ISL_582051; EPI_ISL_582049 | Servicio de Microbiología. Hospital Universitario Donostia. OSI Donostialdea. Área de Enfermedades Infecciosas, Grupo de Infección Respiratoria y Resistencia Antimicrobiana. Instituto de Investigación Sanitaria Biodonostia | SeqCOVID-SPAIN consortium/IBV(CSIC) | Gustavo Cilla et al |
| EPI_ISL_541115; EPI_ISL_541112; EPI_ISL_541100 | Servicio de Microbiología. Hospital Universitario Donostia. OSI Donostialdea. Área de Enfermedades Infecciosas, Grupo de Infección Respiratoria y Resistencia Antimicrobiana. Instituto de Investigación Sanitaria Biodonostia | SeqCOVID-SPAIN consortium/Institute of Biomedicine of Valencia, IBV-CSIC | Gustavo Cilla et al |
| EPI_ISL_452637; EPI_ISL_468899 | Servicio de Microbiología. Hospital Universitario Donostia. OSI Donostialdea. Área de Enfermedades Infecciosas, Grupo de Infección Respiratoria y Resistencia Antimicrobiana. Instituto de Investigación Sanitaria Biodonostia. | SeqCOVID-SPAIN consortium/IBV(CSIC) | Gustavo Cilla et al |
| EPI_ISL_529010; EPI_ISL_529012 | Servizio Igiene Epidemiologia e Sanità Pubblica (SIESP)-L'Aquila | Istituto Zooprofilattico Sperimentale dell'Abruzzo e Molise "G.Caporale" | Lorusso A et al |
| EPI_ISL_529006 | Servizio di igiene e sanità pubblica (SIESP)-Teramo | Istituto Zooprofilattico Sperimentale dell'Abruzzo e Molise "G.Caporale" | Lorusso A et al |
| EPI_ISL_529000; EPI_ISL_528998; EPI_ISL_529024; EPI_ISL_529001; EPI_ISL_529002; EPI_ISL_528997; EPI_ISL_528999; EPI_ISL_528995; EPI_ISL_528996; EPI_ISL_528994; EPI_ISL_529023; EPI_ISL_529005; EPI_ISL_529003; EPI_ISL_529004; EPI_ISL_529025 | Servizio di igiene epidemiologia e sanità pubblica (SIESP)-Chieti | Istituto Zooprofilattico Sperimentale dell'Abruzzo e Molise "G.Caporale" | Lorusso A et al |
| EPI_ISL_710554; EPI_ISL_710547; EPI_ISL_710563 | Sestre Milosrdnice University Hospital Center | Ruđer Boškovic Institute; Forensic Science Centre Ivan Vučetić; University of Zagreb Faculty of Science | Robert Belužić et al |
| EPI_ISL_416377 | Shanghai Public Health Clinical Center, Shanghai Medical College, Fudan University | National Research Center for Translational Medicine (Shanghai), Ruijin Hospital affiliated to Shanghai Jiao Tong University School of Medicine & Shanghai Public Health Clinical Center | Shengyue Wang et al |
| EPI_ISL_497950 | Shaoxing CDC | Zhejiang Provincial Center for Disease Control and Prevention | Yin Chen et al |
| EPI_ISL_636119; EPI_ISL_636113; EPI_ISL_636114; EPI_ISL_730133; EPI_ISL_730130 | Sharp HealthCare Laboratory | Andersen lab at Scripps Research | SEARCH Alliance San Diego with Aaron Harding et al |
| EPI_ISL_582125; EPI_ISL_582126 | Sheikh Khalifa Medical City | Molecular Surveillance lab Sheikh Khalifa Medical City | Amirtharaj Francis et al |
| EPI_ISL_582608; EPI_ISL_582674; EPI_ISL_582675; EPI_ISL_582609; EPI_ISL_582613; EPI_ISL_582624; EPI_ISL_582620; EPI_ISL_582626; EPI_ISL_582644; EPI_ISL_582653; EPI_ISL_582635; EPI_ISL_582655; EPI_ISL_582657; EPI_ISL_582660; EPI_ISL_582659; EPI_ISL_582676; EPI_ISL_582658; EPI_ISL_582650; EPI_ISL_582663; EPI_ISL_582683; EPI_ISL_582679; EPI_ISL_582677; EPI_ISL_582646; EPI_ISL_582648; EPI_ISL_582649; EPI_ISL_582647; EPI_ISL_582645 | Sheikh Khalifa Medical City | Molecular/Surveillance lab Sheikh Khalifa Medical City | Amirtharaj Francis et al |
| EPI_ISL_406593 | Shenzhen Key Laboratory of Pathogen and Immunity, National Clinical Research Center for Infectious Disease, Shenzhen Third People's Hospital | Shenzhen Key Laboratory of Pathogen and Immunity, National Clinical Research Center for Infectious Disease, Shenzhen Third People's Hospital | Yang Yang et al |
| EPI_ISL_437187 | Siloam Hospitals | Institute of Tropical Disease, Universitas Airlangga | Kazufumi Shimizu et al |
| EPI_ISL_482678; EPI_ISL_482679; EPI_ISL_482684; EPI_ISL_482686; EPI_ISL_482683; EPI_ISL_482697; EPI_ISL_509390; EPI_ISL_583895; EPI_ISL_610152; EPI_ISL_482681; EPI_ISL_509385 | Singapore General Hospital | Department of Microbiology | Nurdyana Abdul Rahman et al |
| EPI_ISL_406973 | Singapore General Hospital | National Public Health Laboratory | Mak et al (https://dx.doi.org/10.2807/1560-7917.ES.2020.25.13.2000305) |
| EPI_ISL_407987 | Singapore General Hospital | Programme in Emerging Infectious Diseases, Duke-NUS Medical School | Danielle E Anderson et al (https://dx.doi.org/doi/10.1101/2020.03.11.987222) |
| EPI_ISL_410537 | Singapore General Hospital, Molecular Laboratory, Division of Pathology | Programme in Emerging Infectious Diseases, Duke-NUS Medical School | Danielle E Anderson et al (https://dx.doi.org/doi/10.1101/2020.03.11.987222) |
| EPI_ISL_759957 | Siti Khodijah Hospital | Institute of Tropical Disease, Universitas Airlangga | Rima R Prasetya et al |
| EPI_ISL_534229; EPI_ISL_534228 | Skanes universitetssjukhus Lund | The Public Health Agency of Sweden | Anna-Malin Linde et al |
| EPI_ISL_475099 | Skovde/Unilabs | The Public Health Agency of Sweden | Oskar Karlsson Lindsjo et al |
| EPI_ISL_408431 | Sorbonne Université, Inserm et Assistance Publique-Hôpitaux de Paris (Pitié Salpétrière) | National Reference Center for Viruses of Respiratory Infections, Institut Pasteur, Paris | Mélanie Albert et al B (https://dx.doi.org/10.2807/1560-7917.ES.2020.25.13.2000305) |
| EPI_ISL_455099; EPI_ISL_455092; EPI_ISL_455093 | South Eastern Area Laboratory Services | NSW Health Pathology - Institute of Clinical Pathology and Medical Research; Westmead Hospital; University of Sydney | CIDM-PH et al. et al |
| EPI_ISL_629001; EPI_ISL_593740; EPI_ISL_593738; EPI_ISL_667789; EPI_ISL_593695; EPI_ISL_629030; EPI_ISL_699642; EPI_ISL_593723; EPI_ISL_593722; EPI_ISL_740867; EPI_ISL_740868; EPI_ISL_667791; EPI_ISL_490029; EPI_ISL_593724; EPI_ISL_667781; EPI_ISL_667780 | South Eastern Area Laboratory Services (SEALS) | NSW Health Pathology - Institute of Clinical Pathology and Medical Research; Westmead Hospital; University of Sydney | CIDM-PH et al. et al |
| EPI_ISL_456281; EPI_ISL_456273; EPI_ISL_456282; EPI_ISL_456278; EPI_ISL_456315; EPI_ISL_456313 | Southern Community Labs Dunedin | Institute of Environmental Science and Research (ESR) | Matt Storey et al B |
| EPI_ISL_579138; EPI_ISL_579139 | Southern Community Labs Dunedin | Institute of Environmental Science and Research (ESR) | Xiaoyun Ren et al |
| EPI_ISL_754109; EPI_ISL_754179; EPI_ISL_753700 | Specialized Lab for COVID-19 Detection, Department of Genetic Engineering and Biotechnology | Specialized Lab for COVID-19 Detection, Department of Genetic Engineering and Biotechnology | Hammadul Hoque et al |
| EPI_ISL_754059; EPI_ISL_754173 | Specialized Lab for COVID-19 Detection, Department of Genetic Engineering and Biotechnology | Specialized Lab for COVID-19 Detection, Department of Genetic Engineering and Biotechnology | Md. Nazmul Hasan et al |
| EPI_ISL_753701; EPI_ISL_754177 | Specialized Lab for COVID-19 Detection, Department of Genetic Engineering and Biotechnology | Specialized Lab for COVID-19 Detection, Department of Genetic Engineering and Biotechnology | Nurnabi Azad Jewel et al |
| EPI_ISL_753697; EPI_ISL_754176 | Specialized Lab for COVID-19 Detection, Department of Genetic Engineering and Biotechnology | Specialized Lab for COVID-19 Detection, Department of Genetic Engineering and Biotechnology | Shamsul H. Prodhan et al |
| EPI_ISL_450347; EPI_ISL_450346; EPI_ISL_450352 | St.Olavs hospital/NTNU | Institute of Genomics Core Facility, University of Tartu | Aleksandr Ianevski et al |
| EPI_ISL_412912 | State Health Office Baden-Württemberg | Charité Universitätsmedizin Berlin, Institute of Virology | Victor M Corman et al B (https://dx.doi.org/10.2807/1560-7917.ES.2020.25.13.2000305) |
| EPI_ISL_416046 | State Key Laboratory for Diagnosis and Treatment of Infectious Diseases, National Clinical Research Center for Infectious Diseases, First Affiliated Hospital, Zhejiang University School of Medicine, Hangzhou, China 310003 | State Key Laboratory for Diagnosis and Treatment of Infectious Diseases, National Clinical Research Center for Infectious Diseases, First Affiliated Hospital, Zhejiang University School of Medicine, Hangzhou, China 310003 | Hangping Yao et al (https://dx.doi.org/10.1056/NEJMoa2006100) |
| EPI_ISL_752612 | State Laboratories Division, Hawaii State Department of Health | State Laboratories Division, Hawaii State Department of Health | Pamela O'Brien et al |
| EPI_ISL_428893; EPI_ISL_428883; EPI_ISL_428862; EPI_ISL_428869; EPI_ISL_428889; EPI_ISL_428891; EPI_ISL_428887; EPI_ISL_428911; EPI_ISL_428885 | State Research Center of Virology and Biotechnology VECTOR, Department of Collection of Microorganisms | State Research Center of Virology and Biotechnology VECTOR, Department of Collection of Microorganisms | Oleg V. Pyankov et al |
| EPI_ISL_428898; EPI_ISL_428872; EPI_ISL_428874; EPI_ISL_428917; EPI_ISL_428920; EPI_ISL_428901 | State Research Center of Virology and Biotechnology VECTOR, Department of Collection of Microorganisms | State Research Center of Virology and Biotechnology VECTOR, Department of Collection of Microorganisms | Sergey A. Bodnev et al |
| EPI_ISL_451656; EPI_ISL_451662; EPI_ISL_451664; EPI_ISL_451658; EPI_ISL_451660; EPI_ISL_451659; EPI_ISL_451655 | State Sanitary Inspectorate | Laboratory of Recombinant Vaccines | Lukasz Rabalski et al |
| EPI_ISL_491045; EPI_ISL_491047; EPI_ISL_491038; EPI_ISL_491036; EPI_ISL_491087; EPI_ISL_491064; EPI_ISL_491043; EPI_ISL_491085; EPI_ISL_491046; EPI_ISL_491073; EPI_ISL_491037; EPI_ISL_491090; EPI_ISL_491049 | Suceava County Emergency Hospital | Stefan cel Mare University Metagenomics Lab | Lobiuc Andrei et al |
| EPI_ISL_491066; EPI_ISL_491044; EPI_ISL_491089; EPI_ISL_491053; EPI_ISL_491051; EPI_ISL_491091; EPI_ISL_491067; EPI_ISL_491072; EPI_ISL_491060; EPI_ISL_491086; EPI_ISL_491041; EPI_ISL_491050 | Suceava County Emergency Hospital | Stefan cel Mare University Metagenomics Lab | Lobiuc Andrei et al. et al |
| EPI_ISL_486834 | Suceava County Energency Hospital "Sf. Ioan cel Nou" | SMU Metagenomics lab | Lobiuc Andrei et al |
| EPI_ISL_648192 | Sundsvall | The Public Health Agency of Sweden | Anna-Malin Linde et al |
| EPI_ISL_534244 | Sundsvalls sjukhus | The Public Health Agency of Sweden | Anna-Malin Linde et al |
| EPI_ISL_450747 | Sunnybrook Health Sciences Centre | Department of Laboratory Medicine and Molecular Diagnostics, Sunnybrook Health Sciences Centre | Jalees A. Nasir et al |

| | | | |
|---|---|---|---|
| EPI_ISL_759960 | Surabaya Public Health Office | Institute of Tropical Disease, Universitas Airlangga | Aldise M Nastri et al |
| EPI_ISL_629010; EPI_ISL_708798 | Sydney South West Pathology Service (SSWPS) - Royal Prince Alfred Hospital - NSW Health Pathology | NSW Health Pathology - Institute of Clinical Pathology and Medical Research; Westmead Hospital; University of Sydney | CIDM-PH et al. et al |
| EPI_ISL_435057 | T.C. Sağlık Bakanlığı Adıyaman İl Sağlık Müdürlüğü Adıyaman Eğitim Ve Araştırma Hastanesi | VETAL Animal Health Products Company, BSL3+ Production Laboratuary, Turkey | Fatma Nilay Tutak et al |
| EPI_ISL_694651; EPI_ISL_694865 | TGen North | TGen North | Jolene Bowers et al |
| EPI_ISL_457733; EPI_ISL_447593; EPI_ISL_447257; EPI_ISL_457726; EPI_ISL_447252; EPI_ISL_457730; EPI_ISL_427397; EPI_ISL_427392; EPI_ISL_458029; EPI_ISL_436108; EPI_ISL_436099 | TSGH-CP molecular lab | TSGH-CP molecular lab | Cherng-Lih Perng et al |
| EPI_ISL_411926 | Taiwan Centers for Disease Control | Taiwan Centers for Disease Control | Ji-Rong Yang et al A (https://dx.doi.org/10.1080/22221751.2020.1782271) |
| EPI_ISL_411927 | Taiwan Centers for Disease Control | Taiwan Centers for Disease Control | Ji-Rong Yang et al D (https://dx.doi.org/10.2807/1560-7917.ES.2020.25.13.2000305) |
| EPI_ISL_759892; EPI_ISL_733579; EPI_ISL_733577; EPI_ISL_733573; EPI_ISL_733578; EPI_ISL_733572; EPI_ISL_733576; EPI_ISL_733574; EPI_ISL_759893 | Temporary Specimen Collection Centre | Hong Kong Department of Health | Alan K.L. Tsang et al |
| EPI_ISL_411956 | Texas Department of State Health Services | Pathogen Discovery, Respiratory Viruses Branch, Division of Viral Diseases, Centers for Disease Control and Prevention | Krista Queen et al A |
| EPI_ISL_429089 | The First Affiliated Hospital of Guangzhou Medical University | BGI-shenzhen & The First Affiliated Hospital of Guangzhou Medical University | Yanqun Wang et al |
| EPI_ISL_429080 | The First Affiliated Hospital of Guangzhou Medical University | BGI-shenzhen & The First Affiliated Hospital of Guangzhou Medical University | et al |
| EPI_ISL_693663; EPI_ISL_693668; EPI_ISL_693684; EPI_ISL_577635; EPI_ISL_626586; EPI_ISL_584081; EPI_ISL_626574; EPI_ISL_577634; EPI_ISL_626576; EPI_ISL_560408; EPI_ISL_560409; EPI_ISL_560412; EPI_ISL_560410; EPI_ISL_560411; EPI_ISL_577625; EPI_ISL_547967; EPI_ISL_546436; EPI_ISL_577624; EPI_ISL_547966; EPI_ISL_693675; EPI_ISL_530350; EPI_ISL_737027; EPI_ISL_626605; EPI_ISL_626580; EPI_ISL_626591; EPI_ISL_626606; EPI_ISL_626585; EPI_ISL_693687; EPI_ISL_545580; EPI_ISL_541332; EPI_ISL_541333; EPI_ISL_541334; EPI_ISL_577633; EPI_ISL_545954; EPI_ISL_545957; EPI_ISL_545956; EPI_ISL_546935; EPI_ISL_577627; EPI_ISL_577628; EPI_ISL_626588; EPI_ISL_577637; EPI_ISL_626575; EPI_ISL_547968; EPI_ISL_546936; EPI_ISL_541336; EPI_ISL_541337; EPI_ISL_577623; EPI_ISL_584074; EPI_ISL_584076; EPI_ISL_584077; EPI_ISL_539777; EPI_ISL_626570; EPI_ISL_539493; EPI_ISL_626593; EPI_ISL_577626; EPI_ISL_541082; EPI_ISL_584075; EPI_ISL_584078; EPI_ISL_693680; EPI_ISL_577638; EPI_ISL_626587; EPI_ISL_626617; EPI_ISL_626608; EPI_ISL_626603; EPI_ISL_660585; EPI_ISL_737013; EPI_ISL_693667; EPI_ISL_737029; EPI_ISL_660588; EPI_ISL_626583; EPI_ISL_577636; EPI_ISL_693666; EPI_ISL_693669; EPI_ISL_626620; EPI_ISL_626582; EPI_ISL_626589; EPI_ISL_693686; EPI_ISL_737020; EPI_ISL_660560; EPI_ISL_660567; EPI_ISL_626572; EPI_ISL_693674; EPI_ISL_737011; EPI_ISL_737018; EPI_ISL_660574; EPI_ISL_626602; EPI_ISL_693683; EPI_ISL_626619; EPI_ISL_693672; EPI_ISL_737015; EPI_ISL_693685; EPI_ISL_693661; EPI_ISL_737023; EPI_ISL_626577; EPI_ISL_626571; EPI_ISL_626584; EPI_ISL_693665; EPI_ISL_693677; EPI_ISL_626578; EPI_ISL_626600; EPI_ISL_626599; EPI_ISL_737025; EPI_ISL_626612; EPI_ISL_577629; EPI_ISL_584080; EPI_ISL_577632; EPI_ISL_577631; EPI_ISL_584079; EPI_ISL_584073 | The National Institute of Public Health | State Veterinary Institute Prague | Nagy et al |
| EPI_ISL_471547; EPI_ISL_471553; EPI_ISL_471528; EPI_ISL_471530 | The National Institute of Public Health | State Veterinary Institute Prague and The National Institute of Public Health | Nagy et al |
| EPI_ISL_541335 | The National Institute of Public Health | Sídlištní 136/24 165 03, Prague Czech Republic | Nagy et al |
| EPI_ISL_513512; EPI_ISL_491118; EPI_ISL_491119 | The National Institute of Public Health | The National Institute of Public Health and State Veterinary Institute Prague | Nagy et al |
| EPI_ISL_437519; EPI_ISL_434572 | The National Institute of Public Health Center for Epidemiology and Microbiology | The National Institute of Public Health Center for Epidemiology and Microbiology | Alexander Nagy et al |
| EPI_ISL_417765; EPI_ISL_417574; EPI_ISL_417843; EPI_ISL_417769; EPI_ISL_417862; EPI_ISL_417840; EPI_ISL_417688; EPI_ISL_417828; EPI_ISL_417829; EPI_ISL_417598; EPI_ISL_424598; EPI_ISL_417854; EPI_ISL_417837; EPI_ISL_417804; EPI_ISL_417622; EPI_ISL_417851; EPI_ISL_417569; EPI_ISL_417746; EPI_ISL_417550; EPI_ISL_417770; EPI_ISL_417609 | The National University Hospital of Iceland | deCODE genetics | Daniel F Gudbjartsson et al (https://dx.doi.org/10.1056/NEJMoa2006100) |
| EPI_ISL_648156; EPI_ISL_648197; EPI_ISL_648181; EPI_ISL_648138; EPI_ISL_648134; EPI_ISL_648139; EPI_ISL_648182; EPI_ISL_648191; EPI_ISL_648135; EPI_ISL_648132; EPI_ISL_648185; EPI_ISL_648150; EPI_ISL_648187; EPI_ISL_648188 | The Public Health Agency of Sweden | The Public Health Agency of Sweden | Anna-Malin Linde et al |
| EPI_ISL_710582 | The Public Health Agency of Sweden | The Public Health Agency of Sweden | Department of Microbiology et al |
| EPI_ISL_738133 | The Public Health Agency of Sweden | The Public Health Agency of Sweden | Oskar Karlsson Lindsjo et al |
| EPI_ISL_754229; EPI_ISL_754230; EPI_ISL_754234; EPI_ISL_754235; EPI_ISL_754232; EPI_ISL_754233; EPI_ISL_754231 | The Republican Research and Practical Center for Epidemiology and Microbiology (RRPCEM) | WHO National Influenza Centre Russian Federation | Elena Gasich et al |
| EPI_ISL_480033 | Tochigi Prefectural Institute of Public Health and Environmental Science | Pathogen Genomics Center, National Institute of Infectious Diseases | Tsuyoshi Sekizuka et al C (https://www.medrxiv.org/content/10.1101/2020.07.01.20143958v1) |
| EPI_ISL_479888 | Tokyo Metropolitan Institute of Public Health | Pathogen Genomics Center, National Institute of Infectious Diseases | Tsuyoshi Sekizuka et al C (https://www.medrxiv.org/content/10.1101/2020.07.01.20143958v1) |
| EPI_ISL_755745; EPI_ISL_755744; EPI_ISL_755749; EPI_ISL_755742; EPI_ISL_755748; EPI_ISL_755761; EPI_ISL_755764; EPI_ISL_755803; EPI_ISL_755812; EPI_ISL_755813; EPI_ISL_755804; EPI_ISL_729311; EPI_ISL_729312; EPI_ISL_586277; EPI_ISL_586322; EPI_ISL_591188; EPI_ISL_591195; EPI_ISL_755903; EPI_ISL_755695; EPI_ISL_729274; EPI_ISL_729275; EPI_ISL_755808; EPI_ISL_586420; EPI_ISL_586333; EPI_ISL_729268; EPI_ISL_755670; EPI_ISL_755882; EPI_ISL_755893 | Toronto Invasive Bacterial Diseases Network | McMaster University | Allison McGeer et al |
| EPI_ISL_755736; EPI_ISL_755717; EPI_ISL_755720 | Toronto Invasive Bacterial Diseases Network | McMaster University | David Richardson et al |
| EPI_ISL_542988; EPI_ISL_610258 | TriCore Reference Laboratories | Center for Global Health, University of New Mexico Health Sciences Center | Daryl Domman et al |
| EPI_ISL_717696; EPI_ISL_717698; EPI_ISL_717695; EPI_ISL_717694; EPI_ISL_717700; EPI_ISL_717699; EPI_ISL_756362; EPI_ISL_717692; EPI_ISL_717697; EPI_ISL_756363 | Trinidad Public Health Laboratory | Carrington Lab, Department of PreClinical Sciences, Faculty of Medical Sciences, The University of the West Indies | Nikita S. D. Sahadeo et al |
| EPI_ISL_539819; EPI_ISL_539818 | Tuen Mun Hospital | Hong Kong Department of Health | Alan K.L. Tsang et al |
| EPI_ISL_745140 | Tygerberg Hospital wc TBH | National Health Laboratory Service (NHLS), Tygerberg | Susan Engelbrecht et al |
| EPI_ISL_444610 | U.S. Naval Medical Research Center Biological Defense Research Directorate | U.S. Naval Medical Research Center Biological Defense Research Directorate | Voegtly et al |
| EPI_ISL_483333 | UC San Diego Center for Advanced Laboratory Medicine | Andersen lab at Scripps Research | SEARCH Alliance San Diego with David Pride et al |
| EPI_ISL_418583; EPI_ISL_418582; EPI_ISL_418548 | UCD National Virus Reference Laboratory | UCD National Virus Reference Laboratory | Michael Carr et al |
| EPI_ISL_537558 | UCLA Pathology Clinical Microbiology Lab | Kruglyak Lab | Guo et al. et al |
| EPI_ISL_755179 | UCSD EXCITE lab | Andersen lab at Scripps Research | SEARCH Alliance San Diego et al |
| EPI_ISL_648125; EPI_ISL_648124; EPI_ISL_628760; EPI_ISL_628761 | UHAS COVID-19 Lab | UHAS COVID-19 Lab | Kwabena O. Duedu et al |
| EPI_ISL_747477; EPI_ISL_747481; EPI_ISL_747478 | ULSS 5 Polesana | Istituto Zooprofilattico Sperimentale delle Venezie | Adelaide Milani et al |
| EPI_ISL_636465; EPI_ISL_636466 | ULSS6 Euganea | Istituto Zooprofilattico Sperimentale delle Venezie | Adelaide Milani et al |
| EPI_ISL_747473 | ULSS9 Distretto di Bussolengo | Istituto Zooprofilattico Sperimentale delle Venezie | Adelaide Milani et al |
| EPI_ISL_522859 | ULSS9 Scaligera | Istituto Zooprofilattico Sperimentale delle Venezie | Adelaide Milani et al |
| EPI_ISL_733234; EPI_ISL_732992; EPI_ISL_733225; EPI_ISL_733226; EPI_ISL_732996; EPI_ISL_560311; EPI_ISL_560309; EPI_ISL_733223 | UMMC-Health | WHO National Influenza Centre Russian Federation | Andrey Komissarov et al A |
| EPI_ISL_583956; EPI_ISL_583957; EPI_ISL_583961; EPI_ISL_583965 | UOC Microbiologia e Virologia, Azienda Ospedaliera Universitaria Senese, Siena, Italy | Dipartimento di Biotecnologie Mediche | Maria Grazia Cusi et al |
| EPI_ISL_416723 | UW Virology Lab | UW Virology Lab | Pavitra Roychoudhury et al A (https://doi.dx.org/10.1101/2020.04.02.20051417) |
| EPI_ISL_576422 | UW Virology Lab | UW Virology Lab | Pavitra Roychoudhury et al B |
| EPI_ISL_734553; EPI_ISL_734869; EPI_ISL_735014; EPI_ISL_735012; EPI_ISL_735033; EPI_ISL_735045; EPI_ISL_735075; EPI_ISL_735079; EPI_ISL_735089; EPI_ISL_735161; EPI_ISL_735209; EPI_ISL_735150; EPI_ISL_735105; EPI_ISL_735103; EPI_ISL_735128; EPI_ISL_735170; EPI_ISL_734952; EPI_ISL_734994; EPI_ISL_734992; EPI_ISL_735017; EPI_ISL_735015; EPI_ISL_735011; EPI_ISL_734694; EPI_ISL_734837; EPI_ISL_734803; EPI_ISL_734882; EPI_ISL_734839; EPI_ISL_734857; EPI_ISL_734910; EPI_ISL_734916; EPI_ISL_734817; EPI_ISL_734909; EPI_ISL_734903; EPI_ISL_734864; EPI_ISL_734841; EPI_ISL_734603; EPI_ISL_738401; EPI_ISL_738373; EPI_ISL_738385; EPI_ISL_738490; EPI_ISL_738393; EPI_ISL_738440; EPI_ISL_738403; EPI_ISL_738379; EPI_ISL_738424; EPI_ISL_738454; EPI_ISL_735031; EPI_ISL_734648; EPI_ISL_734580; EPI_ISL_734581; EPI_ISL_734623; EPI_ISL_734769; EPI_ISL_734771; EPI_ISL_734758; EPI_ISL_738220; EPI_ISL_734786; EPI_ISL_734772; EPI_ISL_734778; EPI_ISL_734777; EPI_ISL_734727; EPI_ISL_734738; EPI_ISL_734773; EPI_ISL_734774; EPI_ISL_735238; EPI_ISL_735237; EPI_ISL_734959; EPI_ISL_734930; EPI_ISL_735101; EPI_ISL_738453; EPI_ISL_734748; EPI_ISL_734756; EPI_ISL_734493; EPI_ISL_734768; EPI_ISL_734552; EPI_ISL_734943; EPI_ISL_734886; EPI_ISL_734808; EPI_ISL_734861; EPI_ISL_734790; EPI_ISL_734826; EPI_ISL_734816; EPI_ISL_734859; EPI_ISL_734858; EPI_ISL_734853; EPI_ISL_735042; EPI_ISL_734980; EPI_ISL_735006; EPI_ISL_738489; EPI_ISL_738488; EPI_ISL_738472; EPI_ISL_735084; EPI_ISL_735077; EPI_ISL_735182; EPI_ISL_735180; EPI_ISL_735157; EPI_ISL_735146; EPI_ISL_735198 | UZ Leuven, National Reference Laboratory for Coronaviruses, Laboratory Medicine, Leuven, Belgium | KU Leuven, Rega Institute, Clinical and Epidemiological Virology | Tony Wawina-Bokalanga et al |
| EPI_ISL_737954; EPI_ISL_737951; EPI_ISL_737970; EPI_ISL_738005; EPI_ISL_737986; EPI_ISL_738028; EPI_ISL_738027; EPI_ISL_738031; EPI_ISL_738035; EPI_ISL_738033; EPI_ISL_738037; EPI_ISL_738034; EPI_ISL_738032; EPI_ISL_738036; EPI_ISL_737962; EPI_ISL_737939; EPI_ISL_737985; EPI_ISL_737931; EPI_ISL_738010; EPI_ISL_738011; EPI_ISL_737940; EPI_ISL_737976; EPI_ISL_737990; EPI_ISL_738000; EPI_ISL_737980; EPI_ISL_738006; EPI_ISL_737997; EPI_ISL_737945; EPI_ISL_737974; EPI_ISL_738022; EPI_ISL_737959; EPI_ISL_737956; EPI_ISL_737958; EPI_ISL_737935; EPI_ISL_737953; EPI_ISL_738038; EPI_ISL_738043; EPI_ISL_738042; EPI_ISL_738040; EPI_ISL_738039; EPI_ISL_738041; EPI_ISL_737955; EPI_ISL_737947; EPI_ISL_737967; EPI_ISL_737932; EPI_ISL_737977; EPI_ISL_737979; EPI_ISL_737972; EPI_ISL_737978; EPI_ISL_737948; EPI_ISL_737949; EPI_ISL_737952; EPI_ISL_737966 | Uganda Central Public Health Lab and Uganda Virus Research Institute | MRC/UVRI & LSHTM Uganda Research Unit | Matthew Cotten et al |
| EPI_ISL_451199; EPI_ISL_451183; EPI_ISL_451184; EPI_ISL_451193; EPI_ISL_451192; EPI_ISL_451190; EPI_ISL_451198; EPI_ISL_451201; EPI_ISL_451197 | Uganda Virus Research Institute | MRC/UVRI & LSHTM Uganda Research Unit | Dan Lule Bugembe et al (https://dx.doi.org/10.3201/eid2610.202575) |
| EPI_ISL_615094; EPI_ISL_615095 | Umea klinisk mikrobiologi | The Public Health Agency of Sweden | Anna-Malin Linde et al |
| EPI_ISL_735409 | Unidade de Pronto Atendimento Carlos Lourenco | Instituto Adolfo Lutz, Interdiciplinary Procedures Center, Strategic Laboratory | Claudio Tavares Sacchi et al B |
| EPI_ISL_661298; EPI_ISL_661296; EPI_ISL_661295 | Unilabs | The Public Health Agency of Sweden | Department of Microbiology et al |
| EPI_ISL_594154 | Unilabs Eskilstuna | The Public Health Agency of Sweden | Anna-Malin Linde et al |
| EPI_ISL_510829; EPI_ISL_510828 | Unilabs Eskilstuna | The Public Health Agency of Sweden | Oskar Karlsson Lindsjo et al |
| EPI_ISL_549123; EPI_ISL_635132; EPI_ISL_590882; EPI_ISL_738314; EPI_ISL_420143; EPI_ISL_708063; EPI_ISL_668409 | Unilabs Laboratory Medicine | Norwegian Institute of Public Health, Department of Virology | Kathrine Stene-Johansen et al |

| | | | |
|---|---|---|---|
| EPI_ISL_763015; EPI_ISL_763022; EPI_ISL_763045; EPI_ISL_763049; EPI_ISL_763012; EPI_ISL_763013; EPI_ISL_763018; EPI_ISL_763010; EPI_ISL_763014; EPI_ISL_763046; EPI_ISL_763048; EPI_ISL_763031; EPI_ISL_763047; EPI_ISL_765893; EPI_ISL_763032; EPI_ISL_763033; EPI_ISL_763037; EPI_ISL_763036; EPI_ISL_763051; EPI_ISL_763041; EPI_ISL_763050; EPI_ISL_763040; EPI_ISL_763043; EPI_ISL_763056; EPI_ISL_763053; EPI_ISL_763055; EPI_ISL_763021; EPI_ISL_763026; EPI_ISL_763002; EPI_ISL_763008; EPI_ISL_763006; EPI_ISL_763025; EPI_ISL_765894; EPI_ISL_763023; EPI_ISL_763024; EPI_ISL_763042; EPI_ISL_763027; EPI_ISL_763004; EPI_ISL_763019; EPI_ISL_763011; EPI_ISL_763057; EPI_ISL_763001; EPI_ISL_763007; EPI_ISL_763058; EPI_ISL_763030; EPI_ISL_763060; EPI_ISL_763028; EPI_ISL_763052; EPI_ISL_762999; EPI_ISL_763005; EPI_ISL_763009; EPI_ISL_763000; EPI_ISL_763038; EPI_ISL_763034; EPI_ISL_763059; EPI_ISL_763035; EPI_ISL_763003; EPI_ISL_763039; EPI_ISL_763061; EPI_ISL_763029; EPI_ISL_763044; EPI_ISL_763054 | Unit 17: Influenza & Other Respiratory Viruses, German National Influenza Center | Project group Epidemiology of Highly Pathogenic Microorganisms, Robert Koch-Institute | Ariane Düx et al |
| EPI_ISL_411951 | Unit for Laboratory Development and Technology Transfer, Public Health Agency of Sweden | Unit for Laboratory Development and Technology Transfer, Public Health Agency of Sweden | Bengner et al (https://dx.doi.org/10.2807/1560-7917.ES.2020.25.13.2000305) |
| EPI_ISL_671654; EPI_ISL_671657; EPI_ISL_671666; EPI_ISL_609811; EPI_ISL_671669; EPI_ISL_570019; EPI_ISL_569977; EPI_ISL_569985; EPI_ISL_570009; EPI_ISL_569972; EPI_ISL_569996; EPI_ISL_526275; EPI_ISL_526259; EPI_ISL_570022; EPI_ISL_464011; EPI_ISL_526255; EPI_ISL_569967 | Unity Health Toronto | Ontario Institute for Cancer Research | Ramzi Fattouh et al |
| EPI_ISL_525468; EPI_ISL_525469; EPI_ISL_525467 | Universidad Iberoamericana | International Centre for Genetic Engineering and Biotechnology (ICGEB) and ARGO Open Lab Platform | Robert Paulino-Ramirez et al |
| EPI_ISL_523811; EPI_ISL_523812 | Universidad Iberoamericana, Instituto de Medicina Tropical & Salud Global | International Centre for Genetic Engineering and Biotechnology (ICGEB) and ARGO Open Lab Platform | Robert Paulino-Ramirez et al |
| EPI_ISL_539783; EPI_ISL_697796 | Universidad Regional Amazonica IKIAM | Institute of Microbiology, Universidad San Francisco de Quito | Fabian Aguilar et al |
| EPI_ISL_537341 | Universidad de León | SeqCOVID-SPAIN consortium/IBV(CSIC) | Ana Carvajal et al |
| EPI_ISL_583561 | Universitaetsklinik für Innere Medizin II Innsbruck | Bergthaler laboratory, CeMM Research Center for Molecular Medicine of the Austrian Academy of Sciences | Alexandra Popa et al A |
| EPI_ISL_475932; EPI_ISL_475936 | Universitaetsklinik für Innere Medizin II Innsbruck | Bergthaler laboratory, CeMM Research Center for Molecular Medicine of the Austrian Academy of Sciences | Alexandra Popa et al B (https://dx.doi.org/10.1101/2020.07.15.204339v1) |
| EPI_ISL_437961; EPI_ISL_437969; EPI_ISL_437952; EPI_ISL_437938 | Universitaetsklinik für Innere Medizin II Innsbruck | Bergthaler laboratory, CeMM Research Center for Molecular Medicine of the Austrian Academy of Sciences | Alexandra Popa et al B (https://www.biorxiv.org/content/10.1101/2020.07.15.204339v1) |
| EPI_ISL_529962 | Universitas Airlangga Hospital | Institute of Tropical Disease, Universitas Airlangga | Jezzy R Dewantari et al |
| EPI_ISL_529963 | Universitas Airlangga Hospital | Institute of Tropical Disease, Universitas Airlangga | Rima R Prasetya et al |
| EPI_ISL_534247; EPI_ISL_534224; EPI_ISL_560978 | Universitetssjukhuset i Linkoping | The Public Health Agency of Sweden | Anna-Malin Linde et al |
| EPI_ISL_462990 | University Clinical Centre of the Republic of Srpska | University of Sarajevo, Veterinary Faculty | Teufik et al (https://dx.doi.org/10.1101/2020.06.19.160606) |
| EPI_ISL_462753 | University Clinical Hospital of Mostar | University of Sarajevo Veterinary Faculty | Goletic et al (https://dx.doi.org/10.1101/2020.06.19.160606) |
| EPI_ISL_665162 | University College London Hospital | COVID-19 Genomics UK (COG-UK) Consortium | Judith Heaney et al |
| EPI_ISL_724663 | University College London, Great Ormond Street Hospital for Children NHS Foundation Trust, Imperial College Healthcare NHS Trust | COVID-19 Genomics UK (COG-UK) Consortium | Sergi Castellano et al |
| EPI_ISL_581816; EPI_ISL_528063; EPI_ISL_527917; EPI_ISL_581837; EPI_ISL_528276; EPI_ISL_581762; EPI_ISL_581934; EPI_ISL_581892; EPI_ISL_528065; EPI_ISL_528087; EPI_ISL_528319; EPI_ISL_581897; EPI_ISL_581925; EPI_ISL_581692 | University Hospital Basel, Clinical Virology | University Hospital Basel, Clinical Bacteriology | Madlen Stange et al |
| EPI_ISL_418273 | University Hospital Basel, Clinical Virology | University Hospital Basel, Labormedizin | Hirsch et al |
| EPI_ISL_631381; EPI_ISL_631386; EPI_ISL_631379; EPI_ISL_631378; EPI_ISL_631380; EPI_ISL_631383; EPI_ISL_631384; EPI_ISL_631382; EPI_ISL_631377; EPI_ISL_631385; EPI_ISL_631388 | University Hospital Cologne | Center of Medical Microbiology, Virology, and Hospital Hygiene, University of Duesseldorf | Maximilian Damagnez et al |
| EPI_ISL_710553; EPI_ISL_710561; EPI_ISL_710569; EPI_ISL_710564; EPI_ISL_710574; EPI_ISL_710570; EPI_ISL_710559; EPI_ISL_710572; EPI_ISL_710565; EPI_ISL_710555; EPI_ISL_710568; EPI_ISL_710571; EPI_ISL_710556; EPI_ISL_710560; EPI_ISL_710567; EPI_ISL_710566; EPI_ISL_710558; EPI_ISL_710573; EPI_ISL_710548; EPI_ISL_710557; EPI_ISL_710550; EPI_ISL_710551; EPI_ISL_710552; EPI_ISL_710549; EPI_ISL_710562 | University Hospital Dubrava | Ruđer Boškovic Institute; Forensic Science Centre Ivan Vučetić; University of Zagreb Faculty of Science | Robert Belužić et al |
| EPI_ISL_681739; EPI_ISL_681757; EPI_ISL_681735; EPI_ISL_681744; EPI_ISL_681724; EPI_ISL_681709; EPI_ISL_681734; EPI_ISL_681733; EPI_ISL_681749; EPI_ISL_681771; EPI_ISL_681753; EPI_ISL_681742; EPI_ISL_681741 | University Hospital Limerick | Irish Coronavirus Sequencing Consortium - Teagasc Moorepark | Paul Cotter et al |
| EPI_ISL_483677; EPI_ISL_483670; EPI_ISL_483682 | University Hospital Zurich | Department of Biosystems Science and Engineering, ETH Zürich | Christian Beisel et al |
| EPI_ISL_730634 | University Hospital Zürich | Institute of Medical Virology, University of Zurich | Stefan Schmutz et al A |
| EPI_ISL_454588; EPI_ISL_454581; EPI_ISL_454592; EPI_ISL_454578; EPI_ISL_454583 | University Hospital for Infectious Diseases "Dr. Fran Mihaljević", Research Unit | University of Zagreb, Centre for research and knowledge transfer in biotechnology | Ivan-Christian Kurolt et al |
| EPI_ISL_759987; EPI_ISL_759986; EPI_ISL_420152; EPI_ISL_708067; EPI_ISL_708065; EPI_ISL_708068; EPI_ISL_708066; EPI_ISL_635137; EPI_ISL_635179; EPI_ISL_635180; EPI_ISL_635070; EPI_ISL_635073; EPI_ISL_635181; EPI_ISL_635069 | University Hospital of Northern Norway, Department for Microbiology and Infectious Disease Control | Norwegian Institute of Public Health, Department of Virology | Kathrine Stene-Johansen et al |
| EPI_ISL_415698; EPI_ISL_415699; EPI_ISL_415702; EPI_ISL_415700; EPI_ISL_415457; EPI_ISL_415455; EPI_ISL_415459 | University Hospitals of Geneva Laboratory of Virology | University Hospitals of Geneva Laboratory of Virology | Laubscher F. et al |
| EPI_ISL_738144 | University of Bari Biomedical Sciences and Human Oncology | University of Bari Biomedical Sciences and Human Oncology | Maria Chironna et al |
| EPI_ISL_730652 | University of Bari, Valenzano, Italy | Dept. Food safety, Nutrition and Veterinary Public Health, Istituto superiore di sanità | N. Decaro et al |
| EPI_ISL_730653 | University of Bari, Valenzano, Italy | Istituto Zooprofilattico Sperimentale dell'Abruzzo e del Molise "G. Caporale". | N. Decaro et al |
| EPI_ISL_735500 | University of Chittagong | Central Biological Research Laboratory and Department of Biochemistry and Molecular Biology | H. M. Abdullah Al Masud et al |
| EPI_ISL_735499; EPI_ISL_735498 | University of Chittagong | Central Biological Research Laboratory and Department of Biochemistry and Molecular Biology | Mohammad Omar Faruque et al |
| EPI_ISL_671429; EPI_ISL_671437; EPI_ISL_671426; EPI_ISL_671440; EPI_ISL_671438; EPI_ISL_671433; EPI_ISL_671425; EPI_ISL_671435; EPI_ISL_671452; EPI_ISL_671445; EPI_ISL_671448; EPI_ISL_671455; EPI_ISL_671461; EPI_ISL_671485; EPI_ISL_671470; EPI_ISL_671458; EPI_ISL_671442; EPI_ISL_678392; EPI_ISL_671494; EPI_ISL_671434; EPI_ISL_671424; EPI_ISL_671421; EPI_ISL_671449; EPI_ISL_671444; EPI_ISL_671453; EPI_ISL_671467; EPI_ISL_671484; EPI_ISL_671481; EPI_ISL_671464; EPI_ISL_671463; EPI_ISL_476071; EPI_ISL_477010; EPI_ISL_671422; EPI_ISL_671441; EPI_ISL_671420; EPI_ISL_671423; EPI_ISL_671454; EPI_ISL_671427; EPI_ISL_678389; EPI_ISL_477009; EPI_ISL_476074; EPI_ISL_476075; EPI_ISL_476072; EPI_ISL_476077; EPI_ISL_477012; EPI_ISL_477011; EPI_ISL_476068; EPI_ISL_476073; EPI_ISL_476076; EPI_ISL_477013; EPI_ISL_476070; EPI_ISL_477008 | University of Debrecen, Department of Medical Microbiology | National Laboratory of Virology, Szentágothai Research Centre | Endre Gábor Tóth et al |
| EPI_ISL_471992 | University of Exeter | COVID-19 Genomics UK (COG-UK) Consortium | Ben Temperton et al |
| EPI_ISL_540465; EPI_ISL_540464; EPI_ISL_540447 | University of Liège COVID-19 testing center | GIGA Medical Genomics | Keith Durkin et al A |
| EPI_ISL_707813 | University of Michigan Clinical Microbiology Laboratory | Lauring Lab, University of Michigan, Department of Microbiology and Immunology | Valesano et al |
| EPI_ISL_677742; EPI_ISL_677762; EPI_ISL_677729; EPI_ISL_677730; EPI_ISL_677793; EPI_ISL_677787; EPI_ISL_677816; EPI_ISL_677815; EPI_ISL_677799; EPI_ISL_677807; EPI_ISL_677785; EPI_ISL_677739; EPI_ISL_677766; EPI_ISL_677737; EPI_ISL_677735; EPI_ISL_677736; EPI_ISL_677733; EPI_ISL_677734; EPI_ISL_677738; EPI_ISL_677732; EPI_ISL_677728; EPI_ISL_477623; EPI_ISL_477620; EPI_ISL_477624; EPI_ISL_477618; EPI_ISL_477615; EPI_ISL_677767 | University of Szeged, Institute of Clinical Microbiology | National Laboratory of Virology, Szentágothai Research Centre | Endre Gábor Tóth et al |
| EPI_ISL_485389; EPI_ISL_485396; EPI_ISL_485395 | University of Ulsan College of Medicine and Asan Medical Center | University of Ulsan College of Medicine and Asan Medical Center | Kuenyoul Park et al |
| EPI_ISL_418809 | University of Wisconsin - Madison: Influenza Research Institute | University of Wisconsin Madison, AIDS Vaccine Research Laboratories | Katarina Braun et al |
| EPI_ISL_536669; EPI_ISL_605553; EPI_ISL_677599; EPI_ISL_677446; EPI_ISL_509977; EPI_ISL_516481; EPI_ISL_541617 | University of Wisconsin-Madison AIDS Vaccine Research Laboratories | University of Wisconsin-Madison AIDS Vaccine Research Laboratories | Gage Moreno et al |
| EPI_ISL_445231 | Uppsala Narakut Aleris | The Public Health Agency of Sweden | Annika Nilsson et al |
| EPI_ISL_475516; EPI_ISL_475526 | Uppsala Narakut Aleris | The Public Health Agency of Sweden | Oskar Karlsson Lindsjo et al |
| EPI_ISL_582780; EPI_ISL_623079; EPI_ISL_648130 | Uppsala klinisk mikrobiologi | The Public Health Agency of Sweden | Anna-Malin Linde et al |
| EPI_ISL_676528; EPI_ISL_676525; EPI_ISL_676523 | Uppsala klinisk mikrobiologi | The Public Health Agency of Sweden | Department of Microbiology et al |
| EPI_ISL_475119; EPI_ISL_475151; EPI_ISL_475118 | Uppsala klinisk mikrobiologi | The Public Health Agency of Sweden | Oskar Karlsson Lindsjo et al |
| EPI_ISL_708815; EPI_ISL_708817; EPI_ISL_708816; EPI_ISL_708818 | Urban Institute for Disease Prevention and Control | National Institute of Health, Department of Medical Sciences, Ministry of Public Health, Thailand | Pilailuk Okada et al |
| EPI_ISL_746865; EPI_ISL_417026 | Utah Public Health Laboratory | Utah Public Health Laboratory | Erin Young et al |
| EPI_ISL_710576 | VC Sorgenfrimottagningen | The Public Health Agency of Sweden | Department of Microbiology et al |
| EPI_ISL_708820 | Vajira Hospital | National Institute of Health, Department of Medical Sciences, Ministry of Public Health, Thailand | Pilailuk Okada et al |
| EPI_ISL_700462 | Vanguard CHC wc VGC | NHLS/UCT | Arash Iranzadeh et al |
| EPI_ISL_445233 | Vardcentralen Brinken | The Public Health Agency of Sweden | Agnes Wigh et al |
| EPI_ISL_590919; EPI_ISL_549051; EPI_ISL_590983; EPI_ISL_668390 | Vestfold Hospital, Toensberg Department of Microbiology | Norwegian Institute of Public Health, Department of Virology | Kathrine Stene-Johansen et al |
| EPI_ISL_420137 | Vestfold Hospital, Tonsberg Department of Microbiology | Norwegian Institute of Public Health, Department of Virology | Kathrine Stene-Johansen et al |
| EPI_ISL_437435; EPI_ISL_437436 | Veterinary Specialized Institue Kraljevo | Veterinary Specialized Institue Kraljevo | Dejan Vidanovic. Bojana Tesovic et al |
| EPI_ISL_644572; EPI_ISL_644583; EPI_ISL_582530; EPI_ISL_644571; EPI_ISL_644565; EPI_ISL_582529; EPI_ISL_582527; EPI_ISL_582528; EPI_ISL_582531; EPI_ISL_644574; EPI_ISL_582526; EPI_ISL_644566; EPI_ISL_644580; EPI_ISL_644581; EPI_ISL_582525; EPI_ISL_644579; EPI_ISL_644567; EPI_ISL_644573; EPI_ISL_582532 | Veterinary Specialized Institute "Kraljevo", Serbia | Veterinary Specialized Institute "Kraljevo", Serbia | Vidanovic et al |
| EPI_ISL_678491; EPI_ISL_678489; EPI_ISL_678486 | Veterinary Specialized Institute "Sabac", Serbia | Veterinary Specialized Institute "Kraljevo", Serbia | Vidanovic et al |
| EPI_ISL_454795 | Veterinary Specialized Institute Kraljevo | Veterinary Specialized Institute Kraljevo | Vidanovic et al |

| | | | |
|---|---|---|---|
| EPI_ISL_708823 | Vibharam Hospital | National Institute of Health, Department of Medical Sciences, Ministry of Public Health, Thailand | Pilailuk Okada et al |
| EPI_ISL_700596 | Victoria Hospital wc VHW | NHLS/UCT | Arash Iranzadeh et al |
| EPI_ISL_456420 | Victorian Infectious Diseases Reference Laboratory (VIDRL) | Microbiological Diagnostic Unit Public Health Laboratory and Victorian Infectious Diseases Reference Laboratory, Doherty Institute | Caly L. et al B |
| EPI_ISL_522105; EPI_ISL_520253; EPI_ISL_480728; EPI_ISL_480734; EPI_ISL_480781; EPI_ISL_480780; EPI_ISL_480779; EPI_ISL_480778; EPI_ISL_522060; EPI_ISL_521499; EPI_ISL_521980; EPI_ISL_480705; EPI_ISL_518169; EPI_ISL_522110; EPI_ISL_522109 | Victorian Infectious Diseases Reference Laboratory (VIDRL) | VIDRL and MDU-PHL | Caly L. et al B |
| EPI_ISL_561517 | Victorian Infectious Diseases Reference Laboratory (VIDRL) | VIDRL and MDU-PHL | Caly et al B |
| EPI_ISL_416415; EPI_ISL_419774; EPI_ISL_419888 | Victorian Infectious Diseases Reference Laboratory (VIDRL) | Victorian Infectious Diseases Reference Laboratory and Microbiological Diagnostic Unit Public Health Laboratory, Doherty Institute | Caly L. et al A (https://dx.doi.org/10.1101/2020.05.12.20099929) |
| EPI_ISL_560394; EPI_ISL_560393; EPI_ISL_560402; EPI_ISL_560398; EPI_ISL_560395; EPI_ISL_560399; EPI_ISL_560396; EPI_ISL_560391; EPI_ISL_560404; EPI_ISL_560403; EPI_ISL_560401; EPI_ISL_560392; EPI_ISL_560400; EPI_ISL_560397; EPI_ISL_560405 | Vilnius University Hospital Santaros Klinikos, Vilnius University | Institute of Biotechnology, Life Sciences Center, Vilnius University and Thermo Fisher Scientific | Justinas Slikas et al |
| EPI_ISL_728866; EPI_ISL_737928; EPI_ISL_737668; EPI_ISL_737549; EPI_ISL_737403; EPI_ISL_737524; EPI_ISL_737580; EPI_ISL_728839; EPI_ISL_728846; EPI_ISL_728865; EPI_ISL_729144; EPI_ISL_737832; EPI_ISL_737835; EPI_ISL_737629; EPI_ISL_737730; EPI_ISL_728801; EPI_ISL_729108; EPI_ISL_737422; EPI_ISL_737461; EPI_ISL_729122; EPI_ISL_729094 | Viollier AG | Department of Biosystems Science and Engineering, ETH Zürich | Chaoran Chen et al |
| EPI_ISL_476094; EPI_ISL_574902; EPI_ISL_603330; EPI_ISL_693975; EPI_ISL_603712; EPI_ISL_721971; EPI_ISL_693873; EPI_ISL_693872; EPI_ISL_693868; EPI_ISL_603475; EPI_ISL_535634; EPI_ISL_729164; EPI_ISL_539402; EPI_ISL_539447; EPI_ISL_539436; EPI_ISL_721925; EPI_ISL_539472; EPI_ISL_541464; EPI_ISL_541489; EPI_ISL_560429; EPI_ISL_693886; EPI_ISL_603391; EPI_ISL_693891; EPI_ISL_721800; EPI_ISL_476090; EPI_ISL_560461; EPI_ISL_535612; EPI_ISL_541469; EPI_ISL_512028; EPI_ISL_476132; EPI_ISL_486515; EPI_ISL_467007; EPI_ISL_603553; EPI_ISL_614895; EPI_ISL_614896; EPI_ISL_574944; EPI_ISL_451716; EPI_ISL_560549; EPI_ISL_466984; EPI_ISL_451856; EPI_ISL_468281; EPI_ISL_451751; EPI_ISL_489983; EPI_ISL_489961; EPI_ISL_489962; EPI_ISL_489966; EPI_ISL_486509; EPI_ISL_486521; EPI_ISL_486527; EPI_ISL_489975; EPI_ISL_500930; EPI_ISL_489978; EPI_ISL_489981; EPI_ISL_500935; EPI_ISL_500944; EPI_ISL_511992; EPI_ISL_510732; EPI_ISL_510733; EPI_ISL_489972; EPI_ISL_489965; EPI_ISL_486523; EPI_ISL_486522; EPI_ISL_489971; EPI_ISL_489973; EPI_ISL_498622; EPI_ISL_500899; EPI_ISL_451893; EPI_ISL_486479; EPI_ISL_451835; EPI_ISL_510794; EPI_ISL_512030; EPI_ISL_512035; EPI_ISL_539464; EPI_ISL_541513; EPI_ISL_489964; EPI_ISL_486534; EPI_ISL_489960; EPI_ISL_486510; EPI_ISL_486532; EPI_ISL_500892; EPI_ISL_539345; EPI_ISL_603455; EPI_ISL_721903; EPI_ISL_721676; EPI_ISL_523909; EPI_ISL_535622; EPI_ISL_516592; EPI_ISL_603617; EPI_ISL_603618; EPI_ISL_603643; EPI_ISL_614953; EPI_ISL_523908; EPI_ISL_486488; EPI_ISL_535617; EPI_ISL_451855; EPI_ISL_451906; EPI_ISL_574950; EPI_ISL_468295; EPI_ISL_468212; EPI_ISL_468223; EPI_ISL_476133; EPI_ISL_468286; EPI_ISL_560478; EPI_ISL_560519; EPI_ISL_560425; EPI_ISL_560428; EPI_ISL_560427; EPI_ISL_476103; EPI_ISL_489986; EPI_ISL_498623; EPI_ISL_500891; EPI_ISL_535633; EPI_ISL_560433; EPI_ISL_560544; EPI_ISL_541510; EPI_ISL_560434; EPI_ISL_541523; EPI_ISL_466999; EPI_ISL_467004; EPI_ISL_467016; EPI_ISL_476083; EPI_ISL_476100; EPI_ISL_476099; EPI_ISL_476097; EPI_ISL_476095; EPI_ISL_721695; EPI_ISL_539349; EPI_ISL_451886; EPI_ISL_614969; EPI_ISL_535627; EPI_ISL_467013; EPI_ISL_693922; EPI_ISL_541421; EPI_ISL_574937; EPI_ISL_560455; EPI_ISL_467000; EPI_ISL_468242; EPI_ISL_721728; EPI_ISL_603603; EPI_ISL_603610; EPI_ISL_603615; EPI_ISL_721794; EPI_ISL_729187; EPI_ISL_721787; EPI_ISL_721910; EPI_ISL_721801; EPI_ISL_539357; EPI_ISL_603280; EPI_ISL_721819; EPI_ISL_603588 | Viollier AG | Department of Biosystems Science and Engineering, ETH Zürich | Christian Beisel et al |
| EPI_ISL_447247; EPI_ISL_420841; EPI_ISL_420840; EPI_ISL_437340; EPI_ISL_437343; EPI_ISL_420031; EPI_ISL_435114; EPI_ISL_434678; EPI_ISL_420849; EPI_ISL_435161; EPI_ISL_437355; EPI_ISL_417948; EPI_ISL_417955; EPI_ISL_420035; EPI_ISL_417946; EPI_ISL_420854; EPI_ISL_420838 | Viral Respiratory Lab, National Institute for Biomedical Research (INRB) | Pathogen Sequencing Lab, National Institute for Biomedical Research (INRB) | Placide Mbala-Kingebeni et al A (https://virological.org/t/phylogenetic-analysis-of-sars-cov-2-in-drc/528) |
| EPI_ISL_513597; EPI_ISL_471412; EPI_ISL_471411; EPI_ISL_496498; EPI_ISL_496493; EPI_ISL_527495; EPI_ISL_527503; EPI_ISL_527550; EPI_ISL_495641; EPI_ISL_527540; EPI_ISL_527562; EPI_ISL_527551; EPI_ISL_591084; EPI_ISL_591083; EPI_ISL_527510; EPI_ISL_591085; EPI_ISL_591086 | Viral Respiratory Lab, National Institute for Biomedical Research (INRB) | Pathogen Sequencing Lab, National Institute for Biomedical Research (INRB) | Placide Mbala-Kingebeni et al B |
| EPI_ISL_522796 | Virginia DCLS | Virginia DCLS | Virginia DCLS et al |
| EPI_ISL_416488 | ViroGenetics - BSL3 Laboratory of Virology; Human Genome Variation Research Group & Genomics Centre MCB; Bioinformatics Research Group Department of Virology | ViroGenetics - BSL3 Laboratory of Virology; Human Genome Variation Research Group & Genomics Centre MCB; Bioinformatics Research Group Department of Virology | Aleksandra Milewska et al |
| EPI_ISL_428930 | ViroGenetics - BSL3 Laboratory of Virology; Human Genome Variation Research Group & Genomics Centre MCB; Bioinformatics Research Group; Wojewódzka Stacja Sanitarno-Epidemiologiczna w Krakowie | ViroGenetics - BSL3 Laboratory of Virology; Human Genome Variation Research Group & Genomics Centre MCB; Bioinformatics Research Group | Wojciech Branicki et al |
| EPI_ISL_435416; EPI_ISL_435413; EPI_ISL_435426; EPI_ISL_435427; EPI_ISL_435428; EPI_ISL_435430; EPI_ISL_435415; EPI_ISL_435412; EPI_ISL_435404; EPI_ISL_435403; EPI_ISL_435425; EPI_ISL_435424; EPI_ISL_435423; EPI_ISL_435418; EPI_ISL_435429; EPI_ISL_435421 | Virological Research Group, Szentágothai Research Centre | Bioinformatics Research Group, Szentágothai Research Centre | Péter Urbán et al |
| EPI_ISL_425842 | Virology Department, Royal Infirmary of Edinburgh, NHS Lothian / School of Biological Sciences, University of Edinburgh / Institute of Genetics and Molecular Medicine, University of Edinburgh | COVID-19 Genomics UK (COG-UK) Consortium | McHugh M et al A (https://virological.org/t/preliminary-analysis-of-sars-cov-2-importation-establishment-of-uk-transmission-lineages/507) |
| EPI_ISL_651428; EPI_ISL_493971; EPI_ISL_493973; EPI_ISL_493969; EPI_ISL_493965; EPI_ISL_507184 | Virology Department, Royal Infirmary of Edinburgh, NHS Lothian / School of Biological Sciences, University of Edinburgh / Institute of Genetics and Molecular Medicine, University of Edinburgh | COVID-19 Genomics UK (COG-UK) Consortium | McHugh M et al B |
| EPI_ISL_636987 | Virology Lab, National Institute for Biomedical Research (INRB) | Project group Epidemiology of Highly Pathogenic Microorganisms, Robert Koch-Institute | Jean-Jacques Muyembe Tamfum et al |
| EPI_ISL_494747 | Virology Lab,Department of Pathology, National Cheng Kung University Hospital | Virology Lab,Department of Pathology, National Cheng Kung University Hospital | Huey-Pin Tsai et al |
| EPI_ISL_677635; EPI_ISL_677636; EPI_ISL_677634; EPI_ISL_625456; EPI_ISL_508862; EPI_ISL_508863 | Virology Unit, Institut Pasteur de Madagascar | Virology Unit, Institut Pasteur de Madagascar | Christian Ranaivoson et al |
| EPI_ISL_411902 | Virology Unit, Institut Pasteur du Cambodge. | Virology Unit, Institut Pasteur du Cambodge (Sequencing done by: Jessica E Manning/Jennifer A Bohl at Malaria and Vector Research Research Laboratory, National Institute of Allergy and Infectious Diseases and Vida Ahyong from Chan-Zuckerberg Biohub) | Erik A Karlsson et al (https://dx.doi.org/10.2807/1560-7917.ES.2020.25.13.2000305) |
| EPI_ISL_467666 | Virology lab, NIC, NCCD, Ulaanbaatar, Mongolia | National Centre for Communicable Deseases (NCCD) | Naranzul Ts et al |
| EPI_ISL_416458 | Virology laboratory Ministry of Health Kuwait sequenced at Dasman Diabetes Institute | Dasman Diabetes Institute | Fahd Al-Mulla et al |
| EPI_ISL_610245 | Virology, National Institute for Biological Standards and Control | Virology, National Institute for Biological Standards and Control | Mee et al |
| EPI_ISL_707948; EPI_ISL_708013; EPI_ISL_708015; EPI_ISL_707943; EPI_ISL_707942; EPI_ISL_707991; EPI_ISL_708022; EPI_ISL_707969; EPI_ISL_708023; EPI_ISL_708024; EPI_ISL_708006; EPI_ISL_708007; EPI_ISL_707940; EPI_ISL_708012; EPI_ISL_707989; EPI_ISL_708001; EPI_ISL_707975; EPI_ISL_707980; EPI_ISL_707978; EPI_ISL_707979; EPI_ISL_707970; EPI_ISL_707974; EPI_ISL_707971; EPI_ISL_707999; EPI_ISL_708011; EPI_ISL_707997; EPI_ISL_708019; EPI_ISL_707966; EPI_ISL_707941; EPI_ISL_707946; EPI_ISL_707951; EPI_ISL_707963; EPI_ISL_707968; EPI_ISL_708021; EPI_ISL_707947; EPI_ISL_707949; EPI_ISL_707952; EPI_ISL_707954; EPI_ISL_708003; EPI_ISL_708016; EPI_ISL_708017; EPI_ISL_708009; EPI_ISL_707959; EPI_ISL_708004; EPI_ISL_707957; EPI_ISL_708008; EPI_ISL_708025; EPI_ISL_708018; EPI_ISL_707964; EPI_ISL_707958; EPI_ISL_707967; EPI_ISL_707960; EPI_ISL_707961; EPI_ISL_707965; EPI_ISL_708020; EPI_ISL_707990; EPI_ISL_707993; EPI_ISL_707985; EPI_ISL_707986; EPI_ISL_708014; EPI_ISL_707988; EPI_ISL_707987; EPI_ISL_707983; EPI_ISL_707972; EPI_ISL_708002; EPI_ISL_707992; EPI_ISL_707981; EPI_ISL_707982; EPI_ISL_707995; EPI_ISL_707994; EPI_ISL_707962; EPI_ISL_707996; EPI_ISL_708000; EPI_ISL_708010; EPI_ISL_707976; EPI_ISL_707977; EPI_ISL_707998 | Virology, Universitätsklinikum des Saarlandes | Epigenetics, Saarland University | Kathrin Kattler et al |
| EPI_ISL_568579 | Virus Molecular Laboratory of the Microbiology and Virology Department | INMI Lazzaro Spallanzani IRCCS | Cesare E.M. Gruber et al B |
| EPI_ISL_722201 | Vitalis Mostar | Alea Genetic Center | Salihefendic L. et al |
| EPI_ISL_524015; EPI_ISL_524026; EPI_ISL_524032; EPI_ISL_733015; EPI_ISL_733017; EPI_ISL_733279; EPI_ISL_733259; EPI_ISL_733251; EPI_ISL_733260; EPI_ISL_733274; EPI_ISL_732983; EPI_ISL_732978; EPI_ISL_733021; EPI_ISL_596230; EPI_ISL_733019; EPI_ISL_733300; EPI_ISL_507227; EPI_ISL_507246; EPI_ISL_507236; EPI_ISL_507224; EPI_ISL_507233; EPI_ISL_507256; EPI_ISL_507259; EPI_ISL_733010; EPI_ISL_507258; EPI_ISL_507269; EPI_ISL_524021; EPI_ISL_507279; EPI_ISL_507288; EPI_ISL_507289; EPI_ISL_524039; EPI_ISL_415710; EPI_ISL_733297; EPI_ISL_507226 | WHO National Influenza Centre Russian Federation | WHO National Influenza Centre Russian Federation | Andrey Komissarov et al A |
| EPI_ISL_430070; EPI_ISL_430111; EPI_ISL_427313; EPI_ISL_427339; EPI_ISL_430110; EPI_ISL_450247; EPI_ISL_450264; EPI_ISL_450253; EPI_ISL_450263; EPI_ISL_420080; EPI_ISL_430071 | WHO National Influenza Centre Russian Federation | WHO National Influenza Centre Russian Federation | Andrey Komissarov et al B (https://dx.doi.org/10.1101/2020.07.14.20150979) |
| EPI_ISL_548101; EPI_ISL_548064 | Waikato Hospital | Institute of Environmental Science and Research (ESR) | Xiaoyun Ren et al |
| EPI_ISL_418114; EPI_ISL_420962; EPI_ISL_432421; EPI_ISL_445497; EPI_ISL_419438 | Wales Specialist Virology Centre | Public Health Wales Microbiology Cardiff | Catherine Moore et al A (https://virological.org/t/preliminary-analysis-of-sars-cov-2-importation-establishment-of-uk-transmission-lineages/507) |
| EPI_ISL_577459; EPI_ISL_560262 | Wales Specialist Virology Centre Sequencing lab: Pathogen Genomics Unit | COVID-19 Genomics UK (COG-UK) Consortium | Catherine Moore et al B |
| EPI_ISL_463597; EPI_ISL_463623; EPI_ISL_463592; EPI_ISL_463603 | Washington State Department of Health | Seattle Flu Study | Chu et al et al |
| EPI_ISL_497623; EPI_ISL_495899 | Washington State Department of Health | Seattle Flu Study | Deborah A. Nickerson et al |
| EPI_ISL_456401; EPI_ISL_456161 | Wellington SCL | Institute of Environmental Science and Research (ESR) | Matt Storey et al B |
| EPI_ISL_451346; EPI_ISL_451370; EPI_ISL_451344; EPI_ISL_451334; EPI_ISL_451345 | West China Hospital of Sichuan University | State Key Laboratory of Biotherapy of Sichuan University | Baowen Du et al |
| EPI_ISL_704807; EPI_ISL_724210; EPI_ISL_704804; EPI_ISL_540780; EPI_ISL_484620; EPI_ISL_461597 | West of Scotland Specialist Virology Centre, NHSGGC / MRC-University of Glasgow Centre for Virus Research | COVID-19 Genomics UK (COG-UK) Consortium | Ana da Silva Filipe et al A |
| EPI_ISL_453070; EPI_ISL_439014; EPI_ISL_456824 | West of Scotland Specialist Virology Centre, NHSGGC / MRC-University of Glasgow Centre for Virus Research | COVID-19 Genomics UK (COG-UK) Consortium | Ana da Silva Filipe et al B (https://virological.org/t/preliminary-analysis-of-sars-cov-2-importation-establishment-of-uk-transmission-lineages/507) |
| EPI_ISL_408670 | Wisconsin Department of Health Services | Pathogen Discovery, Respiratory Viruses Branch, Division of Viral Diseases, Centers for Disease Control and Prevention | Jing Zhang et al A (https://dx.doi.org/10.1101/2020.03.09.20032896) |
| EPI_ISL_708812; EPI_ISL_708813 | World Medical Hospital | National Institute of Health, Department of Medical Sciences, Ministry of Public Health, Thailand | Pilailuk Okada et al |
| EPI_ISL_765957 | Worobey Lab, Department of Ecology and Evolutionary Biology, University of Arizona | Worobey Lab, Department of Ecology and Evolutionary Biology, University of Arizona | Brendan Larsen et al |
| EPI_ISL_454981; EPI_ISL_454928; EPI_ISL_454955; EPI_ISL_454956 | Wuhan Chain Medical Labs (CMLabs) | State Key Laboratory of Biotherapy of Sichuan University | Baowen Du et al |
| EPI_ISL_412899 | Wuhan Jinyintan Hospital | Hubei Provincial Center for Disease Control and Prevention | Bin Fang et al (https://dx.doi.org/10.1101/2020.03.04.976662) |
| EPI_ISL_402124; EPI_ISL_402129 | Wuhan Jinyintan Hospital | Wuhan Institute of Virology, Chinese Academy of Sciences | Peng Zhou et al |
| EPI_ISL_614260; EPI_ISL_614261; EPI_ISL_614259; EPI_ISL_614264 | Wyoming Public Health Laboratory | Center for Global Health, University of New Mexico Health Sciences Center | Daryl Domman et al |
| EPI_ISL_765966; EPI_ISL_765965; EPI_ISL_756164; EPI_ISL_756189 | Wyoming Public Health Laboratory | Wyoming Public Health Laboratory | Noah Hull et al |

| | | | |
|---|---|---|---|
| EPI_ISL_631387; EPI_ISL_733524; EPI_ISL_733527; EPI_ISL_733526; EPI_ISL_718042; EPI_ISL_539606; EPI_ISL_539609; EPI_ISL_718045; EPI_ISL_718087; EPI_ISL_718089; EPI_ISL_718098; EPI_ISL_539591; EPI_ISL_666637; EPI_ISL_718061; EPI_ISL_631358; EPI_ISL_718068; EPI_ISL_733517; EPI_ISL_718070; EPI_ISL_718053; EPI_ISL_718058; EPI_ISL_539594; EPI_ISL_539595; EPI_ISL_733525; EPI_ISL_631314; EPI_ISL_631322; EPI_ISL_631315; EPI_ISL_539588; EPI_ISL_539589; EPI_ISL_631375; EPI_ISL_631348; EPI_ISL_631350; EPI_ISL_631351; EPI_ISL_718032; EPI_ISL_631339; EPI_ISL_631320; EPI_ISL_718033; EPI_ISL_718034; EPI_ISL_666639; EPI_ISL_666640; EPI_ISL_666641; EPI_ISL_666642; EPI_ISL_539603; EPI_ISL_539607; EPI_ISL_666624; EPI_ISL_718037; EPI_ISL_718075; EPI_ISL_718039; EPI_ISL_733521; EPI_ISL_718059; EPI_ISL_666635; EPI_ISL_631359; EPI_ISL_718035; EPI_ISL_718030; EPI_ISL_718031; EPI_ISL_718104; EPI_ISL_631323; EPI_ISL_718092; EPI_ISL_718038; EPI_ISL_631344; EPI_ISL_631327; EPI_ISL_539593; EPI_ISL_539596; EPI_ISL_631363; EPI_ISL_718103; EPI_ISL_718071; EPI_ISL_631362; EPI_ISL_631354; EPI_ISL_718080; EPI_ISL_718081; EPI_ISL_718083; EPI_ISL_718094; EPI_ISL_733506; EPI_ISL_718095; EPI_ISL_718076; EPI_ISL_718077; EPI_ISL_718072; EPI_ISL_718078; EPI_ISL_631321; EPI_ISL_631305; EPI_ISL_718082; EPI_ISL_631311; EPI_ISL_631308; EPI_ISL_631309; EPI_ISL_631306; EPI_ISL_666643; EPI_ISL_666632; EPI_ISL_718044; EPI_ISL_539601; EPI_ISL_631316; EPI_ISL_631343; EPI_ISL_666629; EPI_ISL_631310; EPI_ISL_631307; EPI_ISL_631337; EPI_ISL_631334; EPI_ISL_666631; EPI_ISL_631347; EPI_ISL_666633; EPI_ISL_666634; EPI_ISL_539585; EPI_ISL_539598; EPI_ISL_539597; EPI_ISL_539582; EPI_ISL_539584; EPI_ISL_539581; EPI_ISL_539578; EPI_ISL_539580; EPI_ISL_539579; EPI_ISL_539590; EPI_ISL_539583; EPI_ISL_718096; EPI_ISL_718079; EPI_ISL_666636; EPI_ISL_539577; EPI_ISL_666648; EPI_ISL_666644; EPI_ISL_733507; EPI_ISL_733501; EPI_ISL_733514; EPI_ISL_631342; EPI_ISL_631324; EPI_ISL_631325; EPI_ISL_539599; EPI_ISL_631352; EPI_ISL_631326; EPI_ISL_733504; EPI_ISL_733505; EPI_ISL_539586; EPI_ISL_631331; EPI_ISL_718069; EPI_ISL_631361; EPI_ISL_666628; EPI_ISL_631364; EPI_ISL_718036; EPI_ISL_666647; EPI_ISL_631328; EPI_ISL_631356; EPI_ISL_631357; EPI_ISL_631319; EPI_ISL_631346; EPI_ISL_631330; EPI_ISL_631333; EPI_ISL_666630; EPI_ISL_718041; EPI_ISL_666623; EPI_ISL_718097; EPI_ISL_733523; EPI_ISL_631318; EPI_ISL_718074; EPI_ISL_733516; EPI_ISL_733515; EPI_ISL_539608; EPI_ISL_539592; EPI_ISL_631355; EPI_ISL_631349; EPI_ISL_666646; EPI_ISL_539602; EPI_ISL_631353; EPI_ISL_666649; EPI_ISL_539612; EPI_ISL_718102; EPI_ISL_718046; EPI_ISL_666645; EPI_ISL_631341; EPI_ISL_539604; EPI_ISL_733502; EPI_ISL_718084; EPI_ISL_718062; EPI_ISL_718063; EPI_ISL_733503; EPI_ISL_666650; EPI_ISL_631332; EPI_ISL_718028; EPI_ISL_718085; EPI_ISL_718029; EPI_ISL_666627; EPI_ISL_631345; EPI_ISL_631360; EPI_ISL_718099; EPI_ISL_718101; EPI_ISL_733519; EPI_ISL_666625; EPI_ISL_733513; EPI_ISL_718060; EPI_ISL_631340; EPI_ISL_718049; EPI_ISL_718051; EPI_ISL_718056; EPI_ISL_718043; EPI_ISL_718057; EPI_ISL_539613; EPI_ISL_539614; EPI_ISL_539600; EPI_ISL_539587; EPI_ISL_539610; EPI_ISL_539615; EPI_ISL_666651; EPI_ISL_666652; EPI_ISL_718064; EPI_ISL_718065; EPI_ISL_718067; EPI_ISL_718066; EPI_ISL_718040; EPI_ISL_718054; EPI_ISL_718055; EPI_ISL_718100; EPI_ISL_733511; EPI_ISL_733512; EPI_ISL_733509; EPI_ISL_733510; EPI_ISL_733508; EPI_ISL_631312; EPI_ISL_631338; EPI_ISL_718027; EPI_ISL_631313; EPI_ISL_666638; EPI_ISL_631317; EPI_ISL_631372; EPI_ISL_631374; EPI_ISL_631370; EPI_ISL_631365; EPI_ISL_631368; EPI_ISL_631371; EPI_ISL_631369; EPI_ISL_631376; EPI_ISL_631373; EPI_ISL_631367; EPI_ISL_631366; EPI_ISL_718091; EPI_ISL_539605; EPI_ISL_539611; EPI_ISL_631329; EPI_ISL_631304; EPI_ISL_733520; EPI_ISL_733522; EPI_ISL_733518; EPI_ISL_718050; EPI_ISL_666626; EPI_ISL_718073; EPI_ISL_718086; EPI_ISL_718088; EPI_ISL_718090; EPI_ISL_718052; EPI_ISL_718048; EPI_ISL_718047; EPI_ISL_718093 | ZOTZ KLIMAS MVZ Düsseldorf-Centrum GbR ÜBAG für Labormedizin, Genetik, Zytologie, Pathologie | Center of Medical Microbiology, Virology, and Hospital Hygiene, University of Duesseldorf | Maximilian Damagnez et al |
| EPI_ISL_475903; EPI_ISL_475902; EPI_ISL_475904; EPI_ISL_475895 | Zentralinstitut für medizinische und chemische Labordiagnostik, Universitätskliniken Insbruck | Bergthaler laboratory, CeMM Research Center for Molecular Medicine of the Austrian Academy of Sciences | Alexandra Popa et al B (https://dx.doi.org/10.1101/2020.07.15.204339 v1) |
| EPI_ISL_404228 | Zhejiang Provincial Center for Disease Control and Prevention | Department of Microbiology, Zhejiang Provincial Center for Disease Control and Prevention | Yanjun Zhang et al |
| EPI_ISL_660070 | Zurita & Zurita Laboratorios | Zurita & Zurita Laboratorios | Gabriela Sevillano Camilo Zurita-Salinas Karen Loaiza David Ortega-Paredes Jeannete Zurita et al |
| EPI_ISL_681702 | Zurita & Zurita Laboratorios | Zurita & Zurita Laboratorios | Gabriela Sevillano et al |
| EPI_ISL_586541 | Zydus Medical Hospital,Dahod | Gujarat Biotechnology Research Centre | Zuber Saiyed et al |
| EPI_ISL_468148; EPI_ISL_468134; EPI_ISL_468145; EPI_ISL_468149; EPI_ISL_468156; EPI_ISL_468147; EPI_ISL_468151; EPI_ISL_468155; EPI_ISL_468153; EPI_ISL_468152; EPI_ISL_468146; EPI_ISL_468136; EPI_ISL_468137; EPI_ISL_468138; EPI_ISL_468158; EPI_ISL_468140; EPI_ISL_468139; EPI_ISL_468143; EPI_ISL_468141; EPI_ISL_468157; EPI_ISL_468144; EPI_ISL_468142; EPI_ISL_468135 | [Romania, Bucharest] National Institute for Infectious Diseases "Prof. Dr. Matei Balş" | [Romania, Bucharest] National Institute for Infectious Diseases "Prof. Dr. Matei Balş" | Leontina Banica et al |
| EPI_ISL_560578 | hôpital | National Reference Center for Viruses of Respiratory Infections, Institut Pasteur, Paris | Sylvie Behillil et al |
| EPI_ISL_454574 | nstitute for Public Health | Laboratory for advanced genomics | Filip Rokić et al |
| EPI_ISL_524800 | Evandro Chagas Institute | Evandro Chagas Institute | Santos et al |
